# Robustness of Wolbachia-mediated incompatible-insect technique to future climate change scenarios

**DOI:** 10.64898/2026.06.26.26356650

**Authors:** Lin Geng, Perran A. Ross, Cai Yu, Tao Huang, Jo Yi Chow, Zihao Wang, Esther Li Wen Choo, Chia-Chen Chang, Lisa Couper, Xinyue Gu, Ary Hoffmann, Jue Tao Lim

**Author notes:** Correspondence to Ary Hoffmann. Correspondence to Jue Tao Lim. These authors contributed equally (Lin Geng, Perran A. Ross, Cai Yu).

## Abstract

*Wolbachia*-mediated incompatible-insect technique (IIT) via wAlbB, wMel or *w*Pip/*w*AlbA/*w*AlbB strains are promising approaches for suppressing wildtype *Aedes* mosquitoes and therefore *Aedes*-borne diseases. Yet, the effectiveness of this technique under climate change remains uncertain. Here, we evaluate the long-term robustness of male *Wolbachia*-infected mosquito releases to suppress wildtype *Aedes aegypti* and *Ae. albopictus* populations across future climate scenarios across diverse geographical regions. We compiled large publicly available datasets on *Aedes* abundance across Singapore, China, the European Union and the United States, historical and projected climatic conditions in these regions and conducted experiments to test the thermal stability of cytoplasmic incompatibility in *Wolbachia*-infected male *Aedes aegypti* and *albopictus*. A climatically-driven entomological model was developed and calibrated using a Bayesian approach to model observed *Aedes* population dynamics and infer area-specific climate-driven variation in mosquito life-history traits. We back-inferred historical mosquito abundance and projected mosquito abundance in future climate change scenarios incorporating experimental and locally inferred entomological parameters and then simulated the counterfactual implementation of IIT in these regions. We find that *Aedes* populations are projected to increase in most regions across all climate change scenarios from 2050–2100 even under high heat conditions in the absence of interventions. While we found that IIT can suppress wild-type populations effectively across all future scenarios and in high heat conditions, effectiveness was found to depend heavily on mosquito emigration rates, overflooding ratios, release intervals and release strategies Extensive robustness checks confirmed that the model reproduced historical temporal trends, captured the influence of individual parameters on outcome and was sensitive to changes in values of inferred parameters and implement policy. These findings demonstrate that IIT may be a robust vector control tool under future climate conditions.

**Research in context:** *Evidence before this study:* Incompatible Insect Technique (IIT) is a promising strategy for vector control. IIT involves the release of male mosquitoes infected with the *Wolbachia* bacterium; when these males mate with wild females without *Wolbachia* or infected with a different strain, cytoplasmic incompatibility yields non-viable offspring, thereby suppressing mosquito populations and reducing dengue transmission. We searched Embase, MEDLINE, Global Health, and PubMed for publications between database inception and Dec 1, 2025, with the search terms capturing the type of intervention (((“*Wolbachia*”) OR (“incompatible insect technique”)) and (“intervention”)) and (“climate change”)) as well as the type of outcome (“*Aegypti*”) of interest for our study. We also contacted key field experts for relevant articles. The search returned 26 articles. 1 study evaluated the robustness of *Wolbachia* replacement in future heat scenarios. No study has ascertained the long-term robustness of *Wolbachia*-mediated IIT deployments in the context of climate change.

*Added value of this study:* This study ascertains the long-term viability of *Wolbachia*-IIT via wAlbB, wMel or *w*Pip/*w*AlbA/*w*AlbB strains to suppress mosquito populations under future climate change scenarios. Our approach advances the field by providing first-of-its-kind evidence that IIT can be a highly effective climate-resilient, and sustainable *Aedes*-control tool with global applicability.

*Implications of all the available evidence:* Our findings demonstrate strong evidence that IIT can be a robust vector control tool under future climate conditions in the European Union, United States, China and Singapore. Policy makers can consider *Wolbachia*-mediated IIT as a strong tool to combat *Aedes*-borne diseases with long-term efficacy.

## Introduction

Dengue is the fastest-growing vector-borne disease globally [1,2]. Dengue’s principle vectors, *Aedes aegypti* and *Aedes albopictus* mosquitoes, are increasing in abundance and expanding in geographical range, particularly across tropical and subtropical regions [3]. Rapid urbanisation, rising urban populations, and increasingly favourable climatic conditions for mosquito breeding amplify both host and vector densities and intensify host-vector interactions [4]. Collectively, these factors and increasing mobility across the globe elevate transmission potential and the attendant public-health and economic burden [5–12]. Looking ahead, climate change is expected to accelerate mosquito proliferation, with modelling evidence projecting *Aedes* invasion into new temperate regions and continued expansion through 2080 in high emission scenarios [1,13]. Concomitantly, human population growth is expected to concentrate in *Aedes*-established areas, placing roughly half of the global population at high risk of arbovirus disease [14]

Vector control is the primary means of dengue prevention. Other measures, including vaccination, remain constrained by safety concerns, discontinuation of commercial production [15,16], uneven distribution, waning long-term effectiveness [17], and the lack of evidence surrounding tetravalent efficacy [18–21]. Conventional vector control measures deployed for outbreak response are often ineffective in addressing long-term dengue risk; with heterogenous entomological and epidemiological efficacy, concerns with sustainability, insecticide resistance, and often requires prolonged implementation to achieve reductions in mosquito abundance [22–30].

*Wolbachia* Incompatible Insect Technique (IIT) offers a promising complementary strategy for vector control. IIT involves the release of male mosquitoes infected with the *Wolbachia* bacterium; when these males mate with wild females without *Wolbachia* or infected with a different strain, cytoplasmic incompatibility (CI) yields non-viable offspring, thereby suppressing mosquito populations and reducing dengue transmission. However, as sex-sorting is imperfect, the occasional release of females may occur, establishing infection in the wild and undermining suppression [31]. Many operational programmes have therefore combined IIT with the Sterile Insect Technique (SIT) to mitigate this risk [32–34]. IIT-SIT reduces the likelihood of unintentional fertile female release and preserves higher mating competitiveness compared to strategies based on irradiation alone given that a lower dose of radiation is required to sterilize females compared to males [32,35,36]. Trial deployments of IIT and IIT-SIT strategies have reduced *Aedes* abundance in simulations, lab assessments, and field trials across French Polynesia, the United States (US), Mexico, Spain, Italy, Thailand, mainland China and Singapore [34,37–46]. In Singapore, IIT-SIT deployment has also been associated with significant declines in dengue incidence and risk [47–49]. Simulation studies have also proposed strategies to optimise releases for IIT for larger-scale deployment [50].

Yet, rising temperatures and more frequent extreme weather events may alter mosquito dynamics and intervention success [51]. Prior modelling studies have projected increases in *Aedes* abundance and/or interventions against *Aedes*-borne diseases under different climate change scenarios [10,52–62]. For example, Bonnin et al. (2022) developed a climate-sensitive process-based model to show increases in *Ae. aegypti* and *albopictus* abundance in future climate change scenarios across Southeast Asia [62]. Vàsquez et al. (2023) predicted that the effectiveness of *w*Mel *Wolbachia* replacement in *Ae. aegypti* (which also involves deliberate release of mosquitoes) is only robust in the near term to 2050 in Australia and Vietnam, and may fail under prolonged heat waves [63]. However, current models looking at the future robustness of *Wolbachia-*based technologies were calibrated to lab data and are inconsistent with field outcomes [63]. These models relied on possibly overestimated thermal thresholds which can upwardly bias the thermal tolerance of *w*Mel replacement [63]. These existing models are also not calibrated to observed mosquito abundance or to area-specific life-history traits [64]. Moreover, temperature effects on CI during the adult stage are particularly relevant for IIT-based programmes (as opposed to replacement programmes), but have yet to be tested. Consequentially, the extent to which thermal sensitivity may influence the effectiveness of IIT-based vector control programmes is unclear. To support assessment of *Wolbachia*-based IIT rollout globally, an evidence-based entomological and modelling assessment of its climate robustness is needed.

To address this gap, we predicted the long-term robustness of *Wolbachia*-mediated IIT deployments under future climate scenarios. We integrated extensive, routinely collected *Aedes* abundance datasets with historical and projected climate records, linked these to experimental evidence on *Wolbachia*-induced cytoplasmic incompatibility at high temperatures near mortality, and applied a Bayesian climate-driven process-based modelling framework to project intervention performance. We also undertook extensive sensitivity analyses to ascertain the generalisability of our results to changes in parameters and release strategies. By grounding projections in real-world surveillance and experimental validation, this study provides guidance on the long-term viability of IIT under changing climates, informing the future scale-up of such *Wolbachia*-based technologies.

## Methods

### *Aedes* abundance data

We compiled entomological surveillance data across China (CN), Singapore (SG), the European Union (EU), and the United States (US), as detailed in Table 1. In Singapore, Gravitrap *aegypti* index (GAI) data from 2019 to 2024 was obtained from the National Environmental Agency of Singapore. GAI measures adult female *Ae. aegypti* abundance by calculating the weekly number of female adults captured in Gravitraps [65,66] normalized by the total number of functioning Gravitraps deployed in each spatial area. For Mainland China, bi-weekly or monthly Breteau Index (BI) and Mosq-Ovitrap Index (MOI) data from 2016 to 2019 were used as published by China CDC at the province level [9]. Where only graphical results were available, values were extracted using WebPlotDigitizer [67]. BI measures the number of positive containers or other water bodies per house inspected, and MOI reflects the number of positive ovitraps among all effective ovitraps. Species information was not recorded for the China dataset and we assumed to be *Ae. albopictus* which is the dominant *Aedes* species across this region based on prior studies [68]. In Europe, the VectAbundance database was used which compiled weekly *Ae. albopictus* egg counts from ovitraps across multiple areas from Albania, France, Italy and Switzerland between 2010 and 2022. The raw numbers of mosquitoes were collected biweekly or weekly by various institutes using similar protocols, and were standardized through temporal downscaling [69]. In the United States, monthly county-level data of *Ae. aegypti* and *Ae. albopictus* per trap night (ATN) from 2006 to 2019 were used as summarized by U.S. Centers for Disease Control and Prevention [70]. ATN was calculated as the number of adults identified per cumulative number of nights that traps were operational. We used CO_2_ baited traps including CDC and ABC light traps and Fay-Prince traps for the analysis, and half the index value was taken to reflect the female population [71].

**Table 1.**
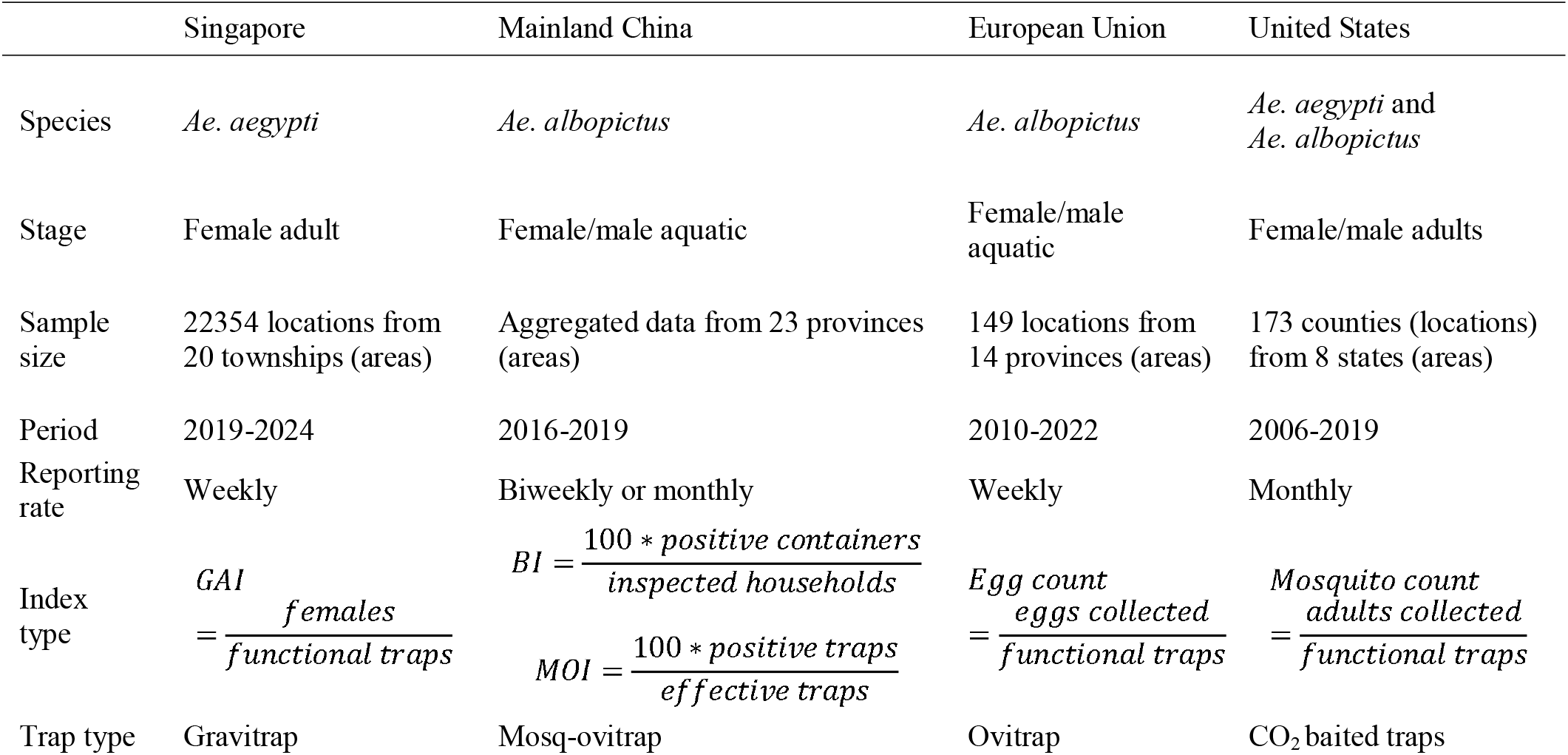
Summary of mosquito abundance datasets across different study regions. Location pertains to the finest unit used to track where traps were deployed. Area pertains to the administrative area that aggregates all trap locations together.

|  | Singapore | Mainland China | European Union | United States |
| --- | --- | --- | --- | --- |
| Species | *Ae. aegypti* | *Ae. albopictus* | *Ae. albopictus* | *Ae. aegypti* and *Ae. albopictus* |
| Stage | Female adult | Female/male aquatic | Female/male aquatic | Female/male adults |
| Sample size | 22354 locations from 20 townships (areas) | Aggregated data from 23 provinces (areas) | 149 locations from 14 provinces (areas) | 173 counties (locations) from 8 states (areas) |
| Period | 2019-2024 | 2016-2019 | 2010-2022 | 2006-2019 |
| Reporting rate | Weekly | Biweekly or monthly | Weekly | Monthly |
| Index type | $GAI = \frac{\text{females}}{\text{functional traps}}$ | $BI = \frac{100 * \text{positive containers}}{\text{inspected households}}$<br>$MOI = \frac{100 * \text{positive traps}}{\text{effective traps}}$ | $\text{Egg count} = \frac{\text{eggs collected}}{\text{functional traps}}$ | $\text{Mosquito count} = \frac{\text{adults collected}}{\text{functional traps}}$ |
| Trap type | Gravitrap | Mosq-ovitraps | Ovitraps | CO<sub>2</sub> baited traps |

We employed several exclusion criteria to ensure data consistency across regions. In Singapore, the area of Tengah was excluded due to an extremely low number of recorded observations. Additionally, ten ongoing IIT-SIT trial sites—Bedok, Bukit Batok, Choa Chu Kang, Geylang, Hougang, Punggol, Sengkang, Tampines, Woodlands, and Yishun—were removed as surveillance data from these areas no longer reflect natural *Aedes* populations. MOI was the only available index for Beijing and Guizhou, and preferred over BI in Guangdong due to its sensitivity in association with dengue outbreaks, since MOI also broadly sampled from traps outside households [9]. For the United States, *Ae. aegypti* abundance data were selected for Arizona, California, Florida, and Texas, while *Ae. albopictus* data were selected for Connecticut, Georgia, New Jersey, New York, North Carolina. The dominant species for each available state was chosen for model calibration and simulations.

### Climate data and SSPs

Shared Socioeconomic Pathways (SSPs) were used to characterise plausible long-term climate scenarios. SSPs 1 to 5 segmented future climate based on demographic trends, human development, economic and lifestyle patterns, the evolution of policies and institutions, technological change, and the state of the environment and natural resources [72]. We used publicly available climate projections from the Coupled Model Intercomparison Project Phase 6 (CMIP6) because it provided the most up-to-date simulations with advanced representations of climate processes and high-resolution global coverage [73]. Daily climate projections from the ISIMIP3b protocol were used as it provided bias-corrected CMIP6 climate projections. The considered SSPs were:

1. SSP126: the low-emissions scenario from the SSP1 “Sustainability” family, where the world shifts toward a more sustainable path. It is associated with a radiative forcing level of 2.6 W/m^2^ by 2100, reflecting strong mitigation efforts and reduced fossil fuel use.
2. SSP370: the medium-to-high emissions scenario from the SSP3 “Regional Rivalry” family, where countries prioritize regional interests, resulting in limited international cooperation, slow economic development, and moderate to high emissions of radiative forcing level at 7.0 W/m^2^ by 2100.
3. SSP585: the highest-emissions scenario and belongs to the SSP5 “Fossil-fuelled Development” family, characterized by rapid economic growth driven by heavy reliance on fossil fuels. It reaches a radiative forcing level of 8.5 W/m^2^ by 2100, marking the upper end of the SSP scenario range.

ISIMIP3b provided bias corrected climate projections of five general circulate models (GCMs) using land data from WFDE5 and ocean data from ERA5 [74]. The historical and future periods were segmented at the year 2015 to align with the CMIP6 data, with ISIMIP3b providing future climate scenarios (SSP126, SSP370, and SSP585) based on GCMs including GFDL-ESM4, IPSL-CM6A-LR, MPI-ESM1-2-HR, MRI-ESM2-0, and UKESM1-0-LL. The historical and future projections were integrated across 2015 till 2100 to create continuous climate time series under each SSP and binned across 2025-2050, 2050-2075 and 2075-2100. Owing to the availability of climate projections and the lack of microclimate surveillance data, we selected near-surface air temperature and precipitation as the key climatic drivers in our climate-driven entomological model. The selection of these variables maximized the realism of the model by enabling integration of empirical laboratory evidence with field-based mosquito abundance surveillance and corresponding climatic conditions. Daily mean temperatures were converted to Celsius and daily precipitation to daily mean kg/m^2^. Data were obtained at the spatial resolution of the geographical grid per 0.5°.

### Stability of cytoplasmic incompatibility in *w*MelM- and *w*AlbB-infected male *Ae. aegypti* mosquitoes

We performed experiments to test the stability of *Wolbachia*-induced cytoplasmic incompatibility in adults under extreme temperatures and suppression by antibiotic treatment. *w*MelM [75], *w*AlbB [76,77] and uninfected *Ae. aegypti* with a Cairns, Australia genetic background were reared under standard laboratory conditions (4L trays containing 400 larvae, diet of Hikari Tropical Sinking Wafers, Kyorin Food, Himeji, Japan) at 29°C with a 12:12 light:dark cycle used to reflect mass-rearing conditions in Singapore [78]. Pupae were sexed and released into cages. All treatments within an experiment were completed in a single block with mosquitoes reared under identical conditions. Mosquito age, treatment and mating history were all controlled between mosquito lines.

In the extreme temperature experiment, adult males (2-3 d old) from each population were exposed to one of three different temperature treatments in PHCbi MIR-254 incubators: (1) acute heat exposure for 1 hr at 41°C, (2)chronic heat exposure for 3 d at 34°C and (3) controls maintained at 29°C. Heat exposure treatments were chosen based on pilot experiments and previous studies [79,80] where temperatures above these conditions resulted in near-complete mortality. Immediately following exposure, males from all treatments were crossed to uninfected females reared at 29°C, with each cross involving 4 replicate cages with 20 males and 20 females per cage. Crosses involving males from the control and the acute heat exposure treatment were undertaken at 29°C while crosses involving males from the chronic heat exposure treatment took place at 34°C. Females were blood fed one day post-mating on the forearm of a human according to procedures approved by the University of Melbourne Human Ethics Committee (project ID 28583). In the chronic heat exposure treatment, oviposition also occurred at 34°C. Eggs were collected from groups of females on sandpaper strips, partially dried and stored at 29°C, then hatched 3 days after collection. CI was measured by scoring the proportion of viable eggs compared to compatible crosses (uninfected female x uninfected male) in each temperature treatment. To account for potential effects of high temperatures during mating and oviposition on fertility, uninfected female x uninfected male crosses were included for all temperature treatments.

One week after the first blood feed, females were blood fed again and the same procedure was repeated to assess CI in a second gonotrophic cycle. Females from the chronic heat exposure treatment were also maintained at 34°C for the second gonotrophic cycle. To test for the long-term durability of CI in males and potential effects of sperm depletion, the males used in the first set of crosses were crossed to additional cages of unmated uninfected females one week after the first set of crosses. In the second set of crosses, males from the chronic heat exposure treatment had been maintained at 34°C for 10 days while males from the acute heat exposure were not treated again after the initial 41°C heat shock. These crosses involved 10 males and 50 females per replicate cage to ensure some level of sperm depletion in the males. To check that females from incompatible crosses were inseminated, we dissected and observed the spermathecae from subsets of females from each cross (∼5 females per treatment).

We then performed an antibiotic treatment experiment to further test the robustness of CI. Treating adults with tetracycline is expected to substantially reduce *Wolbachia* densities, but whether direct suppression of *Wolbachia* during the adult stage leads to decreased CI expression remains to be tested. In thisexperiment, adult males (2-3 d old) from each population were provided with 0 or 8 mg/mL tetracycline hydrochloride in a 10% sucrose solution for one week. This concentration was chosen as it did not cause substantial mortality but was higher than concentrations typically used for curing *Wolbachia* from mosquitoes (1-2 mg/mL). Males from both treatments were then crossed to uninfected females at 29°C with 4 replicate cages with 20 males and 20 females per cage. Eggs were collected and hatch proportions were scored for a single gonotrophic cycle according to the previous experiment. Spermathecae were also dissected according to the previous experiment. After mating, males were stored in absolute ethanol and 24 individuals per treatment were measured for their *Wolbachia* density according to methods described previously [81,82].

### Stability of cytoplasmic incompatibility in triple-infected male *Ae. albopictus* mosquitoes

#### Data integration and normalization

Mosquito abundance and climate data were first aggregated to the weekly resolution to achieve temporal alignment. To align data spatially, the closest climate grid was matched to the latitude and longitude of recorded mosquito abundance data in Singapore and Europe. The matched data was further upscaled and analysed by residential townships of Singapore and provinces of European countries. Whereas regions of mainland China and the U.S. had climate data averaged at the province and state level respectively.

#### Model calibration and projections

We calibrated the database to the model in order to characterise historical life history traits across individual areas in each study region, and projected mosquito abundance according to climate change scenarios.

Model calibration was performed using STAN, using the Hamiltonian Monte Carlo (HMC) algorithm [83]. HMC was used as it can efficiently explore the parameter space especially in models with highly correlated parameters [84] by using the gradient of the log-posterior to move along trajectories that explore high-probability regions efficiently. By avoiding random-walk behaviour from other commonly used approaches (i.e random-walk Metropolis-Hastings), HMC avoids local maxima, enables samples to mix faster and be less correlated. It is therefore more suited to calibrate the high-dimensional entomological model as described below. For fitting parameters with finite support, we set boundaries to ensure that proposal values remained entomologically plausible and assigned uninformative priors using uniform distributions (Supplementary Information Table S1). We fixed the plausible upper bound of temperatures for every biological process based on literature to ensure that the inferred entomological processes were aligned with observed life-history traits for mosquito populations (Supplementary Information Table S2). We ran four independent HMC chains for 2,000 iterations in parallel, discarding the first 1,000 iterations as burn-in. We assessed convergence by computing the Gelman-Rubin (GR) statistic for accepted samples of each parameter and deemed that the algorithm has reached convergence if GR ≤ 1.1 for the respective parameters, and when the effective sample size (ESS) for the combined chains was sufficiently large. To obtain contemporaneous predictions of mosquito abundance, we randomly selected a sample of 1,000 accepted parameter draws to approximate the posterior distribution of the samples to simulate trajectories of mosquito abundance. (Supplementary Information Table S3–6).

Using climate data from ISIMIP3, we projected the mosquito abundance in each area by the respective measurement used in each area with the sampled parameters. We simulated the deployment of IIT in a weekly basis as the intervention scenario, at a release ratio of adult male *Wolbachia*-infected mosquitoes set to six times the historical mosquito abundance for that specific area. We denote this release ratio as the overflooding ratio. Both calibration and projection processes use climate data according to ISIMIP3 protocol, across timeframes and Shared Socioeconomic Pathways of SSP126, SSP370 and SSP585.

#### Reporting metrics

We reported simulation results using the following metrics to illustrate changes in mosquito abundance and the effectiveness of IIT interventions under different climate change scenarios across multiple timeframes. These metrics were calculated at the area level and then aggregated as averages within each study region. Mosquitoes usually reach peak abundance around the summer months in regions with temperate climates. This was taken as the 12 warmest consecutive weeks of the year. By identifying the contiguous period with the highest mean temperatures within each region, the definition captures the interval during which mosquito populations are most consistently exposed to prolonged and elevated heat. Therefore, we compared model-based projections of this heat period with the other weeks (normal) of the year.

1. **Historical mosquito abundance** (HA): HA quantifies the historical weekly mosquito abundance enabling comparison across sub-areas for a specific region. The average HA for a specific location between the start week of observation to the last week of 2024 was taken as the baseline scenario from which future projections in the absence of intervention are compared against. HA reflects temporal heterogeneity in mosquito density at that specific spatial resolution. However, it is not intended for cross-regional comparison, due to differences in surveillance protocols across study regions.
2. **Change in Mosquito abundance** (CA): CA is computed as the projected future female mosquito abundance divided by the historical average female mosquito abundance for a specific area. This quantity was aggregated to the areal or regional level. Multiple future timeframes were considered for future baseline projections. CA was further computed between periods of heat and normal temperatures to examine potential seasonal changes in the effectiveness of IIT under future climate change scenarios. High temperature conditions were defined as the 12 consecutive warmest weeks of each year; normal conditions included all other weeks.
3. **Intervention effectiveness** (IE): IE quantifies the percentage reduction in female mosquito abundance under the IIT interventions compared to the projected baseline scenario without IIT intervention. IE was calculated via comparison of baseline and intervention scenarios under specific future timeframes for a specific location, and is computed as the difference in mosquito abundances between these scenarios, divided by the baseline abundance. The mosquito abundance in the baseline scenario and intervention scenarios were respectively the projected mosquito abundance without and with IIT interventions implemented.

### Climatically forced process-based model

The model comprised uninfected adult female (*F*_*n*_), adult male (*M*_*n*_) and aquatic stages(*A*_*n*_). In the baseline case, only wildtype *Ae. aegypti* or *Ae. albopictus* population from each area were simulated. IIT programmes were incorporated through the release of *Wolbachia* infected male mosquitoes (*M*_*w*)_ to suppress wildtype mosquito populations (Figure 1). We assumed that IIT was conducted via release of *w*MelM or *w*AlbB transinfected male mosquitoes for modelling *Ae. aegypti* and via release of *w*Pip/*w*AlbA/*w*AlbB transinfected mosquitoes for modelling of *Ae. albopictus*. Both approaches result in unidirectional cytoplasmic incompatibility. Cytoplasmic incompatibility is induced by *Wolbachia*-infected male mosquitoes mating with wildtype females, which creates unviable offspring with probability *f*_*c*_. The model characterised important, climatically-forced entomological processes, including aquatic (*f*_*μ*_,*a*) and adult stage (*f*_*μ,m*_,*f*_*μ,f*_), mortality, mosquito reproduction (*f*_*ρ*_) and development (*f*_*τ*_) as separate functions (Figure 1, Equation 1–4). The temperature dependence of these processes was characterised using a trapezoidal function, defined by rising and falling logistic functions at either lateral side. Each logistic function was parameterized by temperature-dependent inverse exponential functions which are described in detail below (Equation 5–9). Their parameters calibrated to historical mosquito abundance and climate data using the HMC approach as described above (Table 1). The aquatic removal rate was determined via the baseline aquatic mortality rate (*f*_*μ,a*_) and the aquatic stage environmental carrying capacity function (*f*_*K*_), which was characterised via the constant parameter ω which represents site-specific geographical features such as larval habitat availability, and also a rate parameter λ to reflect precipitation-induced changes to the carrying capacity (Equation 10).

**Figure 1.**
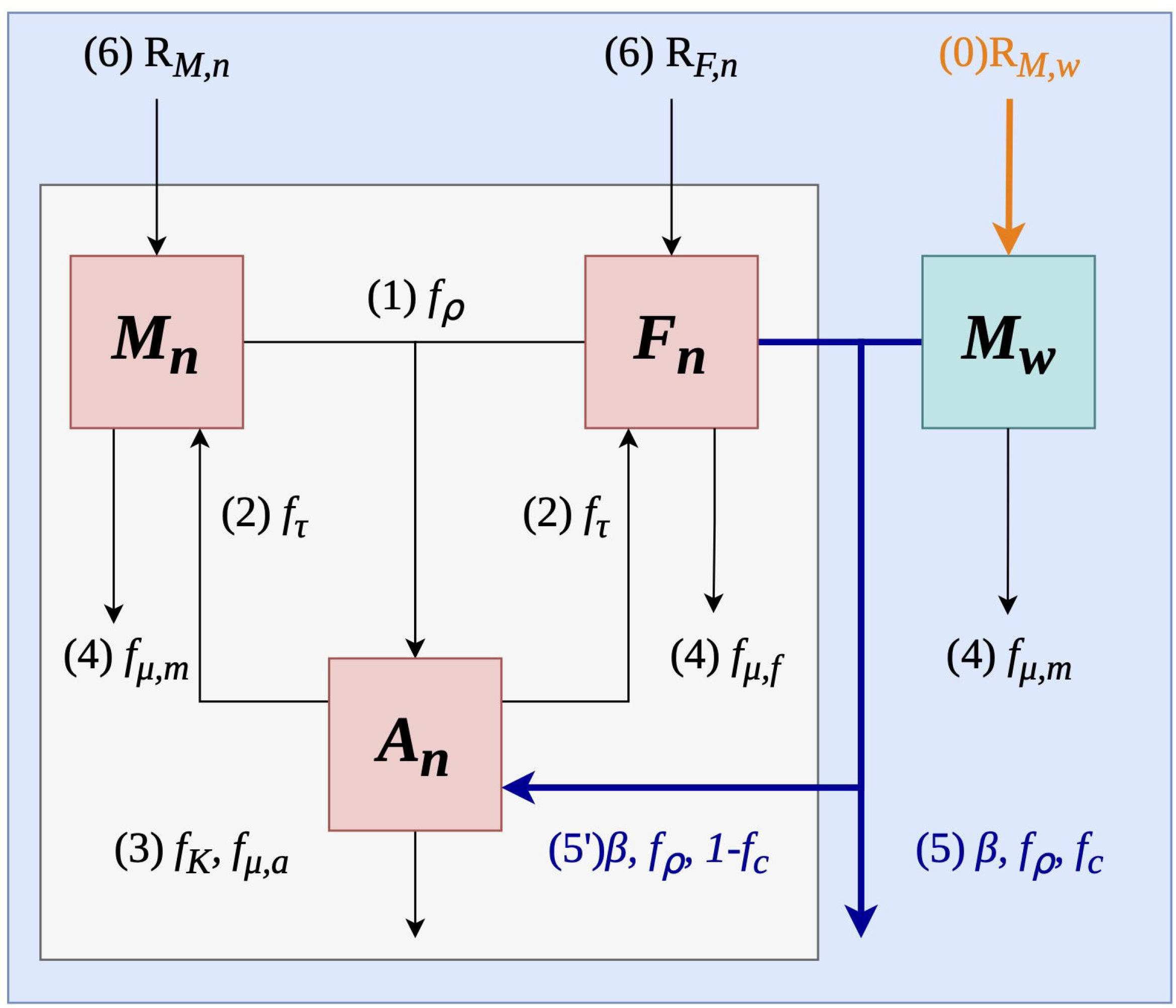
Diagram of climatically driven entomological model. The model comprised two scenarios: baseline mosquito abundance (grey panel) and mosquito abundance under IIT implementation (blue panel). Wildtype adult female, wildtype adult male, *Wolbachia* infected adult male, and wildtype aquatic stage mosquitoes were denoted as *F*_*n*_,*M*_*n*_, *M*_*w*_ and *A*_*n*_ respectively. Entomological functions illustrated the change of mosquito life history traits and can change with climate. The following processes characterized the transitions between mosquito compartments, namely: (0) release of *Wolbachia*-infected male mosquitoes into the environment (1) wildtype females mate with wildtype male mosquitoes and produce wildtype offspring in the aquatic stage, governed by the reproduction function *f*_*ρ*_ . (2) Wildtype offspring mature and emerge as female and male wildtype mosquitoes governed by the development function *f*._*τ*_ Female and male mosquitoes were assumed to emerge at the same ratio. (3) Wildtype aquatic mosquito mortality was characterized by *f*_*μ*_,_*a*_ and the environmental carrying capacity function *f*_*K*_. (4) Adult mosquito mortality was characterized by *f*_*μ*_,_*m*_ and *f*_*μ*_,_*f*_ . (5) IIT intervention was characterized via wildtype females mating (*f*_*ρ*_) with *Wolbachia* infected male mosquitoes and producing eggs that fail to hatch due to cytoplasmic incompatibility (*f*_*c*_). (5’) Imperfect cytoplasmic incompatibility is modelled as (1-*f*_*c*_) (6) Immigration of wild-type mosquitoes into the modelled location (*R*_*M,n*_, *R*_*F,n*_)is governed by the rate of migration *α*, and the current population size.

We employed several climatically-forced process based functions to describe the transition between different compartments (Equation 1–4). For aquatic stage mosquitoes, 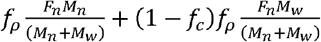 characterised the number of new aquatic stage mosquitoes per week. Via the temperature dependent reproduction rate *f*_*ρ*_, wildtype female mate with wildtype male with probability 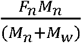 or they mate with *Wolbachia* infected male with probability 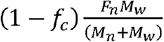, and produce eggs that hatch. The aquatic mosquitoes leave their compartment after maturation (f_*τ*_) or mortality (*f*_*μ,a*_) . Mortality was scaled via 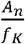, the ratio of current wildtype aquatic stage mosquitoes over the current environmental carrying capacity. We constrained the number of aquatic stage mosquitoes to remain below the environmental capacity at each timepoint via *A*_*n*_ ≤ *f*_*K*_, and restricted the maximum number of aquatic mosquitoes leaving their compartment to be equal or below its total population size via 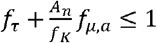:

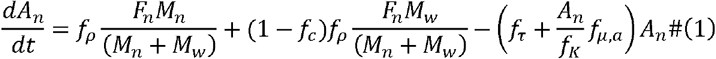

We simulated the deployment of IIT via release of *Wolbachia*-infected male mosquitoes (*R*_*M,w,i*_) from 2025 to 2100 at 6 times the local historical female mosquito abundance. 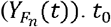 and *t*_1_ denote the start and end of the historically observed period of data:

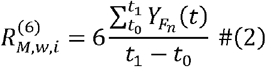

Several alternative deployment strategies were also simulated to reflect different potential implementations strategies for IIT. Namely:

1. Constant release with different overflooding ratios and release intervals: the overflooding ratio (*n*) and release interval (*m*) were varied from 1 to 10. The release volumes were set at times the local historical female mosquito abundance and the *Wolbachia*-infected males were released at per *m* week intervals, from 2025 to 2100.

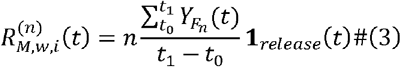

where **1**_*release*_ (*t*) = 1 for a release week and **1**_*release*_ (*t*) = 0 otherwise
2. Releases depended on the level of suppression achieved, with release volumes adjusted when the wildtype female mosquito populations reach 50% and 25% of the baseline projection. We assumed that overflooding ratios are reduced from 6 to 3 and 3 to 1 when wildtype populations reached 50% and 25% respectively. These epochs span *t*_2_ to *t*_3_ and *t*_3_ to *t*_4_.

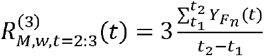, *t* ∈ {*t*_2_ : *t*_3_ }, 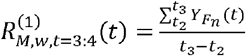, {*t*_3_ : *t*_4_ } characterised the migration of adult mosquitoes between neighbouring locales via a migration term. The migration term depend on some migration rate and the wildtype mosquito abundance 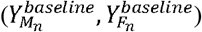 in the previous week (*t* −1)in the absence of intervention. The baseline scenario was used to represent mosquito influx from neighbouring regions without intervention.

To prevent unrealistic projections of mosquito extinction, the migration rate α was constrained to be positive.

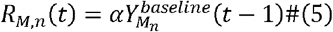

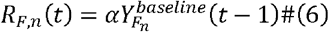

Adult male and female wildtype mosquitoes (*M*_*n*_, *F*_*n*_) emerge at the rate of 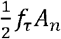 and have a mortality rate of *f*_*μ,m*_, and *f*_*μ,f*_ respectively. We assumed that *Wolbachia*-infected male mosquitoes (*M*_*w*_) have similar life-history traits as wildtype male mosquitoes and are removed at a mortality rate of, *f*_*μ,m*_.

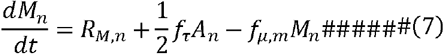

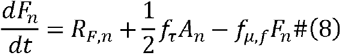

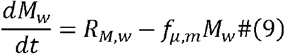

Mosquito development (*τ*), reproduction (*ρ*) and mortality (*μ*) were characterised via trapezoidal functions updated according to temperature *T* in week *t* (Equation 5–9). These functions (*η ϵ*(*τ, ρ, μ*)) comprised two logistic functions parameterised by a pair of midpoint temperatures *T* _1,*η*_, *T*_2,*η*_ taken from literature, rate parameters *k* _1,*η*_, *k*_2,*η*_, and one shared unknown scale parameter *L*_*η*_ (see Supplementary Information Table 1 and Table 2). The aquatic (*µ*_*a*_), adult female (*µ*_*f*_) and adult male (*µ*_*m*_) mortality were characterised as 1 minus the trapezoidal function, as these rates are inversely related to the optimal temperature range. We characterised sex-specific differences in adult stage mortality via the parameter *δ*. Unknown parameters (*T*_2,*η*_, *k* _1,*η*_, *k* _2,*η*_, *L*_*η*_) were calibrated to observed data using the HMC approach described above.

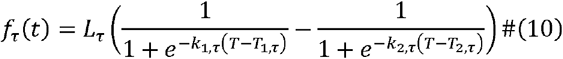

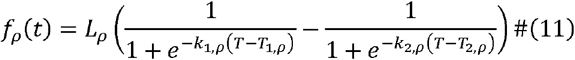

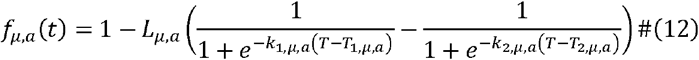

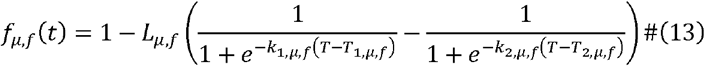

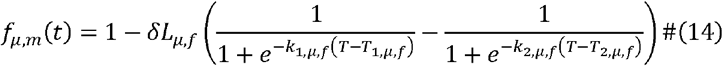

The environmental capacity function *f*_*K*_ is taken to be linearly related to daily mean precipitation (*P*)at week *t*, via the rate parameter *λ* and varies with a constant parameter ω:

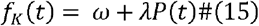

While laboratory evidence strongly suggests complete cytoplasmic incompatibility induced by both *w*AlbB and *w*MelM for *Ae. Aegypti* and *w*Pip/*w*AlbA/*w*AlbB for *Ae. albopictus* up to the lethal temperatures for mosquitoes, we conservatively modelled CI stochastically via a Beta distribution (*f*_*c*_) parameterised from observed egg hatch proportion under both acute and chronic heat stress conditions (Figure 4, X). Probability that CI is successfully induced at some time step was modelled as a stochastic draw from a Beta distribution,

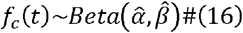

Let the observed egg hatch proportions under *Wolbachia*-infected crosses be denoted as *h*_*i*_ ∈ [0,1] . We define the probability that CI is successfully induced as:

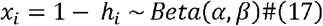

with the sample mean and variance being:

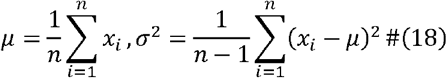

*α, β* are estimated via the method of moments:

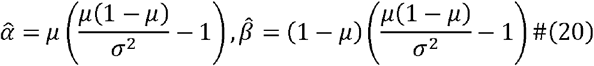

### Observation process

Observed wildtype weekly mosquito abundance was denoted *Z*_*S*_(*t*), where *S* ∈ (*A, F*_*n*_) for wildtype aquatic stage mosquito (*Z*_*A*_) and female adult mosquito 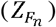 respectively. We used a normal distribution to characterize the observation process and modelled the wildtype weekly mosquito abundance *Z*_*S*_ *(t)* as:

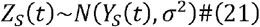

with the mean corresponding to the simulated time series *Y*_*S*_(*t*) generated by the process-based model and the standard deviation σ characterising the spread in the observation process., Increments from *t* to *t* + 1 comprised the simulated values:

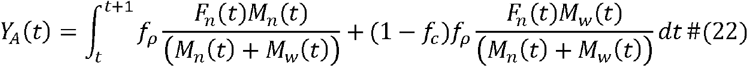

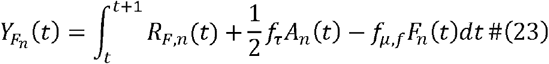

The likelihood was given by the product sum of observation (*Z*_*S*_), given the modelled trajectories (*Y*_*S*_) produced by the parameter set Θ from the first till last recorded week of observed data *t* = 0:*T*.

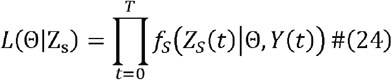

### Robustness checks

We examined whether the model adequately characterized mosquito abundance by examining the differences in mosquito abundance generated by the calibrated models to the historically recorded mosquito abundance per area. To further test the predictive ability of the model, we employed expanding window cross-validation and report the root square mean percentage error. Model validation was performed by splitting the timeseries data into the first 70% of data as the train set and the last 30% of data as the testing set. Model projection accuracy was reported with mean square error, mean absolute error and mean percentage error (Supplementary Information, Figure S1–S10).

Sensitivity analysis were conducted to examine the changes in mosquito abundance and intervention effectiveness when parameters were deviated from calibrated values or when IIT implement programme changed. To account for climate model variability, we included other four climate models, namely, IPSL-CM6A-LR, MPI-ESM1-2-HR, MRI-ESM2-0, and UKESM1-0-LL. To examine the impact of having entomological functions which were different from estimated values, we separately varied the climate dependent entomological functions (*f*_*μ,a*_, *f*_*μ,f*_, *f*_*μ,m*_, *f*_*ρ*_, and *f*_*K*_) and migration rate (*α*) by increasing or decreasing the values by 5% while keeping all other parameters constant. To approximate the complexity of real-world implementation and policy variation, we simulated different IIT programmes to evaluate their impact on IE. Mosquito abundance under baseline and intervention scenarios was subsequently reprojected forward in time to calculate CA and IE, which were analysed at the subarea level for each region using primary climate model GFDL-ESM4 (Supplementary Information, Figure S31–49).

All computations were performed in R version 4.5.1 on a Mac Studio equipped with an M2 pro chip. Analysis was supported by package rstan (version 2.32.7). Visualization of results relied on the following tools: ggplot2 (version 3.5.2), rnaturalearth (version 1.0.1), ggmapcn (version 0.1.2), onemapsgapi (version 2.00), Singapore planning area boundary dataset [85]

## Results

### Historical abundance across regions

We fitted our model to observed abundance data in each region and reconstructed the historical mosquito abundance (HA) over the full period for which data were available. Figure 2 showed that HA varied substantially across regions from the beginning of the observation period until the last week before 2025, under the SSP580 scenario with the GFDL-ESM4 climate model. In mainland China (Figure 2A), Hainan Province exhibited the highest HA, whereas Liaoning Province showed the lowest. In Europe (Figure 2B), the Emilia-Romagna region had the highest HA compared to the Puglia region, which had the lowest HA. In the US (Figure 2C), North Carolina exhibited the highest HA compared to the lowest in California. In Singapore (Figure 2D), Jurong East had the highest HA, whereas Sungei Kadut had the lowest HA.

**Figure 2.**
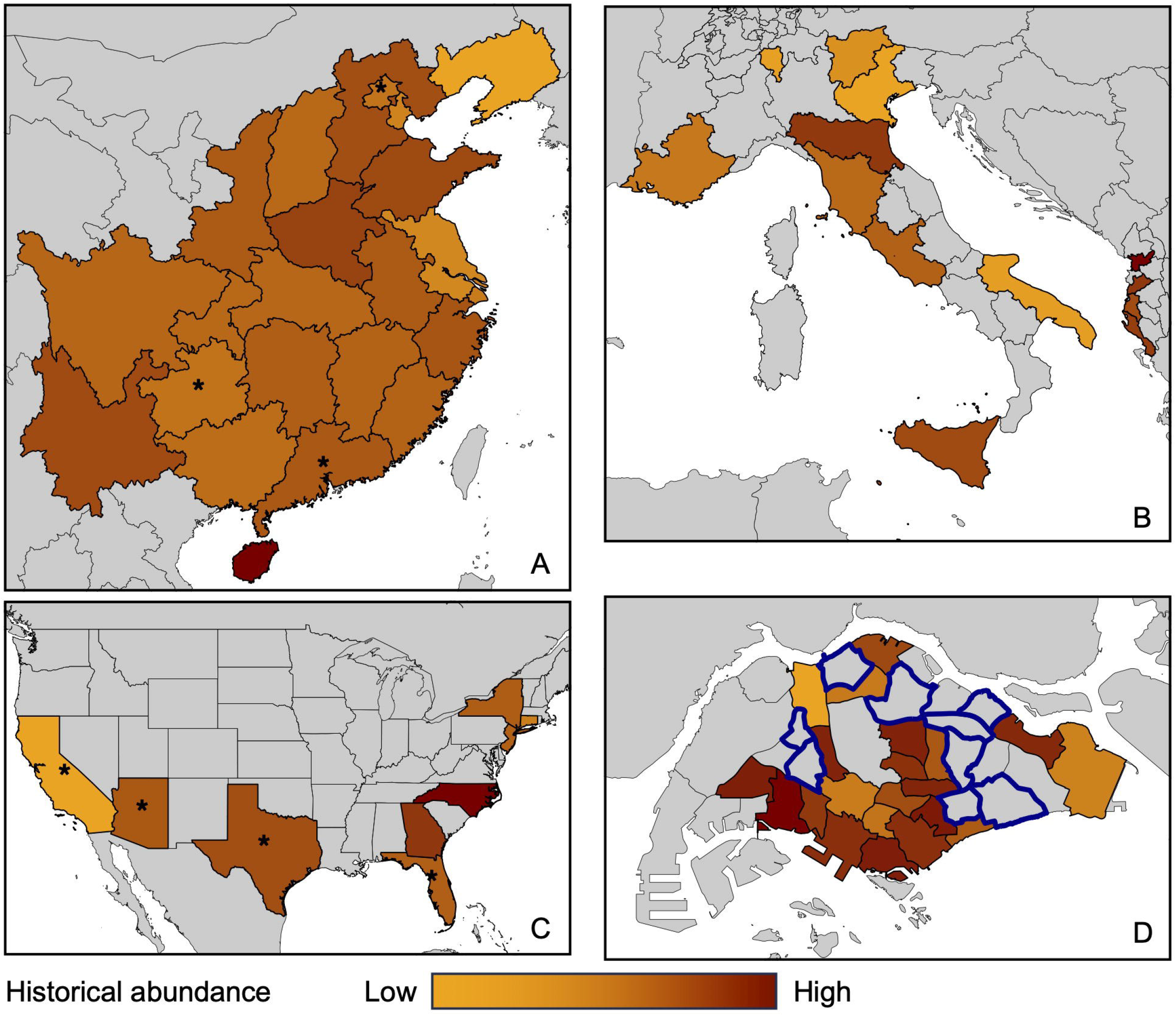
Historical mosquito abundance by region in relative density. Historical mosquito abundance (HA) is shown for four study regions: mainland China (A), Europe (B), the United States (C), and Singapore (D). HA is the weekly average female mosquito abundance in the historical period as projected from the location-specific process-based model calibrated to abundance in that location. Due to differences in entomological surveillance systems and measured indices (Table 1), the abundance values are presented in log scale and are not directly comparable across regions. The stars were annotated for the following items: in mainland China (A), mosquito density was measured using the Mosquito Ovitrap Index (MOI), whereas the other regions annotated in stars used the Breteau Index (BI). In the U.S. (C), the primary vector species analysed was *Ae. albopictus*, while other regions annotated in stars focused on *Ae. aegypti*. In Singapore (D), areas with ongoing IIT-SIT deployment shaded in blue were excluded in the study, as historical data does not reflect natural *Aedes* populations. Regions which were not analysed are denoted in grey due to a lack of data.

### Changes in mosquito abundance in future climate change scenarios

We projected changes in mosquito abundance (CA) to quantify increases in mosquito abundance under future climates compared to historically observed abundance (Figure 3). Overall, projected mosquito abundance exhibited substantial spatial heterogeneity across regions and time periods. The magnitude and direction of change varied considerably among geographic settings. In mainland China (Figure 3, A1–A3), projected changes were generally moderate and spatially heterogeneous. In 2025–2050, several provinces in central and eastern China exhibited abundance levels comparable to or slightly below the historical baseline, whereas Fujian province showed mild increases (1–2×). By 2050–2075 and 2075–2100, we projected elevated abundance in a broad number of provinces, particularly across the northern and southwestern subtropical regions, where increases of 1–2× became more prevalent. In contrast, several provinces in central and eastern China continued to exhibit stable or reduced abundance relative to the historical period.

**Figure 3.**
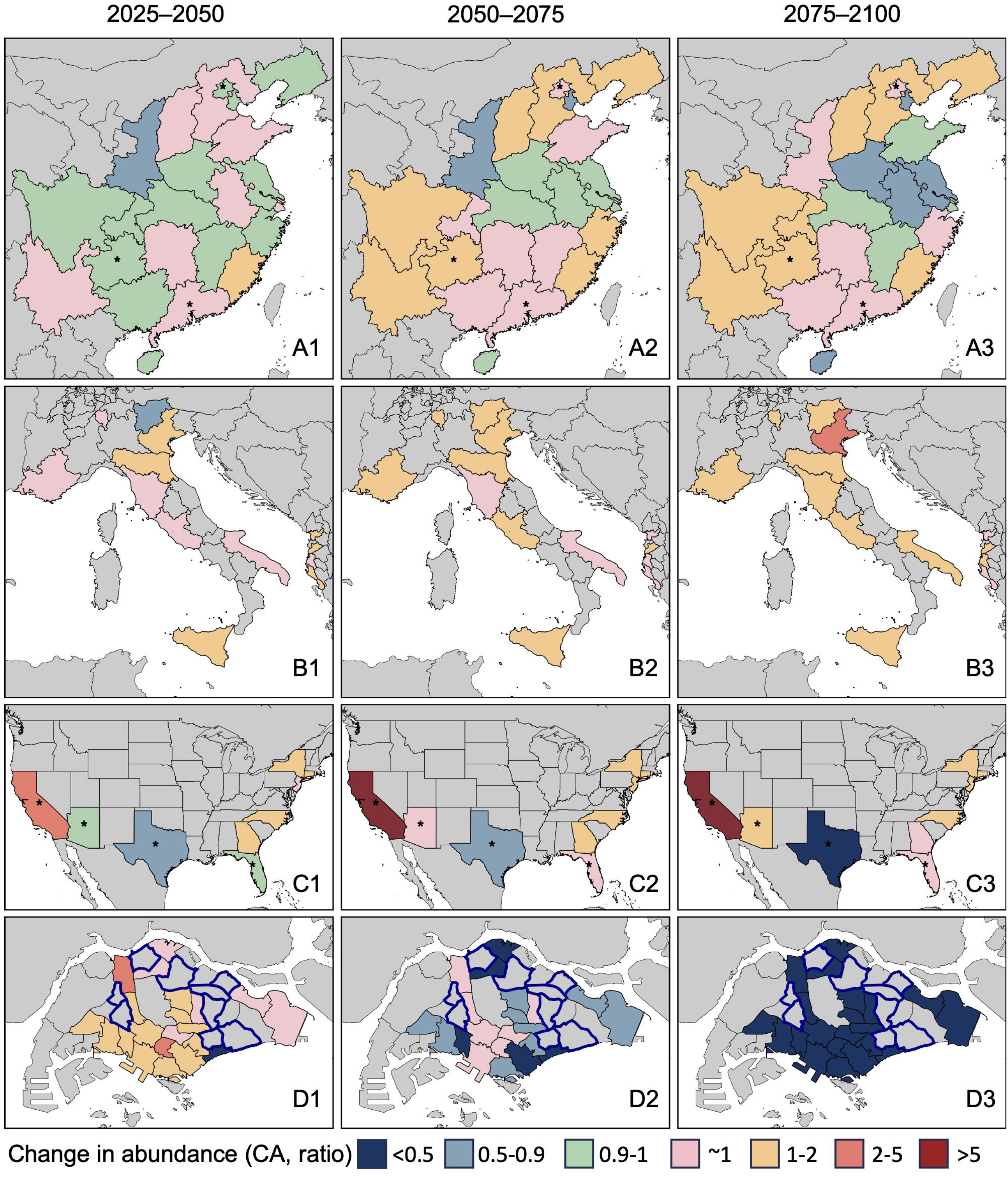
Projected change in mosquito abundance under future climate conditions across different timeframes. Maps show the projected change in mosquito abundance (CA) for each study region across three future time periods: 2025–2050 (column 1), 2050–2075 (column 2) and 2075–2100 (column 3). Projections are based on simulations using the GFDL-ESM4 climate model under SSP585. CA values represent the change in female mosquito abundance during each future period compared to a historical baseline (until 2025). Panels show regional projections for mainland China (A), Europe (B), the United States (C), and Singapore (D). Categorical colour scales represented in blue indicate a decrease in mean CA at 0.5–0.9 and in green at 0.9–1. Pink indicated no significant differences in projected abundance compared to the historical period with 95% credible intervals covering 1. Yellow (1–2, mild), red (2–5, moderate), and dark red (>5, severe) colours represent increasing levels of projected mean CA versus the historical period. The stars are annotated for the following items: in mainland China (A), mosquito density was measured using the Mosquito Ovitrap Index (MOI), whereas in the other regions they indicate use of the Breteau Index (BI). In the U.S. (C), the primary vector species analysed was *Ae. albopictus*, while other annotated regions focused on *Ae. aegypti*. In Singapore (D), areas with ongoing IIT-SIT deployment are excluded in the study and denoted in blue outlines. Regions which were not analysed are denoted in grey due to a lack of data.

In Europe (Figure 3, B1–B3), projected mosquito abundance generally increased over time. During 2025–2050, most regions showed abundance levels close to historical conditions or modest increases. By 2050–2075, mild increases (1–2×) became more widespread across southern Europe. In 2075–2100, several regions exhibited stronger increases, including province of Veneto with projected abundance exceeding 2–5× historical levels, indicating an intensification of mosquito suitability in parts of continental Europe.

In the United States (Figure 3, C1–C3), we projected heterogenous changes in abundance. California consistently showed the largest increases in mosquito abundance across all future periods, with projected abundance exceeding fivefold above the historical baseline by mid- and late century. In contrast, Texas is projected to have lowered abundance relative to historical levels up to the end of the century. Several eastern and north-eastern coastal states showed modest increases (1–2×), whereas Florida and parts of the south-eastern United States are projected to have relatively stable mosquito abundance.

In equatorial Singapore (Figure 3, D1–D3), projected changes displayed strong intra-urban heterogeneity and a marked temporal transition. During 2025–2050, mosquito abundance varied substantially across neighbourhoods, ranging from decreases relative to historical levels to localized moderate increases (2–5×). However, projections for 2050–2075 indicated a broad shift toward lower abundance across much of the island, with many areas exhibiting reductions below historical levels. By 2075–2100, decreases became more widespread and pronounced, with all areas were projected to experience reductions in abundance to less than half of historical levels. These results suggest that future climate-driven mosquito dynamics in Singapore may differ substantially from those projected in temperate regions, with potential declines emerging under environmental conditions later in the century.

### Stability of cytoplasmic incompatibility in *w*MelM- and *w*AlbB-infected male *Ae. aegypti* mosquitoes

We performed experiments to support our assumption that CI induced by *Wolbachia* would be robust to future climate scenarios. Both *w*MelM and *w*AlbB induced near-complete cytoplasmic incompatibility in all control incompatible crosses between *Wolbachia*-infected males and uninfected females (Figure 4). Egg hatch proportions in crosses between uninfected females and uninfected males were high in all crosses and differed significantly from each of the incompatible crosses (Wilcoxon rank sum test, all P < 0.027). The high egg hatch proportions in compatible crosses exposed to chronic heat suggest that fertility was not substantially affected by high temperatures during mating and oviposition. CI remained near-complete for both *Wolbachia* strains following acute and chronic heat exposure in adults (Figure 4A), even when females went through a second gonotrophic cycle (Figure 4B) and when males were remated following sperm depletion (Figure 4C). Some incompatible crosses for both *w*MelM and *w*AlbB in this experiment produced a very small number of viable offspring (1-3 eggs hatched) but there was no relationship with temperature treatment. CI was also complete when *Wolbachia*-infected males were exposed to high concentrations of tetracycline for one week prior to mating (Figure 4D). This concentration reduced the median density of *w*MelM by 15-fold (Figure 4E). While there was only a 2.6-fold reduction in *w*AlbB density in positive males, 33% of treated males tested negative for *Wolbachia*, indicative of complete loss (Figure 4E). Overall, the results suggest that CI induced by mass-reared *Wolbachia*-infected males is likely to remain robust even under extreme temperatures (1-2°C below mosquito lethal limits, assuming immature development at 29°C. Furthermore, CI remains robust even when *Wolbachia* strains are suppressed to artificially low levels through antibiotic treatment at the adult stage. Subsets of dissected females from incompatible crosses all showed evidence of insemination, suggesting that maintenance of complete CI is not due to a lack of mating.

**Figure 4.**
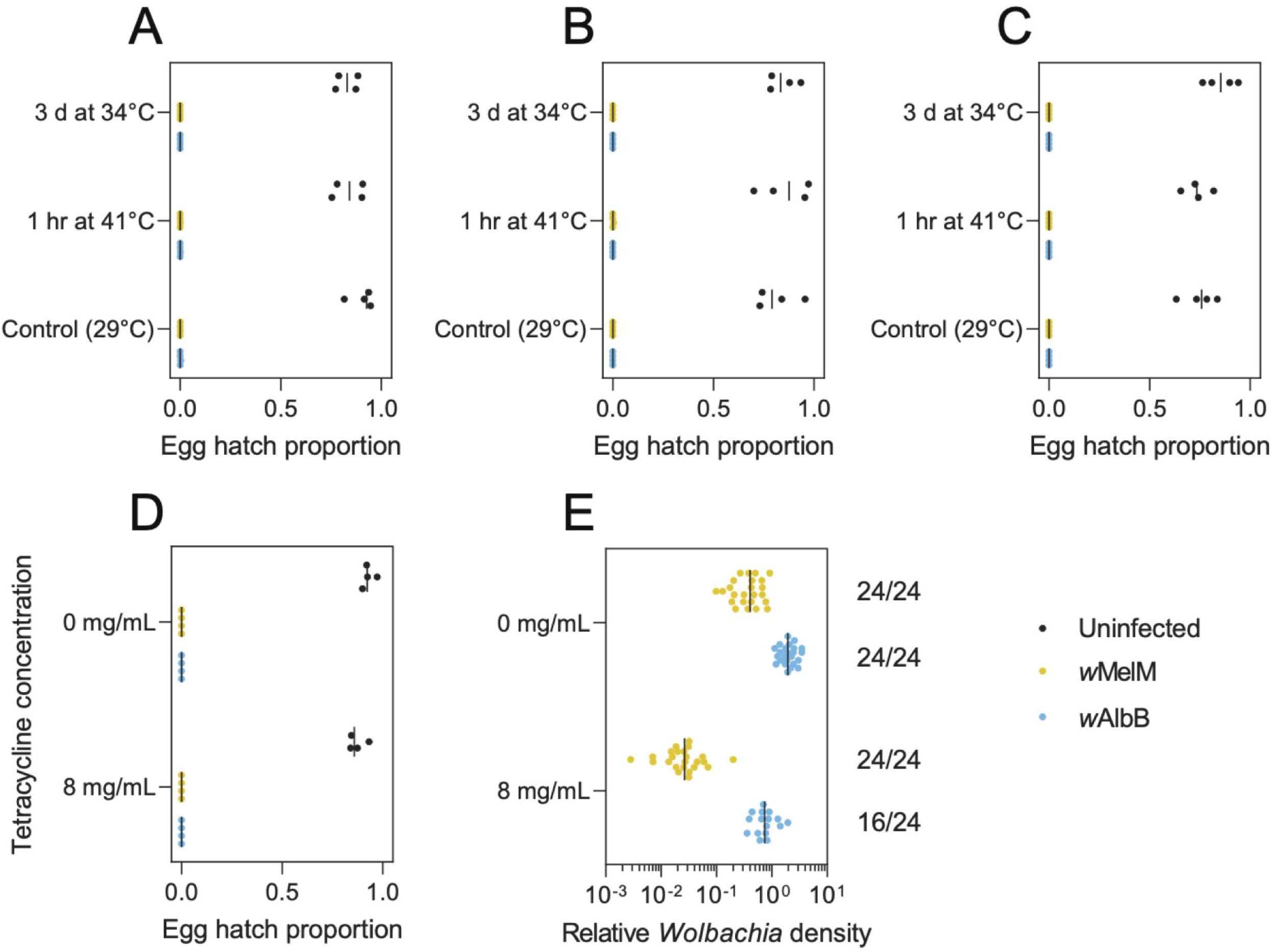
*Wolbachia*-induced cytoplasmic incompatibility remains robust to extreme temperatures and suppression through antibiotic treatment. (A-D) Egg hatch proportions from compatible crosses between uninfected females and uninfected males, or incompatible crosses involving *w*MelM or *w*AlbB males and uninfected females. (A-C) Adult males were either kept under control conditions (29°C), exposed to a 1 hr 41°C heat shock before mating, or maintained at 34°C for 3 d before mating. Hatch proportions were measured over the (A) first and (B) second gonotrophic cycles. (C) Males were then remated to uninfected females at a 10:50 male:female ratio. (D) Adult males were exposed to 0 or 8 mg/mL tetracycline for one week before mating. (E) *Wolbachia* density in adult males exposed to 0 or 8 mg/mL tetracycline for one week. Numbers indicate the number of males testing positive for *Wolbachia* relative to the total tested. Each dot represents one replicate cage (A-D) or individual mosquito (E) while lines show medians.

### Stability of cytoplasmic incompatibility in *XX*-infected male *Ae. albopictus* mosquitoes

#### Simulating future effectiveness of IIT under future climate change scenarios

We simulated IIT deployment through the release of *Wolbachia*-infected male mosquitoes in each area from 2025 to 2100. Intervention effectiveness (IE) was computed to evaluate the robustness of IIT across studied regions under each future climate change scenarios (Figure 5) by comparing the mosquito abundance between the intervention and the baseline case with no interventions. Across all areas, substantial mosquito suppression was achieved due to interactions between wildtype female mosquito populations and released *Wolbachia*-infected male mosquito populations (see Supplementary Information). Taking the GFDL-ESM4 model under the SSP585 scenario as an example, IE sustained at a high level across all regions and future time periods.

**Figure 5.**
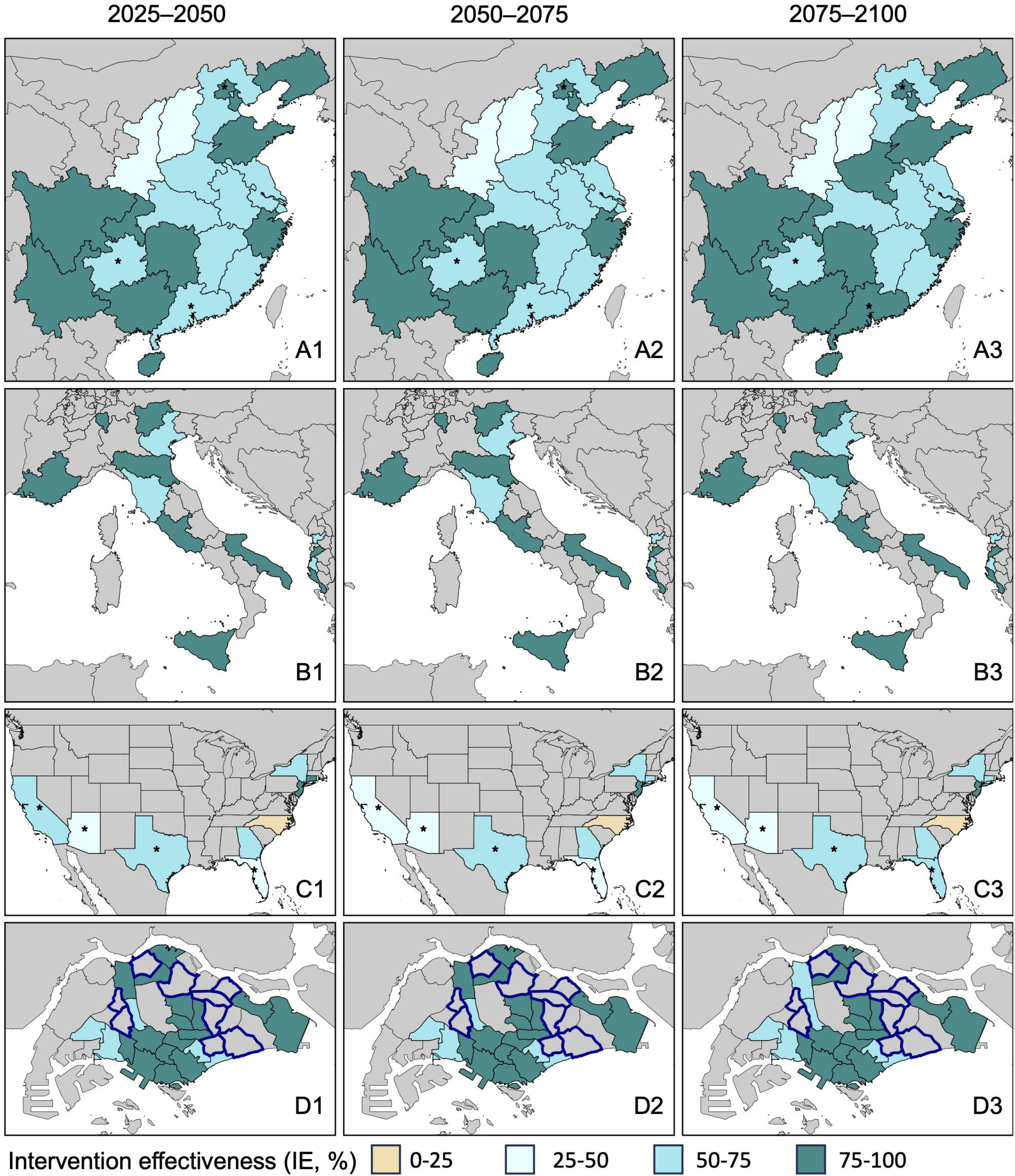
Projected intervention effectiveness of IIT under future climate conditions by different timeframes. Maps show the projected intervention effectiveness (IE) for each study region across three future time periods: 2025–2050 (column 1), 2050–2075 (column 2) and 2075–2100 (column 3). IE defined as the percentage reduction in female mosquito abundance in the intervention scenario compared to a baseline scenario without intervention, under future climate projections. Simulations assume a 6:1 overflooding ratio (OFR) and are driven by the GFDL-ESM4 climate model under SSP580. Map was coloured to show the IE levels in yellow (0–25%), light blue (25–50%), blue (50–75%) and green (75–100%). Panels show regional projections for mainland China (A), Europe (B), the United States (C), and Singapore (D). The stars were annotated for the following items: In mainland China (A), mosquito density was measured using the Mosquito Ovitrap Index (MOI), whereas the other regions annotated in stars used the Breteau Index (BI). In the U.S. (C), the primary vector species analysed was *Ae. albopictus*, while other regions annotated in stars focused on *Ae. aegypti*. In Singapore (D), the areas with ongoing IIT-SIT deployment were excluded in the study, as data do not reflect natural *Aedes* population dynamics.

#### Intervention effectiveness of IIT across different SSPs and the impact of seasonality

We examined IIT effectiveness across different SSPs and climatic conditions (Figure 6). At baseline without intervention, we projected increases in abundance across mainland China, Europe and the United States under all SSPs and future periods, particularly in periods of hotter temperatures. Abundance is projected to increase with worsening SSPs across mainland China, Europe and the United States, with stronger effects observed during heat conditions relative to normal conditions. However, Singapore is projected to have declines in mosquito abundance in SSP 370 and SSP585 (Figure 6, A4–B4).

**Figure 6.**
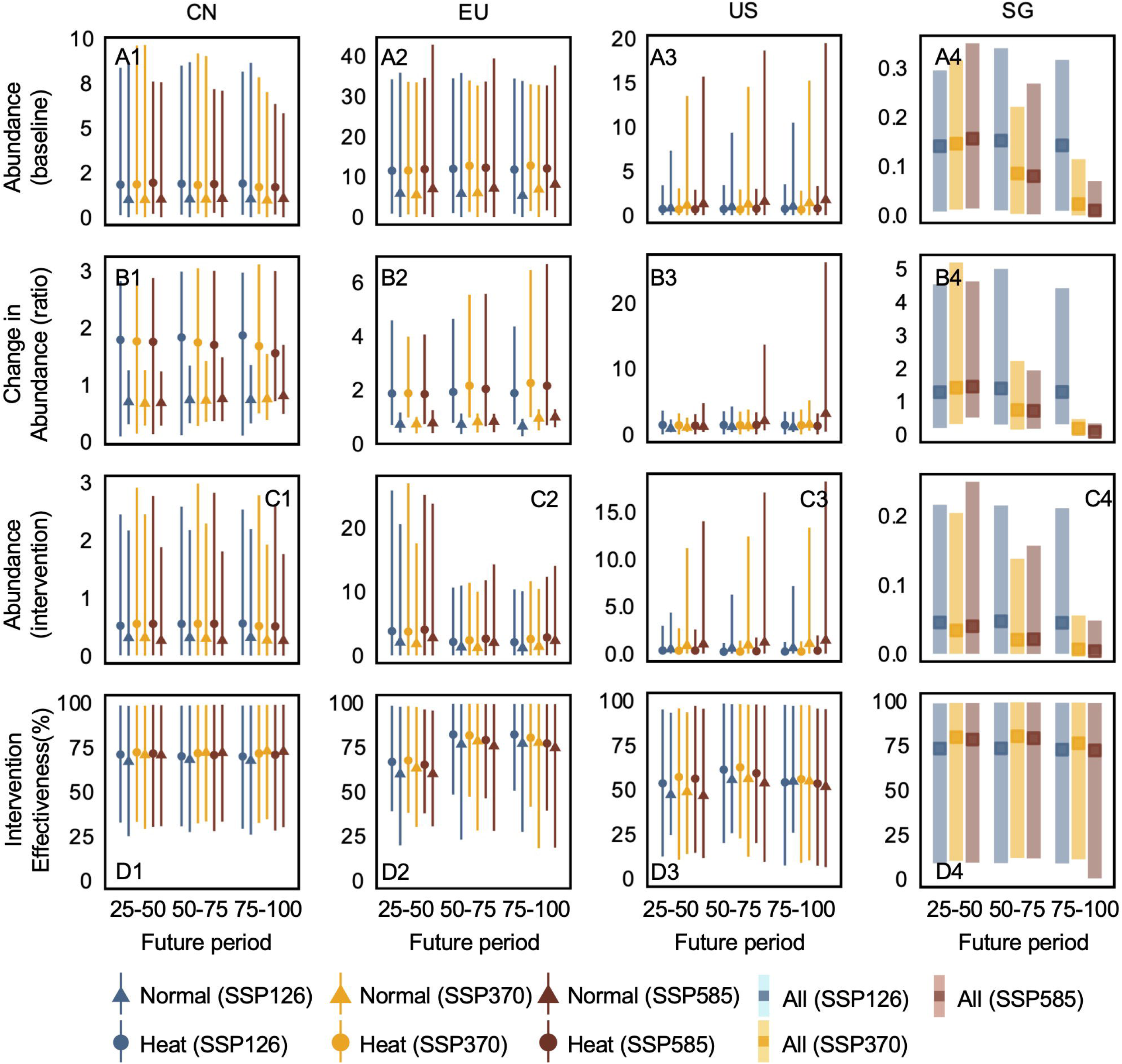
Abundance projections with corresponding intervention effectiveness (IE) under different future climate scenarios. The series of box plots summarize the projected abundance in baseline scenario (A1– 4), change in abundance (B1–4), abundance in intervention scenario (C1–4), and intervention effectiveness (D1–4) for each for mainland China (CN), Europe (EU), the United States (US), and Singapore (SG) across three future time periods: 2025–2050, 2050–2075 and 2075–2100. The change in abundance was taken as the projected future mosquito abundance divided by historical mosquito abundance. Intervention effectiveness was defined as the percentage reduction in female mosquito abundance achieved by implementing IIT interventions relative to the baseline scenario of no IIT interventions. Simulations assumed a 6:1 overflooding ratio. The figure visualizes projections under the GFDL-ESM4 model. Each panel visualizes seasonal changes in high temperatures and normal conditions for EU, US and CN, or for the whole year for SG. High temperature conditions were defined as the 12 consecutive warmest weeks of each year, while normal conditions included all other weeks. 95% credible intervals are shown as error bars. The colour scheme included SSP126 in blue, SSP370 in yellow, and the SPP585 in red to compare projections across different emission scenarios.

We simulated the deployment of IIT programmes in the future period spanning 2025 to 2100. A sustained decline in mosquito abundance was observed across all regions from 2025–2050 onwards (Figure 6, C1–4). We computed intervention effectiveness (Figure 6, D1–4) to quantify the relative effects of IIT compared to the baseline scenario. IE increased rapidly to stable levels from the initial deployment period (2025–2050) and remained consistently high through the end of the century across all regions and SSPs. IE varied only modestly between heat and normal conditions across all future periods and SSPs.

#### Impact of overflooding ratio, release intervals and release strategies on intervention effectiveness

We varied the overflooding ratio and release interval to simulate potentially different IIT release programmes. These simulations were employed across all study regions over all available climate change scenarios to test future IE across 2025 to 2100 (Figure 7). Frequent release schedules were important for maintaining IIT effectiveness, whereas higher overflooding ratios became increasingly important for achieving high IE under severe climate change scenarios in temperate study regions (Figure 7, A–C). Reduced IIT release volumes in mainland China, Europe, and the United States resulted in wider and higher credible intervals for projected mosquito abundance. Reductions in IIT release volumes produced greater heterogeneity in IE between hotter and normal temperature conditions in Europe compared with mainland China and the United States (Supplementary Information, Figure S47–S48). In Singapore, IIT remained robust across most constant-release policies (Figure 7, D1–D3), therefore, we additionally explored suppression-based adaptive IIT release strategies. IE increased rapidly in the near future (2025–2050) and remained stable throughout the century in most areas. However, in some areas, IE was attenuated by climate change and mosquito migration (Figure Supplementary Information, S49). These simulations highlight that intervention outcomes may vary according to the volume and timing of *Wolbachia*-infected male mosquitoes releases. Implementing a constant-release IIT programmes at broad spatial scales may achieve greater effectiveness in temperate regions, whereas releases adapted to observed mosquito abundance could potentially maintain intervention effectiveness while balancing release volumes.

**Figure 7.**
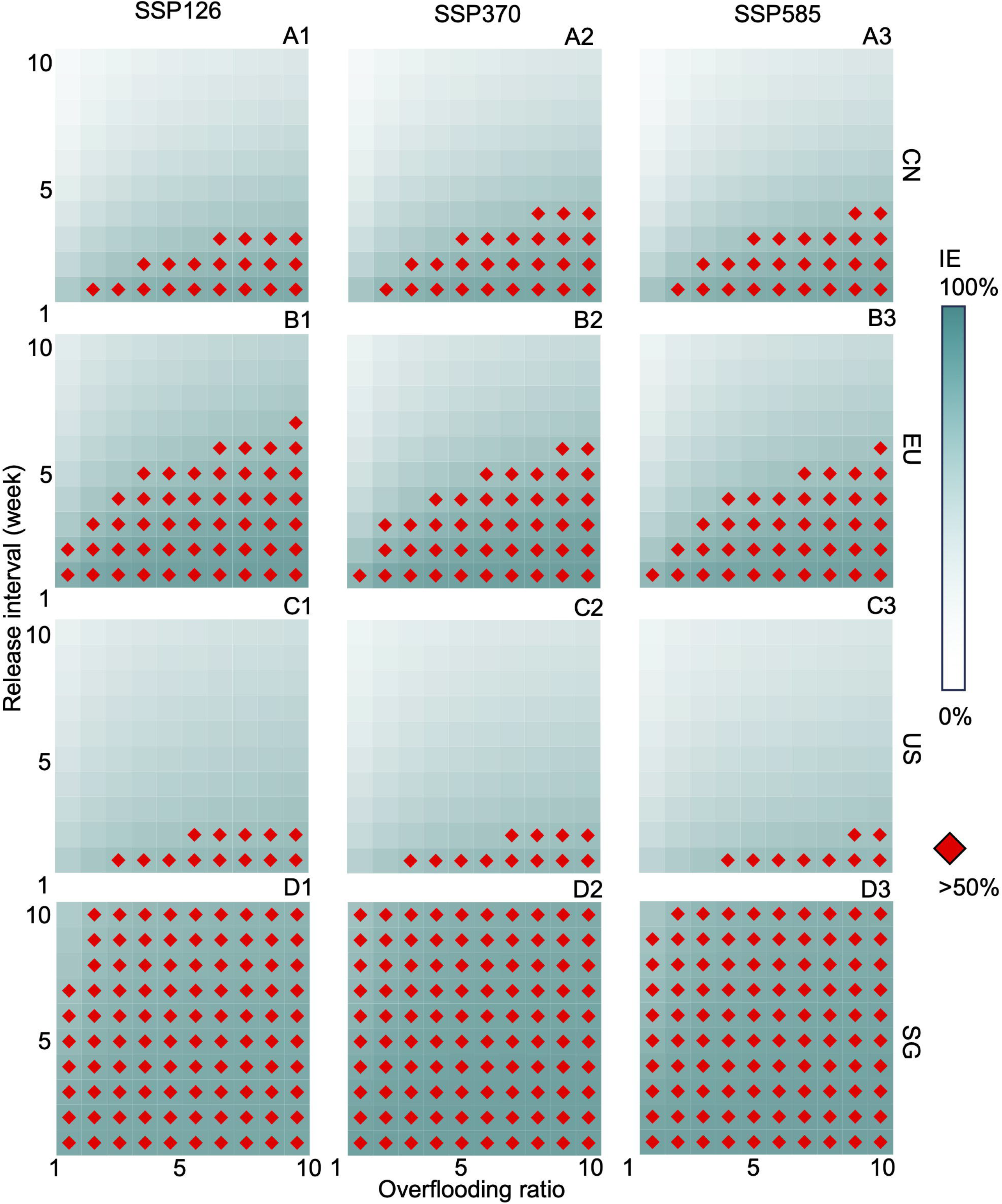
Intervention effectiveness projections by different release programmes. Projected intervention effectiveness in mainland China (A1–3), Europe (B1–3), United States (C1–3), and Singapore (D1–3) across three climate scenarios: SSP126 (column 1), SSP370 (column 2), SSP585 (column 3). The intervention effectiveness was defined as the percentage reduction in mosquito abundance achieved by implementing IIT interventions relative to the baseline scenario of no intervention. Simulations assumed varying overflooding ratios and release intervals. The figure visualizes projections under the GFDL-ESM4 model, with each compartment visualizing the IE from 2025 till 2100 at the regional level by implementing a constant release IIT programme with fixed release intervals or overflooding ratios. Darker shades represent higher IEs, and IE over 50% were denoted with red diamonds.

#### Robustness checks

We conducted expanding-window cross-validation and a comprehensive set of sensitivity analyses to assess the robustness of model projections. Validation results indicated that the model captures local seasonality, magnitude, and long-term trends in mosquito abundance well with low error, although interannual fluctuations were less well captured in equatorial climatic settings (Supplementary Information, Figures S1– S10). Using alternative climate models resulted in variation in projected mosquito abundance. In particular, some climate models projected warmer future temperatures that induced natural declines in baseline mosquito populations. Under these conditions, intervention effectiveness may appear low because projected mosquito abundance under intervention and no-intervention scenarios become close in value. Yet, in these cases, the underlying intervention is expected to remain effective, with IIT-based interventions continuing to suppress the wild-type mosquito populations even under warmer climates (Supplementary Information, Figures S31–34). Varying inferred life-history trait values altered the projected baseline abundance and widened the credible intervals of projections. These results highlight that climate-driven life-history traits are important determinants of future population dynamics (Supplementary Information, Figures S35–S46). These variations did not alter our interpretation on the projected baseline wild-type populations or the robustness of IIT, although mosquito migration was determined to be a key factor attenuating IE.

## Discussion

Our study collated a globally representative set of mosquito abundance and climate data to calibrate a climatically driven entomological model, aimed at understanding how climate influences mosquito life-history traits and at providing region-specific projections of future mosquito abundance and the efficacy of hypothetical suppression programmes based on release of *Wolbachia*-infected male mosquitoes. Our study empirically examined the potential failure of cytoplasmic incompatibility (CI) under extreme adverse heat conditions, despite CI remaining highly effective across most scenarios. Experimental evidence demonstrated that CI was maintained even under near-lethal thermal conditions, supporting the extrapolation of these effects to aggregated climate projections (Figure 4 and XX). We projected heterogenous changes in mosquito abundance across both tropical and non-tropical regions under future emission scenarios, with the magnitude of change varying substantially by geography. *Aedes* abundance was also expected to rise or remain stable during the warmest months of the year (Figure 6). Importantly, releases of *Wolbachia*-infected males retained effectiveness under future emission scenarios, across diverse geographies, and over extended timescales. Yet, effectiveness was found to depend heavily on mosquito emigration rates, overflooding ratios, release intervals and release strategies (Figure 7). These results provide consistent evidence that IIT can function as an effective, climate-resilient, and sustainable *Aedes*-control tool with global applicability.

In contrast, semi-field [86,87], field [86,88], laboratory [75,89–91] and modelling studies [63] suggested that the *w*Mel replacement strategy may falter under thermal stress, owing to the strategy’s dependence on heat-sensitive bacterial strains in infected female mosquitoes and need for long-term persistence of the infection under field conditions across all life stages [92]. In contrast, IIT relies on the release of *Wolbachia*-infected adult males, and accordingly, only mosquitoes in this life stage are subject to thermal stress exposure. We found that CI is preserved even under extreme thermal stress in experiments conducted here. Incorporating this mechanistic distinction into climate-informed process-based modelling at global scale further demonstrated the robustness of IIT as a future-proof vector control strategy. Our findings are also likely to be generalizable to combined approaches that integrate IIT with SIT, given that both IIT and IIT-SIT rely on the same fundamental mechanism of inducing sterility to suppress wild mosquito populations. Moreover, laboratory studies have demonstrated that mosquitoes produced under IIT-SIT retain life history traits and mating competitiveness comparable to, or even exceeding, those of mosquitoes generated through standalone IIT, further supporting the applicability of our results to IIT-SIT strategies [78].

This work represents the first global, climate-integrated entomological model to project both *Aedes* abundance and IIT effectiveness under climate change. Our approach advances the field by calibrating to real world mosquito abundance data, capturing regional variation in species composition and life-history traits through an extensive dataset assembled from diverse climatic and urban contexts. Study sites were selected to reflect a wide range of climate and geographic regimes—from tropical equatorial Singapore, Mediterranean Europe and the highly diverse continental climates of mainland China and the United States. Contrasting prior work [63], our model adopted an intuitive, climate-informed compartmental framework of mosquito abundance and IIT in which transitions were explicitly modulated by local temperature and precipitation, and it accommodated heterogeneous spatial-temporal input data. Detailed climate projections were integrated with field-inferred parameters, calibrated through a Bayesian HMC scheme [93]. Extensive robustness checks also ensured the internal validity of our modelling framework and insensitivity to changes in model parameters.

Nevertheless, our work has limitations. While the primary purpose of historical data was to infer the region-specific life-history traits of mosquito populations and also conduct validation of model prediction, inference and tests for predictive accuracy may be limited due to the limited length and resolution of available surveillance data. In particular, the model was not able to fully validate model performance over decadal time-scales or during individual extreme climate events. Our modelling framework was designed to evaluate broad climate-associated trends in mosquito abundance and IIT effectiveness over long-term future. Therefore, climate variables were harmonised to the temporal and spatial resolution of observed *Aedes* abundance. Humidity was excluded as modelling variable to avoid nesting large uncertainties from climate projections into our modelling framework. Experimental evidence demonstrated that cytoplasmic incompatibility remained highly effective under both acute and chronic heat stress conditions up to temperatures associated with mosquito mortality, supporting the extrapolation of CI outcomes under projected future climates. However, aggregated climate data may not fully capture the influence of rare extreme weather events on mosquito dynamics and intervention outcomes. In addition, limited surveillance coverage restricted our ability to assess species interactions and potential range shifts, including the expansion of *Ae. aegypti* into regions currently dominated by *Ae. albopictus*. Future work should evaluate these processes as higher-resolution environmental and surveillance data become available.

Changes in biogeographic and anthropogenic factors such as river systems, monsoons, urbanisation levels, and existing vector control infrastructure can further influence the aquatic carrying capacity of mosquitoes [94–96], this information was not available for the future climate change scenarios considered here. The aquatic carrying capacity was thus proxied using a data-informed function and allowed to vary with precipitation. Prevailing vector control practices across regions could have influenced past entomological patterns but cannot be accounted for due to a lack of data– this may led to biases in inferred life-history traits and potentially underestimates future *Aedes* abundance. However, this is unlikely to influence our overall results as extensive sensitivity analyses indicated that deviations from entomological functions are unlikely to influence our projections significantly. We also assumed no major behavioural or physiological differences between released *Wolbachia*-infected male mosquitoes and wild-type males—a reasonable assumption supported by laboratory and semi□field trials showing that *w*AlbB□SG transinfected *aegypti* males or triple-wPip/wAlbA/wAlbB transinfected *albopictus* males exhibit comparable lifespan and mating capacity to their wild-type counterparts [35,78]. However, other studies have shown that thermal sensitivity may be increased in *Wolbachia*-infected *Ae. aegypti* adults [97]. Additionally, no major differences in life-history traits are assumed to develop between wildtype and *Wolbachia*-infected male mosquitoes, consistent with current data [78,99]. This may be primarily due to the use of field-caught mosquitoes which are reared and then used to produce *Wolbachia*-infected mosquitoes. Regular renewal of strains in IIT programmes is also necessary to ensure maintenance of field ability. Thermal response parameters were derived from laboratory experiments and assumed to be approximate to field conditions, and translatable to *Ae. albopictus* species. Our assumption of complete CI is reasonable as experiments done under this study showed that even antibiotic-treated *Wolbachia*-infected *aegypti* males maintained CI despite substantial decreases in density (for *w*MelM) and complete loss from some individuals (for *w*AlbB). This suggests that sperm modification by CI is already completed during the aquatic and young adult stages when males are in the mass-rearing facility and just before release–this is consistent with data from Drosophila suggesting that sperm modification by *Wolbachia* occurs during early spermatogenesis [100] Finally, the inferred area-specific entomological features were also obtained through calibration of models to abundance in different areas. Future work should aim to validate these model-derived estimates against independent field studies. However, sensitivity analyses of entomological parameters showed no major differences in the robustness of the intervention (See Supplementary Information Figures S31-42).

## Conclusion

Our evidence showed that IIT can be a highly effective, climate-resilient and sustainable *Aedes*-control tool and can be adopted globally.

## Data Availability

All data produced in the present study are available upon reasonable request to the authors.

https://github.com/lineliott/IITRobust

## Acknowledgement

We thank Apeksha Warusawithana and Lizzy Li for assistance with the *Wolbachia* thermal stability experiments and Mason for molecular screening of *Wolbachia* in the antibiotic treatment experiment.

