## Supplementary Information for "Robustness of Wolbachia-mediated incompatible-insect technique to future climate change scenarios"

**1. Model calibration**

**1.1 Summary of calibrated parameters**

We calibrated the recorded time series of mosquito abundance in the finest spatial resolution to the process-based model using Hamilton Monte Carlo (HMC). Priors were set according to prevailing literature for the model parameters that reflected existing knowledge and uncertainty about their likely range of values (Table S1–S2). Laboratory and semi-field studies have reported significant thermal sensitivity differences in life-history traits between *Ae. Aegypti* and *Ae. Albopictus*. Empirically, the longevity of female mosquitoes is observed to be longer than that of male mosquitoes due to blood acquisition. These differences were incorporated into the prior information according to the mosquito stage, sex, and species while calibrating the model. The climatically forced parameters and initial values for each of the model’s compartments were assigned broad parameter spaces to reflect uncertainty. Transition rate parameters ($L_{\eta}$, $\eta\in(\mu_{a}, \mu_{f}, \tau, \rho,\alpha)$) are presented on a weekly basis.

**Table S1 Fitting parameters**

| $\Theta$ | Definition | Parameter boundary | Prior information |
| --- | --- | --- | --- |
| $L_{\mu,a}$ | Aquatic mortality scale parameter | [0,0.9] | U (0, 0.9) |
| $T_{1,\mu,a}$ | Aquatic mortality lower midpoint temperature parameter | *Ae. Aegypti*: [10, 20]  *Ae. Albopictus*: [-15, -5] | *Ae. Aegypti*: N (15, 0.1) [1–4]  *Ae. Albopictus*: N (-10, 0.1) [1,4,5] |
| $k_{1,\mu,a}$ | Aquatic mortality lower rate parameter | [0.5, 1] | U (0.5, 1) |
| $k_{2,\mu,a}$ | Aquatic mortality higher rate parameter | [0.5, 1] | U (0.5, 1) |
| $L_{\mu,f}$ | Female mortality scale parameter | [0, 0.9] | *Ae. Aegypti*: N (0.67, 0.1) [1,2,6,7]  *Ae. Albopictus*: N (0.75, 0.1) [1,5,6,8–10] |
| $T_{1,\mu,f}$ | Female mortality lower midpoint temperature parameter | *Ae. Aegypti*: [10, 20]  *Ae. Albopictus*: [5, 15] | *Ae. Aegypti*: N (15, 0.1) [1,2,7]  *Ae. Albopictus*: N (10, 0.1) [1,5,8–10] |
| $k_{1,\mu,f}$ | Female mortality lower rate parameter | [0.5, 1] | U (0.5, 1) |
| $k_{2,\mu,f}$ | Female mortality higher rate parameter | [0.5, 1] | U (0.5, 1) |
| $L_{\tau}$ | Development scale parameter | [0.1, 1] | *Ae. Aegypti*: N (1, 0.1) [1–3,11,12]  *Ae. Albopictus*: N (0.5, 0.1) [1,5,8–10] |
| $T_{1,\tau}$ | Development lower midpoint temperature parameter | *Ae. Aegypti*: [10, 20]  *Ae. Albopictus*: [5, 15] | *Ae. Aegypti*: N (15, 0.1) [1–3,11,12]  *Ae. Albopictus*: N (10, 0.1) [1,5,8–10] |
| $k_{1,\tau}$ | Development lower rate parameter | [0.5, 1] | U (0.5, 1) |
| $k_{2,\tau}$ | Development higher rate parameter | [0.5, 1] | U (0.5, 1) |
| $L_{\rho}$ | Reproduction scale parameter | [1, 10] | U (1, 10) |
| $T_{1,\rho}$ | Reproduction lower midpoint temperature parameter | *Ae. Aegypti*: [20, 30]  *Ae. Albopictus*: [10, 20] | *Ae. Aegypti*: N (25, 0.1) [1,2,7,11]  *Ae. Albopictus*: N (15, 0.1) [1,5,8–10] |
| $k_{1,\rho}$ | Reproduction lower rate parameter | [0.5, 1] | U (0.5, 1) |
| $k_{2,\rho}$ | Reproduction higher rate parameter | [0.5, 1] | U (0.5, 1) |
| $\alpha$ | Migration rate | [0, 0.2] | U (0, 1) |
| $\delta$ | Mortality difference by sex | [0.5, 1] | *Ae. Aegypti*: N (0.67, 0.1) [6]  *Ae. Albopictus*: N (0.67, 0.1) [5,6,8,10] |
| $\omega$ | Environmental capacity constant | CN:[0.01, 10];  EU:[0.01, 40]  US:[0.01, 1];  SG:[0.01, 0.1] | CN:U (0.01, 10);  EU:U (0.01, 40)  US:U (0.01, 1);  SG:U (0.01, 0.1) |
| $\lambda$ | Environmental capacity rate parameter | CN:[0.01, 3];  EU:[0.01, 20]  US:[0.01, 0.3];  SG:[0.01, 0.1] | CN: U (0.01, 3);  EU: U (0.01, 20)  US:U (0.01, 0.3);  SG:U (0.01, 0.1) |
| $F_{n,0}$ | Initial value of $F_{n}$ | CN:[0, 20];  EU:[0, 100]  US:[0,100];  SG:[0, 1] | CN: U (0, 20);  EU: U (0, 100)  US: U (0,100);  SG: U (0, 1) |
| $M_{n,0}$ | Initial value of $M_{n}$ | CN:[0, 20];  EU:[0, 100]  US:[0,100];  SG:[0, 1] | CN: U (0, 20);  EU: U (0, 100)  US: U (0,100);  SG: U (0, 1) |
| $A_{n,0}$ | Initial value of $A_{n}$ | CN:[0, 20];  EU:[0, 100]  US:[0,100];  SG:[0, 1] | CN: U (0, 20);  EU: U (0, 100)  US: U (0,100);  SG: U (0, 1) |
| $\sigma$ | Observation process standard deviation | [0, 5] | U(0, 5) |

**Table S2 Fitted parameters**

| $\epsilon$ | Definition | *Aegypti* | *Albopictus* | Reference |
| --- | --- | --- | --- | --- |
| $T_{2,\mu,a}$ | Aquatic mosquito mortality higher midpoint temperature parameter | $34℃$ | $34℃$ | [1–3,11] |
| $T_{2,\mu,m}$ | Mature mosquito mortality higher midpoint temperature parameter | $34℃$ | $34℃$ |  |
| $T_{2,\tau}$ | Development higher midpoint temperature parameter | $32℃$ | $32℃$ |  |
| $T_{2,\rho}$ | Reproduction higher midpoint temperature parameter | $32℃$ | $32℃$ |  |

**1.2 Calibration results**

We performed calibration by location using the GFDL-ESM4 climate model for the main results. We compiled the estimates by study region for each parameter according to their SSPs and areas in Table S3–S6. Point estimates were reported as posterior means with accompanying 95% credible intervals in parentheses.

**Table S3 Posterior summary of mainland China by different SSPs and areas.**

| SSP | Area | $A_{n,0}$ | $F_{n,0}$ | $M_{n,0}$ | $\delta$ | $T_{1,\mu,a}$ | $T_{1,\mu,f}$ | $T_{1,\tau}$ | $T_{1,\rho}$ | $L_{\mu,a}$ | $L_{\mu,f}$ | $L_{\tau}$ | $L_{\rho}$ | $k_{1,\mu,a}$ | $k_{1,\mu,f}$ | $k_{1,\tau}$ | $k_{1,\rho}$ | $k_{2,\mu,a}$ | $k_{2,\mu,f}$ | $k_{2,\tau}$ | $k_{2,\rho}$ | $\omega$ | $\lambda$ | $\sigma$ | $\alpha$ |
| --- | --- | --- | --- | --- | --- | --- | --- | --- | --- | --- | --- | --- | --- | --- | --- | --- | --- | --- | --- | --- | --- | --- | --- | --- | --- |
| ssp126 | Anhui | 9.64  (0.24-19.50) | 0.16  (0.01-0.50) | 9.81  (0.47-19.48) | 0.68  (0.52-0.86) | -10.00  (-10.19--9.80) | 9.99  (9.79-10.19) | 10.00  (9.81-10.21) | 15.01  (14.83-15.20) | 0.85  (0.72-0.90) | 0.71  (0.61-0.81) | 0.45  (0.26-0.65) | 8.59  (6.19-9.94) | 0.75  (0.51-0.99) | 0.76  (0.52-0.99) | 0.82  (0.54-0.99) | 0.53  (0.50-0.60) | 0.75  (0.51-0.99) | 0.92  (0.76-1.00) | 0.89  (0.64-1.00) | 0.93  (0.78-1.00) | 0.91  (0.47-1.82) | 0.10  (0.05-0.19) | 3.04  (2.64-3.51) | 0.11  (0.01-0.20) |
| ssp126 | Chongqing | 9.75  (0.42-19.49) | 0.64  (0.28-1.04) | 9.85  (0.42-19.40) | 0.68  (0.52-0.87) | -10.00  (-10.21--9.81) | 9.99  (9.79-10.20) | 10.01  (9.82-10.20) | 14.94  (14.74-15.13) | 0.86  (0.75-0.90) | 0.61  (0.49-0.71) | 0.47  (0.29-0.66) | 8.89  (7.04-9.95) | 0.75  (0.51-0.99) | 0.80  (0.53-0.99) | 0.86  (0.59-1.00) | 0.54  (0.50-0.64) | 0.75  (0.51-0.99) | 0.81  (0.54-0.99) | 0.83  (0.54-0.99) | 0.89  (0.65-1.00) | 1.43  (0.81-2.46) | 0.05  (0.02-0.09) | 2.02  (1.76-2.32) | 0.07  (0.00-0.18) |
| ssp126 | Fujian | 9.73  (0.27-19.46) | 0.23  (0.00-0.86) | 9.97  (0.53-19.36) | 0.68  (0.52-0.88) | -10.00  (-10.20--9.80) | 9.98  (9.78-10.18) | 9.99  (9.79-10.19) | 14.95  (14.76-15.14) | 0.87  (0.79-0.90) | 0.67  (0.58-0.78) | 0.51  (0.33-0.70) | 9.41  (8.16-9.98) | 0.75  (0.51-0.99) | 0.66  (0.50-0.96) | 0.74  (0.51-0.99) | 0.52  (0.50-0.57) | 0.75  (0.51-0.99) | 0.81  (0.55-0.99) | 0.79  (0.53-0.99) | 0.86  (0.61-1.00) | 0.66  (0.33-1.21) | 0.06  (0.04-0.10) | 2.96  (2.63-3.33) | 0.14  (0.02-0.20) |
| ssp126 | Guangxi | 10.18  (0.85-19.43) | 0.92  (0.40-1.76) | 10.04  (0.58-19.61) | 0.68  (0.53-0.87) | -10.00  (-10.19--9.81) | 9.99  (9.79-10.18) | 10.00  (9.80-10.20) | 15.01  (14.82-15.20) | 0.77  (0.38-0.90) | 0.67  (0.56-0.78) | 0.47  (0.26-0.66) | 6.92  (3.65-9.81) | 0.75  (0.51-0.99) | 0.76  (0.51-0.99) | 0.75  (0.51-0.99) | 0.54  (0.50-0.68) | 0.75  (0.51-0.99) | 0.88  (0.66-1.00) | 0.88  (0.62-1.00) | 0.91  (0.73-1.00) | 0.63  (0.26-1.45) | 0.05  (0.02-0.10) | 1.51  (1.34-1.71) | 0.10  (0.01-0.19) |
| ssp126 | Hainan | 10.71  (0.75-19.62) | 10.20  (2.80-17.52) | 10.00  (0.57-19.48) | 0.68  (0.52-0.87) | -10.00  (-10.19--9.81) | 10.01  (9.81-10.20) | 10.01  (9.81-10.21) | 15.03  (14.84-15.24) | 0.45  (0.03-0.86) | 0.67  (0.53-0.80) | 0.49  (0.31-0.67) | 1.75  (1.02-4.50) | 0.75  (0.51-0.99) | 0.63  (0.50-0.97) | 0.71  (0.51-0.99) | 0.55  (0.50-0.74) | 0.76  (0.51-0.99) | 0.95  (0.83-1.00) | 0.95  (0.84-1.00) | 0.96  (0.86-1.00) | 7.82  (2.86-9.95) | 0.47  (0.07-1.32) | 3.86  (3.45-4.32) | 0.08  (0.00-0.19) |
| ssp126 | Hebei | 9.52  (0.34-19.29) | 1.08  (0.09-2.86) | 9.85  (0.48-19.53) | 0.68  (0.53-0.86) | -10.00  (-10.19--9.80) | 10.00  (9.81-10.20) | 10.00  (9.80-10.20) | 15.00  (14.80-15.20) | 0.82  (0.51-0.90) | 0.72  (0.59-0.85) | 0.46  (0.27-0.66) | 6.41  (3.30-9.69) | 0.75  (0.52-0.99) | 0.76  (0.52-0.99) | 0.77  (0.51-0.99) | 0.65  (0.50-0.95) | 0.75  (0.51-0.99) | 0.76  (0.51-0.99) | 0.76  (0.52-0.99) | 0.75  (0.51-0.99) | 3.30  (1.20-7.76) | 0.09  (0.01-0.28) | 6.84  (5.79-8.12) | 0.09  (0.01-0.19) |
| ssp126 | Henan | 9.66  (0.36-19.49) | 1.08  (0.24-2.25) | 10.15  (0.40-19.58) | 0.68  (0.52-0.86) | -10.00  (-10.19--9.81) | 9.99  (9.79-10.19) | 10.00  (9.80-10.20) | 15.00  (14.81-15.20) | 0.86  (0.73-0.90) | 0.69  (0.58-0.80) | 0.44  (0.25-0.63) | 8.09  (4.80-9.91) | 0.75  (0.51-0.99) | 0.76  (0.52-0.99) | 0.78  (0.52-0.99) | 0.53  (0.50-0.61) | 0.75  (0.51-0.99) | 0.84  (0.60-0.99) | 0.82  (0.56-0.99) | 0.88  (0.66-1.00) | 2.84  (1.35-6.62) | 0.16  (0.06-0.35) | 5.51  (4.81-6.33) | 0.10  (0.01-0.19) |
| ssp126 | Hubei | 10.07  (0.58-19.45) | 1.48  (0.96-2.15) | 10.07  (0.51-19.59) | 0.68  (0.53-0.87) | -10.00  (-10.19--9.80) | 9.99  (9.79-10.18) | 10.01  (9.81-10.20) | 14.97  (14.78-15.17) | 0.87  (0.76-0.90) | 0.70  (0.58-0.82) | 0.45  (0.25-0.64) | 8.89  (6.68-9.96) | 0.75  (0.51-0.99) | 0.77  (0.52-0.99) | 0.82  (0.54-0.99) | 0.55  (0.50-0.68) | 0.75  (0.51-0.99) | 0.69  (0.51-0.98) | 0.71  (0.51-0.98) | 0.85  (0.58-0.99) | 1.01  (0.47-2.11) | 0.06  (0.02-0.12) | 3.47  (3.02-4.01) | 0.10  (0.01-0.19) |
| ssp126 | Hunan | 10.12  (0.50-19.50) | 2.01  (1.03-3.07) | 9.93  (0.37-19.55) | 0.68  (0.53-0.86) | -10.00  (-10.19--9.80) | 9.99  (9.80-10.18) | 10.01  (9.81-10.20) | 14.98  (14.78-15.17) | 0.85  (0.72-0.90) | 0.62  (0.50-0.72) | 0.46  (0.28-0.64) | 9.24  (7.67-9.98) | 0.75  (0.51-0.99) | 0.78  (0.52-0.99) | 0.82  (0.55-0.99) | 0.54  (0.50-0.64) | 0.75  (0.51-0.99) | 0.76  (0.51-0.99) | 0.73  (0.51-0.99) | 0.85  (0.58-1.00) | 1.01  (0.42-2.07) | 0.28  (0.16-0.48) | 5.93  (5.14-6.84) | 0.07  (0.00-0.18) |
| ssp126 | Jiangsu | 9.91  (0.49-19.57) | 0.33  (0.10-0.64) | 9.83  (0.41-19.45) | 0.68  (0.52-0.87) | -10.00  (-10.20--9.80) | 9.99  (9.80-10.19) | 10.01  (9.81-10.20) | 15.00  (14.80-15.20) | 0.86  (0.71-0.90) | 0.67  (0.54-0.79) | 0.41  (0.21-0.61) | 8.17  (5.22-9.93) | 0.75  (0.51-0.98) | 0.77  (0.51-0.99) | 0.82  (0.54-0.99) | 0.54  (0.50-0.66) | 0.75  (0.51-0.99) | 0.85  (0.61-0.99) | 0.84  (0.56-0.99) | 0.91  (0.73-1.00) | 0.68  (0.31-1.55) | 0.02  (0.01-0.04) | 1.34  (1.16-1.55) | 0.09  (0.00-0.19) |
| ssp126 | Jiangxi | 10.14  (0.57-19.52) | 1.92  (1.41-2.49) | 10.12  (0.48-19.53) | 0.68  (0.52-0.87) | -10.00  (-10.20--9.81) | 9.98  (9.77-10.18) | 9.99  (9.79-10.19) | 14.96  (14.76-15.14) | 0.84  (0.68-0.90) | 0.70  (0.61-0.80) | 0.51  (0.34-0.69) | 9.37  (7.97-9.98) | 0.75  (0.51-0.99) | 0.75  (0.51-0.99) | 0.74  (0.51-0.99) | 0.53  (0.50-0.63) | 0.75  (0.51-0.99) | 0.72  (0.51-0.98) | 0.72  (0.51-0.98) | 0.85  (0.58-0.99) | 0.41  (0.13-0.86) | 0.09  (0.06-0.15) | 4.00  (3.45-4.65) | 0.15  (0.04-0.20) |
| ssp126 | Liaoning | 9.85  (0.37-19.49) | 0.11  (0.00-0.34) | 9.99  (0.48-19.54) | 0.68  (0.52-0.86) | -10.00  (-10.20--9.80) | 10.00  (9.81-10.19) | 10.00  (9.80-10.20) | 15.00  (14.80-15.19) | 0.46  (0.03-0.86) | 0.65  (0.50-0.80) | 0.43  (0.23-0.63) | 5.53  (3.12-9.23) | 0.74  (0.51-0.98) | 0.76  (0.51-0.99) | 0.76  (0.52-0.99) | 0.62  (0.50-0.93) | 0.75  (0.51-0.99) | 0.76  (0.51-0.99) | 0.76  (0.51-0.99) | 0.77  (0.52-0.99) | 0.56  (0.22-1.11) | 0.02  (0.01-0.06) | 0.94  (0.80-1.12) | 0.07  (0.00-0.18) |
| ssp126 | Shaanxi | 10.06  (0.53-19.56) | 7.09  (2.83-16.43) | 9.81  (0.41-19.48) | 0.68  (0.52-0.86) | -10.00  (-10.19--9.81) | 10.00  (9.81-10.19) | 10.01  (9.81-10.21) | 15.01  (14.81-15.22) | 0.74  (0.18-0.90) | 0.80  (0.71-0.89) | 0.45  (0.23-0.66) | 4.66  (1.52-9.25) | 0.75  (0.51-0.99) | 0.71  (0.51-0.98) | 0.78  (0.52-0.99) | 0.65  (0.50-0.96) | 0.76  (0.51-0.99) | 0.75  (0.52-0.99) | 0.75  (0.51-0.99) | 0.77  (0.52-0.99) | 0.92  (0.10-5.13) | 0.37  (0.03-2.35) | 5.35  (4.46-6.51) | 0.13  (0.04-0.20) |
| ssp126 | Shandong | 2.49  (0.06-16.81) | 0.29  (0.01-0.91) | 9.92  (0.52-19.41) | 0.68  (0.52-0.87) | -10.00  (-10.19--9.81) | 10.00  (9.80-10.19) | 10.00  (9.81-10.20) | 14.99  (14.79-15.19) | 0.77  (0.26-0.90) | 0.66  (0.49-0.82) | 0.41  (0.21-0.60) | 3.40  (2.20-6.96) | 0.75  (0.51-0.98) | 0.76  (0.52-0.99) | 0.76  (0.52-0.99) | 0.67  (0.50-0.97) | 0.75  (0.51-0.99) | 0.78  (0.53-0.99) | 0.79  (0.52-0.99) | 0.83  (0.56-0.99) | 5.29  (0.89-9.64) | 0.30  (0.02-1.19) | 3.43  (2.86-4.14) | 0.07  (0.00-0.19) |
| ssp126 | Shanghai | 9.87  (0.36-19.50) | 0.22  (0.02-0.51) | 10.17  (0.49-19.51) | 0.68  (0.52-0.87) | -10.00  (-10.20--9.80) | 9.99  (9.79-10.19) | 10.01  (9.82-10.20) | 14.99  (14.79-15.19) | 0.86  (0.76-0.90) | 0.63  (0.51-0.74) | 0.44  (0.23-0.63) | 8.95  (6.81-9.96) | 0.75  (0.51-0.99) | 0.77  (0.52-0.99) | 0.81  (0.54-0.99) | 0.53  (0.50-0.63) | 0.75  (0.51-0.99) | 0.91  (0.70-1.00) | 0.87  (0.58-1.00) | 0.93  (0.76-1.00) | 0.81  (0.41-1.66) | 0.05  (0.02-0.10) | 1.78  (1.54-2.06) | 0.09  (0.01-0.19) |
| ssp126 | Shanxi | 9.96  (0.64-19.35) | 0.35  (0.06-0.79) | 10.05  (0.41-19.60) | 0.68  (0.52-0.87) | -10.00  (-10.20--9.81) | 10.00  (9.81-10.19) | 10.00  (9.81-10.18) | 14.99  (14.79-15.18) | 0.64  (0.09-0.89) | 0.76  (0.64-0.87) | 0.44  (0.23-0.64) | 4.87  (2.55-8.85) | 0.75  (0.51-0.98) | 0.74  (0.51-0.99) | 0.73  (0.51-0.99) | 0.55  (0.50-0.68) | 0.75  (0.51-0.99) | 0.75  (0.51-0.99) | 0.75  (0.51-0.99) | 0.76  (0.51-0.99) | 1.00  (0.28-2.77) | 0.07  (0.01-0.20) | 1.24  (1.05-1.47) | 0.11  (0.01-0.20) |
| ssp126 | Sichuan | 9.67  (0.36-19.48) | 1.90  (0.07-5.17) | 10.01  (0.47-19.45) | 0.68  (0.52-0.86) | -10.00  (-10.19--9.80) | 9.98  (9.78-10.18) | 10.00  (9.81-10.20) | 14.91  (14.72-15.11) | 0.89  (0.84-0.90) | 0.57  (0.43-0.70) | 0.49  (0.30-0.67) | 9.49  (8.21-9.99) | 0.76  (0.52-0.99) | 0.79  (0.52-0.99) | 0.86  (0.60-1.00) | 0.52  (0.50-0.57) | 0.75  (0.51-0.99) | 0.75  (0.51-0.99) | 0.75  (0.51-0.99) | 0.75  (0.51-0.99) | 2.99  (1.59-5.19) | 0.07  (0.01-0.19) | 6.66  (5.81-7.68) | 0.07  (0.00-0.18) |
| ssp126 | Tianjin | 9.93  (0.59-19.51) | 0.79  (0.19-1.56) | 10.03  (0.61-19.49) | 0.68  (0.52-0.87) | -10.00  (-10.20--9.80) | 10.00  (9.82-10.19) | 10.00  (9.80-10.20) | 15.00  (14.80-15.21) | 0.79  (0.37-0.90) | 0.60  (0.44-0.75) | 0.47  (0.27-0.66) | 2.80  (1.87-4.67) | 0.77  (0.52-0.99) | 0.76  (0.52-0.99) | 0.78  (0.52-0.99) | 0.76  (0.51-0.99) | 0.75  (0.51-0.99) | 0.82  (0.54-0.99) | 0.84  (0.56-0.99) | 0.87  (0.62-1.00) | 1.55  (0.63-2.74) | 0.02  (0.01-0.06) | 1.10  (0.92-1.32) | 0.05  (0.00-0.16) |
| ssp126 | Yunnan | 10.27  (0.73-19.49) | 15.31  (6.37-19.81) | 9.95  (0.59-19.42) | 0.68  (0.52-0.87) | -10.00  (-10.19--9.81) | 9.97  (9.78-10.17) | 9.99  (9.80-10.18) | 14.95  (14.77-15.14) | 0.60  (0.03-0.90) | 0.67  (0.57-0.78) | 0.43  (0.24-0.62) | 2.94  (2.09-4.45) | 0.75  (0.51-0.99) | 0.73  (0.51-0.98) | 0.77  (0.52-0.99) | 0.53  (0.50-0.60) | 0.75  (0.51-0.99) | 0.75  (0.51-0.98) | 0.75  (0.52-0.99) | 0.76  (0.51-0.99) | 1.71  (0.61-5.01) | 0.27  (0.15-0.54) | 1.32  (1.19-1.47) | 0.14  (0.03-0.20) |
| ssp126 | Zhejiang | 9.45  (0.27-19.40) | 1.34  (0.11-3.02) | 10.06  (0.53-19.55) | 0.68  (0.52-0.87) | -10.00  (-10.19--9.81) | 9.96  (9.76-10.16) | 10.02  (9.83-10.22) | 14.85  (14.67-15.04) | 0.89  (0.88-0.90) | 0.53  (0.42-0.65) | 0.41  (0.25-0.57) | 9.77  (9.17-9.99) | 0.75  (0.51-0.99) | 0.79  (0.52-0.99) | 0.89  (0.67-1.00) | 0.51  (0.50-0.54) | 0.75  (0.51-0.98) | 0.89  (0.68-1.00) | 0.91  (0.72-1.00) | 0.95  (0.84-1.00) | 3.39  (1.93-5.95) | 0.06  (0.02-0.13) | 3.52  (3.08-4.02) | 0.13  (0.01-0.20) |
| ssp370 | Anhui | 9.77  (0.30-19.40) | 0.22  (0.01-0.60) | 10.09  (0.48-19.47) | 0.68  (0.52-0.87) | -10.00  (-10.20--9.80) | 9.99  (9.80-10.19) | 10.00  (9.80-10.19) | 15.01  (14.83-15.20) | 0.84  (0.65-0.90) | 0.71  (0.61-0.82) | 0.47  (0.28-0.66) | 8.31  (5.77-9.92) | 0.75  (0.51-0.99) | 0.73  (0.51-0.98) | 0.78  (0.52-0.99) | 0.54  (0.50-0.65) | 0.75  (0.51-0.99) | 0.88  (0.66-1.00) | 0.83  (0.54-0.99) | 0.92  (0.73-1.00) | 0.81  (0.38-1.66) | 0.10  (0.05-0.19) | 2.87  (2.49-3.33) | 0.11  (0.01-0.20) |
| ssp370 | Chongqing | 9.57  (0.29-19.50) | 0.76  (0.35-1.30) | 10.01  (0.59-19.47) | 0.68  (0.52-0.86) | -10.00  (-10.19--9.80) | 9.99  (9.79-10.19) | 10.00  (9.81-10.19) | 14.99  (14.80-15.18) | 0.83  (0.66-0.90) | 0.62  (0.50-0.73) | 0.47  (0.27-0.66) | 8.33  (6.15-9.89) | 0.75  (0.51-0.99) | 0.78  (0.52-0.99) | 0.80  (0.53-0.99) | 0.53  (0.50-0.61) | 0.75  (0.51-0.99) | 0.78  (0.53-0.99) | 0.79  (0.52-0.99) | 0.86  (0.61-0.99) | 1.56  (0.87-2.91) | 0.03  (0.01-0.07) | 1.90  (1.66-2.19) | 0.07  (0.00-0.18) |
| ssp370 | Fujian | 9.56  (0.25-19.41) | 0.48  (0.01-1.70) | 9.91  (0.43-19.45) | 0.68  (0.52-0.86) | -10.00  (-10.19--9.81) | 9.99  (9.79-10.18) | 10.00  (9.81-10.20) | 14.95  (14.75-15.14) | 0.85  (0.70-0.90) | 0.68  (0.57-0.78) | 0.48  (0.28-0.67) | 9.05  (7.22-9.97) | 0.75  (0.52-0.98) | 0.71  (0.51-0.98) | 0.74  (0.51-0.99) | 0.52  (0.50-0.59) | 0.75  (0.51-0.99) | 0.69  (0.50-0.98) | 0.68  (0.51-0.98) | 0.80  (0.53-0.99) | 0.88  (0.47-1.62) | 0.06  (0.03-0.11) | 2.74  (2.45-3.07) | 0.10  (0.01-0.19) |
| ssp370 | Guangxi | 10.02  (0.56-19.54) | 2.04  (1.15-3.81) | 10.12  (0.56-19.40) | 0.68  (0.52-0.86) | -10.00  (-10.20--9.80) | 9.98  (9.79-10.17) | 10.00  (9.80-10.20) | 15.00  (14.80-15.19) | 0.72  (0.22-0.90) | 0.69  (0.59-0.80) | 0.48  (0.29-0.67) | 7.25  (3.87-9.86) | 0.74  (0.51-0.98) | 0.82  (0.55-0.99) | 0.77  (0.52-0.99) | 0.54  (0.50-0.67) | 0.75  (0.51-0.99) | 0.86  (0.62-1.00) | 0.79  (0.52-0.99) | 0.91  (0.73-1.00) | 0.44  (0.20-0.98) | 0.05  (0.02-0.10) | 1.44  (1.29-1.62) | 0.12  (0.01-0.20) |
| ssp370 | Hainan | 11.14  (1.10-19.45) | 12.16  (5.34-17.87) | 8.74  (0.70-19.22) | 0.66  (0.50-0.87) | -10.01  (-10.20--9.82) | 10.01  (9.83-10.22) | 10.00  (9.81-10.19) | 15.03  (14.84-15.23) | 0.24  (0.01-0.69) | 0.69  (0.53-0.82) | 0.51  (0.33-0.68) | 1.40  (1.02-2.44) | 0.75  (0.51-0.98) | 0.62  (0.50-0.97) | 0.74  (0.51-0.98) | 0.61  (0.50-0.93) | 0.74  (0.52-0.99) | 0.94  (0.80-1.00) | 0.94  (0.78-1.00) | 0.94  (0.84-1.00) | 7.64  (0.49-9.97) | 0.36  (0.01-0.69) | 4.00  (3.58-4.44) | 0.10  (0.00-0.19) |
| ssp370 | Hebei | 9.89  (0.55-19.44) | 1.62  (0.16-4.12) | 10.05  (0.60-19.41) | 0.68  (0.52-0.87) | -10.00  (-10.20--9.80) | 10.00  (9.80-10.20) | 10.00  (9.80-10.20) | 14.99  (14.79-15.18) | 0.76  (0.26-0.90) | 0.73  (0.60-0.86) | 0.44  (0.25-0.65) | 5.11  (2.39-9.39) | 0.76  (0.52-0.99) | 0.76  (0.51-0.99) | 0.76  (0.51-0.99) | 0.60  (0.50-0.91) | 0.75  (0.51-0.99) | 0.75  (0.52-0.99) | 0.75  (0.51-0.99) | 0.75  (0.51-0.99) | 4.12  (1.17-9.10) | 0.24  (0.01-1.00) | 7.46  (6.30-8.89) | 0.10  (0.01-0.19) |
| ssp370 | Henan | 9.67  (0.41-19.35) | 1.50  (0.24-3.49) | 9.98  (0.49-19.45) | 0.68  (0.52-0.87) | -10.00  (-10.20--9.81) | 9.99  (9.79-10.19) | 10.00  (9.81-10.19) | 15.01  (14.83-15.21) | 0.71  (0.20-0.90) | 0.71  (0.58-0.83) | 0.42  (0.23-0.63) | 7.22  (3.93-9.87) | 0.75  (0.51-0.99) | 0.73  (0.51-0.99) | 0.75  (0.51-0.99) | 0.53  (0.50-0.60) | 0.75  (0.51-0.99) | 0.85  (0.60-0.99) | 0.84  (0.56-0.99) | 0.88  (0.64-1.00) | 3.52  (1.33-8.42) | 0.06  (0.01-0.18) | 6.42  (5.58-7.39) | 0.10  (0.01-0.19) |
| ssp370 | Hubei | 10.06  (0.51-19.54) | 1.95  (1.18-3.23) | 9.99  (0.48-19.51) | 0.68  (0.52-0.87) | -10.00  (-10.20--9.81) | 9.99  (9.79-10.18) | 10.00  (9.81-10.19) | 14.99  (14.78-15.19) | 0.84  (0.62-0.90) | 0.70  (0.56-0.83) | 0.44  (0.25-0.64) | 8.08  (5.03-9.95) | 0.75  (0.51-0.99) | 0.77  (0.52-0.99) | 0.79  (0.53-0.99) | 0.57  (0.50-0.80) | 0.75  (0.51-0.99) | 0.75  (0.51-0.99) | 0.75  (0.51-0.99) | 0.82  (0.55-0.99) | 1.23  (0.48-2.94) | 0.03  (0.01-0.08) | 3.67  (3.18-4.23) | 0.10  (0.01-0.19) |
| ssp370 | Hunan | 9.92  (0.42-19.42) | 2.33  (1.29-3.71) | 10.09  (0.67-19.43) | 0.68  (0.52-0.87) | -10.00  (-10.19--9.81) | 9.99  (9.79-10.19) | 10.01  (9.81-10.20) | 14.99  (14.79-15.18) | 0.84  (0.68-0.90) | 0.62  (0.49-0.72) | 0.46  (0.27-0.65) | 8.73  (6.36-9.96) | 0.75  (0.51-0.99) | 0.75  (0.51-0.99) | 0.81  (0.53-0.99) | 0.61  (0.50-0.91) | 0.75  (0.52-0.99) | 0.73  (0.51-0.98) | 0.70  (0.51-0.98) | 0.81  (0.54-0.99) | 1.29  (0.57-2.75) | 0.27  (0.14-0.52) | 5.18  (4.49-5.96) | 0.07  (0.00-0.18) |
| ssp370 | Jiangsu | 10.00  (0.49-19.48) | 0.38  (0.12-0.69) | 9.94  (0.51-19.43) | 0.68  (0.53-0.87) | -10.00  (-10.21--9.81) | 9.99  (9.80-10.18) | 10.00  (9.81-10.19) | 14.99  (14.79-15.18) | 0.85  (0.70-0.90) | 0.65  (0.53-0.77) | 0.46  (0.28-0.66) | 8.31  (5.56-9.92) | 0.76  (0.51-0.99) | 0.76  (0.52-0.98) | 0.80  (0.53-0.99) | 0.57  (0.50-0.81) | 0.75  (0.52-0.99) | 0.80  (0.54-0.99) | 0.80  (0.54-0.99) | 0.89  (0.67-1.00) | 0.52  (0.25-1.04) | 0.03  (0.01-0.06) | 1.18  (1.03-1.36) | 0.08  (0.00-0.19) |
| ssp370 | Jiangxi | 10.12  (0.64-19.45) | 1.94  (1.13-3.01) | 9.97  (0.51-19.42) | 0.68  (0.52-0.87) | -10.00  (-10.19--9.80) | 9.99  (9.79-10.19) | 10.01  (9.82-10.20) | 14.99  (14.79-15.18) | 0.79  (0.46-0.90) | 0.71  (0.57-0.83) | 0.42  (0.23-0.62) | 8.31  (5.72-9.93) | 0.75  (0.51-0.99) | 0.77  (0.52-0.99) | 0.77  (0.52-0.99) | 0.63  (0.50-0.95) | 0.75  (0.51-0.99) | 0.64  (0.50-0.96) | 0.68  (0.51-0.97) | 0.79  (0.53-0.99) | 1.17  (0.45-2.57) | 0.10  (0.04-0.21) | 4.16  (3.61-4.79) | 0.08  (0.00-0.19) |
| ssp370 | Liaoning | 9.80  (0.45-19.47) | 0.11  (0.01-0.34) | 10.10  (0.47-19.42) | 0.68  (0.52-0.87) | -10.00  (-10.19--9.81) | 10.00  (9.81-10.19) | 9.99  (9.80-10.19) | 14.98  (14.79-15.18) | 0.54  (0.03-0.88) | 0.67  (0.52-0.80) | 0.47  (0.28-0.66) | 6.06  (3.06-9.64) | 0.75  (0.51-0.99) | 0.77  (0.52-0.99) | 0.74  (0.51-0.99) | 0.64  (0.50-0.94) | 0.75  (0.51-0.99) | 0.75  (0.51-0.99) | 0.75  (0.51-0.99) | 0.75  (0.51-0.99) | 0.41  (0.14-0.95) | 0.02  (0.01-0.05) | 0.87  (0.73-1.03) | 0.07  (0.00-0.18) |
| ssp370 | Shaanxi | 10.09  (0.49-19.53) | 6.59  (2.79-15.04) | 9.99  (0.61-19.47) | 0.68  (0.52-0.87) | -10.00  (-10.20--9.80) | 10.00  (9.80-10.20) | 10.00  (9.81-10.19) | 15.00  (14.80-15.20) | 0.70  (0.15-0.90) | 0.81  (0.74-0.89) | 0.45  (0.25-0.65) | 5.20  (1.82-9.60) | 0.76  (0.51-0.99) | 0.76  (0.51-0.99) | 0.76  (0.52-0.99) | 0.63  (0.50-0.94) | 0.75  (0.51-0.99) | 0.75  (0.51-0.99) | 0.74  (0.51-0.99) | 0.77  (0.52-0.99) | 0.77  (0.14-2.56) | 0.09  (0.01-0.33) | 5.73  (4.82-6.87) | 0.14  (0.06-0.20) |
| ssp370 | Shandong | 8.78  (0.28-19.44) | 0.38  (0.01-1.28) | 10.05  (0.51-19.46) | 0.68  (0.52-0.86) | -10.00  (-10.20--9.81) | 10.00  (9.80-10.20) | 10.01  (9.82-10.21) | 15.00  (14.81-15.20) | 0.64  (0.07-0.90) | 0.71  (0.56-0.83) | 0.44  (0.25-0.64) | 5.03  (2.33-9.40) | 0.75  (0.51-0.99) | 0.75  (0.51-0.99) | 0.78  (0.52-0.99) | 0.68  (0.50-0.98) | 0.75  (0.51-0.99) | 0.76  (0.52-0.99) | 0.75  (0.51-0.99) | 0.79  (0.53-0.99) | 2.22  (0.48-8.24) | 0.16  (0.04-0.38) | 3.55  (3.02-4.21) | 0.09  (0.00-0.19) |
| ssp370 | Shanghai | 9.71  (0.28-19.44) | 0.20  (0.01-0.53) | 10.06  (0.60-19.55) | 0.68  (0.52-0.87) | -10.00  (-10.19--9.81) | 9.99  (9.80-10.18) | 10.01  (9.81-10.20) | 14.99  (14.80-15.18) | 0.86  (0.74-0.90) | 0.62  (0.49-0.73) | 0.44  (0.24-0.63) | 8.71  (6.34-9.96) | 0.75  (0.51-0.99) | 0.75  (0.51-0.99) | 0.80  (0.53-0.99) | 0.54  (0.50-0.69) | 0.75  (0.51-0.99) | 0.87  (0.63-0.99) | 0.85  (0.56-0.99) | 0.93  (0.77-1.00) | 0.92  (0.46-1.88) | 0.02  (0.01-0.06) | 1.54  (1.34-1.78) | 0.08  (0.00-0.18) |
| ssp370 | Shanxi | 9.95  (0.64-19.47) | 0.45  (0.03-1.21) | 10.00  (0.52-19.45) | 0.68  (0.53-0.87) | -10.00  (-10.19--9.81) | 10.00  (9.79-10.19) | 10.00  (9.80-10.20) | 14.98  (14.79-15.18) | 0.77  (0.22-0.90) | 0.71  (0.56-0.84) | 0.38  (0.16-0.59) | 3.06  (1.85-6.17) | 0.77  (0.52-0.99) | 0.77  (0.52-0.99) | 0.75  (0.51-0.99) | 0.55  (0.50-0.68) | 0.75  (0.51-0.99) | 0.75  (0.52-0.99) | 0.76  (0.51-0.99) | 0.75  (0.51-0.99) | 2.89  (0.71-7.05) | 0.08  (0.01-0.44) | 1.52  (1.27-1.80) | 0.09  (0.01-0.19) |
| ssp370 | Sichuan | 9.94  (0.39-19.60) | 8.22  (0.56-18.36) | 10.10  (0.48-19.49) | 0.68  (0.52-0.88) | -10.00  (-10.20--9.80) | 9.99  (9.79-10.18) | 10.01  (9.82-10.21) | 14.93  (14.73-15.12) | 0.89  (0.86-0.90) | 0.56  (0.42-0.69) | 0.49  (0.32-0.67) | 9.50  (8.30-9.99) | 0.76  (0.52-0.99) | 0.77  (0.52-0.99) | 0.89  (0.66-1.00) | 0.53  (0.50-0.60) | 0.75  (0.51-0.98) | 0.75  (0.51-0.99) | 0.75  (0.51-0.99) | 0.75  (0.51-0.99) | 3.04  (1.57-5.18) | 0.11  (0.01-0.28) | 6.15  (5.36-7.07) | 0.05  (0.00-0.16) |
| ssp370 | Tianjin | 10.04  (0.61-19.43) | 0.50  (0.27-0.93) | 10.02  (0.47-19.55) | 0.68  (0.52-0.87) | -10.00  (-10.19--9.81) | 10.00  (9.80-10.20) | 10.00  (9.81-10.20) | 15.00  (14.80-15.19) | 0.80  (0.45-0.90) | 0.75  (0.63-0.87) | 0.47  (0.26-0.66) | 6.84  (3.47-9.84) | 0.75  (0.52-0.99) | 0.76  (0.51-0.99) | 0.78  (0.52-0.99) | 0.72  (0.51-0.98) | 0.75  (0.51-0.99) | 0.73  (0.51-0.99) | 0.70  (0.51-0.98) | 0.77  (0.52-0.99) | 0.14  (0.04-0.38) | 0.06  (0.02-0.13) | 0.86  (0.72-1.03) | 0.10  (0.01-0.19) |
| ssp370 | Yunnan | 12.70  (4.79-19.69) | 13.13  (1.83-19.76) | 9.96  (0.54-19.58) | 0.68  (0.52-0.87) | -10.00  (-10.21--9.80) | 9.98  (9.78-10.17) | 9.99  (9.79-10.18) | 15.01  (14.81-15.20) | 0.85  (0.69-0.90) | 0.53  (0.38-0.66) | 0.38  (0.23-0.59) | 3.01  (2.41-3.82) | 0.75  (0.52-0.98) | 0.75  (0.51-0.99) | 0.70  (0.51-0.97) | 0.54  (0.50-0.67) | 0.75  (0.52-0.99) | 0.75  (0.52-0.99) | 0.76  (0.51-0.99) | 0.75  (0.51-0.99) | 6.03  (2.92-9.58) | 0.06  (0.01-0.16) | 1.55  (1.39-1.73) | 0.06  (0.00-0.18) |
| ssp370 | Zhejiang | 9.02  (0.20-19.50) | 0.90  (0.06-2.23) | 9.99  (0.59-19.40) | 0.68  (0.52-0.86) | -10.00  (-10.19--9.80) | 9.97  (9.77-10.17) | 10.02  (9.82-10.21) | 14.87  (14.69-15.06) | 0.89  (0.87-0.90) | 0.53  (0.41-0.65) | 0.44  (0.29-0.61) | 9.72  (9.08-9.99) | 0.75  (0.51-0.99) | 0.75  (0.51-0.99) | 0.91  (0.72-1.00) | 0.51  (0.50-0.54) | 0.75  (0.51-0.99) | 0.83  (0.57-0.99) | 0.86  (0.61-0.99) | 0.92  (0.76-1.00) | 3.34  (2.13-5.15) | 0.04  (0.01-0.08) | 3.15  (2.77-3.57) | 0.10  (0.01-0.19) |
| ssp585 | Anhui | 8.69  (0.22-19.31) | 0.34  (0.01-1.17) | 9.83  (0.52-19.48) | 0.68  (0.52-0.86) | -10.00  (-10.20--9.80) | 10.00  (9.81-10.19) | 10.01  (9.81-10.20) | 15.02  (14.82-15.21) | 0.79  (0.43-0.90) | 0.68  (0.56-0.80) | 0.44  (0.23-0.64) | 6.65  (3.35-9.79) | 0.75  (0.51-0.99) | 0.73  (0.51-0.99) | 0.78  (0.52-0.99) | 0.57  (0.50-0.77) | 0.75  (0.51-0.99) | 0.93  (0.77-1.00) | 0.88  (0.59-1.00) | 0.92  (0.73-1.00) | 2.20  (0.76-6.56) | 0.05  (0.01-0.16) | 3.83  (3.32-4.44) | 0.10  (0.01-0.19) |
| ssp585 | Chongqing | 9.61  (0.33-19.43) | 0.66  (0.31-1.06) | 10.04  (0.57-19.52) | 0.68  (0.52-0.86) | -10.00  (-10.19--9.81) | 10.00  (9.80-10.20) | 10.01  (9.82-10.20) | 14.99  (14.80-15.18) | 0.86  (0.77-0.90) | 0.64  (0.52-0.75) | 0.41  (0.23-0.60) | 8.67  (6.56-9.95) | 0.75  (0.51-0.99) | 0.75  (0.51-0.99) | 0.81  (0.53-0.99) | 0.53  (0.50-0.62) | 0.75  (0.51-0.99) | 0.80  (0.54-0.99) | 0.79  (0.52-0.99) | 0.90  (0.69-1.00) | 1.71  (0.93-3.25) | 0.02  (0.01-0.06) | 1.87  (1.63-2.16) | 0.07  (0.00-0.18) |
| ssp585 | Fujian | 8.01  (0.03-19.34) | 0.19  (0.00-0.80) | 10.10  (0.58-19.57) | 0.68  (0.52-0.87) | -10.00  (-10.19--9.81) | 9.96  (9.77-10.16) | 9.99  (9.79-10.18) | 14.95  (14.76-15.15) | 0.87  (0.78-0.90) | 0.60  (0.50-0.71) | 0.45  (0.24-0.65) | 9.40  (8.15-9.98) | 0.75  (0.51-0.99) | 0.69  (0.50-0.98) | 0.69  (0.50-0.98) | 0.52  (0.50-0.57) | 0.75  (0.51-0.99) | 0.76  (0.52-0.99) | 0.77  (0.52-0.99) | 0.81  (0.56-0.99) | 0.96  (0.49-1.93) | 0.11  (0.06-0.21) | 2.98  (2.65-3.35) | 0.14  (0.03-0.20) |
| ssp585 | Guangxi | 10.17  (0.61-19.50) | 0.79  (0.27-1.54) | 9.89  (0.44-19.52) | 0.68  (0.52-0.87) | -10.00  (-10.20--9.80) | 9.99  (9.79-10.19) | 10.00  (9.80-10.19) | 15.00  (14.80-15.19) | 0.71  (0.19-0.90) | 0.61  (0.49-0.73) | 0.46  (0.26-0.66) | 6.99  (4.03-9.80) | 0.75  (0.51-0.99) | 0.74  (0.51-0.98) | 0.75  (0.51-0.99) | 0.56  (0.50-0.74) | 0.75  (0.51-0.99) | 0.86  (0.61-1.00) | 0.84  (0.58-0.99) | 0.89  (0.66-1.00) | 0.78  (0.35-1.73) | 0.07  (0.03-0.14) | 1.54  (1.37-1.74) | 0.08  (0.00-0.19) |
| ssp585 | Hainan | 10.40  (0.81-19.48) | 9.09  (2.70-17.02) | 9.91  (0.53-19.45) | 0.68  (0.52-0.87) | -10.00  (-10.19--9.80) | 10.01  (9.82-10.20) | 10.00  (9.80-10.20) | 15.04  (14.84-15.24) | 0.34  (0.01-0.81) | 0.64  (0.49-0.77) | 0.50  (0.31-0.69) | 1.91  (1.02-4.53) | 0.75  (0.51-0.99) | 0.65  (0.50-0.97) | 0.71  (0.51-0.98) | 0.55  (0.50-0.73) | 0.75  (0.51-0.99) | 0.94  (0.80-1.00) | 0.95  (0.85-1.00) | 0.95  (0.85-1.00) | 8.51  (3.97-9.94) | 0.21  (0.05-0.50) | 3.99  (3.57-4.47) | 0.07  (0.00-0.18) |
| ssp585 | Hebei | 9.88  (0.46-19.54) | 1.32  (0.17-3.30) | 10.09  (0.50-19.43) | 0.68  (0.53-0.86) | -10.00  (-10.20--9.80) | 9.99  (9.80-10.19) | 10.00  (9.82-10.19) | 14.99  (14.80-15.18) | 0.82  (0.50-0.90) | 0.76  (0.63-0.88) | 0.44  (0.25-0.63) | 6.37  (2.99-9.80) | 0.77  (0.52-0.99) | 0.72  (0.51-0.98) | 0.74  (0.51-0.99) | 0.61  (0.50-0.89) | 0.75  (0.51-0.98) | 0.77  (0.52-0.99) | 0.76  (0.52-0.99) | 0.76  (0.51-0.99) | 2.99  (0.89-8.10) | 0.06  (0.01-0.19) | 7.24  (6.09-8.65) | 0.12  (0.01-0.20) |
| ssp585 | Henan | 9.09  (0.24-19.45) | 1.66  (0.27-3.98) | 9.96  (0.36-19.52) | 0.68  (0.52-0.86) | -10.00  (-10.20--9.80) | 10.00  (9.80-10.19) | 10.00  (9.80-10.21) | 15.01  (14.81-15.20) | 0.82  (0.57-0.90) | 0.70  (0.57-0.80) | 0.44  (0.25-0.65) | 5.99  (3.29-9.66) | 0.76  (0.51-0.99) | 0.74  (0.51-0.99) | 0.76  (0.51-0.99) | 0.53  (0.50-0.62) | 0.75  (0.51-0.99) | 0.84  (0.60-0.99) | 0.83  (0.55-0.99) | 0.89  (0.67-1.00) | 4.37  (1.50-9.30) | 0.17  (0.05-0.42) | 5.53  (4.82-6.37) | 0.09  (0.00-0.19) |
| ssp585 | Hubei | 10.08  (0.68-19.51) | 1.90  (1.11-3.15) | 10.02  (0.51-19.45) | 0.68  (0.52-0.88) | -10.00  (-10.20--9.80) | 10.00  (9.79-10.19) | 10.00  (9.81-10.20) | 14.99  (14.81-15.19) | 0.85  (0.68-0.90) | 0.69  (0.55-0.81) | 0.44  (0.25-0.63) | 8.13  (5.06-9.94) | 0.75  (0.51-0.99) | 0.75  (0.52-0.99) | 0.79  (0.53-0.99) | 0.58  (0.50-0.83) | 0.75  (0.51-0.99) | 0.77  (0.52-0.99) | 0.77  (0.52-0.99) | 0.86  (0.61-1.00) | 1.38  (0.56-3.19) | 0.04  (0.01-0.10) | 3.53  (3.07-4.09) | 0.09  (0.00-0.19) |
| ssp585 | Hunan | 10.13  (0.59-19.45) | 2.05  (1.04-3.24) | 9.96  (0.51-19.42) | 0.68  (0.52-0.86) | -10.00  (-10.20--9.80) | 10.00  (9.80-10.19) | 10.02  (9.82-10.21) | 14.99  (14.79-15.19) | 0.86  (0.74-0.90) | 0.61  (0.49-0.72) | 0.45  (0.27-0.63) | 8.98  (6.99-9.97) | 0.75  (0.51-0.99) | 0.75  (0.51-0.98) | 0.83  (0.56-0.99) | 0.58  (0.50-0.79) | 0.75  (0.51-0.99) | 0.75  (0.51-0.99) | 0.73  (0.51-0.98) | 0.83  (0.55-0.99) | 1.45  (0.61-2.83) | 0.25  (0.13-0.45) | 5.65  (4.92-6.54) | 0.07  (0.00-0.18) |
| ssp585 | Jiangsu | 9.83  (0.45-19.42) | 0.37  (0.13-0.66) | 9.92  (0.54-19.50) | 0.68  (0.52-0.87) | -10.00  (-10.19--9.80) | 9.99  (9.80-10.18) | 10.01  (9.81-10.20) | 15.00  (14.80-15.21) | 0.86  (0.74-0.90) | 0.71  (0.59-0.82) | 0.46  (0.27-0.65) | 8.43  (5.29-9.94) | 0.75  (0.51-0.99) | 0.76  (0.51-0.99) | 0.82  (0.54-0.99) | 0.54  (0.50-0.68) | 0.75  (0.51-0.99) | 0.88  (0.67-1.00) | 0.82  (0.54-0.99) | 0.89  (0.65-1.00) | 0.40  (0.19-0.87) | 0.02  (0.01-0.05) | 1.19  (1.03-1.38) | 0.10  (0.01-0.19) |
| ssp585 | Jiangxi | 10.38  (0.69-19.60) | 1.76  (1.15-2.48) | 10.07  (0.47-19.50) | 0.68  (0.52-0.87) | -10.00  (-10.20--9.80) | 9.99  (9.78-10.19) | 10.02  (9.82-10.21) | 14.97  (14.78-15.16) | 0.86  (0.74-0.90) | 0.70  (0.59-0.82) | 0.44  (0.25-0.64) | 9.23  (7.45-9.98) | 0.75  (0.51-0.99) | 0.72  (0.51-0.98) | 0.75  (0.51-0.99) | 0.54  (0.50-0.67) | 0.75  (0.51-0.99) | 0.63  (0.50-0.94) | 0.67  (0.51-0.96) | 0.80  (0.53-0.99) | 0.92  (0.38-1.98) | 0.08  (0.04-0.16) | 4.29  (3.70-4.96) | 0.12  (0.01-0.20) |
| ssp585 | Liaoning | 10.03  (0.49-19.52) | 0.11  (0.01-0.31) | 10.02  (0.61-19.38) | 0.68  (0.52-0.87) | -10.00  (-10.20--9.81) | 10.00  (9.80-10.19) | 10.00  (9.80-10.20) | 14.98  (14.79-15.18) | 0.50  (0.04-0.88) | 0.68  (0.53-0.82) | 0.46  (0.28-0.64) | 6.28  (3.37-9.73) | 0.75  (0.51-0.99) | 0.76  (0.51-0.99) | 0.77  (0.52-0.99) | 0.71  (0.51-0.98) | 0.75  (0.51-0.99) | 0.75  (0.51-0.99) | 0.76  (0.51-0.99) | 0.76  (0.52-0.98) | 0.37  (0.10-0.87) | 0.02  (0.01-0.05) | 0.88  (0.74-1.05) | 0.08  (0.00-0.18) |
| ssp585 | Shaanxi | 9.98  (0.51-19.57) | 6.49  (2.65-15.13) | 9.85  (0.40-19.44) | 0.68  (0.52-0.87) | -10.00  (-10.20--9.80) | 9.99  (9.80-10.19) | 10.00  (9.81-10.20) | 15.01  (14.81-15.20) | 0.71  (0.14-0.90) | 0.82  (0.76-0.89) | 0.46  (0.27-0.65) | 4.80  (1.69-9.36) | 0.75  (0.51-0.98) | 0.75  (0.51-0.99) | 0.76  (0.51-0.99) | 0.59  (0.50-0.87) | 0.75  (0.51-0.99) | 0.76  (0.51-0.99) | 0.75  (0.51-0.99) | 0.76  (0.51-0.99) | 0.91  (0.22-2.92) | 0.04  (0.01-0.13) | 5.37  (4.46-6.44) | 0.15  (0.08-0.20) |
| ssp585 | Shandong | 2.14  (0.06-14.37) | 0.39  (0.01-1.09) | 9.94  (0.55-19.45) | 0.68  (0.52-0.86) | -10.00  (-10.20--9.80) | 9.99  (9.79-10.20) | 10.00  (9.81-10.21) | 15.02  (14.82-15.21) | 0.78  (0.29-0.90) | 0.60  (0.44-0.76) | 0.43  (0.28-0.60) | 2.89  (1.99-4.92) | 0.75  (0.52-0.99) | 0.76  (0.52-0.99) | 0.77  (0.52-0.99) | 0.69  (0.50-0.98) | 0.75  (0.51-0.99) | 0.80  (0.53-0.99) | 0.83  (0.55-0.99) | 0.84  (0.58-0.99) | 7.07  (2.28-9.86) | 0.13  (0.02-0.38) | 3.39  (2.83-4.11) | 0.06  (0.00-0.17) |
| ssp585 | Shanghai | 9.43  (0.18-19.63) | 0.22  (0.01-0.64) | 10.21  (0.76-19.36) | 0.68  (0.52-0.87) | -10.00  (-10.20--9.79) | 10.00  (9.80-10.20) | 10.01  (9.81-10.20) | 15.00  (14.80-15.19) | 0.86  (0.71-0.90) | 0.65  (0.53-0.76) | 0.45  (0.27-0.64) | 8.29  (5.50-9.93) | 0.75  (0.51-0.98) | 0.75  (0.51-0.99) | 0.81  (0.54-0.99) | 0.58  (0.50-0.83) | 0.75  (0.51-0.99) | 0.91  (0.69-1.00) | 0.82  (0.54-0.99) | 0.91  (0.70-1.00) | 0.78  (0.38-1.63) | 0.04  (0.01-0.08) | 1.93  (1.68-2.23) | 0.10  (0.01-0.19) |
| ssp585 | Shanxi | 10.00  (0.58-19.59) | 0.55  (0.16-1.06) | 10.02  (0.54-19.51) | 0.68  (0.52-0.87) | -10.00  (-10.19--9.81) | 9.99  (9.80-10.18) | 10.00  (9.80-10.19) | 14.95  (14.76-15.14) | 0.76  (0.22-0.90) | 0.79  (0.66-0.88) | 0.42  (0.20-0.63) | 3.57  (1.97-7.46) | 0.77  (0.51-0.99) | 0.77  (0.52-0.99) | 0.72  (0.51-0.98) | 0.53  (0.50-0.61) | 0.75  (0.51-0.99) | 0.76  (0.51-0.99) | 0.76  (0.51-0.99) | 0.76  (0.51-0.99) | 1.37  (0.27-4.27) | 0.04  (0.01-0.13) | 1.39  (1.16-1.67) | 0.15  (0.05-0.20) |
| ssp585 | Sichuan | 9.82  (0.37-19.46) | 2.10  (0.13-5.19) | 10.03  (0.48-19.50) | 0.68  (0.52-0.86) | -10.00  (-10.20--9.80) | 9.98  (9.79-10.18) | 10.01  (9.81-10.20) | 14.91  (14.71-15.10) | 0.89  (0.85-0.90) | 0.54  (0.40-0.67) | 0.49  (0.31-0.68) | 9.48  (8.22-9.99) | 0.75  (0.52-0.99) | 0.79  (0.52-0.99) | 0.87  (0.62-1.00) | 0.52  (0.50-0.58) | 0.75  (0.51-0.99) | 0.75  (0.51-0.99) | 0.75  (0.51-0.99) | 0.75  (0.51-0.98) | 3.17  (1.83-5.30) | 0.06  (0.01-0.18) | 6.02  (5.21-6.92) | 0.05  (0.00-0.16) |
| ssp585 | Tianjin | 9.97  (0.54-19.45) | 0.69  (0.20-1.43) | 10.08  (0.52-19.50) | 0.68  (0.52-0.87) | -10.00  (-10.20--9.81) | 10.00  (9.80-10.19) | 10.00  (9.80-10.20) | 15.01  (14.81-15.20) | 0.71  (0.17-0.90) | 0.66  (0.51-0.81) | 0.43  (0.22-0.64) | 3.93  (1.96-7.73) | 0.77  (0.52-0.99) | 0.76  (0.51-0.99) | 0.77  (0.52-0.99) | 0.66  (0.50-0.97) | 0.76  (0.52-0.99) | 0.80  (0.53-0.99) | 0.78  (0.52-0.99) | 0.83  (0.54-0.99) | 0.91  (0.27-2.16) | 0.03  (0.01-0.16) | 1.12  (0.95-1.32) | 0.08  (0.00-0.19) |
| ssp585 | Yunnan | 12.40  (4.07-19.58) | 11.76  (1.10-19.68) | 10.07  (0.54-19.48) | 0.68  (0.52-0.87) | -10.00  (-10.20--9.80) | 9.99  (9.79-10.18) | 10.00  (9.81-10.19) | 14.98  (14.79-15.17) | 0.82  (0.58-0.90) | 0.62  (0.49-0.75) | 0.33  (0.21-0.51) | 2.54  (1.96-3.41) | 0.75  (0.51-0.99) | 0.78  (0.52-0.99) | 0.75  (0.51-0.98) | 0.54  (0.50-0.63) | 0.75  (0.51-0.99) | 0.75  (0.51-0.99) | 0.75  (0.52-0.99) | 0.75  (0.51-0.99) | 6.28  (3.20-9.53) | 0.06  (0.01-0.17) | 1.40  (1.25-1.56) | 0.08  (0.00-0.19) |
| ssp585 | Zhejiang | 9.30  (0.34-19.52) | 1.52  (0.13-3.77) | 10.02  (0.51-19.55) | 0.68  (0.52-0.87) | -10.00  (-10.20--9.81) | 9.99  (9.79-10.18) | 10.03  (9.84-10.23) | 14.92  (14.73-15.10) | 0.89  (0.88-0.90) | 0.53  (0.41-0.65) | 0.34  (0.21-0.50) | 9.66  (8.79-9.99) | 0.75  (0.51-0.99) | 0.76  (0.51-0.99) | 0.92  (0.75-1.00) | 0.52  (0.50-0.56) | 0.75  (0.52-0.98) | 0.87  (0.64-0.99) | 0.91  (0.72-1.00) | 0.94  (0.79-1.00) | 4.66  (2.62-7.88) | 0.07  (0.02-0.15) | 3.51  (3.09-4.00) | 0.08  (0.00-0.19) |
| ssp126 | Beijing | 9.53  (0.22-19.54) | 0.51  (0.04-1.43) | 9.93  (0.59-19.37) | 0.68  (0.52-0.87) | -10.00  (-10.20--9.80) | 10.00  (9.80-10.19) | 10.00  (9.80-10.19) | 15.00  (14.81-15.20) | 0.50  (0.04-0.88) | 0.62  (0.48-0.76) | 0.47  (0.28-0.67) | 5.20  (2.90-8.95) | 0.75  (0.51-0.99) | 0.75  (0.51-0.99) | 0.75  (0.51-0.99) | 0.65  (0.50-0.96) | 0.75  (0.51-0.99) | 0.78  (0.53-0.99) | 0.81  (0.54-0.99) | 0.80  (0.54-0.99) | 1.54  (0.55-3.14) | 0.07  (0.01-0.17) | 2.34  (1.98-2.78) | 0.06  (0.00-0.18) |
| ssp126 | Guangdong | 10.10  (0.63-19.44) | 0.46  (0.09-1.24) | 10.02  (0.44-19.56) | 0.68  (0.52-0.86) | -10.00  (-10.20--9.79) | 10.00  (9.80-10.21) | 10.00  (9.81-10.19) | 15.02  (14.82-15.22) | 0.75  (0.23-0.90) | 0.71  (0.60-0.81) | 0.45  (0.12-0.67) | 6.30  (2.55-9.74) | 0.75  (0.51-0.99) | 0.61  (0.50-0.91) | 0.70  (0.51-0.98) | 0.53  (0.50-0.63) | 0.75  (0.51-0.99) | 0.91  (0.75-1.00) | 0.89  (0.65-1.00) | 0.94  (0.80-1.00) | 0.94  (0.27-4.96) | 0.07  (0.02-0.45) | 1.39  (1.24-1.56) | 0.11  (0.01-0.19) |
| ssp126 | Guizhou | 10.12  (0.57-19.53) | 0.66  (0.38-1.01) | 10.07  (0.45-19.53) | 0.68  (0.52-0.87) | -10.00  (-10.20--9.81) | 9.99  (9.81-10.20) | 10.01  (9.81-10.21) | 14.96  (14.77-15.15) | 0.84  (0.70-0.90) | 0.68  (0.57-0.79) | 0.46  (0.27-0.65) | 8.64  (6.44-9.94) | 0.75  (0.52-0.99) | 0.77  (0.51-0.99) | 0.79  (0.53-0.99) | 0.53  (0.50-0.63) | 0.75  (0.52-0.99) | 0.78  (0.52-0.99) | 0.77  (0.52-0.99) | 0.80  (0.54-0.99) | 0.73  (0.38-1.40) | 0.05  (0.02-0.10) | 1.57  (1.37-1.80) | 0.09  (0.01-0.19) |
| ssp370 | Beijing | 9.98  (0.43-19.53) | 0.61  (0.09-1.38) | 9.95  (0.51-19.47) | 0.68  (0.52-0.87) | -10.00  (-10.20--9.80) | 10.00  (9.81-10.19) | 10.00  (9.81-10.20) | 14.99  (14.80-15.19) | 0.72  (0.20-0.90) | 0.63  (0.50-0.73) | 0.43  (0.25-0.63) | 5.16  (3.18-8.51) | 0.76  (0.51-0.99) | 0.75  (0.51-0.99) | 0.76  (0.51-0.99) | 0.68  (0.51-0.97) | 0.74  (0.51-0.99) | 0.76  (0.51-0.99) | 0.76  (0.51-0.99) | 0.76  (0.52-0.99) | 0.77  (0.22-1.77) | 0.42  (0.20-0.80) | 1.91  (1.61-2.28) | 0.06  (0.00-0.17) |
| ssp370 | Guangdong | 9.91  (0.51-19.45) | 0.82  (0.23-1.82) | 10.07  (0.49-19.55) | 0.68  (0.52-0.87) | -10.00  (-10.20--9.80) | 10.00  (9.81-10.19) | 10.00  (9.80-10.20) | 15.00  (14.81-15.19) | 0.59  (0.07-0.89) | 0.70  (0.59-0.80) | 0.48  (0.28-0.68) | 5.69  (2.66-9.50) | 0.75  (0.51-0.99) | 0.73  (0.51-0.98) | 0.74  (0.51-0.99) | 0.54  (0.50-0.67) | 0.75  (0.51-0.99) | 0.84  (0.60-0.99) | 0.81  (0.54-0.99) | 0.92  (0.76-1.00) | 0.66  (0.28-1.40) | 0.07  (0.03-0.15) | 1.19  (1.07-1.33) | 0.09  (0.01-0.19) |
| ssp370 | Guizhou | 9.88  (0.35-19.44) | 0.80  (0.37-1.41) | 10.11  (0.47-19.45) | 0.68  (0.52-0.86) | -10.00  (-10.19--9.80) | 10.00  (9.80-10.20) | 10.00  (9.80-10.21) | 15.00  (14.80-15.19) | 0.79  (0.47-0.90) | 0.67  (0.54-0.77) | 0.45  (0.25-0.65) | 7.16  (4.47-9.79) | 0.75  (0.51-0.99) | 0.75  (0.51-0.99) | 0.78  (0.52-0.99) | 0.54  (0.50-0.67) | 0.75  (0.51-0.99) | 0.80  (0.54-0.99) | 0.80  (0.54-0.99) | 0.83  (0.57-0.99) | 1.28  (0.60-2.70) | 0.02  (0.01-0.05) | 1.64  (1.44-1.88) | 0.08  (0.00-0.19) |
| ssp585 | Beijing | 9.44  (0.27-19.38) | 0.64  (0.05-1.58) | 10.11  (0.53-19.52) | 0.68  (0.52-0.87) | -10.00  (-10.21--9.80) | 10.00  (9.81-10.19) | 10.00  (9.81-10.20) | 15.02  (14.83-15.22) | 0.58  (0.05-0.88) | 0.54  (0.40-0.70) | 0.45  (0.27-0.65) | 4.59  (3.06-7.02) | 0.75  (0.51-0.99) | 0.72  (0.51-0.98) | 0.74  (0.51-0.98) | 0.71  (0.51-0.98) | 0.75  (0.51-0.99) | 0.77  (0.52-0.99) | 0.79  (0.52-0.99) | 0.80  (0.53-0.99) | 1.99  (0.96-3.42) | 0.19  (0.03-0.36) | 2.11  (1.77-2.53) | 0.04  (0.00-0.14) |
| ssp585 | Guangdong | 9.72  (0.35-19.55) | 0.39  (0.03-1.08) | 9.95  (0.51-19.50) | 0.68  (0.52-0.87) | -10.00  (-10.19--9.81) | 9.99  (9.80-10.18) | 9.99  (9.81-10.19) | 14.99  (14.79-15.19) | 0.79  (0.48-0.90) | 0.63  (0.52-0.74) | 0.41  (0.20-0.65) | 4.84  (2.45-9.41) | 0.75  (0.51-0.99) | 0.76  (0.51-0.99) | 0.76  (0.52-0.99) | 0.54  (0.50-0.66) | 0.75  (0.51-0.99) | 0.88  (0.66-1.00) | 0.87  (0.59-1.00) | 0.94  (0.80-1.00) | 1.44  (0.42-3.11) | 0.13  (0.04-0.27) | 1.32  (1.18-1.49) | 0.10  (0.01-0.19) |
| ssp585 | Guizhou | 10.01  (0.53-19.51) | 0.62  (0.38-0.91) | 10.03  (0.42-19.49) | 0.68  (0.52-0.86) | -10.00  (-10.20--9.80) | 9.99  (9.80-10.18) | 10.00  (9.81-10.20) | 14.94  (14.73-15.14) | 0.87  (0.78-0.90) | 0.67  (0.57-0.78) | 0.43  (0.25-0.62) | 9.04  (7.01-9.97) | 0.75  (0.51-0.99) | 0.79  (0.52-0.99) | 0.79  (0.53-0.99) | 0.54  (0.50-0.65) | 0.75  (0.52-0.99) | 0.73  (0.51-0.99) | 0.72  (0.51-0.98) | 0.78  (0.53-0.99) | 0.74  (0.41-1.41) | 0.05  (0.02-0.09) | 1.48  (1.28-1.70) | 0.11  (0.01-0.19) |

**Table S4 Posterior summary of Europe by different SSPs and areas.**

| SSP | Area | $A_{n,0}$ | $F_{n,0}$ | $M_{n,0}$ | $\delta$ | $T_{1,\mu,a}$ | $T_{1,\mu,f}$ | $T_{1,\tau}$ | $T_{1,\rho}$ | $L_{\mu,a}$ | $L_{\mu,f}$ | $L_{\tau}$ | $L_{\rho}$ | $k_{1,\mu,a}$ | $k_{1,\mu,f}$ | $k_{1,\tau}$ | $k_{1,\rho}$ | $k_{2,\mu,a}$ | $k_{2,\mu,f}$ | $k_{2,\tau}$ | $k_{2,\rho}$ | $\omega$ | $\lambda$ | $\sigma$ | $\alpha$ |
| --- | --- | --- | --- | --- | --- | --- | --- | --- | --- | --- | --- | --- | --- | --- | --- | --- | --- | --- | --- | --- | --- | --- | --- | --- | --- |
| ssp126 | Autonomous Province of Trento | 37.47  (0.16-96.86) | 49.28  (2.65-96.69) | 50.04  (2.88-97.15) | 0.68  (0.52-0.87) | -10.00  (-10.21--9.80) | 9.98  (9.79-10.17) | 10.01  (9.81-10.19) | 14.81  (14.62-14.99) | 0.87  (0.81-0.90) | 0.63  (0.44-0.76) | 0.40  (0.16-0.60) | 9.30  (8.06-9.97) | 0.77  (0.52-0.99) | 0.68  (0.51-0.98) | 0.83  (0.55-0.99) | 0.51  (0.50-0.53) | 0.75  (0.52-0.99) | 0.75  (0.51-0.99) | 0.75  (0.51-0.99) | 0.75  (0.51-0.99) | 8.81  (3.76-29.23) | 2.64  (1.13-10.92) | 15.16  (14.26-16.11) | 0.10  (0.01-0.19) |
| ssp126 | Canton of Ticino | 0.01  (0.00-0.02) | 0.53  (0.01-2.36) | 49.67  (2.39-97.17) | 0.68  (0.52-0.88) | -10.01  (-10.19--9.82) | 9.98  (9.78-10.17) | 9.98  (9.78-10.17) | 14.82  (14.64-15.01) | 0.90  (0.88-0.90) | 0.55  (0.43-0.66) | 0.12  (0.10-0.19) | 7.66  (6.03-9.20) | 0.82  (0.55-0.99) | 0.86  (0.58-1.00) | 0.85  (0.60-0.99) | 0.52  (0.50-0.56) | 0.75  (0.51-0.99) | 0.75  (0.51-0.99) | 0.75  (0.51-0.99) | 0.74  (0.51-0.99) | 34.43  (26.17-39.75) | 6.71  (2.59-10.83) | 11.81  (11.07-12.64) | 0.06  (0.00-0.17) |
| ssp126 | Cote Azur | 48.31  (1.57-97.21) | 48.78  (2.51-96.78) | 49.59  (2.78-97.08) | 0.68  (0.52-0.87) | -10.00  (-10.19--9.80) | 10.01  (9.81-10.20) | 10.02  (9.83-10.22) | 15.07  (14.87-15.26) | 0.57  (0.10-0.88) | 0.58  (0.45-0.71) | 0.39  (0.20-0.58) | 8.67  (6.35-9.95) | 0.75  (0.51-0.99) | 0.67  (0.50-0.97) | 0.73  (0.51-0.99) | 0.55  (0.50-0.67) | 0.75  (0.51-0.99) | 0.77  (0.52-0.99) | 0.78  (0.53-0.99) | 0.78  (0.53-0.99) | 14.47  (7.21-28.99) | 0.08  (0.01-0.26) | 24.49  (22.56-26.72) | 0.06  (0.00-0.17) |
| ssp126 | Emilia-Romagna | 26.57  (0.14-94.99) | 42.28  (1.62-96.58) | 49.84  (3.11-97.20) | 0.68  (0.52-0.87) | -10.00  (-10.20--9.80) | 10.03  (9.82-10.23) | 10.04  (9.84-10.23) | 15.21  (15.01-15.40) | 0.57  (0.05-0.89) | 0.62  (0.50-0.72) | 0.50  (0.35-0.66) | 7.36  (4.98-9.81) | 0.75  (0.51-0.99) | 0.58  (0.50-0.86) | 0.82  (0.53-0.99) | 0.58  (0.50-0.78) | 0.75  (0.51-0.99) | 0.80  (0.54-0.99) | 0.82  (0.56-0.99) | 0.89  (0.69-1.00) | 34.84  (25.16-39.81) | 1.13  (0.11-2.60) | 53.41  (50.22-56.97) | 0.04  (0.00-0.13) |
| ssp126 | Fier | 49.29  (2.00-97.25) | 48.46  (2.51-97.46) | 50.08  (2.85-97.06) | 0.68  (0.52-0.87) | -10.00  (-10.20--9.81) | 10.00  (9.81-10.19) | 10.00  (9.81-10.19) | 15.02  (14.82-15.23) | 0.40  (0.02-0.86) | 0.68  (0.51-0.84) | 0.46  (0.27-0.66) | 3.49  (1.81-6.97) | 0.75  (0.51-0.99) | 0.73  (0.51-0.98) | 0.74  (0.51-0.98) | 0.70  (0.51-0.97) | 0.75  (0.51-0.99) | 0.77  (0.52-0.99) | 0.77  (0.52-0.99) | 0.75  (0.51-0.99) | 20.62  (4.91-38.41) | 0.62  (0.02-2.40) | 37.89  (32.19-44.71) | 0.08  (0.00-0.19) |
| ssp126 | Lazio | 49.66  (2.73-97.31) | 46.17  (2.26-96.80) | 49.88  (2.70-97.55) | 0.68  (0.53-0.88) | -10.00  (-10.20--9.81) | 10.01  (9.81-10.20) | 10.02  (9.83-10.21) | 15.16  (14.97-15.36) | 0.23  (0.01-0.65) | 0.57  (0.42-0.71) | 0.35  (0.19-0.57) | 6.74  (5.04-8.99) | 0.75  (0.52-0.99) | 0.61  (0.50-0.94) | 0.61  (0.50-0.94) | 0.53  (0.50-0.61) | 0.75  (0.51-0.98) | 0.81  (0.54-0.99) | 0.85  (0.59-1.00) | 0.85  (0.60-0.99) | 25.52  (13.09-37.87) | 0.05  (0.01-0.15) | 19.32  (17.55-21.31) | 0.04  (0.00-0.14) |
| ssp126 | Lezhe | 43.95  (0.39-96.98) | 48.60  (1.65-98.47) | 51.26  (2.49-97.07) | 0.68  (0.53-0.86) | -9.99  (-10.20--9.80) | 10.01  (9.81-10.20) | 10.00  (9.79-10.19) | 15.02  (14.82-15.24) | 0.26  (0.01-0.82) | 0.74  (0.58-0.87) | 0.54  (0.35-0.74) | 2.68  (1.27-6.12) | 0.75  (0.51-0.99) | 0.70  (0.50-0.98) | 0.74  (0.51-0.98) | 0.82  (0.56-0.99) | 0.74  (0.51-0.98) | 0.76  (0.52-0.99) | 0.76  (0.51-0.99) | 0.71  (0.51-0.98) | 23.95  (2.49-39.46) | 12.36  (2.26-19.57) | 25.47  (18.76-34.76) | 0.09  (0.01-0.19) |
| ssp126 | Lushnje | 49.50  (2.16-97.71) | 49.99  (2.20-97.94) | 50.09  (2.66-97.53) | 0.68  (0.52-0.86) | -10.00  (-10.20--9.80) | 10.00  (9.81-10.20) | 10.00  (9.80-10.20) | 15.02  (14.83-15.22) | 0.51  (0.04-0.89) | 0.63  (0.45-0.80) | 0.47  (0.29-0.66) | 6.64  (3.68-9.73) | 0.75  (0.51-0.99) | 0.73  (0.51-0.99) | 0.74  (0.51-0.99) | 0.72  (0.51-0.98) | 0.75  (0.51-0.99) | 0.73  (0.51-0.98) | 0.74  (0.51-0.99) | 0.72  (0.51-0.98) | 21.04  (8.20-37.47) | 2.00  (0.07-7.50) | 55.47  (46.04-67.27) | 0.06  (0.00-0.17) |
| ssp126 | Puglia | 49.84  (2.56-97.59) | 49.94  (5.11-97.09) | 49.58  (2.52-97.90) | 0.68  (0.52-0.87) | -10.00  (-10.20--9.79) | 9.99  (9.80-10.19) | 10.00  (9.79-10.19) | 15.02  (14.83-15.21) | 0.48  (0.03-0.88) | 0.65  (0.49-0.79) | 0.47  (0.26-0.67) | 4.71  (2.72-8.09) | 0.75  (0.51-0.99) | 0.72  (0.51-0.98) | 0.73  (0.51-0.99) | 0.72  (0.51-0.98) | 0.75  (0.51-0.99) | 0.82  (0.55-0.99) | 0.82  (0.55-0.99) | 0.83  (0.55-0.99) | 4.88  (1.57-10.30) | 0.09  (0.01-0.35) | 12.37  (11.42-13.44) | 0.08  (0.00-0.19) |
| ssp126 | Sicily | 49.53  (2.65-97.15) | 38.67  (4.20-89.71) | 50.14  (3.19-96.84) | 0.68  (0.52-0.87) | -10.00  (-10.19--9.81) | 9.99  (9.80-10.19) | 10.00  (9.80-10.19) | 15.03  (14.84-15.23) | 0.32  (0.01-0.78) | 0.65  (0.49-0.79) | 0.47  (0.29-0.65) | 2.42  (1.59-3.44) | 0.75  (0.51-0.99) | 0.73  (0.51-0.99) | 0.70  (0.51-0.98) | 0.65  (0.51-0.94) | 0.75  (0.51-0.99) | 0.79  (0.53-0.99) | 0.79  (0.53-0.99) | 0.75  (0.51-0.99) | 28.14  (15.46-38.84) | 0.26  (0.02-0.98) | 30.55  (28.44-32.90) | 0.07  (0.00-0.18) |
| ssp126 | Tirane | 48.11  (3.37-97.32) | 51.44  (2.82-97.94) | 49.87  (2.87-97.26) | 0.68  (0.52-0.88) | -10.00  (-10.20--9.80) | 10.00  (9.80-10.20) | 10.00  (9.80-10.20) | 15.06  (14.86-15.25) | 0.27  (0.01-0.72) | 0.68  (0.55-0.79) | 0.51  (0.33-0.69) | 5.91  (4.14-8.50) | 0.75  (0.51-0.99) | 0.69  (0.51-0.98) | 0.71  (0.51-0.98) | 0.62  (0.50-0.89) | 0.75  (0.51-0.99) | 0.82  (0.57-0.99) | 0.84  (0.56-0.99) | 0.84  (0.58-0.99) | 32.61  (19.67-39.77) | 0.29  (0.02-1.03) | 107.93  (100.13-116.84) | 0.07  (0.00-0.18) |
| ssp126 | Tuscany | 49.42  (1.58-97.96) | 49.68  (2.69-97.54) | 50.06  (2.41-97.35) | 0.68  (0.52-0.86) | -10.00  (-10.20--9.80) | 10.00  (9.81-10.20) | 10.01  (9.81-10.20) | 15.04  (14.85-15.23) | 0.57  (0.05-0.89) | 0.65  (0.51-0.78) | 0.46  (0.27-0.65) | 7.38  (4.47-9.80) | 0.75  (0.51-0.99) | 0.70  (0.51-0.98) | 0.74  (0.51-0.99) | 0.65  (0.51-0.94) | 0.75  (0.51-0.99) | 0.78  (0.52-0.99) | 0.78  (0.52-0.99) | 0.78  (0.52-0.99) | 14.31  (6.37-28.94) | 0.16  (0.01-0.62) | 24.90  (22.22-28.05) | 0.06  (0.00-0.18) |
| ssp126 | Veneto | 52.55  (6.97-97.96) | 51.36  (2.31-97.83) | 50.64  (2.82-97.51) | 0.68  (0.52-0.85) | -10.01  (-10.19--9.81) | 9.91  (9.71-10.11) | 9.98  (9.79-10.17) | 14.80  (14.59-15.01) | 0.89  (0.87-0.90) | 0.73  (0.60-0.85) | 0.51  (0.33-0.69) | 9.72  (8.97-9.99) | 0.80  (0.53-0.99) | 0.84  (0.55-0.99) | 0.89  (0.67-1.00) | 0.51  (0.50-0.52) | 0.75  (0.51-0.99) | 0.75  (0.51-0.99) | 0.75  (0.51-0.99) | 0.75  (0.51-0.99) | 8.42  (4.56-15.76) | 0.30  (0.02-0.95) | 13.73  (12.43-15.20) | 0.17  (0.09-0.20) |
| ssp126 | Vlore | 49.09  (1.78-97.37) | 49.49  (2.54-97.99) | 49.90  (3.05-97.22) | 0.68  (0.52-0.87) | -10.00  (-10.20--9.80) | 10.00  (9.80-10.20) | 10.00  (9.81-10.19) | 15.02  (14.83-15.23) | 0.40  (0.02-0.86) | 0.67  (0.50-0.83) | 0.46  (0.27-0.65) | 3.59  (2.03-6.23) | 0.75  (0.51-0.99) | 0.73  (0.51-0.99) | 0.74  (0.51-0.99) | 0.75  (0.52-0.99) | 0.75  (0.51-0.99) | 0.78  (0.53-0.99) | 0.78  (0.52-0.99) | 0.76  (0.52-0.99) | 20.61  (6.23-36.73) | 0.51  (0.02-2.01) | 35.74  (30.46-41.79) | 0.07  (0.00-0.18) |
| ssp370 | Autonomous Province of Trento | 38.24  (1.90-96.17) | 49.44  (2.10-97.40) | 50.00  (2.61-97.28) | 0.68  (0.52-0.86) | -10.01  (-10.22--9.81) | 9.99  (9.79-10.20) | 9.98  (9.78-10.18) | 14.83  (14.64-15.02) | 0.88  (0.81-0.90) | 0.57  (0.40-0.76) | 0.29  (0.14-0.57) | 8.16  (6.23-9.86) | 0.82  (0.54-0.99) | 0.72  (0.51-0.99) | 0.74  (0.51-0.98) | 0.51  (0.50-0.53) | 0.75  (0.51-0.99) | 0.75  (0.51-0.99) | 0.75  (0.51-0.99) | 0.75  (0.51-0.99) | 16.58  (3.77-32.04) | 7.48  (1.47-14.71) | 15.29  (14.38-16.29) | 0.06  (0.00-0.18) |
| ssp370 | Canton of Ticino | 13.03  (0.00-89.67) | 0.50  (0.00-2.93) | 49.89  (2.86-97.24) | 0.68  (0.52-0.87) | -10.00  (-10.19--9.81) | 10.00  (9.81-10.20) | 10.04  (9.84-10.24) | 14.90  (14.70-15.10) | 0.67  (0.00-0.90) | 0.61  (0.48-0.74) | 0.25  (0.10-0.69) | 8.14  (5.92-9.98) | 0.75  (0.51-0.99) | 0.69  (0.51-0.98) | 0.86  (0.56-1.00) | 0.53  (0.50-0.61) | 0.75  (0.51-0.99) | 0.75  (0.51-0.98) | 0.75  (0.51-0.98) | 0.75  (0.52-0.98) | 28.90  (4.32-39.87) | 0.94  (0.01-5.66) | 12.91  (11.82-14.56) | 0.07  (0.00-0.19) |
| ssp370 | Cote Azur | 48.58  (1.61-97.70) | 48.09  (2.05-97.83) | 50.14  (2.53-96.89) | 0.68  (0.52-0.86) | -10.00  (-10.20--9.80) | 10.00  (9.80-10.20) | 10.02  (9.82-10.21) | 15.05  (14.85-15.24) | 0.54  (0.07-0.88) | 0.57  (0.43-0.70) | 0.43  (0.24-0.62) | 8.45  (6.00-9.93) | 0.75  (0.51-0.99) | 0.68  (0.50-0.98) | 0.74  (0.51-0.99) | 0.54  (0.50-0.66) | 0.75  (0.51-0.99) | 0.76  (0.52-0.98) | 0.78  (0.52-0.99) | 0.77  (0.52-0.99) | 14.04  (6.96-28.34) | 0.07  (0.01-0.24) | 24.28  (22.28-26.45) | 0.05  (0.00-0.16) |
| ssp370 | Emilia-Romagna | 8.43  (0.06-80.99) | 30.09  (1.10-90.13) | 50.25  (2.48-97.48) | 0.68  (0.52-0.87) | -10.00  (-10.21--9.80) | 10.01  (9.82-10.21) | 10.02  (9.83-10.22) | 15.15  (14.95-15.34) | 0.80  (0.49-0.90) | 0.63  (0.51-0.72) | 0.52  (0.36-0.69) | 6.76  (5.23-9.56) | 0.75  (0.51-0.99) | 0.66  (0.50-0.97) | 0.87  (0.60-1.00) | 0.54  (0.50-0.64) | 0.75  (0.51-0.99) | 0.83  (0.57-0.99) | 0.85  (0.60-0.99) | 0.89  (0.69-1.00) | 33.56  (22.10-39.78) | 2.29  (0.74-4.11) | 53.21  (49.98-56.67) | 0.04  (0.00-0.14) |
| ssp370 | Fier | 49.47  (2.43-97.18) | 46.66  (1.70-97.71) | 50.00  (2.54-97.97) | 0.68  (0.52-0.87) | -10.00  (-10.21--9.79) | 10.01  (9.82-10.20) | 10.01  (9.81-10.20) | 15.03  (14.84-15.23) | 0.42  (0.02-0.87) | 0.68  (0.50-0.83) | 0.46  (0.28-0.65) | 3.48  (2.03-5.93) | 0.75  (0.51-0.99) | 0.70  (0.51-0.98) | 0.73  (0.51-0.98) | 0.76  (0.52-0.99) | 0.75  (0.51-0.99) | 0.76  (0.51-0.99) | 0.77  (0.52-0.99) | 0.76  (0.52-0.99) | 24.08  (8.07-39.13) | 1.31  (0.05-4.32) | 35.17  (29.93-41.86) | 0.07  (0.00-0.19) |
| ssp370 | Lazio | 50.15  (2.26-97.71) | 40.81  (1.42-95.72) | 49.73  (2.62-97.29) | 0.68  (0.52-0.87) | -10.00  (-10.20--9.80) | 10.02  (9.82-10.21) | 10.02  (9.83-10.21) | 15.16  (14.97-15.36) | 0.41  (0.03-0.83) | 0.57  (0.42-0.69) | 0.38  (0.20-0.60) | 7.41  (5.28-9.61) | 0.75  (0.51-0.99) | 0.57  (0.50-0.87) | 0.59  (0.50-0.89) | 0.57  (0.50-0.73) | 0.75  (0.51-0.99) | 0.79  (0.53-0.99) | 0.83  (0.57-0.99) | 0.81  (0.55-0.99) | 22.68  (11.74-37.59) | 0.08  (0.01-0.26) | 18.67  (17.04-20.51) | 0.04  (0.00-0.14) |
| ssp370 | Lezhe | 50.53  (3.11-97.53) | 49.48  (2.56-97.40) | 49.90  (3.20-97.54) | 0.68  (0.52-0.87) | -10.00  (-10.19--9.79) | 10.00  (9.81-10.20) | 10.01  (9.81-10.21) | 15.00  (14.80-15.22) | 0.51  (0.03-0.88) | 0.65  (0.49-0.81) | 0.50  (0.33-0.69) | 7.43  (3.36-9.93) | 0.75  (0.51-0.99) | 0.73  (0.51-0.99) | 0.75  (0.50-0.99) | 0.82  (0.56-0.99) | 0.75  (0.51-0.99) | 0.75  (0.51-0.99) | 0.74  (0.51-0.99) | 0.72  (0.51-0.98) | 3.81  (0.49-19.32) | 13.62  (7.66-19.65) | 17.54  (13.22-24.09) | 0.06  (0.00-0.18) |
| ssp370 | Lushnje | 50.53  (2.94-97.08) | 44.90  (2.04-96.41) | 50.22  (2.90-97.49) | 0.68  (0.52-0.87) | -10.00  (-10.19--9.81) | 10.00  (9.81-10.20) | 10.00  (9.81-10.19) | 15.02  (14.82-15.21) | 0.56  (0.06-0.89) | 0.65  (0.49-0.81) | 0.47  (0.28-0.66) | 7.21  (4.07-9.85) | 0.75  (0.52-0.99) | 0.72  (0.51-0.98) | 0.74  (0.51-0.99) | 0.80  (0.53-0.99) | 0.75  (0.51-0.99) | 0.74  (0.51-0.99) | 0.74  (0.51-0.99) | 0.75  (0.51-0.99) | 19.61  (7.69-36.93) | 2.38  (0.13-7.38) | 54.05  (44.41-65.96) | 0.07  (0.00-0.18) |
| ssp370 | Puglia | 50.99  (3.01-97.61) | 45.30  (3.05-96.51) | 50.88  (2.89-97.33) | 0.68  (0.52-0.86) | -10.00  (-10.19--9.80) | 10.00  (9.81-10.20) | 10.01  (9.81-10.21) | 15.03  (14.83-15.22) | 0.56  (0.04-0.89) | 0.67  (0.50-0.81) | 0.41  (0.19-0.62) | 6.05  (2.37-9.57) | 0.75  (0.51-0.99) | 0.70  (0.51-0.98) | 0.74  (0.51-0.99) | 0.76  (0.52-0.98) | 0.75  (0.51-0.99) | 0.83  (0.56-0.99) | 0.82  (0.54-0.99) | 0.83  (0.54-0.99) | 4.95  (1.32-19.93) | 0.11  (0.01-0.58) | 12.51  (11.53-13.64) | 0.09  (0.00-0.19) |
| ssp370 | Sicily | 50.23  (2.51-97.81) | 32.55  (2.58-86.38) | 49.98  (2.87-97.27) | 0.68  (0.52-0.87) | -10.00  (-10.19--9.81) | 9.99  (9.80-10.19) | 9.99  (9.78-10.19) | 15.03  (14.84-15.22) | 0.41  (0.02-0.86) | 0.65  (0.51-0.79) | 0.52  (0.35-0.72) | 2.31  (1.54-3.26) | 0.75  (0.51-0.99) | 0.78  (0.51-0.99) | 0.68  (0.51-0.97) | 0.64  (0.51-0.93) | 0.75  (0.51-0.98) | 0.80  (0.53-0.99) | 0.80  (0.53-0.99) | 0.77  (0.52-0.99) | 28.41  (16.61-38.94) | 0.28  (0.02-1.02) | 30.15  (28.08-32.55) | 0.07  (0.00-0.19) |
| ssp370 | Tirane | 49.41  (2.49-96.95) | 50.20  (2.73-97.59) | 50.25  (2.21-97.34) | 0.68  (0.52-0.87) | -10.00  (-10.19--9.81) | 10.01  (9.81-10.20) | 10.00  (9.80-10.21) | 15.04  (14.85-15.24) | 0.37  (0.02-0.83) | 0.68  (0.55-0.80) | 0.50  (0.33-0.67) | 6.81  (4.59-9.50) | 0.75  (0.51-0.99) | 0.69  (0.50-0.98) | 0.74  (0.51-0.99) | 0.67  (0.51-0.96) | 0.75  (0.51-0.99) | 0.81  (0.55-0.99) | 0.82  (0.54-0.99) | 0.85  (0.59-0.99) | 31.23  (17.41-39.62) | 0.35  (0.02-1.28) | 104.67  (97.03-112.98) | 0.07  (0.00-0.18) |
| ssp370 | Tuscany | 50.03  (2.46-97.41) | 35.90  (1.48-91.68) | 50.12  (2.73-97.18) | 0.68  (0.52-0.87) | -10.00  (-10.20--9.80) | 10.00  (9.81-10.20) | 10.01  (9.81-10.20) | 15.01  (14.81-15.21) | 0.71  (0.21-0.89) | 0.66  (0.51-0.80) | 0.46  (0.26-0.65) | 8.24  (5.55-9.93) | 0.75  (0.51-0.98) | 0.68  (0.51-0.98) | 0.75  (0.51-0.99) | 0.60  (0.50-0.83) | 0.75  (0.51-0.99) | 0.77  (0.52-0.99) | 0.77  (0.52-0.99) | 0.74  (0.51-0.99) | 12.61  (6.04-25.24) | 0.57  (0.03-2.00) | 25.31  (22.65-28.35) | 0.07  (0.00-0.18) |
| ssp370 | Veneto | 53.10  (7.54-97.61) | 50.30  (2.13-97.67) | 50.20  (2.70-97.42) | 0.68  (0.52-0.87) | -10.00  (-10.20--9.81) | 9.97  (9.77-10.17) | 9.98  (9.79-10.17) | 14.86  (14.67-15.06) | 0.88  (0.84-0.90) | 0.68  (0.53-0.82) | 0.47  (0.30-0.65) | 9.53  (8.41-9.99) | 0.78  (0.52-0.99) | 0.76  (0.51-0.99) | 0.82  (0.56-0.99) | 0.51  (0.50-0.54) | 0.75  (0.51-0.99) | 0.75  (0.51-0.99) | 0.75  (0.51-0.99) | 0.75  (0.51-0.99) | 9.92  (4.74-18.55) | 0.32  (0.02-1.00) | 13.91  (12.66-15.36) | 0.13  (0.02-0.20) |
| ssp370 | Vlore | 49.14  (1.97-97.53) | 45.46  (2.05-96.98) | 49.71  (1.80-97.52) | 0.68  (0.52-0.86) | -10.00  (-10.19--9.80) | 10.00  (9.81-10.20) | 10.00  (9.80-10.20) | 15.02  (14.84-15.22) | 0.43  (0.03-0.87) | 0.71  (0.54-0.86) | 0.46  (0.27-0.67) | 3.73  (1.99-6.51) | 0.75  (0.51-0.99) | 0.72  (0.51-0.98) | 0.74  (0.51-0.99) | 0.72  (0.51-0.98) | 0.75  (0.51-0.99) | 0.78  (0.52-0.99) | 0.77  (0.52-0.99) | 0.74  (0.51-0.99) | 21.06  (6.55-37.60) | 0.84  (0.03-3.18) | 32.58  (27.81-38.49) | 0.08  (0.00-0.19) |
| ssp585 | Autonomous Province of Trento | 1.39  (0.03-3.93) | 44.02  (2.45-95.15) | 50.11  (2.25-97.40) | 0.68  (0.52-0.87) | -10.01  (-10.20--9.82) | 9.99  (9.80-10.18) | 9.99  (9.79-10.19) | 14.91  (14.73-15.10) | 0.87  (0.79-0.90) | 0.64  (0.47-0.78) | 0.39  (0.21-0.60) | 8.20  (6.51-9.86) | 0.81  (0.53-0.99) | 0.62  (0.50-0.95) | 0.62  (0.50-0.94) | 0.51  (0.50-0.54) | 0.75  (0.51-0.99) | 0.75  (0.52-0.99) | 0.75  (0.51-0.99) | 0.75  (0.51-0.99) | 17.49  (8.45-34.29) | 0.47  (0.03-1.46) | 15.01  (14.19-15.89) | 0.08  (0.00-0.19) |
| ssp585 | Canton of Ticino | 0.00  (0.00-0.02) | 0.24  (0.00-1.24) | 49.90  (3.25-97.05) | 0.68  (0.52-0.88) | -10.00  (-10.19--9.80) | 9.97  (9.78-10.16) | 9.97  (9.77-10.17) | 14.89  (14.69-15.09) | 0.89  (0.87-0.90) | 0.57  (0.42-0.72) | 0.13  (0.10-0.23) | 8.72  (6.92-9.92) | 0.74  (0.51-0.99) | 0.79  (0.51-0.99) | 0.78  (0.51-0.99) | 0.51  (0.50-0.55) | 0.75  (0.51-0.99) | 0.75  (0.51-0.99) | 0.75  (0.52-0.99) | 0.75  (0.52-0.99) | 31.77  (12.78-39.69) | 2.38  (0.13-6.49) | 12.66  (11.86-13.52) | 0.09  (0.00-0.19) |
| ssp585 | Cote Azur | 49.15  (1.80-98.01) | 49.79  (2.66-97.72) | 49.50  (2.88-96.64) | 0.68  (0.52-0.87) | -10.00  (-10.19--9.81) | 10.00  (9.81-10.20) | 10.01  (9.81-10.21) | 15.07  (14.88-15.27) | 0.60  (0.08-0.88) | 0.58  (0.44-0.70) | 0.46  (0.28-0.65) | 7.60  (5.13-9.83) | 0.75  (0.51-0.99) | 0.70  (0.51-0.98) | 0.77  (0.52-0.99) | 0.57  (0.50-0.77) | 0.75  (0.51-0.99) | 0.76  (0.52-0.99) | 0.77  (0.52-0.99) | 0.79  (0.54-0.99) | 14.10  (7.42-26.02) | 0.10  (0.01-0.32) | 24.14  (22.17-26.27) | 0.05  (0.00-0.14) |
| ssp585 | Emilia-Romagna | 21.58  (0.09-93.68) | 39.03  (1.55-95.74) | 49.89  (2.69-97.31) | 0.68  (0.52-0.87) | -10.00  (-10.19--9.81) | 10.01  (9.83-10.21) | 10.00  (9.81-10.20) | 15.13  (14.94-15.32) | 0.63  (0.06-0.89) | 0.60  (0.48-0.70) | 0.56  (0.41-0.72) | 7.14  (4.97-9.78) | 0.75  (0.51-0.99) | 0.66  (0.50-0.97) | 0.75  (0.51-0.99) | 0.57  (0.50-0.74) | 0.75  (0.51-0.99) | 0.81  (0.55-0.99) | 0.83  (0.57-0.99) | 0.88  (0.67-1.00) | 34.27  (23.02-39.83) | 0.51  (0.03-1.58) | 53.87  (50.53-57.47) | 0.04  (0.00-0.14) |
| ssp585 | Fier | 49.84  (2.66-97.22) | 47.56  (1.77-96.76) | 50.22  (2.49-96.88) | 0.68  (0.52-0.87) | -10.00  (-10.20--9.81) | 10.00  (9.81-10.20) | 10.00  (9.80-10.20) | 15.01  (14.82-15.20) | 0.45  (0.03-0.88) | 0.71  (0.54-0.85) | 0.45  (0.26-0.65) | 4.08  (2.10-7.81) | 0.75  (0.51-0.99) | 0.76  (0.51-0.99) | 0.75  (0.51-0.99) | 0.68  (0.51-0.97) | 0.75  (0.51-0.99) | 0.75  (0.51-0.99) | 0.76  (0.51-0.99) | 0.76  (0.52-0.99) | 17.32  (4.00-36.57) | 1.98  (0.04-15.80) | 36.77  (31.31-43.24) | 0.08  (0.00-0.19) |
| ssp585 | Lazio | 50.47  (2.75-97.52) | 39.08  (1.72-93.55) | 50.25  (2.87-97.53) | 0.68  (0.52-0.87) | -10.00  (-10.20--9.81) | 10.00  (9.81-10.20) | 10.01  (9.81-10.20) | 15.14  (14.93-15.33) | 0.32  (0.01-0.78) | 0.54  (0.41-0.66) | 0.44  (0.26-0.63) | 6.36  (4.76-7.97) | 0.75  (0.51-0.99) | 0.67  (0.50-0.97) | 0.65  (0.50-0.97) | 0.52  (0.50-0.59) | 0.75  (0.51-0.99) | 0.83  (0.58-0.99) | 0.88  (0.66-1.00) | 0.88  (0.66-1.00) | 23.23  (14.44-35.77) | 0.06  (0.01-0.21) | 18.34  (16.68-20.25) | 0.04  (0.00-0.14) |
| ssp585 | Lezhe | 48.46  (1.66-96.40) | 50.02  (2.30-97.80) | 49.70  (2.24-97.26) | 0.68  (0.53-0.87) | -10.00  (-10.20--9.80) | 10.00  (9.81-10.19) | 10.00  (9.81-10.19) | 15.00  (14.80-15.20) | 0.36  (0.01-0.85) | 0.80  (0.65-0.90) | 0.50  (0.30-0.69) | 2.27  (1.11-5.22) | 0.75  (0.51-0.99) | 0.75  (0.51-0.99) | 0.74  (0.51-0.99) | 0.79  (0.52-0.99) | 0.75  (0.51-0.99) | 0.75  (0.51-0.99) | 0.74  (0.51-0.99) | 0.75  (0.51-0.99) | 20.89  (0.95-39.30) | 11.30  (1.00-19.65) | 24.89  (18.30-34.03) | 0.12  (0.01-0.20) |
| ssp585 | Lushnje | 49.92  (2.64-97.28) | 49.50  (2.59-97.41) | 49.78  (2.70-97.11) | 0.68  (0.52-0.87) | -10.00  (-10.20--9.81) | 10.00  (9.80-10.19) | 10.01  (9.81-10.19) | 14.99  (14.79-15.18) | 0.56  (0.05-0.89) | 0.69  (0.52-0.85) | 0.46  (0.27-0.65) | 7.63  (4.28-9.90) | 0.75  (0.51-0.99) | 0.78  (0.52-0.99) | 0.76  (0.51-0.99) | 0.69  (0.51-0.97) | 0.75  (0.51-0.99) | 0.75  (0.51-0.99) | 0.75  (0.51-0.99) | 0.75  (0.51-0.99) | 16.40  (4.43-34.72) | 1.72  (0.07-6.36) | 53.88  (44.49-65.64) | 0.08  (0.00-0.19) |
| ssp585 | Puglia | 49.44  (2.73-96.82) | 47.73  (5.07-96.41) | 49.81  (2.21-97.40) | 0.68  (0.52-0.87) | -10.00  (-10.20--9.80) | 10.00  (9.80-10.19) | 10.00  (9.81-10.20) | 15.01  (14.82-15.21) | 0.53  (0.05-0.89) | 0.66  (0.50-0.80) | 0.45  (0.25-0.65) | 5.26  (2.88-8.67) | 0.75  (0.51-0.99) | 0.73  (0.51-0.98) | 0.75  (0.51-0.99) | 0.74  (0.52-0.98) | 0.75  (0.51-0.99) | 0.83  (0.54-0.99) | 0.82  (0.54-0.99) | 0.83  (0.54-0.99) | 4.47  (1.65-9.59) | 0.15  (0.01-0.62) | 12.44  (11.47-13.49) | 0.08  (0.00-0.19) |
| ssp585 | Sicily | 50.48  (2.45-97.64) | 33.11  (2.41-84.01) | 49.97  (2.76-97.43) | 0.68  (0.52-0.87) | -10.00  (-10.20--9.81) | 10.00  (9.82-10.19) | 10.00  (9.80-10.20) | 15.03  (14.84-15.21) | 0.34  (0.01-0.84) | 0.66  (0.51-0.82) | 0.41  (0.25-0.59) | 2.69  (1.69-3.96) | 0.75  (0.51-0.99) | 0.70  (0.51-0.98) | 0.71  (0.51-0.98) | 0.67  (0.51-0.96) | 0.75  (0.51-0.99) | 0.78  (0.53-0.99) | 0.78  (0.52-0.99) | 0.76  (0.51-0.99) | 27.21  (9.79-39.46) | 0.29  (0.02-1.01) | 30.93  (28.84-33.23) | 0.07  (0.00-0.18) |
| ssp585 | Tirane | 49.88  (2.69-97.49) | 49.83  (3.24-97.01) | 49.87  (2.79-97.11) | 0.68  (0.52-0.87) | -10.00  (-10.19--9.81) | 10.01  (9.81-10.20) | 10.01  (9.81-10.21) | 15.05  (14.86-15.25) | 0.28  (0.01-0.76) | 0.67  (0.53-0.79) | 0.48  (0.32-0.65) | 6.97  (4.80-9.50) | 0.75  (0.52-0.98) | 0.68  (0.50-0.98) | 0.71  (0.51-0.98) | 0.62  (0.50-0.90) | 0.76  (0.51-0.99) | 0.84  (0.59-0.99) | 0.85  (0.59-1.00) | 0.88  (0.64-1.00) | 31.55  (18.21-39.69) | 0.35  (0.02-1.26) | 106.33  (98.50-115.04) | 0.07  (0.00-0.18) |
| ssp585 | Tuscany | 49.22  (1.85-96.96) | 49.47  (2.69-97.43) | 49.93  (2.77-96.91) | 0.68  (0.52-0.87) | -10.00  (-10.20--9.80) | 10.00  (9.80-10.20) | 10.01  (9.81-10.20) | 15.05  (14.86-15.24) | 0.69  (0.20-0.89) | 0.72  (0.59-0.83) | 0.46  (0.26-0.65) | 8.19  (5.60-9.93) | 0.75  (0.51-0.99) | 0.72  (0.51-0.98) | 0.77  (0.52-0.99) | 0.66  (0.51-0.92) | 0.75  (0.51-0.99) | 0.74  (0.51-0.99) | 0.74  (0.51-0.98) | 0.77  (0.52-0.99) | 6.11  (2.49-13.01) | 2.75  (1.15-5.40) | 21.97  (19.60-24.68) | 0.09  (0.00-0.19) |
| ssp585 | Veneto | 53.74  (9.02-98.32) | 49.95  (2.36-97.44) | 49.82  (2.13-97.48) | 0.68  (0.52-0.87) | -10.01  (-10.20--9.81) | 9.96  (9.76-10.15) | 10.01  (9.82-10.20) | 14.79  (14.60-14.99) | 0.89  (0.88-0.90) | 0.61  (0.47-0.75) | 0.48  (0.32-0.65) | 9.75  (9.12-9.99) | 0.82  (0.54-0.99) | 0.77  (0.51-0.99) | 0.91  (0.73-1.00) | 0.51  (0.50-0.53) | 0.75  (0.51-0.99) | 0.75  (0.51-0.99) | 0.75  (0.51-0.99) | 0.75  (0.52-0.99) | 13.87  (7.85-23.50) | 0.34  (0.03-1.08) | 12.69  (11.50-14.05) | 0.12  (0.01-0.20) |
| ssp585 | Vlore | 49.77  (2.71-97.43) | 50.47  (2.72-97.86) | 50.20  (2.77-97.23) | 0.68  (0.52-0.86) | -10.00  (-10.20--9.80) | 10.00  (9.79-10.20) | 10.01  (9.82-10.20) | 15.02  (14.83-15.22) | 0.49  (0.04-0.88) | 0.72  (0.57-0.85) | 0.42  (0.24-0.61) | 3.73  (2.35-5.94) | 0.75  (0.52-0.99) | 0.76  (0.51-0.99) | 0.76  (0.51-0.99) | 0.60  (0.50-0.85) | 0.75  (0.51-0.99) | 0.76  (0.52-0.99) | 0.76  (0.51-0.99) | 0.78  (0.52-0.99) | 9.66  (0.90-25.00) | 14.88  (0.85-19.86) | 33.10  (28.21-39.25) | 0.09  (0.00-0.19) |

**Table S5 Posterior summary of the United States by different SSPs and areas.**

| SSP | Area | $A_{n,0}$ | $F_{n,0}$ | $M_{n,0}$ | $\delta$ | $T_{1,\mu,a}$ | $T_{1,\mu,f}$ | $T_{1,\tau}$ | $T_{1,\rho}$ | $L_{\mu,a}$ | $L_{\mu,f}$ | $L_{\tau}$ | $L_{\rho}$ | $k_{1,\mu,a}$ | $k_{1,\mu,f}$ | $k_{1,\tau}$ | $k_{1,\rho}$ | $k_{2,\mu,a}$ | $k_{2,\mu,f}$ | $k_{2,\tau}$ | $k_{2,\rho}$ | $\omega$ | $\lambda$ | $\sigma$ | $\alpha$ |
| --- | --- | --- | --- | --- | --- | --- | --- | --- | --- | --- | --- | --- | --- | --- | --- | --- | --- | --- | --- | --- | --- | --- | --- | --- | --- |
| ssp126 | Connecticut | 48.38  (0.33-97.94) | 0.05  (0.00-0.19) | 49.29  (2.73-96.49) | 0.68  (0.52-0.86) | -10.00  (-10.20--9.81) | 10.00  (9.81-10.20) | 10.01  (9.82-10.21) | 15.01  (14.81-15.20) | 0.41  (0.00-0.88) | 0.64  (0.49-0.79) | 0.38  (0.22-0.58) | 7.80  (4.95-9.84) | 0.75  (0.51-0.99) | 0.71  (0.51-0.98) | 0.74  (0.51-0.98) | 0.76  (0.51-0.99) | 0.75  (0.51-0.99) | 0.74  (0.51-0.99) | 0.76  (0.52-0.99) | 0.75  (0.51-0.98) | 0.10  (0.02-0.27) | 0.14  (0.05-0.29) | 0.26  (0.24-0.29) | 0.06  (0.00-0.18) |
| ssp126 | Georgia | 50.54  (2.32-97.53) | 0.55  (0.02-2.01) | 50.23  (2.50-97.45) | 0.68  (0.52-0.86) | -10.00  (-10.20--9.80) | 10.03  (9.83-10.23) | 10.01  (9.81-10.21) | 15.00  (14.81-15.20) | 0.39  (0.03-0.83) | 0.75  (0.64-0.83) | 0.63  (0.50-0.77) | 7.10  (4.59-9.71) | 0.75  (0.51-0.99) | 0.56  (0.50-0.80) | 0.71  (0.51-0.98) | 0.74  (0.51-0.99) | 0.75  (0.52-0.99) | 0.89  (0.69-1.00) | 0.88  (0.64-1.00) | 0.75  (0.51-0.99) | 0.80  (0.43-0.99) | 0.27  (0.20-0.30) | 1.78  (1.66-1.91) | 0.07  (0.00-0.18) |
| ssp126 | New Jersey | 49.36  (2.36-97.42) | 0.38  (0.02-1.05) | 50.32  (2.68-97.49) | 0.68  (0.52-0.86) | -10.00  (-10.20--9.81) | 10.00  (9.81-10.20) | 10.01  (9.81-10.21) | 15.00  (14.81-15.20) | 0.45  (0.03-0.87) | 0.66  (0.51-0.79) | 0.48  (0.31-0.67) | 6.70  (3.64-9.76) | 0.75  (0.51-0.99) | 0.69  (0.51-0.98) | 0.74  (0.51-0.99) | 0.77  (0.52-0.99) | 0.75  (0.51-0.99) | 0.77  (0.52-0.99) | 0.78  (0.52-0.99) | 0.75  (0.51-0.99) | 0.67  (0.23-0.99) | 0.09  (0.01-0.25) | 0.60  (0.52-0.71) | 0.07  (0.00-0.18) |
| ssp126 | New York | 49.38  (1.66-97.60) | 0.23  (0.01-0.71) | 49.61  (2.68-97.35) | 0.68  (0.52-0.87) | -10.00  (-10.20--9.81) | 10.00  (9.81-10.20) | 9.99  (9.81-10.19) | 15.00  (14.82-15.20) | 0.73  (0.28-0.90) | 0.73  (0.61-0.85) | 0.55  (0.40-0.73) | 8.13  (5.16-9.92) | 0.76  (0.51-0.99) | 0.71  (0.51-0.98) | 0.71  (0.51-0.97) | 0.76  (0.52-0.98) | 0.75  (0.51-0.99) | 0.74  (0.51-0.98) | 0.74  (0.51-0.99) | 0.75  (0.51-0.99) | 0.80  (0.48-0.99) | 0.04  (0.01-0.11) | 0.59  (0.54-0.65) | 0.09  (0.00-0.19) |
| ssp126 | North Carolina | 50.13  (2.83-97.05) | 35.02  (21.24-50.24) | 49.22  (2.31-97.31) | 0.68  (0.52-0.87) | -10.00  (-10.19--9.80) | 10.01  (9.81-10.21) | 9.97  (9.77-10.16) | 14.98  (14.79-15.17) | 0.08  (0.00-0.23) | 0.88  (0.86-0.90) | 0.84  (0.66-0.99) | 1.36  (1.06-1.66) | 0.75  (0.51-0.99) | 0.53  (0.50-0.64) | 0.65  (0.50-0.96) | 0.92  (0.70-1.00) | 0.75  (0.51-0.99) | 0.81  (0.55-0.99) | 0.76  (0.51-0.99) | 0.73  (0.51-0.98) | 0.80  (0.40-0.99) | 0.28  (0.21-0.30) | 11.53  (10.03-13.29) | 0.19  (0.18-0.20) |
| ssp370 | Connecticut | 38.66  (0.00-97.05) | 0.06  (0.00-0.28) | 52.13  (2.53-98.05) | 0.67  (0.52-0.86) | -10.00  (-10.18--9.81) | 10.00  (9.82-10.18) | 10.00  (9.83-10.19) | 14.99  (14.81-15.19) | 0.55  (0.03-0.90) | 0.62  (0.47-0.76) | 0.42  (0.21-0.65) | 5.95  (1.27-9.74) | 0.76  (0.51-1.00) | 0.71  (0.51-0.98) | 0.73  (0.51-0.98) | 0.75  (0.51-0.99) | 0.74  (0.51-0.98) | 0.77  (0.52-0.99) | 0.76  (0.52-0.99) | 0.74  (0.52-0.99) | 0.35  (0.05-0.95) | 0.07  (0.02-0.24) | 0.26  (0.21-0.30) | 0.07  (0.00-0.17) |
| ssp370 | Georgia | 49.96  (2.44-97.60) | 0.76  (0.02-2.25) | 50.89  (2.45-97.17) | 0.68  (0.52-0.86) | -10.00  (-10.20--9.80) | 10.04  (9.84-10.24) | 10.01  (9.82-10.20) | 14.99  (14.80-15.18) | 0.59  (0.12-0.89) | 0.77  (0.66-0.87) | 0.69  (0.54-0.85) | 8.04  (5.05-9.93) | 0.75  (0.51-0.99) | 0.54  (0.50-0.66) | 0.69  (0.51-0.98) | 0.73  (0.51-0.98) | 0.75  (0.51-0.99) | 0.90  (0.71-1.00) | 0.87  (0.63-1.00) | 0.75  (0.51-0.99) | 0.94  (0.80-1.00) | 0.09  (0.02-0.23) | 1.86  (1.74-2.00) | 0.09  (0.00-0.19) |
| ssp370 | New Jersey | 49.89  (2.31-97.47) | 0.30  (0.01-0.89) | 49.13  (2.29-97.34) | 0.68  (0.52-0.88) | -10.00  (-10.20--9.81) | 10.01  (9.82-10.20) | 10.00  (9.81-10.20) | 15.01  (14.80-15.20) | 0.46  (0.04-0.87) | 0.69  (0.55-0.83) | 0.49  (0.31-0.67) | 6.31  (2.95-9.78) | 0.75  (0.51-0.99) | 0.69  (0.51-0.98) | 0.74  (0.51-0.99) | 0.76  (0.51-0.99) | 0.75  (0.51-0.99) | 0.77  (0.52-0.99) | 0.77  (0.52-0.99) | 0.75  (0.51-0.99) | 0.72  (0.30-0.99) | 0.04  (0.01-0.13) | 0.61  (0.52-0.71) | 0.07  (0.00-0.18) |
| ssp370 | New York | 49.95  (1.93-97.53) | 0.25  (0.01-0.79) | 49.53  (2.20-96.95) | 0.68  (0.52-0.86) | -10.00  (-10.19--9.81) | 10.01  (9.80-10.21) | 10.01  (9.81-10.20) | 14.99  (14.79-15.18) | 0.74  (0.35-0.90) | 0.69  (0.58-0.80) | 0.57  (0.43-0.73) | 8.64  (6.02-9.95) | 0.76  (0.51-0.99) | 0.69  (0.51-0.98) | 0.77  (0.52-0.99) | 0.71  (0.51-0.98) | 0.75  (0.52-0.99) | 0.75  (0.51-0.99) | 0.75  (0.51-0.99) | 0.75  (0.51-0.99) | 0.88  (0.65-1.00) | 0.02  (0.01-0.06) | 0.62  (0.57-0.67) | 0.08  (0.00-0.19) |
| ssp370 | North Carolina | 50.45  (1.85-97.76) | 27.64  (8.77-48.33) | 49.73  (2.29-97.75) | 0.68  (0.53-0.87) | -10.00  (-10.19--9.80) | 9.99  (9.80-10.19) | 10.00  (9.80-10.19) | 14.98  (14.78-15.17) | 0.66  (0.02-0.90) | 0.89  (0.86-0.90) | 0.65  (0.47-0.88) | 3.99  (1.07-8.96) | 0.75  (0.51-0.99) | 0.70  (0.51-0.98) | 0.76  (0.52-0.99) | 0.69  (0.51-0.98) | 0.75  (0.51-0.99) | 0.79  (0.53-0.99) | 0.73  (0.51-0.99) | 0.75  (0.51-0.99) | 0.23  (0.01-0.97) | 0.26  (0.14-0.30) | 14.33  (12.34-16.67) | 0.19  (0.17-0.20) |
| ssp585 | Connecticut | 50.46  (1.86-98.04) | 0.08  (0.00-0.30) | 50.48  (2.22-97.82) | 0.68  (0.52-0.87) | -10.00  (-10.20--9.80) | 10.00  (9.80-10.19) | 10.00  (9.81-10.21) | 15.00  (14.80-15.20) | 0.49  (0.04-0.88) | 0.66  (0.50-0.82) | 0.42  (0.22-0.62) | 7.84  (4.81-9.90) | 0.75  (0.51-0.99) | 0.73  (0.51-0.98) | 0.73  (0.51-0.99) | 0.70  (0.51-0.98) | 0.76  (0.51-0.99) | 0.75  (0.51-0.99) | 0.75  (0.51-0.99) | 0.75  (0.51-0.99) | 0.29  (0.10-0.68) | 0.02  (0.01-0.06) | 0.28  (0.26-0.31) | 0.08  (0.00-0.19) |
| ssp585 | Georgia | 50.09  (2.66-96.95) | 0.83  (0.03-2.40) | 50.04  (2.69-97.47) | 0.68  (0.52-0.87) | -10.00  (-10.20--9.81) | 10.03  (9.83-10.23) | 10.01  (9.81-10.21) | 14.99  (14.79-15.18) | 0.67  (0.21-0.89) | 0.77  (0.67-0.87) | 0.72  (0.57-0.88) | 8.54  (5.87-9.95) | 0.75  (0.51-0.99) | 0.55  (0.50-0.72) | 0.72  (0.51-0.99) | 0.67  (0.51-0.96) | 0.75  (0.51-0.98) | 0.93  (0.78-1.00) | 0.89  (0.67-1.00) | 0.75  (0.51-0.99) | 0.95  (0.83-1.00) | 0.05  (0.01-0.14) | 1.91  (1.79-2.06) | 0.09  (0.01-0.19) |
| ssp585 | New Jersey | 42.84  (0.12-96.88) | 0.13  (0.00-0.47) | 48.89  (2.98-96.99) | 0.68  (0.52-0.88) | -10.00  (-10.19--9.80) | 10.00  (9.81-10.19) | 10.00  (9.81-10.19) | 15.00  (14.81-15.20) | 0.51  (0.04-0.88) | 0.61  (0.49-0.71) | 0.63  (0.48-0.77) | 5.64  (3.36-9.33) | 0.75  (0.51-0.99) | 0.72  (0.51-0.98) | 0.76  (0.51-0.99) | 0.81  (0.53-0.99) | 0.75  (0.51-0.99) | 0.76  (0.52-0.99) | 0.79  (0.53-0.99) | 0.75  (0.51-0.99) | 0.17  (0.02-0.46) | 0.27  (0.21-0.30) | 0.45  (0.38-0.54) | 0.05  (0.00-0.17) |
| ssp585 | New York | 49.74  (2.17-97.83) | 0.25  (0.01-0.80) | 49.99  (2.10-97.99) | 0.68  (0.52-0.87) | -10.00  (-10.20--9.80) | 10.00  (9.81-10.20) | 10.01  (9.81-10.21) | 14.98  (14.78-15.18) | 0.75  (0.40-0.90) | 0.72  (0.60-0.84) | 0.53  (0.39-0.67) | 8.98  (6.84-9.98) | 0.75  (0.51-0.99) | 0.63  (0.50-0.94) | 0.68  (0.51-0.98) | 0.71  (0.51-0.98) | 0.75  (0.51-0.99) | 0.75  (0.51-0.98) | 0.76  (0.52-0.99) | 0.75  (0.51-0.99) | 0.87  (0.63-1.00) | 0.03  (0.01-0.07) | 0.61  (0.56-0.66) | 0.10  (0.01-0.19) |
| ssp585 | North Carolina | 50.20  (2.00-98.08) | 28.63  (12.28-48.87) | 50.58  (2.76-96.80) | 0.68  (0.52-0.87) | -10.00  (-10.18--9.81) | 9.98  (9.79-10.17) | 9.99  (9.80-10.17) | 14.98  (14.78-15.17) | 0.44  (0.01-0.88) | 0.89  (0.86-0.90) | 0.64  (0.48-0.81) | 7.84  (4.20-9.89) | 0.75  (0.51-0.99) | 0.58  (0.50-0.89) | 0.73  (0.51-0.99) | 0.59  (0.50-0.90) | 0.75  (0.51-0.99) | 0.80  (0.54-0.99) | 0.75  (0.51-0.99) | 0.75  (0.52-0.99) | 0.10  (0.01-0.73) | 0.27  (0.20-0.30) | 13.48  (11.80-15.54) | 0.19  (0.17-0.20) |
| ssp126 | Arizona | 0.18  (0.04-0.42) | 0.36  (0.01-1.01) | 5.01  (0.22-9.78) | 0.68  (0.52-0.87) | 15.00  (14.80-15.20) | 14.93  (14.74-15.12) | 15.00  (14.80-15.20) | 24.98  (24.78-25.18) | 0.47  (0.02-0.89) | 0.85  (0.78-0.90) | 0.90  (0.74-1.00) | 8.69  (6.47-9.95) | 0.75  (0.51-0.98) | 0.87  (0.66-0.99) | 0.82  (0.54-0.99) | 0.57  (0.50-0.74) | 0.75  (0.51-0.99) | 0.74  (0.51-0.99) | 0.75  (0.51-0.99) | 0.75  (0.51-0.99) | 0.37  (0.19-0.65) | 0.04  (0.01-0.09) | 0.43  (0.37-0.49) | 0.18  (0.14-0.20) |
| ssp126 | California | 5.11  (0.35-9.78) | 0.02  (0.00-0.05) | 5.03  (0.31-9.73) | 0.67  (0.52-0.86) | 15.00  (14.81-15.20) | 14.86  (14.67-15.04) | 15.04  (14.85-15.23) | 24.99  (24.79-25.19) | 0.45  (0.03-0.88) | 0.82  (0.74-0.89) | 0.84  (0.70-0.97) | 6.10  (4.64-8.04) | 0.84  (0.56-0.99) | 0.77  (0.51-0.99) | 0.94  (0.79-1.00) | 0.52  (0.50-0.59) | 0.74  (0.51-0.99) | 0.74  (0.51-0.99) | 0.75  (0.51-0.99) | 0.73  (0.51-0.99) | 0.03  (0.02-0.06) | 0.25  (0.13-0.30) | 0.03  (0.02-0.03) | 0.19  (0.15-0.20) |
| ssp126 | Florida | 5.08  (0.35-9.70) | 2.08  (1.61-2.55) | 5.02  (0.22-9.80) | 0.68  (0.52-0.87) | 14.99  (14.79-15.19) | 14.89  (14.69-15.09) | 15.03  (14.84-15.23) | 24.79  (24.59-24.98) | 0.70  (0.17-0.89) | 0.64  (0.59-0.69) | 0.89  (0.72-1.00) | 9.82  (9.30-10.00) | 0.73  (0.51-0.98) | 0.73  (0.51-0.98) | 0.77  (0.52-0.99) | 0.51  (0.50-0.52) | 0.75  (0.51-0.99) | 0.69  (0.51-0.98) | 0.75  (0.51-0.99) | 0.80  (0.55-0.99) | 0.14  (0.07-0.21) | 0.03  (0.01-0.04) | 0.41  (0.39-0.43) | 0.19  (0.15-0.20) |
| ssp126 | Texas | 4.86  (0.14-9.79) | 0.67  (0.01-2.49) | 4.94  (0.19-9.75) | 0.68  (0.52-0.87) | 15.00  (14.80-15.20) | 14.99  (14.80-15.19) | 15.00  (14.81-15.20) | 24.99  (24.79-25.20) | 0.48  (0.03-0.88) | 0.72  (0.58-0.86) | 0.91  (0.76-1.00) | 8.18  (5.15-9.93) | 0.75  (0.51-0.99) | 0.75  (0.51-0.99) | 0.76  (0.52-0.99) | 0.68  (0.51-0.96) | 0.75  (0.51-0.98) | 0.70  (0.51-0.98) | 0.75  (0.51-0.99) | 0.76  (0.52-0.99) | 0.72  (0.25-0.99) | 0.14  (0.02-0.29) | 2.66  (2.32-3.06) | 0.14  (0.02-0.20) |
| ssp370 | Arizona | 0.42  (0.04-4.85) | 0.37  (0.02-1.02) | 5.06  (0.23-9.70) | 0.68  (0.52-0.87) | 15.00  (14.81-15.19) | 14.86  (14.67-15.05) | 14.99  (14.80-15.19) | 25.00  (24.81-25.20) | 0.46  (0.03-0.88) | 0.87  (0.81-0.90) | 0.91  (0.74-1.00) | 5.10  (1.97-9.38) | 0.74  (0.51-0.99) | 0.89  (0.65-1.00) | 0.78  (0.52-0.99) | 0.86  (0.52-1.00) | 0.76  (0.52-0.99) | 0.75  (0.51-0.99) | 0.75  (0.51-0.99) | 0.71  (0.50-0.98) | 0.39  (0.23-0.78) | 0.07  (0.01-0.27) | 0.45  (0.38-0.52) | 0.19  (0.16-0.20) |
| ssp370 | California | 5.03  (0.43-9.70) | 0.00  (0.00-0.01) | 5.02  (0.25-9.78) | 0.64  (0.51-0.82) | 14.97  (14.77-15.16) | 14.87  (14.67-15.06) | 15.02  (14.83-15.21) | 24.92  (24.74-25.10) | 0.88  (0.81-0.90) | 0.58  (0.52-0.67) | 0.48  (0.36-0.61) | 9.50  (8.55-9.98) | 0.92  (0.77-1.00) | 0.87  (0.60-1.00) | 0.71  (0.55-0.87) | 0.52  (0.50-0.56) | 0.74  (0.51-0.99) | 0.74  (0.51-0.99) | 0.72  (0.51-0.98) | 0.73  (0.51-0.98) | 0.15  (0.11-0.20) | 0.24  (0.14-0.30) | 0.03  (0.02-0.03) | 0.18  (0.12-0.20) |
| ssp370 | Florida | 5.04  (0.31-9.72) | 2.13  (1.61-2.65) | 4.96  (0.29-9.77) | 0.68  (0.52-0.87) | 14.99  (14.79-15.18) | 14.89  (14.69-15.09) | 15.05  (14.86-15.25) | 24.77  (24.57-24.96) | 0.73  (0.28-0.90) | 0.63  (0.58-0.69) | 0.84  (0.67-0.99) | 9.84  (9.41-10.00) | 0.71  (0.51-0.98) | 0.70  (0.51-0.97) | 0.78  (0.53-0.99) | 0.50  (0.50-0.52) | 0.75  (0.51-0.99) | 0.76  (0.51-0.99) | 0.81  (0.53-0.99) | 0.81  (0.56-0.99) | 0.21  (0.14-0.31) | 0.01  (0.01-0.02) | 0.43  (0.41-0.46) | 0.19  (0.15-0.20) |
| ssp370 | Texas | 4.89  (0.15-9.74) | 1.05  (0.03-3.71) | 4.98  (0.26-9.74) | 0.68  (0.52-0.87) | 15.00  (14.80-15.20) | 14.99  (14.80-15.19) | 15.01  (14.82-15.20) | 24.99  (24.80-25.19) | 0.46  (0.03-0.88) | 0.67  (0.51-0.82) | 0.92  (0.78-1.00) | 7.93  (4.63-9.93) | 0.75  (0.51-0.99) | 0.77  (0.51-0.99) | 0.78  (0.52-0.99) | 0.67  (0.51-0.95) | 0.75  (0.51-0.99) | 0.76  (0.52-0.99) | 0.77  (0.52-0.99) | 0.76  (0.52-0.99) | 0.73  (0.25-0.99) | 0.12  (0.01-0.28) | 2.84  (2.48-3.25) | 0.11  (0.01-0.20) |
| ssp585 | Arizona | 0.21  (0.05-0.46) | 0.37  (0.02-1.00) | 4.83  (0.31-9.79) | 0.67  (0.52-0.86) | 14.99  (14.78-15.18) | 14.95  (14.76-15.14) | 15.00  (14.81-15.17) | 24.99  (24.80-25.16) | 0.41  (0.03-0.84) | 0.82  (0.69-0.89) | 0.92  (0.79-1.00) | 8.91  (6.46-9.96) | 0.72  (0.51-0.98) | 0.83  (0.56-0.99) | 0.82  (0.55-0.99) | 0.86  (0.51-1.00) | 0.76  (0.52-0.99) | 0.73  (0.51-0.99) | 0.74  (0.51-0.99) | 0.59  (0.50-0.96) | 0.41  (0.21-0.71) | 0.06  (0.01-0.18) | 0.43  (0.37-0.51) | 0.17  (0.11-0.20) |
| ssp585 | California | 5.32  (1.01-9.79) | 0.02  (0.00-0.07) | 5.08  (0.18-9.79) | 0.68  (0.53-0.86) | 15.00  (14.80-15.19) | 14.92  (14.72-15.11) | 15.07  (14.87-15.26) | 24.99  (24.80-25.19) | 0.32  (0.01-0.85) | 0.77  (0.57-0.90) | 0.32  (0.18-0.49) | 5.68  (2.92-9.79) | 0.83  (0.55-0.99) | 0.59  (0.50-0.89) | 0.87  (0.59-1.00) | 0.58  (0.50-0.76) | 0.75  (0.51-0.99) | 0.75  (0.51-0.99) | 0.75  (0.52-0.99) | 0.74  (0.51-0.99) | 0.91  (0.69-1.00) | 0.24  (0.12-0.30) | 0.03  (0.03-0.03) | 0.19  (0.15-0.20) |
| ssp585 | Florida | 4.97  (0.29-9.75) | 2.14  (1.66-2.64) | 5.03  (0.28-9.75) | 0.68  (0.52-0.87) | 14.98  (14.79-15.18) | 14.90  (14.70-15.10) | 15.07  (14.86-15.26) | 24.78  (24.58-24.96) | 0.76  (0.34-0.90) | 0.62  (0.57-0.68) | 0.82  (0.63-0.98) | 9.83  (9.41-9.99) | 0.73  (0.51-0.98) | 0.74  (0.52-0.98) | 0.78  (0.52-0.99) | 0.51  (0.50-0.52) | 0.75  (0.51-0.99) | 0.68  (0.51-0.97) | 0.76  (0.52-0.99) | 0.80  (0.56-0.99) | 0.20  (0.12-0.31) | 0.02  (0.01-0.04) | 0.42  (0.40-0.44) | 0.18  (0.14-0.20) |
| ssp585 | Texas | 4.90  (0.21-9.73) | 0.73  (0.02-2.60) | 4.97  (0.23-9.69) | 0.68  (0.52-0.86) | 15.00  (14.80-15.20) | 14.99  (14.80-15.19) | 15.00  (14.81-15.19) | 24.99  (24.80-25.19) | 0.47  (0.03-0.88) | 0.71  (0.57-0.85) | 0.93  (0.79-1.00) | 7.98  (5.18-9.93) | 0.75  (0.51-0.99) | 0.77  (0.52-0.99) | 0.76  (0.51-0.99) | 0.70  (0.51-0.98) | 0.75  (0.51-0.99) | 0.71  (0.51-0.98) | 0.76  (0.51-0.99) | 0.76  (0.51-0.99) | 0.55  (0.10-0.97) | 0.24  (0.13-0.30) | 2.64  (2.30-3.03) | 0.13  (0.02-0.20) |

**Table S6 Posterior summary of Singapore by different SSPs and areas.**

| SSP | Area | $A_{n,0}$ | $F_{n,0}$ | $M_{n,0}$ | $\delta$ | $T_{1,\mu,a}$ | $T_{1,\mu,f}$ | $T_{1,\tau}$ | $T_{1,\rho}$ | $L_{\mu,a}$ | $L_{\mu,f}$ | $L_{\tau}$ | $L_{\rho}$ | $k_{1,\mu,a}$ | $k_{1,\mu,f}$ | $k_{1,\tau}$ | $k_{1,\rho}$ | $k_{2,\mu,a}$ | $k_{2,\mu,f}$ | $k_{2,\tau}$ | $k_{2,\rho}$ | $\omega$ | $\lambda$ | $\sigma$ | $\alpha$ |
| --- | --- | --- | --- | --- | --- | --- | --- | --- | --- | --- | --- | --- | --- | --- | --- | --- | --- | --- | --- | --- | --- | --- | --- | --- | --- |
| ssp126 | ANG MO KIO | 0.05  (0.00-0.14) | 0.06  (0.01-0.09) | 0.50  (0.02-0.97) | 0.68  (0.52-0.87) | 15.00  (14.81-15.20) | 15.00  (14.81-15.20) | 14.99  (14.80-15.19) | 25.06  (24.86-25.26) | 0.26  (0.01-0.68) | 0.60  (0.49-0.68) | 0.99  (0.97-1.00) | 1.01  (1.00-1.04) | 0.75  (0.51-0.99) | 0.76  (0.51-0.99) | 0.76  (0.52-0.99) | 0.51  (0.50-0.56) | 0.75  (0.51-0.99) | 0.79  (0.55-0.99) | 0.83  (0.65-0.99) | 0.52  (0.50-0.56) | 0.10  (0.09-0.10) | 0.09  (0.08-0.10) | 0.06  (0.06-0.07) | 0.08  (0.00-0.19) |
| ssp126 | BISHAN | 0.12  (0.00-0.84) | 0.02  (0.00-0.05) | 0.48  (0.02-0.97) | 0.68  (0.53-0.86) | 15.00  (14.82-15.20) | 15.00  (14.81-15.20) | 15.00  (14.81-15.20) | 25.03  (24.85-25.22) | 0.21  (0.00-0.63) | 0.65  (0.49-0.87) | 0.77  (0.10-1.00) | 1.02  (1.00-1.08) | 0.76  (0.51-0.99) | 0.74  (0.51-0.99) | 0.76  (0.51-0.99) | 0.54  (0.50-0.66) | 0.75  (0.51-0.99) | 0.86  (0.63-0.99) | 0.83  (0.51-1.00) | 0.55  (0.50-0.72) | 0.08  (0.01-0.10) | 0.06  (0.01-0.10) | 0.05  (0.04-0.05) | 0.10  (0.01-0.20) |
| ssp126 | BUKIT MERAH | 0.24  (0.00-0.94) | 0.05  (0.02-0.09) | 0.50  (0.03-0.98) | 0.68  (0.52-0.87) | 15.00  (14.80-15.19) | 15.00  (14.81-15.20) | 15.00  (14.80-15.19) | 25.05  (24.85-25.26) | 0.36  (0.01-0.84) | 0.71  (0.50-0.87) | 0.55  (0.10-1.00) | 1.04  (1.00-1.22) | 0.75  (0.51-0.99) | 0.73  (0.51-0.98) | 0.75  (0.51-0.99) | 0.59  (0.50-0.94) | 0.76  (0.52-0.99) | 0.94  (0.79-1.00) | 0.80  (0.51-1.00) | 0.59  (0.50-0.94) | 0.06  (0.01-0.10) | 0.05  (0.01-0.10) | 0.05  (0.04-0.05) | 0.13  (0.01-0.20) |
| ssp126 | BUKIT PANJANG | 0.13  (0.00-0.88) | 0.02  (0.00-0.04) | 0.49  (0.03-0.97) | 0.68  (0.52-0.86) | 15.00  (14.81-15.19) | 15.01  (14.80-15.20) | 15.00  (14.80-15.20) | 25.15  (24.90-25.35) | 0.16  (0.00-0.59) | 0.66  (0.52-0.86) | 0.73  (0.10-0.99) | 1.02  (1.00-1.10) | 0.75  (0.51-0.99) | 0.74  (0.51-0.98) | 0.75  (0.51-0.98) | 0.53  (0.50-0.72) | 0.75  (0.51-0.99) | 0.95  (0.82-1.00) | 0.90  (0.54-1.00) | 0.55  (0.50-0.89) | 0.09  (0.05-0.10) | 0.02  (0.01-0.02) | 0.04  (0.04-0.05) | 0.12  (0.01-0.20) |
| ssp126 | BUKIT TIMAH | 0.05  (0.00-0.12) | 0.04  (0.00-0.07) | 0.50  (0.03-0.98) | 0.68  (0.53-0.87) | 15.00  (14.80-15.20) | 15.00  (14.81-15.19) | 15.00  (14.81-15.20) | 25.00  (24.81-25.19) | 0.29  (0.01-0.87) | 0.49  (0.34-0.61) | 0.96  (0.83-1.00) | 1.31  (1.02-1.73) | 0.75  (0.51-0.99) | 0.74  (0.51-0.99) | 0.75  (0.51-0.99) | 0.76  (0.52-0.99) | 0.75  (0.51-0.99) | 0.73  (0.51-0.98) | 0.81  (0.55-0.99) | 0.71  (0.51-0.97) | 0.02  (0.01-0.03) | 0.02  (0.01-0.03) | 0.03  (0.03-0.03) | 0.05  (0.00-0.15) |
| ssp126 | CENTRAL AREA | 0.07  (0.00-0.18) | 0.06  (0.01-0.09) | 0.51  (0.02-0.98) | 0.68  (0.52-0.88) | 15.00  (14.81-15.20) | 15.00  (14.79-15.20) | 15.00  (14.80-15.20) | 25.01  (24.81-25.20) | 0.29  (0.01-0.74) | 0.60  (0.49-0.68) | 0.98  (0.92-1.00) | 1.01  (1.00-1.05) | 0.76  (0.51-0.99) | 0.76  (0.52-0.99) | 0.75  (0.51-0.99) | 0.52  (0.50-0.58) | 0.75  (0.51-0.99) | 0.83  (0.57-0.99) | 0.78  (0.54-0.98) | 0.52  (0.50-0.58) | 0.10  (0.09-0.10) | 0.06  (0.02-0.10) | 0.05  (0.05-0.06) | 0.09  (0.00-0.19) |
| ssp126 | CHANGI | 0.04  (0.00-0.10) | 0.02  (0.00-0.04) | 0.51  (0.02-0.98) | 0.68  (0.52-0.87) | 15.00  (14.81-15.20) | 15.00  (14.81-15.20) | 15.00  (14.81-15.21) | 24.99  (24.79-25.18) | 0.32  (0.01-0.82) | 0.53  (0.40-0.64) | 0.90  (0.75-0.99) | 1.17  (1.00-1.55) | 0.75  (0.51-0.99) | 0.75  (0.51-0.99) | 0.75  (0.51-0.99) | 0.73  (0.51-0.99) | 0.75  (0.51-0.99) | 0.72  (0.51-0.98) | 0.72  (0.51-0.98) | 0.68  (0.51-0.97) | 0.06  (0.02-0.10) | 0.04  (0.01-0.09) | 0.04  (0.04-0.05) | 0.05  (0.00-0.16) |
| ssp126 | CLEMENTI | 0.05  (0.00-0.14) | 0.06  (0.02-0.08) | 0.50  (0.02-0.98) | 0.68  (0.53-0.87) | 15.00  (14.79-15.20) | 15.01  (14.81-15.21) | 15.00  (14.82-15.20) | 25.06  (24.86-25.23) | 0.49  (0.03-0.86) | 0.58  (0.47-0.66) | 0.98  (0.91-1.00) | 1.02  (1.00-1.09) | 0.76  (0.51-0.99) | 0.74  (0.51-0.99) | 0.76  (0.52-0.98) | 0.52  (0.50-0.59) | 0.77  (0.51-0.99) | 0.92  (0.76-1.00) | 0.94  (0.80-1.00) | 0.56  (0.50-0.71) | 0.10  (0.09-0.10) | 0.06  (0.01-0.10) | 0.04  (0.04-0.04) | 0.08  (0.00-0.19) |
| ssp126 | JURONG EAST | 0.09  (0.01-0.18) | 0.04  (0.00-0.09) | 0.50  (0.03-0.97) | 0.68  (0.52-0.87) | 15.00  (14.80-15.19) | 15.00  (14.81-15.19) | 15.00  (14.80-15.19) | 25.36  (25.15-25.56) | 0.83  (0.32-0.90) | 0.58  (0.49-0.68) | 0.99  (0.96-1.00) | 1.01  (1.00-1.03) | 0.75  (0.51-0.99) | 0.72  (0.51-0.98) | 0.75  (0.51-0.99) | 0.51  (0.50-0.55) | 0.84  (0.56-0.99) | 0.96  (0.87-1.00) | 0.96  (0.86-1.00) | 0.59  (0.50-0.97) | 0.10  (0.10-0.10) | 0.08  (0.05-0.10) | 0.07  (0.06-0.07) | 0.10  (0.01-0.19) |
| ssp126 | JURONG WEST | 0.27  (0.00-0.95) | 0.03  (0.00-0.07) | 0.50  (0.03-0.98) | 0.68  (0.52-0.87) | 15.00  (14.81-15.19) | 15.00  (14.81-15.21) | 15.00  (14.80-15.20) | 25.15  (24.87-25.41) | 0.28  (0.00-0.89) | 0.70  (0.51-0.86) | 0.54  (0.10-1.00) | 1.02  (1.00-1.10) | 0.75  (0.51-0.99) | 0.72  (0.50-0.98) | 0.75  (0.51-0.98) | 0.54  (0.50-0.71) | 0.77  (0.51-0.99) | 0.97  (0.87-1.00) | 0.87  (0.53-1.00) | 0.58  (0.50-0.92) | 0.09  (0.06-0.10) | 0.03  (0.01-0.07) | 0.05  (0.04-0.06) | 0.13  (0.02-0.20) |
| ssp126 | KALLANG | 0.29  (0.01-0.95) | 0.05  (0.01-0.09) | 0.51  (0.03-0.98) | 0.68  (0.52-0.87) | 15.00  (14.81-15.19) | 15.00  (14.79-15.19) | 15.00  (14.80-15.19) | 25.05  (24.85-25.26) | 0.23  (0.00-0.72) | 0.71  (0.51-0.88) | 0.54  (0.10-1.00) | 1.03  (1.00-1.14) | 0.74  (0.51-0.99) | 0.76  (0.51-0.99) | 0.75  (0.51-0.99) | 0.56  (0.50-0.85) | 0.75  (0.51-0.99) | 0.91  (0.71-1.00) | 0.71  (0.50-0.98) | 0.57  (0.50-0.89) | 0.06  (0.01-0.10) | 0.05  (0.01-0.10) | 0.06  (0.05-0.07) | 0.13  (0.01-0.20) |
| ssp126 | MANDAI | 0.20  (0.00-0.90) | 0.05  (0.00-0.10) | 0.51  (0.03-0.97) | 0.68  (0.52-0.86) | 15.00  (14.80-15.19) | 15.00  (14.81-15.20) | 15.00  (14.80-15.19) | 25.01  (24.80-25.20) | 0.31  (0.01-0.87) | 0.51  (0.37-0.61) | 0.95  (0.86-1.00) | 1.19  (1.01-1.64) | 0.75  (0.51-0.98) | 0.75  (0.51-0.99) | 0.76  (0.51-0.99) | 0.70  (0.51-0.97) | 0.75  (0.51-0.99) | 0.71  (0.51-0.98) | 0.82  (0.55-0.99) | 0.73  (0.51-0.98) | 0.07  (0.04-0.10) | 0.03  (0.01-0.09) | 0.04  (0.04-0.05) | 0.05  (0.00-0.15) |
| ssp126 | MARINE PARADE | 0.41  (0.19-0.90) | 0.16  (0.08-0.25) | 0.50  (0.02-0.98) | 0.68  (0.52-0.87) | 15.00  (14.80-15.19) | 15.00  (14.81-15.19) | 15.00  (14.80-15.20) | 25.11  (24.91-25.31) | 0.34  (0.00-0.88) | 0.60  (0.43-0.85) | 0.74  (0.10-0.99) | 1.06  (1.00-1.23) | 0.75  (0.51-0.99) | 0.73  (0.51-0.99) | 0.74  (0.51-0.99) | 0.56  (0.50-0.70) | 0.75  (0.51-0.99) | 0.90  (0.70-1.00) | 0.84  (0.51-1.00) | 0.83  (0.50-1.00) | 0.07  (0.01-0.10) | 0.05  (0.01-0.10) | 0.03  (0.03-0.04) | 0.08  (0.00-0.19) |
| ssp126 | NOVENA | 0.24  (0.06-0.42) | 0.11  (0.03-0.18) | 0.50  (0.03-0.98) | 0.68  (0.52-0.87) | 15.00  (14.81-15.19) | 15.00  (14.80-15.20) | 15.00  (14.79-15.20) | 24.95  (24.76-25.14) | 0.29  (0.01-0.76) | 0.56  (0.42-0.67) | 0.92  (0.78-1.00) | 1.17  (1.01-1.47) | 0.75  (0.51-0.99) | 0.75  (0.51-0.99) | 0.75  (0.51-0.99) | 0.81  (0.51-0.99) | 0.75  (0.51-0.99) | 0.72  (0.51-0.99) | 0.67  (0.51-0.96) | 0.62  (0.50-0.89) | 0.04  (0.02-0.07) | 0.03  (0.02-0.04) | 0.04  (0.04-0.05) | 0.06  (0.00-0.18) |
| ssp126 | PASIR RIS | 0.57  (0.21-0.97) | 0.03  (0.00-0.10) | 0.50  (0.03-0.98) | 0.68  (0.52-0.87) | 15.00  (14.80-15.19) | 15.00  (14.81-15.20) | 15.00  (14.80-15.21) | 25.03  (24.84-25.22) | 0.78  (0.19-0.90) | 0.59  (0.48-0.68) | 0.85  (0.73-0.96) | 1.07  (1.00-1.21) | 0.75  (0.52-0.98) | 0.75  (0.51-0.99) | 0.75  (0.51-0.99) | 0.58  (0.50-0.78) | 0.80  (0.53-0.99) | 0.91  (0.73-1.00) | 0.90  (0.67-1.00) | 0.84  (0.55-1.00) | 0.09  (0.08-0.10) | 0.01  (0.01-0.02) | 0.04  (0.04-0.04) | 0.08  (0.00-0.19) |
| ssp126 | QUEENSTOWN | 0.07  (0.00-0.20) | 0.07  (0.01-0.11) | 0.51  (0.03-0.97) | 0.68  (0.51-0.87) | 15.00  (14.80-15.20) | 15.00  (14.81-15.19) | 15.00  (14.80-15.20) | 25.18  (25.00-25.37) | 0.75  (0.10-0.90) | 0.58  (0.48-0.67) | 0.98  (0.88-1.00) | 1.01  (1.00-1.04) | 0.74  (0.51-0.98) | 0.74  (0.51-0.99) | 0.75  (0.51-0.99) | 0.52  (0.50-0.55) | 0.80  (0.53-0.99) | 0.94  (0.82-1.00) | 0.94  (0.80-1.00) | 0.53  (0.50-0.67) | 0.10  (0.10-0.10) | 0.07  (0.01-0.10) | 0.05  (0.05-0.06) | 0.09  (0.01-0.19) |
| ssp126 | SEMBAWANG | 0.25  (0.00-0.95) | 0.03  (0.02-0.06) | 0.49  (0.02-0.97) | 0.68  (0.52-0.87) | 15.01  (14.82-15.23) | 15.00  (14.82-15.19) | 15.00  (14.81-15.19) | 25.01  (24.81-25.20) | 0.21  (0.00-0.78) | 0.69  (0.49-0.87) | 0.54  (0.10-1.00) | 1.05  (1.00-1.17) | 0.76  (0.51-0.99) | 0.74  (0.51-0.97) | 0.74  (0.51-0.98) | 0.57  (0.50-0.86) | 0.74  (0.51-0.99) | 0.92  (0.71-1.00) | 0.77  (0.51-0.99) | 0.60  (0.50-0.92) | 0.06  (0.01-0.10) | 0.03  (0.01-0.09) | 0.02  (0.02-0.03) | 0.11  (0.00-0.20) |
| ssp126 | SERANGOON | 0.32  (0.19-0.44) | 0.07  (0.02-0.12) | 0.49  (0.02-0.97) | 0.68  (0.52-0.86) | 15.00  (14.81-15.20) | 15.00  (14.80-15.20) | 15.00  (14.80-15.20) | 25.03  (24.83-25.22) | 0.33  (0.02-0.77) | 0.56  (0.44-0.65) | 0.92  (0.84-0.99) | 1.11  (1.00-1.32) | 0.75  (0.52-0.99) | 0.75  (0.51-0.99) | 0.75  (0.51-0.99) | 0.63  (0.50-0.89) | 0.75  (0.51-0.99) | 0.86  (0.60-0.99) | 0.85  (0.60-0.99) | 0.77  (0.52-0.99) | 0.03  (0.02-0.04) | 0.03  (0.02-0.04) | 0.03  (0.03-0.03) | 0.07  (0.00-0.18) |
| ssp126 | SUNGEI KADUT | 0.00  (0.00-0.01) | 0.00  (0.00-0.01) | 0.50  (0.03-0.98) | 0.68  (0.53-0.86) | 15.00  (14.81-15.19) | 15.00  (14.81-15.19) | 15.00  (14.82-15.19) | 24.95  (24.77-25.14) | 0.53  (0.03-0.89) | 0.56  (0.43-0.67) | 0.92  (0.77-1.00) | 1.10  (1.00-1.33) | 0.75  (0.51-0.99) | 0.74  (0.51-0.99) | 0.75  (0.51-0.99) | 0.78  (0.51-0.99) | 0.76  (0.51-0.99) | 0.64  (0.50-0.97) | 0.76  (0.51-0.99) | 0.60  (0.50-0.90) | 0.05  (0.01-0.10) | 0.06  (0.01-0.10) | 0.03  (0.03-0.03) | 0.05  (0.00-0.16) |
| ssp126 | TANGLIN | 0.00  (0.00-0.02) | 0.00  (0.00-0.01) | 0.50  (0.02-0.97) | 0.68  (0.52-0.87) | 15.00  (14.80-15.20) | 15.00  (14.80-15.19) | 15.00  (14.79-15.19) | 24.95  (24.75-25.15) | 0.38  (0.01-0.89) | 0.56  (0.43-0.66) | 0.97  (0.86-1.00) | 1.11  (1.00-1.39) | 0.75  (0.52-0.99) | 0.75  (0.51-0.99) | 0.75  (0.51-0.99) | 0.80  (0.51-0.99) | 0.76  (0.51-0.99) | 0.64  (0.50-0.96) | 0.82  (0.54-0.99) | 0.60  (0.50-0.87) | 0.07  (0.04-0.10) | 0.04  (0.01-0.10) | 0.05  (0.04-0.05) | 0.05  (0.00-0.16) |
| ssp126 | TOA PAYOH | 0.15  (0.00-0.88) | 0.03  (0.00-0.06) | 0.49  (0.02-0.97) | 0.68  (0.53-0.87) | 15.00  (14.80-15.21) | 15.00  (14.81-15.20) | 15.00  (14.80-15.19) | 25.06  (24.86-25.25) | 0.24  (0.00-0.71) | 0.65  (0.49-0.87) | 0.77  (0.10-1.00) | 1.02  (1.00-1.09) | 0.75  (0.51-0.99) | 0.75  (0.51-0.99) | 0.75  (0.52-0.98) | 0.53  (0.50-0.68) | 0.75  (0.51-0.99) | 0.91  (0.73-1.00) | 0.85  (0.51-1.00) | 0.54  (0.50-0.75) | 0.08  (0.01-0.10) | 0.07  (0.01-0.10) | 0.06  (0.05-0.07) | 0.10  (0.01-0.20) |
| ssp370 | ANG MO KIO | 0.16  (0.00-0.87) | 0.09  (0.03-0.14) | 0.50  (0.02-0.98) | 0.68  (0.53-0.87) | 15.00  (14.81-15.20) | 15.00  (14.80-15.19) | 15.00  (14.80-15.20) | 25.16  (24.94-25.37) | 0.22  (0.00-0.65) | 0.65  (0.49-0.87) | 0.77  (0.10-1.00) | 1.02  (1.00-1.07) | 0.74  (0.51-0.98) | 0.73  (0.51-0.99) | 0.75  (0.51-0.99) | 0.55  (0.50-0.67) | 0.74  (0.51-0.99) | 0.90  (0.72-1.00) | 0.85  (0.51-1.00) | 0.58  (0.50-0.84) | 0.08  (0.01-0.10) | 0.07  (0.01-0.10) | 0.06  (0.05-0.08) | 0.10  (0.00-0.20) |
| ssp370 | BISHAN | 0.07  (0.00-0.16) | 0.05  (0.00-0.09) | 0.49  (0.02-0.97) | 0.68  (0.52-0.87) | 15.00  (14.80-15.19) | 15.00  (14.80-15.19) | 15.00  (14.79-15.21) | 25.09  (24.90-25.28) | 0.29  (0.01-0.71) | 0.56  (0.45-0.64) | 0.98  (0.95-1.00) | 1.03  (1.00-1.10) | 0.75  (0.51-0.99) | 0.75  (0.51-0.99) | 0.75  (0.51-0.99) | 0.56  (0.50-0.69) | 0.75  (0.51-0.99) | 0.88  (0.67-1.00) | 0.93  (0.78-1.00) | 0.84  (0.56-0.99) | 0.10  (0.09-0.10) | 0.08  (0.05-0.10) | 0.04  (0.04-0.04) | 0.07  (0.00-0.18) |
| ssp370 | BUKIT MERAH | 0.38  (0.01-0.96) | 0.05  (0.03-0.09) | 0.51  (0.03-0.98) | 0.68  (0.52-0.87) | 15.00  (14.80-15.19) | 15.00  (14.81-15.19) | 15.00  (14.81-15.20) | 25.07  (24.87-25.27) | 0.09  (0.00-0.55) | 0.77  (0.53-0.89) | 0.32  (0.10-0.99) | 1.02  (1.00-1.09) | 0.75  (0.51-0.99) | 0.70  (0.50-0.98) | 0.75  (0.51-0.99) | 0.55  (0.50-0.72) | 0.75  (0.51-0.99) | 0.93  (0.76-1.00) | 0.71  (0.50-0.99) | 0.58  (0.50-0.89) | 0.03  (0.01-0.10) | 0.02  (0.01-0.04) | 0.05  (0.04-0.05) | 0.14  (0.02-0.20) |
| ssp370 | BUKIT PANJANG | 0.04  (0.00-0.12) | 0.04  (0.01-0.07) | 0.50  (0.02-0.98) | 0.68  (0.53-0.87) | 15.00  (14.81-15.20) | 15.00  (14.80-15.20) | 15.00  (14.80-15.20) | 25.16  (24.98-25.35) | 0.30  (0.01-0.75) | 0.61  (0.50-0.69) | 0.96  (0.89-1.00) | 1.01  (1.00-1.05) | 0.75  (0.51-0.99) | 0.74  (0.51-0.99) | 0.74  (0.51-0.99) | 0.53  (0.50-0.60) | 0.75  (0.51-0.99) | 0.90  (0.73-1.00) | 0.93  (0.78-1.00) | 0.56  (0.50-0.75) | 0.10  (0.09-0.10) | 0.02  (0.02-0.03) | 0.04  (0.04-0.04) | 0.09  (0.00-0.19) |
| ssp370 | BUKIT TIMAH | 0.05  (0.00-0.13) | 0.05  (0.01-0.08) | 0.50  (0.03-0.98) | 0.68  (0.52-0.87) | 15.00  (14.81-15.19) | 15.00  (14.81-15.20) | 15.00  (14.80-15.20) | 25.11  (24.90-25.32) | 0.35  (0.01-0.88) | 0.42  (0.25-0.56) | 0.97  (0.88-1.00) | 1.41  (1.05-1.88) | 0.75  (0.51-0.99) | 0.75  (0.51-0.99) | 0.74  (0.51-0.98) | 0.76  (0.53-0.98) | 0.75  (0.51-0.99) | 0.77  (0.52-0.99) | 0.86  (0.57-0.99) | 0.84  (0.57-0.99) | 0.07  (0.02-0.10) | 0.04  (0.01-0.10) | 0.03  (0.03-0.03) | 0.04  (0.00-0.13) |
| ssp370 | CENTRAL AREA | 0.10  (0.01-0.24) | 0.09  (0.02-0.13) | 0.49  (0.03-0.98) | 0.68  (0.52-0.86) | 15.00  (14.80-15.21) | 15.00  (14.81-15.19) | 15.00  (14.80-15.19) | 25.06  (24.86-25.25) | 0.35  (0.02-0.79) | 0.57  (0.46-0.66) | 0.98  (0.93-1.00) | 1.03  (1.00-1.12) | 0.75  (0.51-0.99) | 0.74  (0.51-0.99) | 0.76  (0.51-0.99) | 0.57  (0.50-0.72) | 0.74  (0.51-0.99) | 0.85  (0.61-0.99) | 0.89  (0.68-1.00) | 0.66  (0.50-0.96) | 0.10  (0.09-0.10) | 0.08  (0.04-0.10) | 0.05  (0.04-0.05) | 0.07  (0.00-0.18) |
| ssp370 | CHANGI | 0.10  (0.00-0.77) | 0.02  (0.00-0.05) | 0.51  (0.02-0.97) | 0.68  (0.52-0.87) | 15.00  (14.80-15.20) | 15.00  (14.80-15.19) | 15.00  (14.81-15.18) | 24.99  (24.79-25.19) | 0.37  (0.02-0.84) | 0.53  (0.41-0.64) | 0.90  (0.75-1.00) | 1.17  (1.01-1.55) | 0.75  (0.51-0.98) | 0.76  (0.51-0.98) | 0.75  (0.51-0.99) | 0.74  (0.51-0.99) | 0.75  (0.51-0.99) | 0.71  (0.51-0.98) | 0.71  (0.51-0.98) | 0.68  (0.51-0.96) | 0.05  (0.01-0.10) | 0.04  (0.01-0.10) | 0.04  (0.04-0.05) | 0.06  (0.00-0.17) |
| ssp370 | CLEMENTI | 0.08  (0.00-0.20) | 0.08  (0.02-0.12) | 0.49  (0.02-0.97) | 0.68  (0.53-0.86) | 15.00  (14.81-15.19) | 15.00  (14.80-15.19) | 15.00  (14.81-15.21) | 25.05  (24.86-25.23) | 0.43  (0.02-0.87) | 0.55  (0.43-0.63) | 0.96  (0.90-1.00) | 1.08  (1.00-1.26) | 0.75  (0.51-0.99) | 0.75  (0.51-0.99) | 0.75  (0.51-0.99) | 0.61  (0.50-0.84) | 0.74  (0.51-0.98) | 0.86  (0.63-1.00) | 0.87  (0.64-1.00) | 0.86  (0.62-0.99) | 0.09  (0.07-0.10) | 0.07  (0.03-0.10) | 0.04  (0.03-0.04) | 0.06  (0.00-0.17) |
| ssp370 | JURONG EAST | 0.13  (0.01-0.25) | 0.06  (0.00-0.12) | 0.50  (0.03-0.97) | 0.68  (0.52-0.86) | 15.00  (14.81-15.20) | 15.00  (14.79-15.20) | 15.00  (14.79-15.20) | 25.37  (25.17-25.57) | 0.31  (0.01-0.86) | 0.59  (0.49-0.69) | 0.94  (0.88-0.99) | 1.01  (1.00-1.03) | 0.75  (0.51-0.99) | 0.73  (0.51-0.99) | 0.74  (0.51-0.99) | 0.52  (0.50-0.56) | 0.76  (0.52-0.99) | 0.94  (0.82-1.00) | 0.96  (0.86-1.00) | 0.81  (0.50-1.00) | 0.10  (0.10-0.10) | 0.05  (0.04-0.07) | 0.06  (0.06-0.07) | 0.09  (0.00-0.19) |
| ssp370 | JURONG WEST | 0.08  (0.00-0.18) | 0.05  (0.00-0.09) | 0.49  (0.02-0.97) | 0.68  (0.52-0.86) | 15.00  (14.80-15.20) | 15.00  (14.80-15.20) | 15.00  (14.80-15.19) | 25.19  (25.01-25.36) | 0.31  (0.01-0.86) | 0.60  (0.50-0.70) | 0.94  (0.87-1.00) | 1.01  (1.00-1.04) | 0.75  (0.51-0.99) | 0.74  (0.51-0.99) | 0.74  (0.51-0.99) | 0.52  (0.50-0.57) | 0.76  (0.51-0.99) | 0.92  (0.77-1.00) | 0.94  (0.81-1.00) | 0.56  (0.50-0.80) | 0.10  (0.10-0.10) | 0.03  (0.02-0.04) | 0.04  (0.04-0.05) | 0.09  (0.01-0.19) |
| ssp370 | KALLANG | 0.12  (0.01-0.26) | 0.07  (0.00-0.12) | 0.51  (0.02-0.97) | 0.68  (0.52-0.87) | 15.00  (14.80-15.20) | 15.00  (14.81-15.20) | 15.00  (14.81-15.19) | 25.08  (24.90-25.28) | 0.31  (0.01-0.75) | 0.60  (0.50-0.69) | 0.97  (0.93-1.00) | 1.01  (1.00-1.05) | 0.75  (0.51-0.99) | 0.75  (0.51-0.99) | 0.76  (0.51-0.99) | 0.52  (0.50-0.58) | 0.75  (0.51-0.99) | 0.84  (0.60-0.99) | 0.83  (0.59-0.99) | 0.53  (0.50-0.60) | 0.10  (0.09-0.10) | 0.09  (0.07-0.10) | 0.06  (0.05-0.06) | 0.08  (0.00-0.19) |
| ssp370 | MANDAI | 0.27  (0.01-0.93) | 0.06  (0.01-0.13) | 0.50  (0.03-0.97) | 0.68  (0.52-0.87) | 15.00  (14.81-15.20) | 15.00  (14.80-15.19) | 15.00  (14.81-15.19) | 24.99  (24.80-25.18) | 0.45  (0.01-0.89) | 0.53  (0.40-0.63) | 0.94  (0.80-1.00) | 1.13  (1.00-1.43) | 0.75  (0.51-0.99) | 0.76  (0.51-0.99) | 0.75  (0.51-0.99) | 0.69  (0.51-0.98) | 0.75  (0.51-0.99) | 0.72  (0.51-0.98) | 0.77  (0.52-0.99) | 0.70  (0.51-0.98) | 0.04  (0.01-0.09) | 0.05  (0.01-0.10) | 0.04  (0.04-0.05) | 0.05  (0.00-0.16) |
| ssp370 | MARINE PARADE | 0.36  (0.09-0.89) | 0.17  (0.09-0.26) | 0.50  (0.03-0.98) | 0.68  (0.52-0.86) | 15.00  (14.80-15.19) | 15.00  (14.80-15.20) | 15.00  (14.81-15.19) | 25.14  (24.91-25.35) | 0.18  (0.00-0.64) | 0.60  (0.42-0.85) | 0.77  (0.10-1.00) | 1.04  (1.00-1.13) | 0.75  (0.51-0.99) | 0.74  (0.51-0.99) | 0.74  (0.51-0.99) | 0.60  (0.50-0.79) | 0.75  (0.51-0.98) | 0.89  (0.69-1.00) | 0.86  (0.51-1.00) | 0.83  (0.51-1.00) | 0.08  (0.01-0.10) | 0.06  (0.01-0.10) | 0.03  (0.03-0.04) | 0.08  (0.00-0.19) |
| ssp370 | NOVENA | 0.31  (0.10-0.85) | 0.11  (0.03-0.19) | 0.50  (0.02-0.98) | 0.68  (0.52-0.86) | 15.00  (14.81-15.20) | 15.00  (14.80-15.19) | 15.00  (14.80-15.19) | 25.17  (24.95-25.38) | 0.33  (0.02-0.79) | 0.48  (0.33-0.60) | 0.92  (0.83-1.00) | 1.27  (1.03-1.66) | 0.75  (0.52-0.99) | 0.75  (0.51-0.99) | 0.74  (0.51-0.99) | 0.79  (0.57-0.98) | 0.75  (0.51-0.99) | 0.80  (0.54-0.99) | 0.86  (0.61-1.00) | 0.85  (0.58-0.99) | 0.09  (0.05-0.10) | 0.06  (0.02-0.10) | 0.04  (0.04-0.05) | 0.05  (0.00-0.14) |
| ssp370 | PASIR RIS | 0.60  (0.22-0.98) | 0.09  (0.04-0.12) | 0.50  (0.02-0.98) | 0.68  (0.52-0.86) | 15.00  (14.80-15.20) | 15.00  (14.80-15.19) | 15.00  (14.81-15.20) | 25.12  (24.93-25.31) | 0.53  (0.03-0.89) | 0.59  (0.48-0.67) | 0.93  (0.82-1.00) | 1.02  (1.00-1.08) | 0.75  (0.51-0.99) | 0.74  (0.51-0.98) | 0.74  (0.51-0.99) | 0.54  (0.50-0.64) | 0.76  (0.52-0.99) | 0.88  (0.66-1.00) | 0.85  (0.56-1.00) | 0.79  (0.52-0.99) | 0.10  (0.09-0.10) | 0.02  (0.01-0.03) | 0.04  (0.04-0.04) | 0.08  (0.00-0.19) |
| ssp370 | QUEENSTOWN | 0.11  (0.01-0.28) | 0.09  (0.02-0.15) | 0.49  (0.02-0.97) | 0.68  (0.52-0.87) | 15.00  (14.80-15.20) | 15.00  (14.80-15.20) | 15.00  (14.81-15.20) | 25.10  (24.91-25.30) | 0.50  (0.03-0.88) | 0.58  (0.47-0.67) | 0.91  (0.81-0.99) | 1.03  (1.00-1.09) | 0.75  (0.51-0.99) | 0.74  (0.51-0.99) | 0.75  (0.52-0.99) | 0.54  (0.50-0.62) | 0.77  (0.52-0.99) | 0.91  (0.74-1.00) | 0.83  (0.54-1.00) | 0.83  (0.52-1.00) | 0.10  (0.09-0.10) | 0.06  (0.01-0.10) | 0.05  (0.05-0.06) | 0.08  (0.00-0.19) |
| ssp370 | SEMBAWANG | 0.15  (0.00-0.87) | 0.05  (0.01-0.08) | 0.50  (0.03-0.97) | 0.68  (0.52-0.87) | 15.00  (14.80-15.20) | 15.00  (14.81-15.19) | 15.00  (14.80-15.20) | 25.04  (24.85-25.25) | 0.41  (0.00-0.88) | 0.59  (0.40-0.86) | 0.75  (0.10-1.00) | 1.12  (1.00-1.43) | 0.75  (0.51-0.99) | 0.73  (0.51-0.99) | 0.75  (0.51-0.99) | 0.69  (0.50-0.98) | 0.75  (0.51-0.99) | 0.88  (0.60-1.00) | 0.82  (0.51-0.99) | 0.69  (0.51-0.98) | 0.07  (0.01-0.10) | 0.04  (0.01-0.10) | 0.02  (0.02-0.03) | 0.08  (0.00-0.19) |
| ssp370 | SERANGOON | 0.45  (0.23-0.97) | 0.07  (0.02-0.12) | 0.50  (0.03-0.97) | 0.68  (0.52-0.87) | 15.00  (14.80-15.19) | 15.00  (14.80-15.19) | 15.00  (14.80-15.20) | 25.19  (24.99-25.39) | 0.26  (0.01-0.66) | 0.55  (0.43-0.64) | 0.90  (0.83-0.95) | 1.07  (1.00-1.23) | 0.75  (0.51-0.99) | 0.73  (0.51-0.98) | 0.75  (0.51-0.98) | 0.70  (0.53-0.92) | 0.74  (0.51-0.98) | 0.87  (0.63-1.00) | 0.90  (0.69-1.00) | 0.89  (0.69-1.00) | 0.10  (0.09-0.10) | 0.05  (0.02-0.10) | 0.03  (0.03-0.03) | 0.06  (0.00-0.18) |
| ssp370 | SUNGEI KADUT | 0.00  (0.00-0.01) | 0.00  (0.00-0.01) | 0.50  (0.03-0.97) | 0.68  (0.53-0.86) | 15.00  (14.81-15.20) | 15.00  (14.80-15.20) | 15.00  (14.80-15.19) | 24.96  (24.76-25.16) | 0.51  (0.02-0.89) | 0.55  (0.42-0.65) | 0.95  (0.80-1.00) | 1.11  (1.00-1.38) | 0.75  (0.51-0.99) | 0.75  (0.51-0.99) | 0.75  (0.52-0.98) | 0.79  (0.51-0.99) | 0.75  (0.51-0.99) | 0.65  (0.50-0.97) | 0.79  (0.52-0.99) | 0.61  (0.50-0.91) | 0.05  (0.01-0.10) | 0.06  (0.01-0.10) | 0.03  (0.03-0.03) | 0.05  (0.00-0.16) |
| ssp370 | TANGLIN | 0.00  (0.00-0.02) | 0.00  (0.00-0.01) | 0.50  (0.02-0.97) | 0.68  (0.52-0.87) | 15.00  (14.81-15.20) | 15.00  (14.80-15.20) | 15.00  (14.80-15.20) | 24.98  (24.78-25.18) | 0.39  (0.01-0.89) | 0.55  (0.42-0.65) | 0.98  (0.89-1.00) | 1.12  (1.00-1.38) | 0.75  (0.51-0.99) | 0.75  (0.51-0.99) | 0.76  (0.51-0.99) | 0.74  (0.51-0.99) | 0.75  (0.51-0.99) | 0.69  (0.51-0.98) | 0.85  (0.58-1.00) | 0.65  (0.50-0.95) | 0.07  (0.04-0.10) | 0.05  (0.01-0.10) | 0.05  (0.04-0.05) | 0.06  (0.00-0.17) |
| ssp370 | TOA PAYOH | 0.07  (0.00-0.18) | 0.05  (0.01-0.09) | 0.49  (0.02-0.97) | 0.68  (0.52-0.87) | 15.00  (14.80-15.20) | 15.00  (14.81-15.20) | 15.00  (14.79-15.20) | 25.14  (24.94-25.34) | 0.31  (0.01-0.77) | 0.60  (0.49-0.68) | 0.98  (0.94-1.00) | 1.02  (1.00-1.06) | 0.75  (0.51-0.99) | 0.74  (0.51-0.99) | 0.75  (0.51-0.99) | 0.53  (0.50-0.62) | 0.75  (0.51-0.99) | 0.90  (0.70-1.00) | 0.93  (0.76-1.00) | 0.58  (0.50-0.89) | 0.10  (0.09-0.10) | 0.06  (0.04-0.10) | 0.06  (0.05-0.06) | 0.08  (0.00-0.19) |
| ssp585 | ANG MO KIO | 0.05  (0.00-0.16) | 0.07  (0.01-0.10) | 0.50  (0.02-0.98) | 0.68  (0.52-0.87) | 15.00  (14.81-15.20) | 15.00  (14.81-15.19) | 15.00  (14.80-15.20) | 25.30  (25.11-25.51) | 0.20  (0.01-0.57) | 0.61  (0.51-0.69) | 0.98  (0.96-1.00) | 1.01  (1.00-1.03) | 0.75  (0.51-0.99) | 0.73  (0.51-0.99) | 0.75  (0.52-0.99) | 0.51  (0.50-0.55) | 0.75  (0.51-0.99) | 0.92  (0.75-1.00) | 0.94  (0.82-1.00) | 0.52  (0.50-0.57) | 0.10  (0.10-0.10) | 0.09  (0.07-0.10) | 0.06  (0.06-0.07) | 0.09  (0.00-0.19) |
| ssp585 | BISHAN | 0.47  (0.01-0.97) | 0.01  (0.00-0.05) | 0.50  (0.03-0.97) | 0.67  (0.51-0.86) | 15.00  (14.80-15.19) | 15.00  (14.80-15.20) | 15.00  (14.80-15.20) | 25.22  (25.04-25.41) | 0.21  (0.01-0.59) | 0.60  (0.49-0.68) | 0.99  (0.96-1.00) | 1.01  (1.00-1.03) | 0.75  (0.51-0.99) | 0.74  (0.51-0.99) | 0.74  (0.51-0.99) | 0.52  (0.50-0.56) | 0.75  (0.51-0.99) | 0.92  (0.75-1.00) | 0.95  (0.85-1.00) | 0.52  (0.50-0.59) | 0.10  (0.10-0.10) | 0.08  (0.04-0.10) | 0.04  (0.04-0.05) | 0.09  (0.00-0.19) |
| ssp585 | BUKIT MERAH | 0.46  (0.01-0.98) | 0.05  (0.03-0.09) | 0.50  (0.03-0.98) | 0.68  (0.52-0.86) | 15.00  (14.80-15.19) | 15.00  (14.80-15.21) | 15.00  (14.81-15.20) | 25.28  (25.10-25.46) | 0.21  (0.01-0.62) | 0.61  (0.51-0.69) | 0.98  (0.95-1.00) | 1.01  (1.00-1.02) | 0.75  (0.52-0.99) | 0.74  (0.51-0.98) | 0.75  (0.51-0.99) | 0.51  (0.50-0.54) | 0.75  (0.51-0.99) | 0.88  (0.68-1.00) | 0.92  (0.77-1.00) | 0.51  (0.50-0.55) | 0.10  (0.10-0.10) | 0.09  (0.08-0.10) | 0.05  (0.04-0.05) | 0.09  (0.01-0.19) |
| ssp585 | BUKIT PANJANG | 0.14  (0.00-0.88) | 0.04  (0.00-0.07) | 0.50  (0.03-0.97) | 0.68  (0.52-0.88) | 15.00  (14.79-15.20) | 15.00  (14.81-15.19) | 15.00  (14.81-15.20) | 25.13  (24.90-25.34) | 0.24  (0.00-0.80) | 0.66  (0.51-0.86) | 0.76  (0.10-1.00) | 1.02  (1.00-1.08) | 0.75  (0.51-0.99) | 0.72  (0.51-0.99) | 0.74  (0.51-0.99) | 0.54  (0.50-0.70) | 0.76  (0.51-0.99) | 0.89  (0.67-1.00) | 0.86  (0.52-1.00) | 0.55  (0.50-0.84) | 0.08  (0.01-0.10) | 0.07  (0.01-0.10) | 0.04  (0.04-0.05) | 0.10  (0.01-0.20) |
| ssp585 | BUKIT TIMAH | 0.08  (0.01-0.18) | 0.05  (0.01-0.10) | 0.50  (0.03-0.97) | 0.68  (0.52-0.87) | 15.00  (14.81-15.19) | 15.00  (14.81-15.20) | 15.00  (14.81-15.19) | 25.10  (24.89-25.32) | 0.62  (0.09-0.89) | 0.44  (0.28-0.58) | 0.89  (0.75-0.99) | 1.39  (1.06-1.85) | 0.74  (0.51-0.99) | 0.74  (0.51-0.99) | 0.75  (0.50-0.99) | 0.66  (0.51-0.93) | 0.76  (0.52-0.99) | 0.82  (0.55-0.99) | 0.80  (0.52-0.99) | 0.89  (0.63-1.00) | 0.05  (0.01-0.10) | 0.06  (0.02-0.10) | 0.03  (0.03-0.03) | 0.04  (0.00-0.14) |
| ssp585 | CENTRAL AREA | 0.48  (0.01-0.97) | 0.07  (0.05-0.11) | 0.52  (0.03-0.98) | 0.68  (0.53-0.87) | 15.00  (14.80-15.20) | 15.00  (14.80-15.19) | 15.00  (14.80-15.20) | 25.16  (24.97-25.34) | 0.24  (0.01-0.64) | 0.60  (0.49-0.68) | 0.98  (0.95-1.00) | 1.01  (1.00-1.04) | 0.75  (0.51-0.99) | 0.74  (0.51-0.99) | 0.75  (0.51-0.99) | 0.52  (0.50-0.57) | 0.75  (0.51-0.98) | 0.87  (0.66-1.00) | 0.92  (0.77-1.00) | 0.53  (0.50-0.60) | 0.10  (0.10-0.10) | 0.08  (0.05-0.10) | 0.05  (0.04-0.05) | 0.08  (0.00-0.19) |
| ssp585 | CHANGI | 0.50  (0.02-0.98) | 0.02  (0.00-0.05) | 0.50  (0.02-0.98) | 0.68  (0.53-0.86) | 15.00  (14.80-15.20) | 15.00  (14.80-15.20) | 15.00  (14.78-15.19) | 24.99  (24.80-25.18) | 0.37  (0.02-0.85) | 0.54  (0.40-0.64) | 0.88  (0.72-0.99) | 1.15  (1.00-1.52) | 0.75  (0.51-0.99) | 0.75  (0.51-0.99) | 0.75  (0.51-0.99) | 0.72  (0.51-0.99) | 0.75  (0.51-0.99) | 0.71  (0.51-0.98) | 0.70  (0.51-0.98) | 0.68  (0.51-0.97) | 0.05  (0.02-0.09) | 0.05  (0.01-0.10) | 0.04  (0.04-0.05) | 0.05  (0.00-0.16) |
| ssp585 | CLEMENTI | 0.06  (0.00-0.16) | 0.07  (0.02-0.11) | 0.49  (0.02-0.97) | 0.68  (0.52-0.86) | 15.00  (14.81-15.20) | 15.00  (14.80-15.20) | 15.00  (14.80-15.20) | 25.06  (24.88-25.24) | 0.28  (0.01-0.74) | 0.59  (0.47-0.67) | 0.96  (0.86-1.00) | 1.03  (1.00-1.10) | 0.75  (0.51-0.99) | 0.74  (0.51-0.99) | 0.74  (0.51-0.99) | 0.56  (0.50-0.70) | 0.75  (0.51-0.99) | 0.86  (0.63-0.99) | 0.91  (0.73-1.00) | 0.62  (0.50-0.90) | 0.10  (0.09-0.10) | 0.05  (0.01-0.10) | 0.04  (0.03-0.04) | 0.07  (0.00-0.18) |
| ssp585 | JURONG EAST | 0.50  (0.03-0.97) | 0.05  (0.03-0.08) | 0.50  (0.02-0.98) | 0.68  (0.52-0.87) | 15.00  (14.80-15.20) | 15.00  (14.81-15.20) | 15.00  (14.81-15.20) | 25.27  (24.87-25.52) | 0.30  (0.01-0.83) | 0.67  (0.51-0.88) | 0.75  (0.10-1.00) | 2.23  (1.00-8.94) | 0.75  (0.51-0.99) | 0.71  (0.51-0.98) | 0.74  (0.51-0.99) | 0.57  (0.50-0.95) | 0.75  (0.51-0.98) | 0.94  (0.79-1.00) | 0.88  (0.52-1.00) | 0.57  (0.50-0.95) | 0.08  (0.01-0.10) | 0.07  (0.01-0.10) | 0.06  (0.06-0.07) | 0.11  (0.01-0.20) |
| ssp585 | JURONG WEST | 0.48  (0.02-0.97) | 0.03  (0.01-0.06) | 0.50  (0.02-0.98) | 0.68  (0.52-0.86) | 15.00  (14.81-15.19) | 15.00  (14.82-15.20) | 15.00  (14.80-15.20) | 25.17  (24.90-25.38) | 0.20  (0.00-0.64) | 0.66  (0.51-0.86) | 0.76  (0.10-1.00) | 1.01  (1.00-1.07) | 0.75  (0.51-0.99) | 0.73  (0.51-0.99) | 0.75  (0.52-0.99) | 0.53  (0.50-0.69) | 0.75  (0.51-0.99) | 0.91  (0.72-1.00) | 0.86  (0.52-1.00) | 0.54  (0.50-0.75) | 0.08  (0.01-0.10) | 0.05  (0.01-0.10) | 0.04  (0.04-0.05) | 0.11  (0.01-0.20) |
| ssp585 | KALLANG | 0.50  (0.02-0.97) | 0.04  (0.02-0.07) | 0.50  (0.03-0.98) | 0.68  (0.52-0.86) | 15.00  (14.80-15.20) | 15.00  (14.81-15.19) | 15.00  (14.80-15.20) | 25.16  (24.92-25.36) | 0.15  (0.00-0.54) | 0.67  (0.51-0.87) | 0.76  (0.10-1.00) | 1.01  (1.00-1.06) | 0.75  (0.51-0.99) | 0.74  (0.51-0.99) | 0.75  (0.51-0.99) | 0.52  (0.50-0.61) | 0.75  (0.51-0.99) | 0.88  (0.63-1.00) | 0.83  (0.51-1.00) | 0.52  (0.50-0.63) | 0.08  (0.01-0.10) | 0.08  (0.02-0.10) | 0.06  (0.06-0.07) | 0.10  (0.01-0.20) |
| ssp585 | MANDAI | 0.14  (0.01-0.40) | 0.09  (0.03-0.15) | 0.50  (0.03-0.97) | 0.68  (0.52-0.87) | 15.00  (14.81-15.20) | 15.00  (14.81-15.20) | 15.00  (14.81-15.20) | 24.92  (24.73-25.11) | 0.23  (0.01-0.86) | 0.47  (0.32-0.60) | 0.98  (0.88-1.00) | 1.39  (1.04-1.86) | 0.75  (0.51-0.99) | 0.75  (0.51-0.99) | 0.76  (0.52-0.99) | 0.87  (0.55-1.00) | 0.75  (0.51-0.99) | 0.63  (0.50-0.96) | 0.82  (0.56-0.99) | 0.60  (0.50-0.85) | 0.01  (0.01-0.02) | 0.06  (0.03-0.10) | 0.04  (0.04-0.05) | 0.04  (0.00-0.13) |
| ssp585 | MARINE PARADE | 0.52  (0.05-0.97) | 0.24  (0.22-0.27) | 0.49  (0.03-0.97) | 0.68  (0.52-0.87) | 15.00  (14.80-15.20) | 15.00  (14.80-15.20) | 15.00  (14.81-15.19) | 25.11  (24.87-25.35) | 0.14  (0.00-0.59) | 0.67  (0.43-0.88) | 0.55  (0.10-1.00) | 1.03  (1.00-1.12) | 0.75  (0.51-0.99) | 0.73  (0.51-0.99) | 0.75  (0.51-0.99) | 0.59  (0.50-0.78) | 0.75  (0.51-0.99) | 0.91  (0.73-1.00) | 0.78  (0.50-1.00) | 0.75  (0.50-0.99) | 0.05  (0.01-0.10) | 0.04  (0.01-0.10) | 0.03  (0.03-0.04) | 0.11  (0.00-0.20) |
| ssp585 | NOVENA | 0.53  (0.08-0.97) | 0.20  (0.16-0.23) | 0.50  (0.03-0.97) | 0.68  (0.52-0.86) | 15.00  (14.80-15.21) | 15.00  (14.81-15.20) | 15.00  (14.80-15.21) | 25.10  (24.91-25.30) | 0.50  (0.09-0.81) | 0.54  (0.42-0.63) | 0.85  (0.76-0.93) | 1.10  (1.00-1.29) | 0.75  (0.51-0.99) | 0.74  (0.51-0.99) | 0.74  (0.51-0.99) | 0.62  (0.51-0.84) | 0.76  (0.52-0.99) | 0.88  (0.66-1.00) | 0.87  (0.60-1.00) | 0.91  (0.69-1.00) | 0.09  (0.08-0.10) | 0.08  (0.04-0.10) | 0.04  (0.04-0.05) | 0.06  (0.00-0.17) |
| ssp585 | PASIR RIS | 0.54  (0.10-0.97) | 0.13  (0.12-0.15) | 0.50  (0.02-0.97) | 0.68  (0.52-0.87) | 15.00  (14.80-15.20) | 15.00  (14.81-15.20) | 15.00  (14.81-15.18) | 25.11  (24.91-25.31) | 0.27  (0.01-0.74) | 0.60  (0.50-0.70) | 0.89  (0.74-0.99) | 1.02  (1.00-1.08) | 0.75  (0.51-0.99) | 0.74  (0.51-0.99) | 0.74  (0.51-0.99) | 0.55  (0.50-0.68) | 0.75  (0.51-0.99) | 0.89  (0.67-1.00) | 0.90  (0.70-1.00) | 0.62  (0.50-0.93) | 0.10  (0.09-0.10) | 0.06  (0.02-0.10) | 0.04  (0.04-0.04) | 0.08  (0.00-0.19) |
| ssp585 | QUEENSTOWN | 0.09  (0.01-0.23) | 0.09  (0.02-0.13) | 0.51  (0.02-0.98) | 0.68  (0.52-0.88) | 15.00  (14.81-15.20) | 15.00  (14.81-15.20) | 15.00  (14.80-15.19) | 25.12  (24.93-25.30) | 0.34  (0.01-0.81) | 0.60  (0.49-0.68) | 0.93  (0.84-0.99) | 1.02  (1.00-1.06) | 0.75  (0.51-0.99) | 0.74  (0.51-0.99) | 0.74  (0.51-0.99) | 0.54  (0.50-0.62) | 0.75  (0.51-0.99) | 0.91  (0.73-1.00) | 0.93  (0.78-1.00) | 0.63  (0.50-0.96) | 0.10  (0.09-0.10) | 0.06  (0.03-0.10) | 0.05  (0.05-0.05) | 0.08  (0.00-0.19) |
| ssp585 | SEMBAWANG | 0.04  (0.00-0.10) | 0.05  (0.03-0.07) | 0.50  (0.03-0.97) | 0.68  (0.53-0.86) | 15.00  (14.81-15.19) | 15.01  (14.81-15.20) | 14.99  (14.81-15.18) | 25.01  (24.82-25.18) | 0.27  (0.01-0.69) | 0.54  (0.43-0.63) | 0.97  (0.89-1.00) | 1.08  (1.00-1.26) | 0.75  (0.51-0.99) | 0.75  (0.51-0.99) | 0.74  (0.51-0.98) | 0.76  (0.52-0.97) | 0.76  (0.52-0.98) | 0.81  (0.54-0.99) | 0.90  (0.70-1.00) | 0.66  (0.51-0.93) | 0.09  (0.08-0.10) | 0.04  (0.01-0.09) | 0.02  (0.02-0.02) | 0.06  (0.00-0.17) |
| ssp585 | SERANGOON | 0.54  (0.11-0.97) | 0.18  (0.16-0.20) | 0.50  (0.03-0.97) | 0.68  (0.52-0.87) | 15.00  (14.80-15.19) | 15.00  (14.81-15.20) | 15.00  (14.80-15.19) | 25.15  (24.96-25.35) | 0.40  (0.02-0.81) | 0.55  (0.44-0.63) | 0.90  (0.84-0.95) | 1.06  (1.00-1.20) | 0.75  (0.51-0.99) | 0.74  (0.51-0.98) | 0.74  (0.51-0.99) | 0.61  (0.51-0.80) | 0.75  (0.51-0.99) | 0.91  (0.73-1.00) | 0.92  (0.75-1.00) | 0.92  (0.74-1.00) | 0.10  (0.09-0.10) | 0.06  (0.02-0.10) | 0.03  (0.03-0.03) | 0.07  (0.00-0.18) |
| ssp585 | SUNGEI KADUT | 0.00  (0.00-0.02) | 0.00  (0.00-0.01) | 0.50  (0.02-0.98) | 0.68  (0.52-0.87) | 15.00  (14.80-15.19) | 15.00  (14.80-15.20) | 15.00  (14.81-15.20) | 24.96  (24.76-25.16) | 0.47  (0.02-0.88) | 0.55  (0.43-0.66) | 0.93  (0.77-1.00) | 1.11  (1.00-1.38) | 0.75  (0.51-0.99) | 0.75  (0.52-0.99) | 0.75  (0.51-0.99) | 0.74  (0.51-0.99) | 0.75  (0.51-0.99) | 0.67  (0.50-0.97) | 0.76  (0.51-0.99) | 0.63  (0.50-0.94) | 0.05  (0.01-0.10) | 0.06  (0.01-0.10) | 0.03  (0.03-0.03) | 0.05  (0.00-0.16) |
| ssp585 | TANGLIN | 0.00  (0.00-0.01) | 0.00  (0.00-0.01) | 0.50  (0.02-0.98) | 0.68  (0.53-0.87) | 15.00  (14.81-15.19) | 15.00  (14.81-15.20) | 15.00  (14.81-15.19) | 24.98  (24.77-25.17) | 0.40  (0.02-0.89) | 0.56  (0.43-0.66) | 0.98  (0.90-1.00) | 1.11  (1.00-1.36) | 0.75  (0.51-0.99) | 0.75  (0.51-0.99) | 0.75  (0.51-0.99) | 0.69  (0.50-0.98) | 0.75  (0.51-0.99) | 0.71  (0.51-0.98) | 0.85  (0.59-1.00) | 0.64  (0.50-0.96) | 0.07  (0.04-0.10) | 0.05  (0.01-0.10) | 0.05  (0.04-0.05) | 0.06  (0.00-0.17) |
| ssp585 | TOA PAYOH | 0.48  (0.01-0.97) | 0.02  (0.00-0.06) | 0.51  (0.02-0.97) | 0.68  (0.51-0.87) | 15.00  (14.80-15.20) | 15.00  (14.81-15.20) | 15.00  (14.80-15.21) | 25.18  (24.99-25.37) | 0.20  (0.01-0.57) | 0.61  (0.50-0.69) | 0.99  (0.96-1.00) | 1.01  (1.00-1.03) | 0.75  (0.51-0.99) | 0.75  (0.51-0.99) | 0.75  (0.51-0.99) | 0.52  (0.50-0.56) | 0.74  (0.51-0.99) | 0.89  (0.68-1.00) | 0.94  (0.81-1.00) | 0.52  (0.50-0.58) | 0.10  (0.09-0.10) | 0.09  (0.07-0.10) | 0.06  (0.05-0.06) | 0.08  (0.00-0.19) |

**2. Validation of model robustness and predictive accuracy**

**2.1 Cross validation on original model** To test model predictive accuracy, we split each location’s recorded mosquito abundance data into training and testing sets (70%, 30%). The model was calibrated to the training set and mosquito abundance was projected forward in time to assess concordance with the test-set observations. The mean of the predicted abundance and the ground truth during the testing period was calculated, as well as the root mean square error (RMSE) and mean absolute error (MAE) (Figure S1–S10). This assessment check wass done using the GFDL-ESM4 climate model, which was the main climate model used to obtain the primary results of the study.

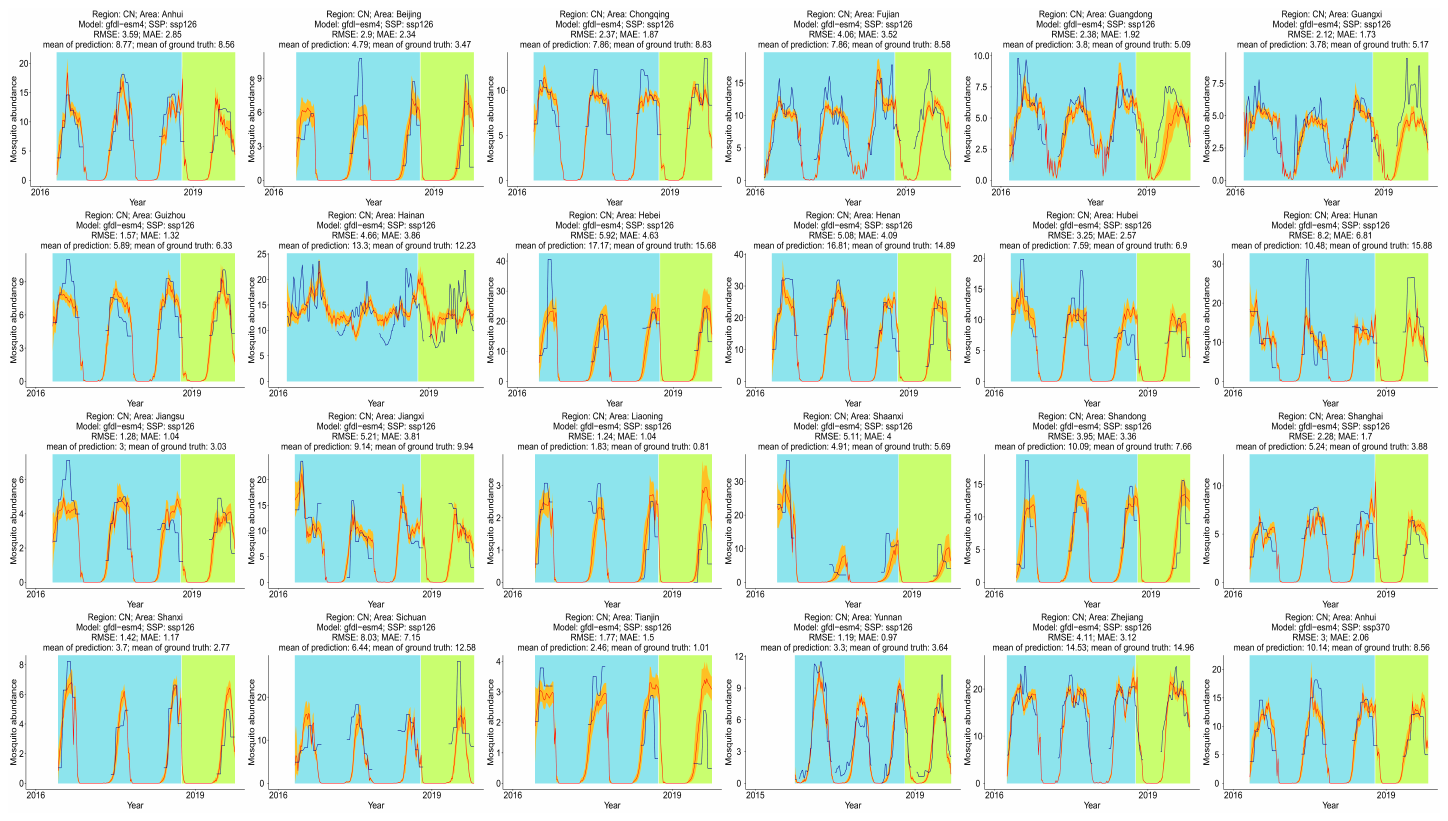

**Figure S1. Fitted and predicted values of mosquito abundance from the calibrated model by area in mainland China (1 of 3).** The observed data is shown in black, while the model projection was visualized in the red line with the yellow band representing the 95% Crl. The model was calibrated on the first 70% of observed data and is represented in the blue shaded region. The model was then validated on the remaining 30% of test data represented in the green shaded area. Root mean square error (RMSE) and mean square error (MSE), as well as the mean of the predicted and observed values for the test set were reported in each panel.

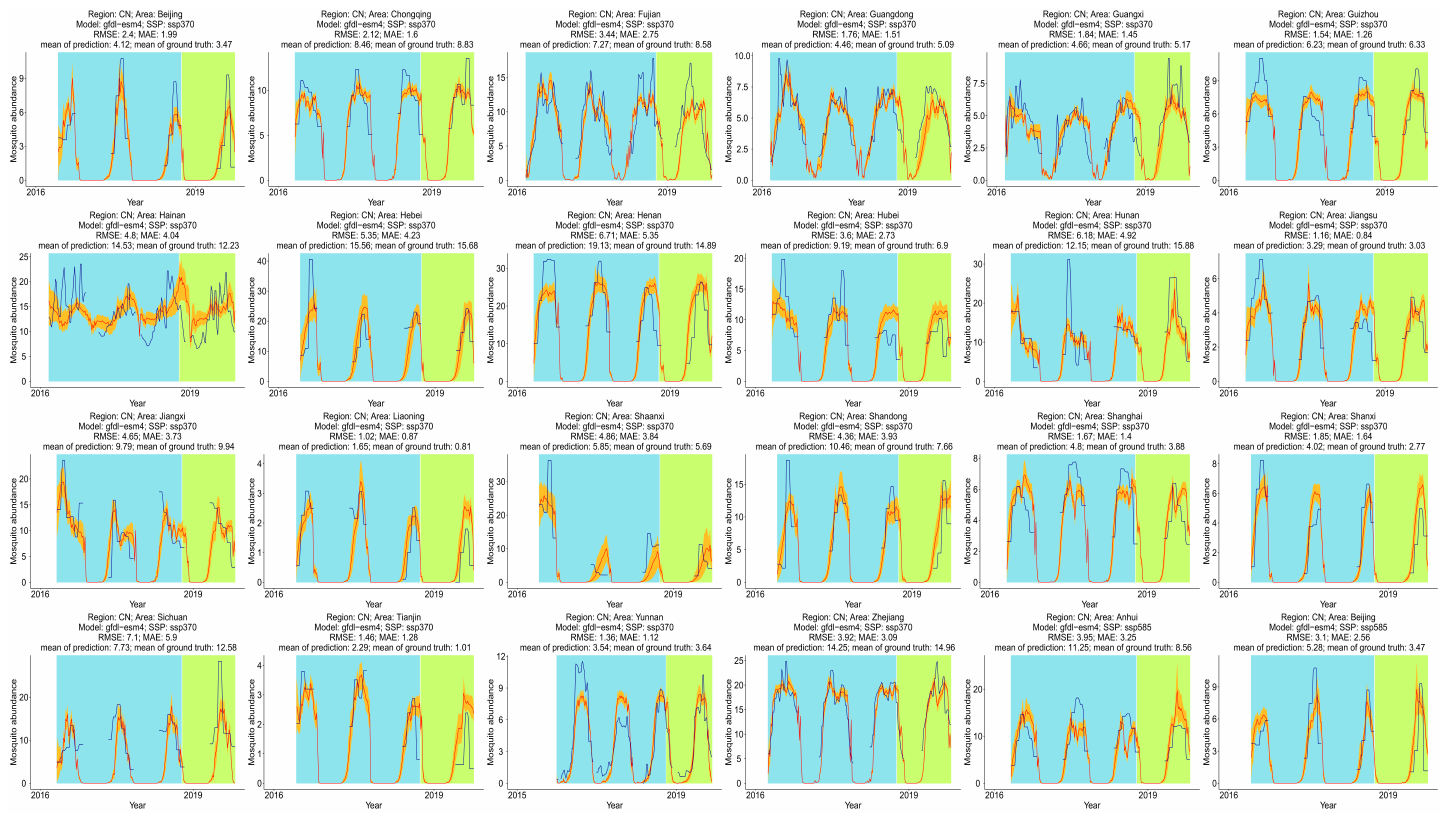

**Figure S2. Fitted and predicted values of mosquito abundance from the calibrated model by area in mainland China (2 of 3).** The observed data is shown in black, while the model projection was visualized in the red line with the yellow band representing the 95% Crl. The model was calibrated on the first 70% of observed data and is represented in the blue shaded region. The model was then validated on the remaining 30% of test data represented in the green shaded area. Root mean square error (RMSE) and mean square error (MSE), as well as the mean of the predicted and observed values for the test set were reported in each panel.

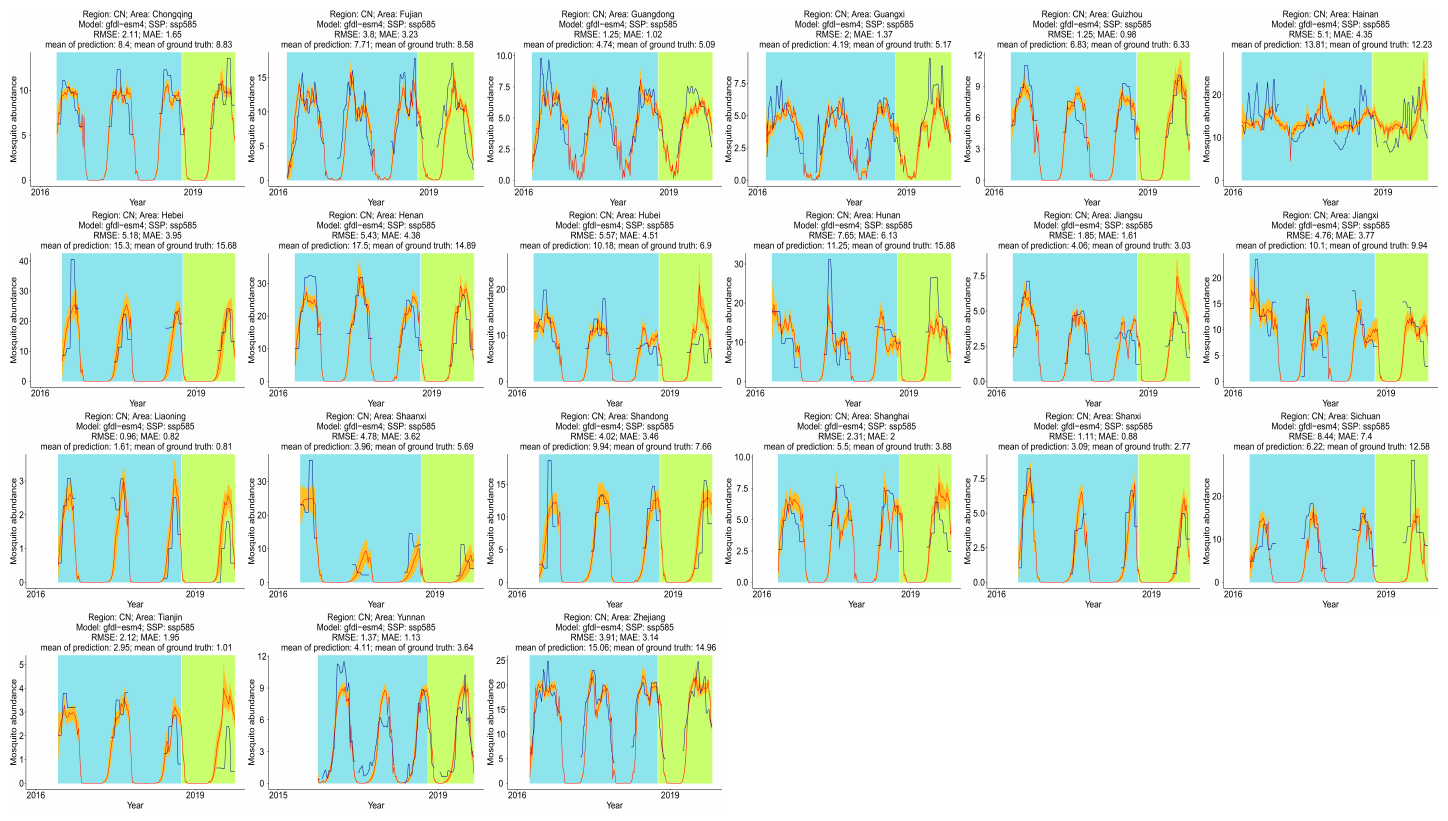

**Figure S3. Fitted and predicted values of mosquito abundance from the calibrated model by area in mainland China (3 of 3).** The observed data is shown in black, while the model projection was visualized in the red line with the yellow band representing the 95% Crl. The model was calibrated on the first 70% of observed data and is represented in the blue shaded region. The model was then validated on the remaining 30% of test data represented in the green shaded area. Root mean square error (RMSE) and mean square error (MSE), as well as the mean of the predicted and observed values for the test set were reported in each panel.

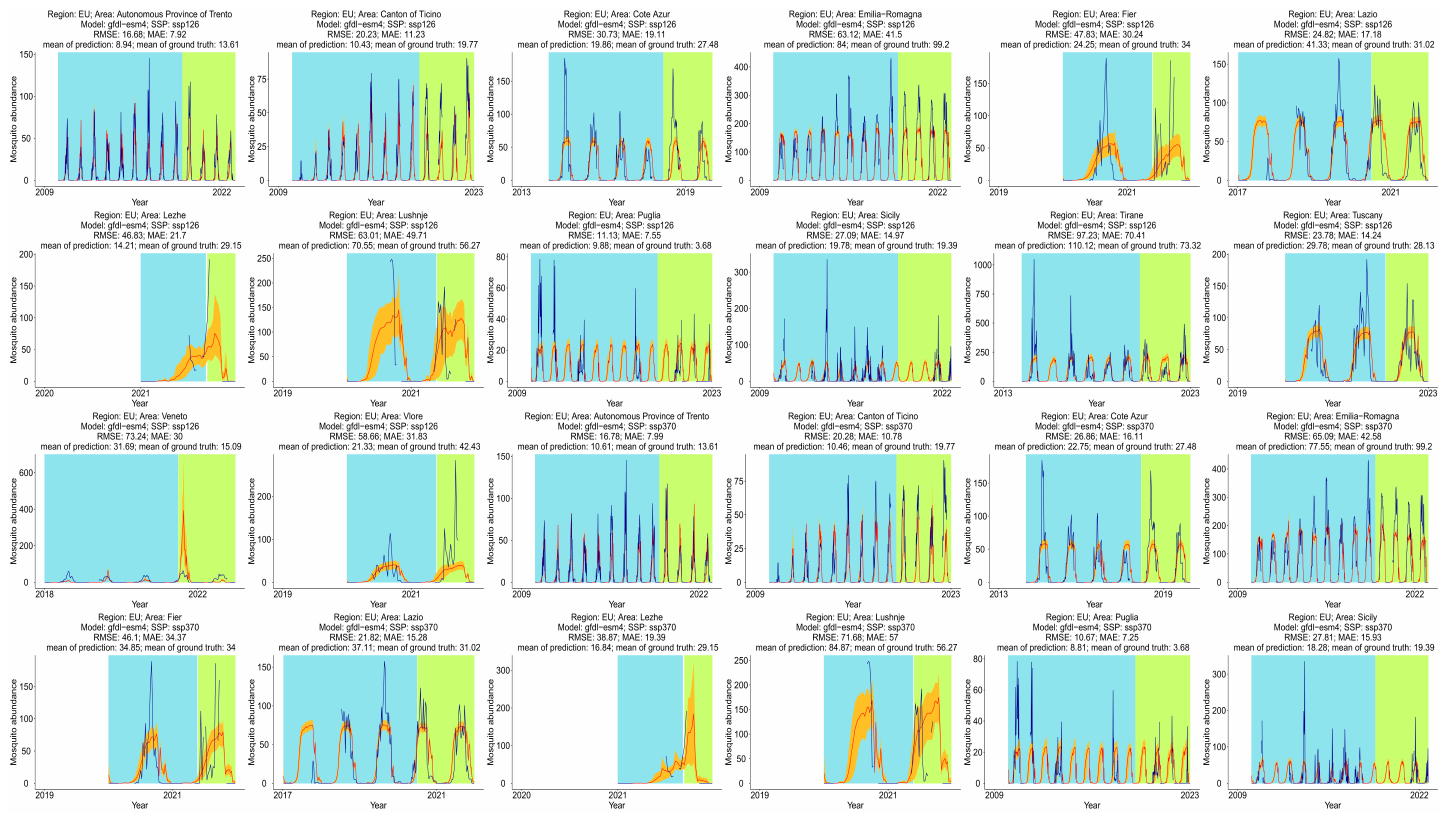

**Figure S4. Fitted and predicted values of mosquito abundance from the calibrated model by area in Europe (1 of 2).** The observed data is shown in black, while the model projection was visualized in the red line with the yellow band representing the 95% Crl. The model was calibrated on the first 70% of observed data and is represented in the blue shaded region. The model was then validated on the remaining 30% of test data represented in the green shaded area. Root mean square error (RMSE) and mean square error (MSE), as well as the mean of the predicted and observed values for the test set were reported in each panel.

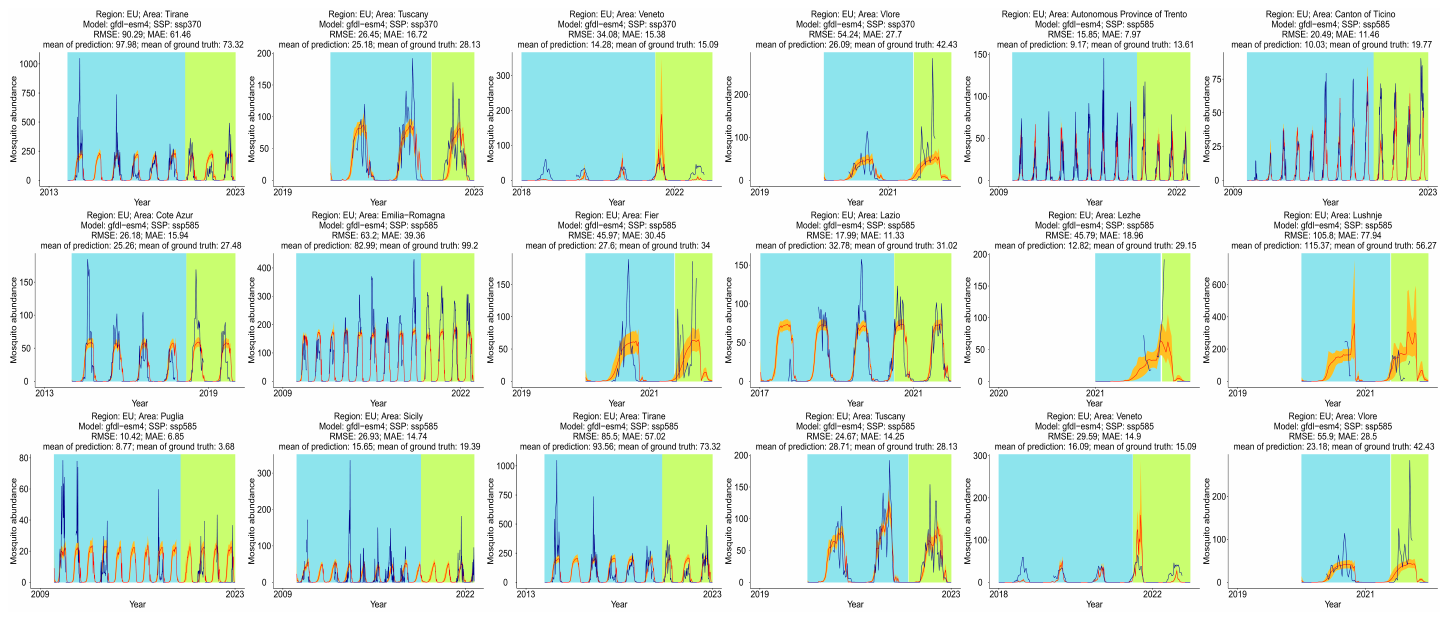

**Figure S5. Fitted and predicted values of mosquito abundance from the calibrated model by area in Europe (2 of 2).** The observed data is shown in black, while the model projection was visualized in the red line with the yellow band representing the 95% Crl. The model was calibrated on the first 70% of observed data and is represented in the blue shaded region. The model was then validated on the remaining 30% of test data represented in the green shaded area. Root mean square error (RMSE) and mean square error (MSE), as well as the mean of the predicted and observed values for the test set were reported in each panel.

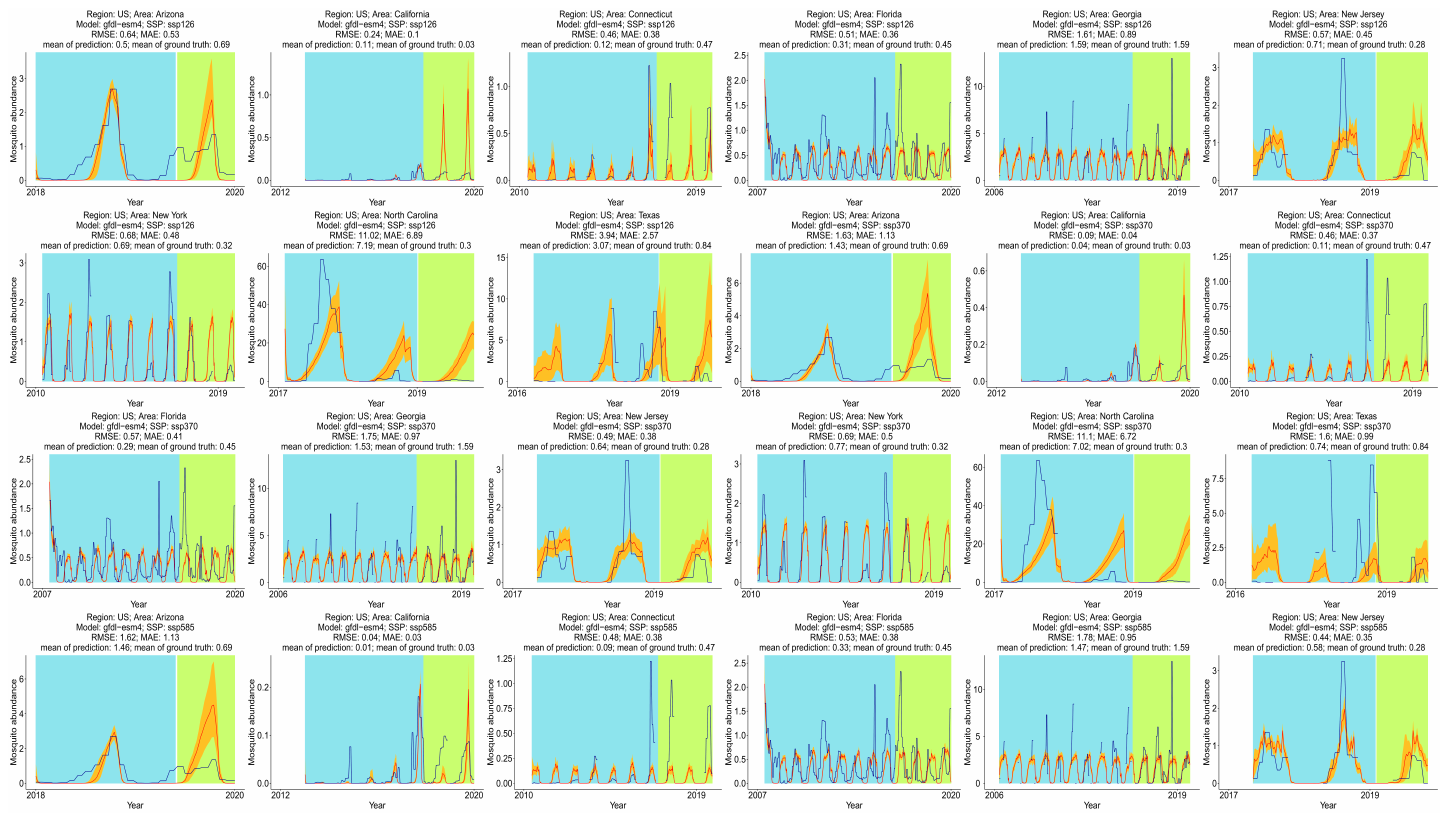

**Figure S6. Fitted and predicted values of mosquito abundance from the calibrated model by area in the United States (1 of 2).** The observed data is shown in black, while the model projection was visualized in the red line with the yellow band representing the 95% Crl. The model was calibrated on the first 70% of observed data and is represented in the blue shaded region. The model was then validated on the remaining 30% of test data represented in the green shaded area. Root mean square error (RMSE) and mean square error (MSE), as well as the mean of the predicted and observed values for the test set were reported in each panel.

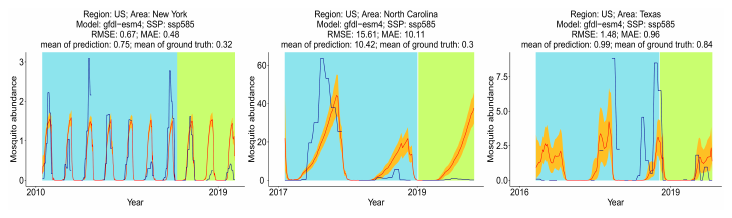

**Figure S7. Fitted and predicted values of mosquito abundance from the calibrated model by area in the United States (2 of 2).** The observed data is shown in black, while the model projection was visualized in the red line with the yellow band representing the 95% Crl. The model was calibrated on the first 70% of observed data and is represented in the blue shaded region. The model was then validated on the remaining 30% of test data represented in the green shaded area. Root mean square error (RMSE) and mean square error (MSE), as well as the mean of the predicted and observed values for the test set were reported in each panel.

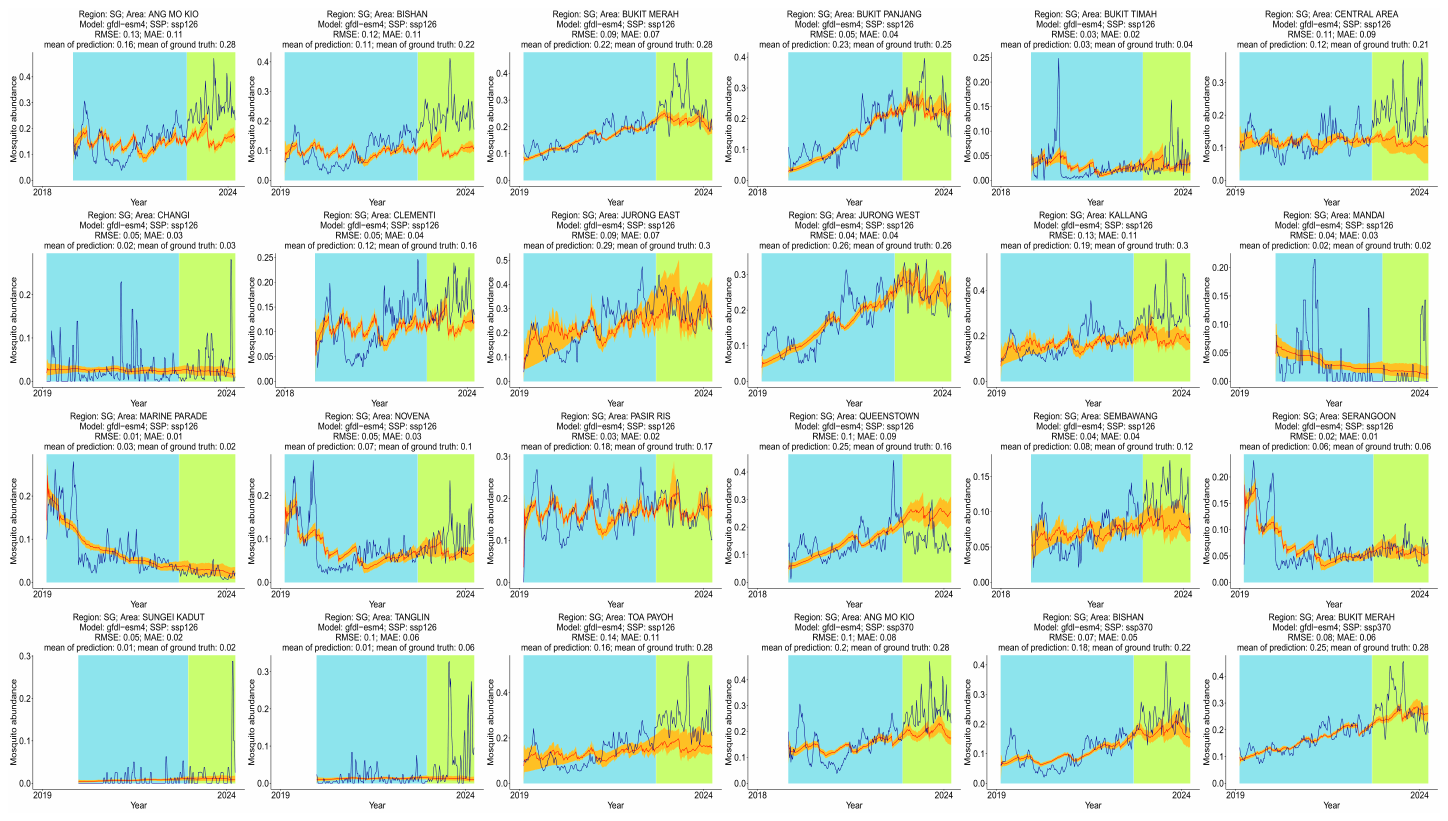

**Figure S8. Fitted and predicted values of mosquito abundance from the calibrated model by area in Singapore (1 of 3).** The observed data is shown in black, while the model projection was visualized in the red line with the yellow band representing the 95% Crl. The model was calibrated on the first 70% of observed data and is represented in the blue shaded region. The model was then validated on the remaining 30% of test data represented in the green shaded area. Root mean square error (RMSE) and mean square error (MSE), as well as the mean of the predicted and observed values for the test set were reported in each panel.

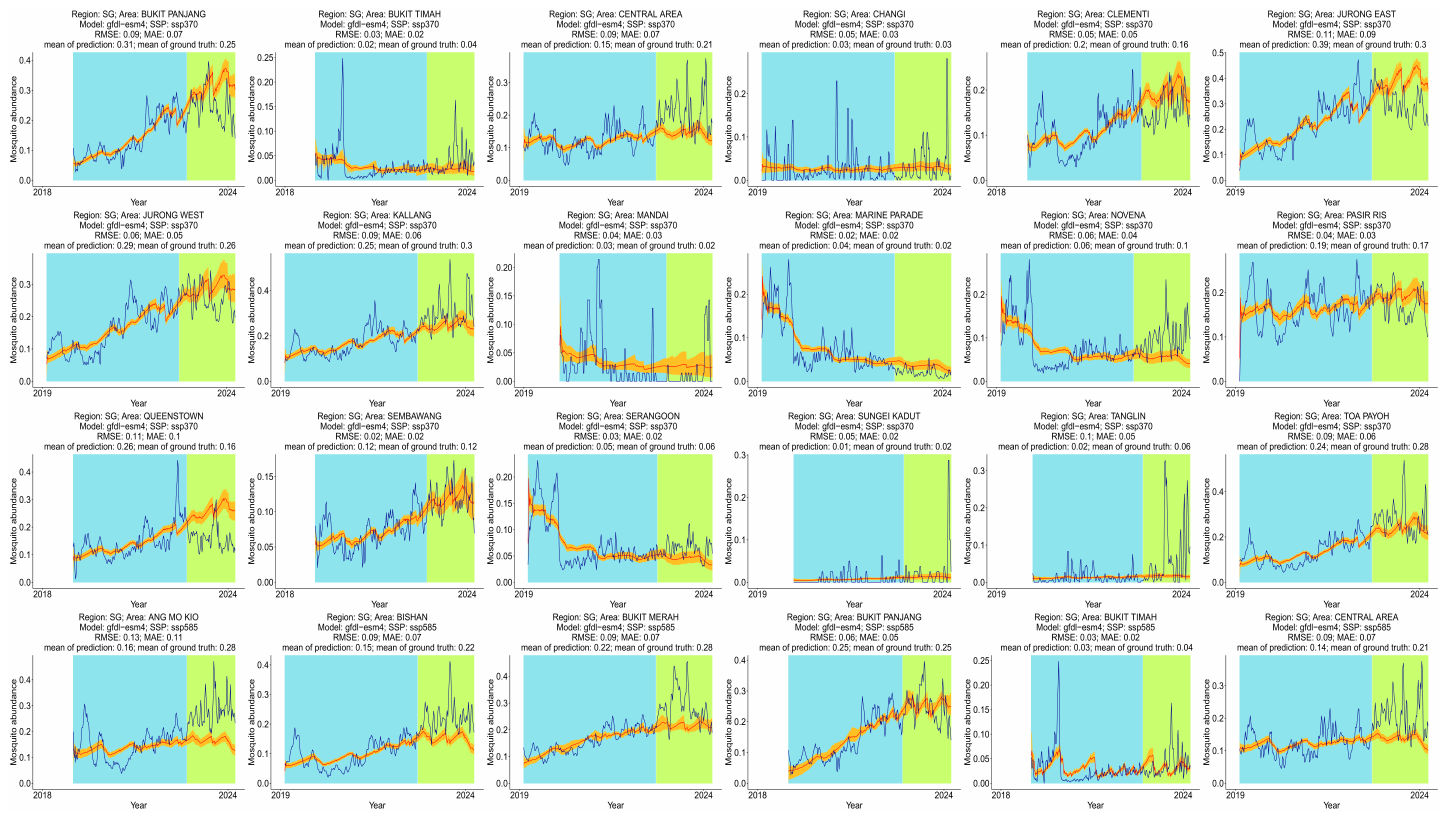

**Figure S9. Fitted and predicted values of mosquito abundance from the calibrated model by area in Singapore (2 of 3).** The observed data is shown in black, while the model projection was visualized in the red line with the yellow band representing the 95% Crl. The model was calibrated on the first 70% of observed data and is represented in the blue shaded region. The model was then validated on the remaining 30% of test data represented in the green shaded area. Root mean square error (RMSE) and mean square error (MSE), as well as the mean of the predicted and observed values for the test set were reported in each panel.

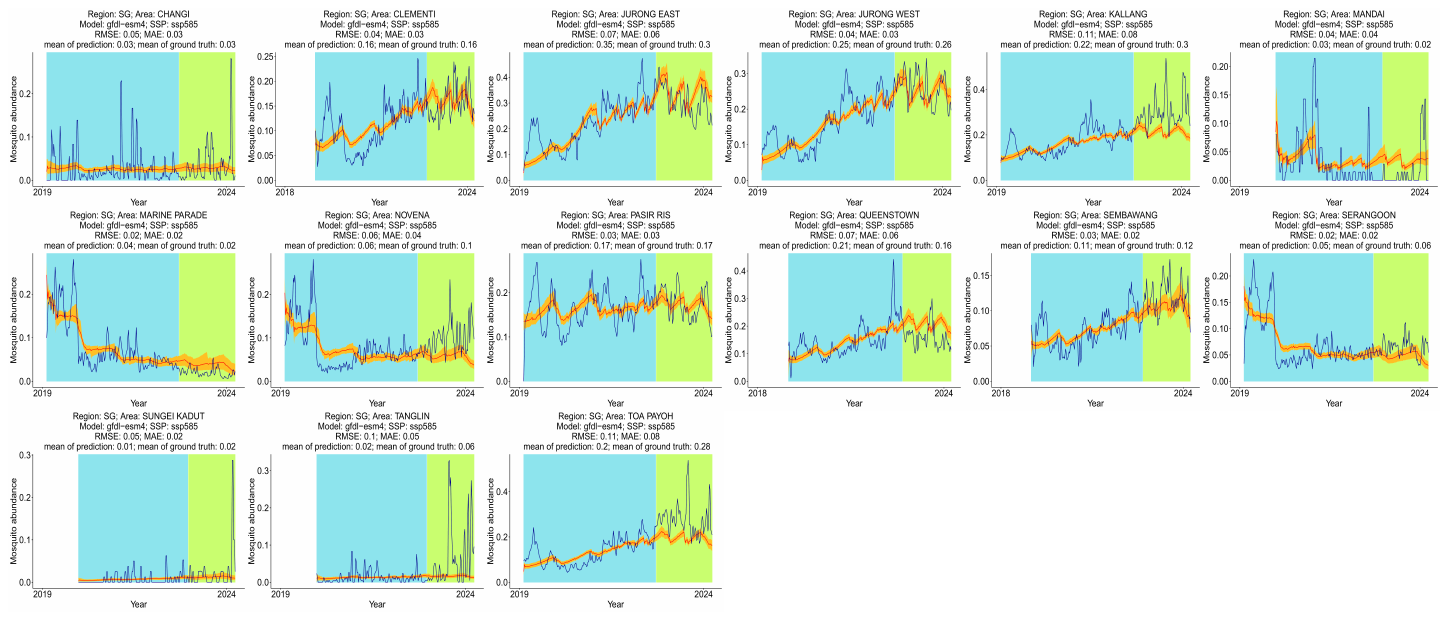

**Figure S10. Fitted and predicted values of mosquito abundance from the calibrated model by area in Singapore (3 of 3).** The observed data is shown in black, while the model projection was visualized in the red line with the yellow band representing the 95% Crl. The model was calibrated on the first 70% of observed data and is represented in the blue shaded region. The model was then validated on the remaining 30% of test data represented in the green shaded area. Root mean square error (RMSE) and mean square error (MSE), as well as the mean of the predicted and observed values for the test set were reported in each panel.

**2.2 Cross Validation on alternative likelihood function** To understand if the specification of the likelihood influences calibration results, we amended the specification of the observation process as an additional sensitivity analysis. As mosquito abundance data are positive and right-skewed, we modelled the alternative observation process of weekly mosquito abundance $Z_{S}\left( t \right)$ using a lognormal distribution:

$$\begin{aligned} Z_{S}\left( t \right)\sim Lognormal\left( {log(Y}_{S}\left( t \right)),\sigma^{2} \right) \end{aligned}$$

where $Y_{S}\left( t \right)$ represents the simulated abundance generated by the process-based model and $\sigma$ characterises observation variability. A small constant (${10}^{-6}$) was added to both observed and simulated abundance values to avoid numerical instability when abundance approached zero.

To test model predictive accuracy, we split each location’s recorded mosquito abundance data into training and testing sets (70%, 30%). The model was calibrated to the training set and mosquito abundance was projected forward in time to assess concordance with the test-set observations. The mean of the predicted abundance and the ground truth during the testing period was calculated, as well as the root mean square error (RMSE) and mean absolute error (MAE) (Figure S11–S20). This assessment check was done using the GFDL-ESM4 climate model, which was the main climate model used to obtain the primary results of the study.

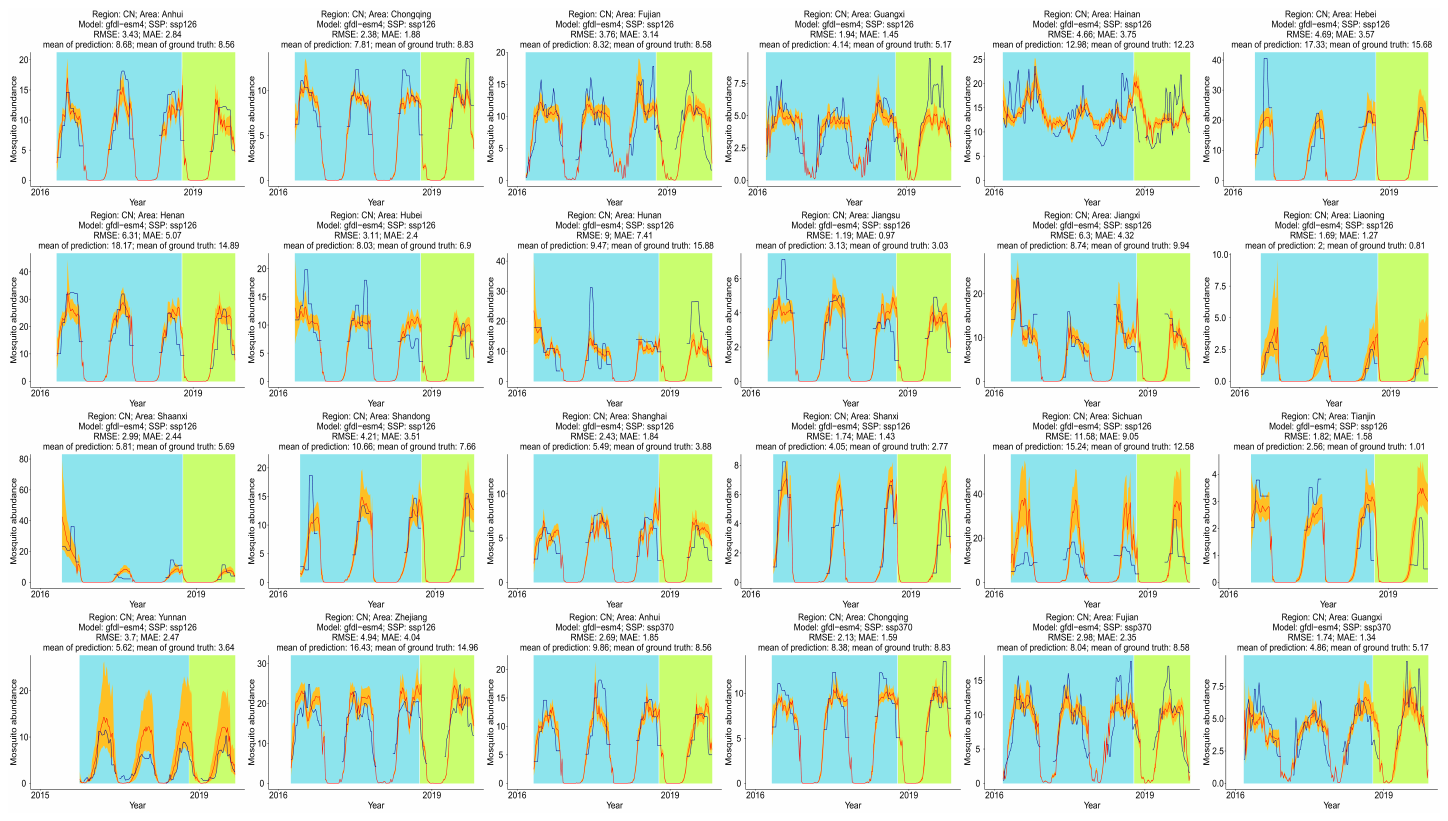

**Figure S11. Fitted and predicted values of mosquito abundance from lognormal likelihood model by area in mainland China (1 of 3).** The observed data is shown in black, while the model projection was visualized in the red line with the yellow band representing the 95% Crl. The model was calibrated on the first 70% of observed data and is represented in the blue shaded region. The model was then validated on the remaining 30% of test data represented in the green shaded area. Root mean square error (RMSE) and mean square error (MSE), as well as the mean of the predicted and observed values for the test set were reported in each panel.

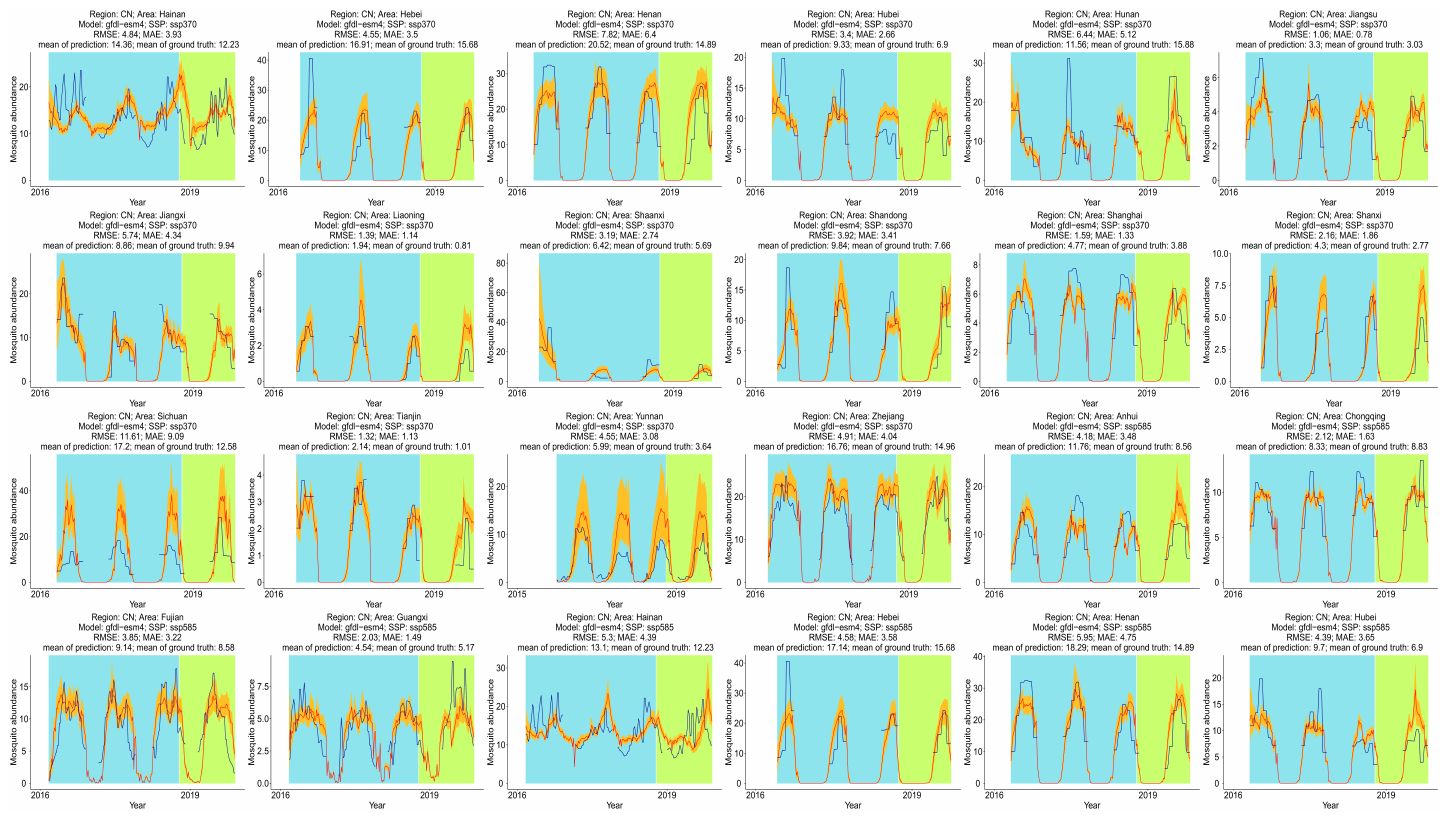

**Figure S12. Fitted and predicted values of mosquito abundance from lognormal likelihood model by area in mainland China (2 of 3).** The observed data is shown in black, while the model projection was visualized in the red line with the yellow band representing the 95% Crl. The model was calibrated on the first 70% of observed data and is represented in the blue shaded region. The model was then validated on the remaining 30% of test data represented in the green shaded area. Root mean square error (RMSE) and mean square error (MSE), as well as the mean of the predicted and observed values for the test set were reported in each panel.

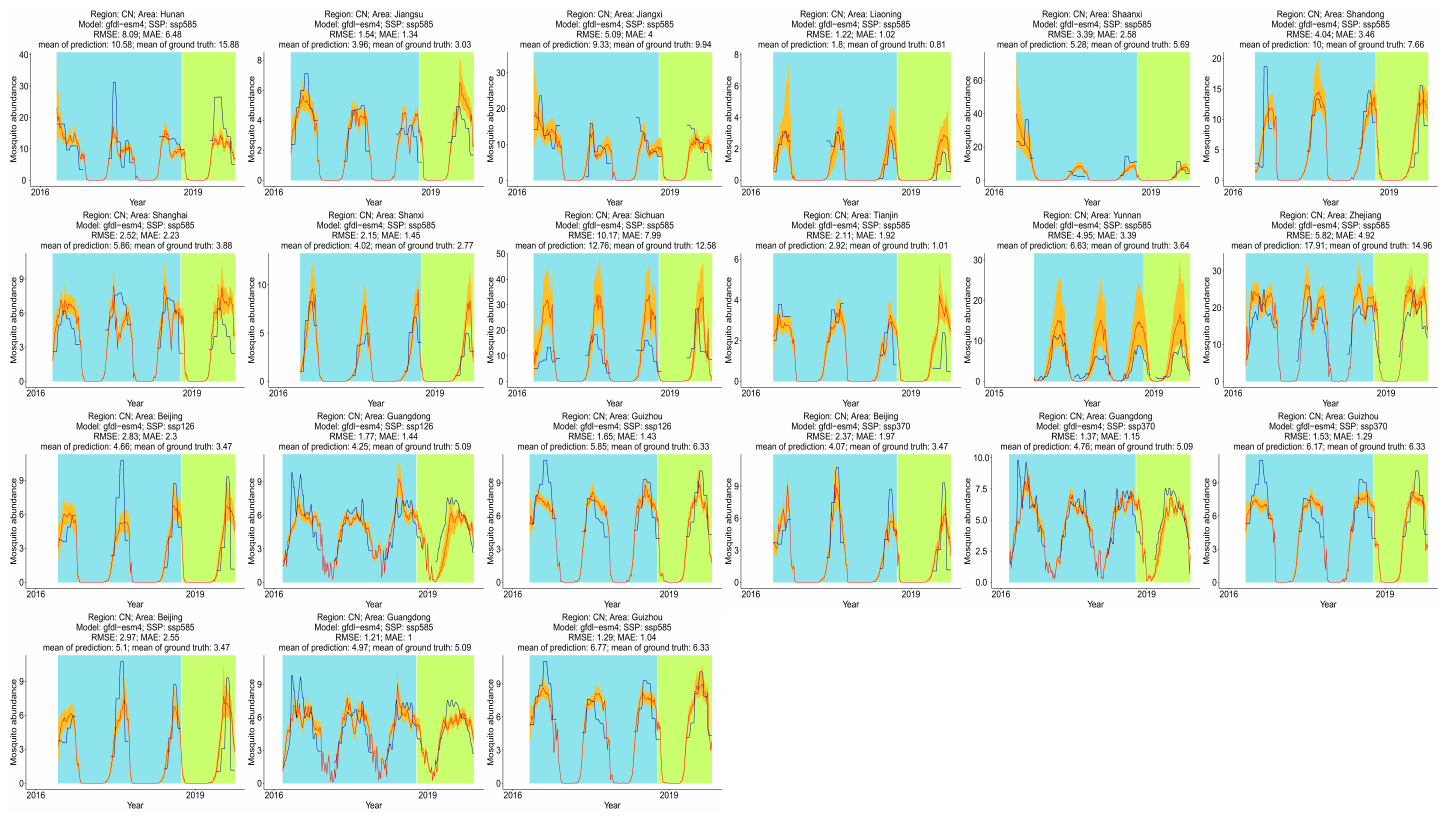

**Figure S13. Fitted and predicted values of mosquito abundance from lognormal likelihood model by area in mainland China (3 of 3).** The observed data is shown in black, while the model projection was visualized in the red line with the yellow band representing the 95% Crl. The model was calibrated on the first 70% of observed data and is represented in the blue shaded region. The model was then validated on the remaining 30% of test data represented in the green shaded area. Root mean square error (RMSE) and mean square error (MSE), as well as the mean of the predicted and observed values for the test set were reported in each panel.

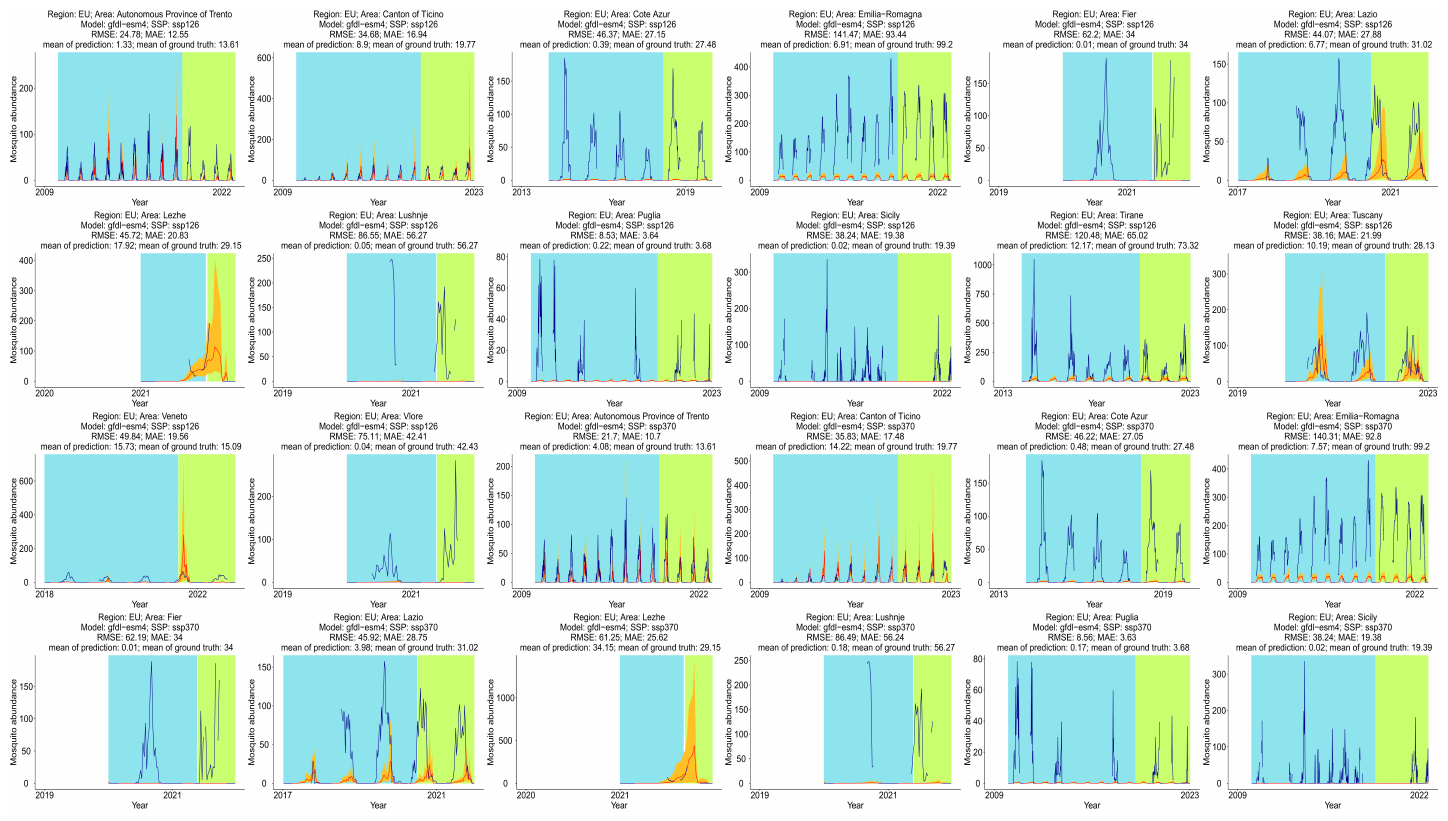

**Figure S14. Fitted and predicted values of mosquito abundance from lognormal likelihood model by area in Europe (1 of 2).** The observed data is shown in black, while the model projection was visualized in the red line with the yellow band representing the 95% Crl. The model was calibrated on the first 70% of observed data and is represented in the blue shaded region. The model was then validated on the remaining 30% of test data represented in the green shaded area. Root mean square error (RMSE) and mean square error (MSE), as well as the mean of the predicted and observed values for the test set were reported in each panel.

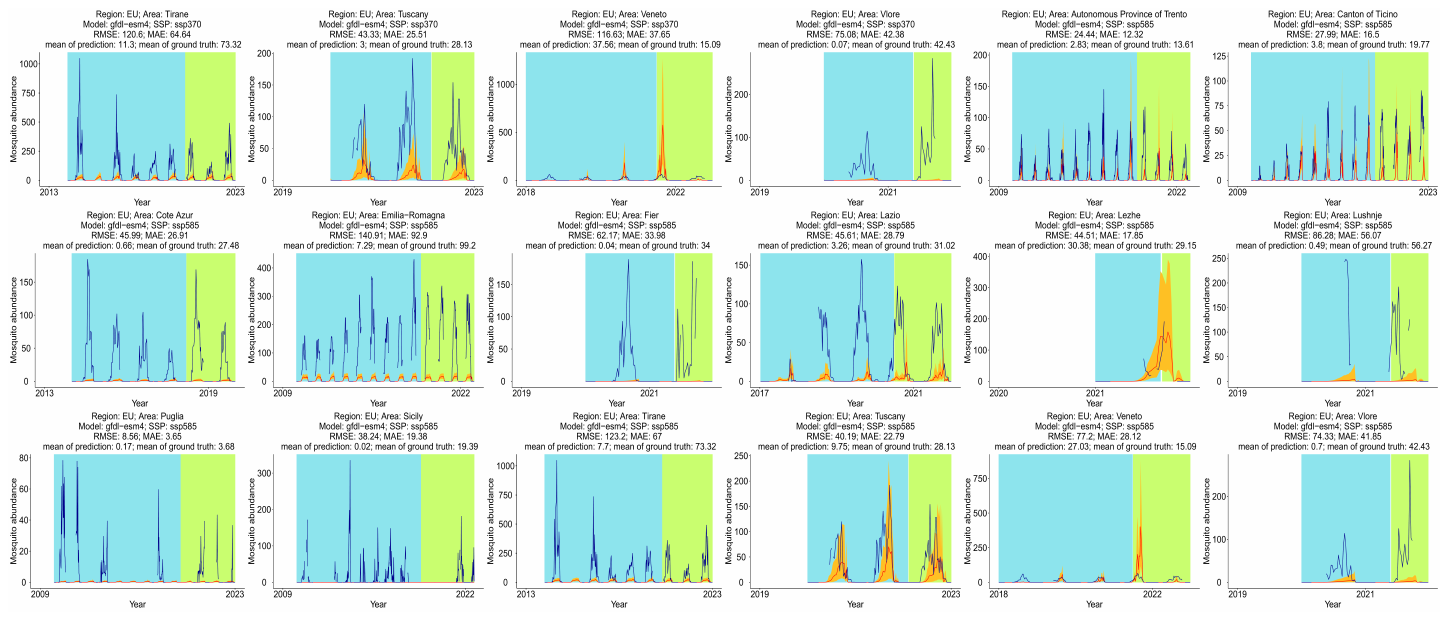

**Figure S15. Fitted and predicted values of mosquito abundance from lognormal likelihood model by area in Europe (2 of 2).** The observed data is shown in black, while the model projection was visualized in the red line with the yellow band representing the 95% Crl. The model was calibrated on the first 70% of observed data and is represented in the blue shaded region. The model was then validated on the remaining 30% of test data represented in the green shaded area. Root mean square error (RMSE) and mean square error (MSE), as well as the mean of the predicted and observed values for the test set were reported in each panel.

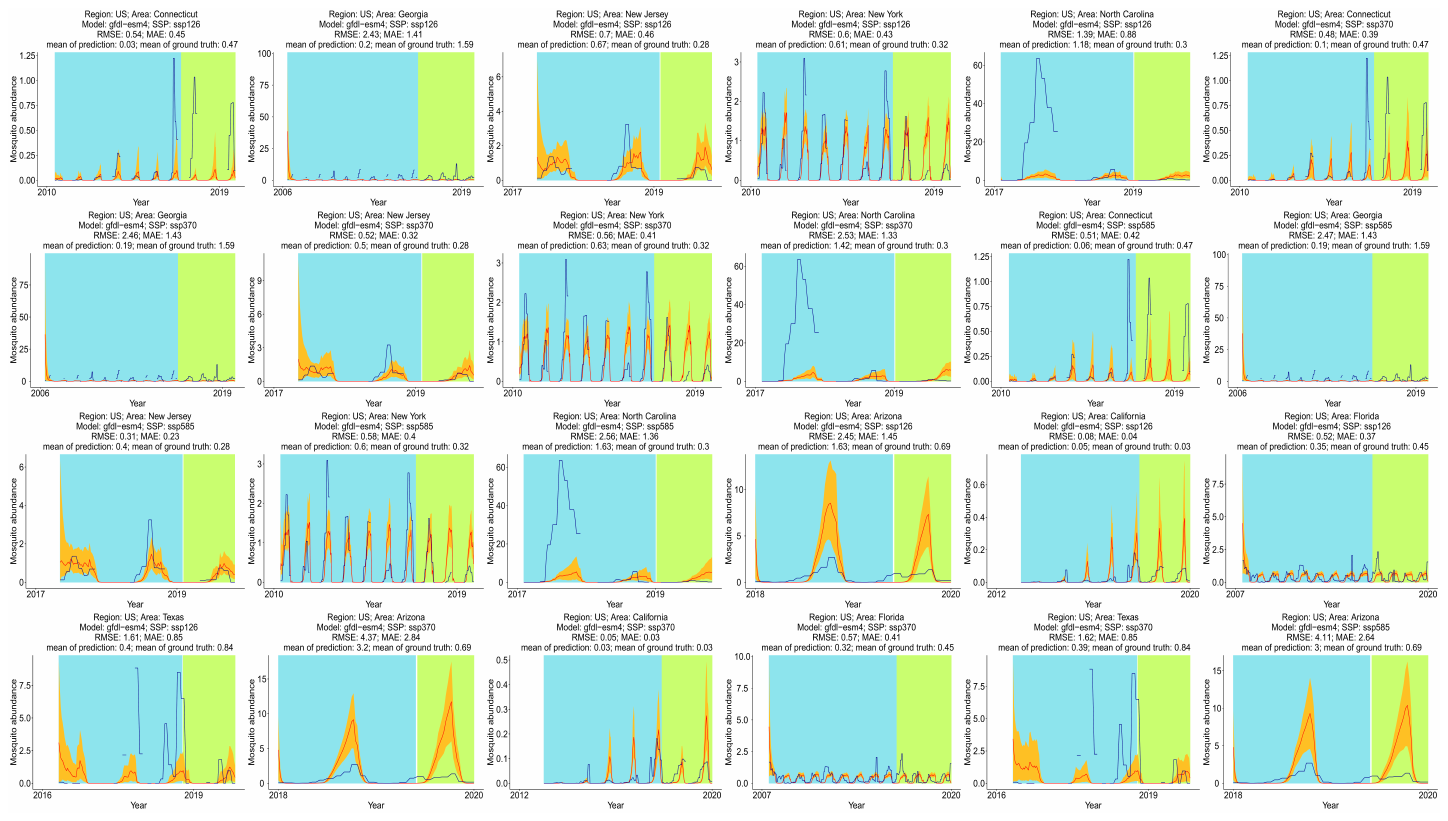

**Figure S16. Fitted and predicted values of mosquito abundance from lognormal likelihood model by area in the United States (1 of 2).** The observed data is shown in black, while the model projection was visualized in the red line with the yellow band representing the 95% Crl. The model was calibrated on the first 70% of observed data and is represented in the blue shaded region. The model was then validated on the remaining 30% of test data represented in the green shaded area. Root mean square error (RMSE) and mean square error (MSE), as well as the mean of the predicted and observed values for the test set were reported in each panel.

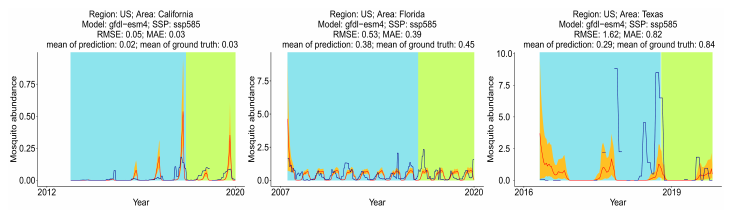

**Figure S17. Fitted and predicted values of mosquito abundance from lognormal likelihood model by area in the United States (2 of 2).** The observed data is shown in black, while the model projection was visualized in the red line with the yellow band representing the 95% Crl. The model was calibrated on the first 70% of observed data and is represented in the blue shaded region. The model was then validated on the remaining 30% of test data represented in the green shaded area. Root mean square error (RMSE) and mean square error (MSE), as well as the mean of the predicted and observed values for the test set were reported in each panel.

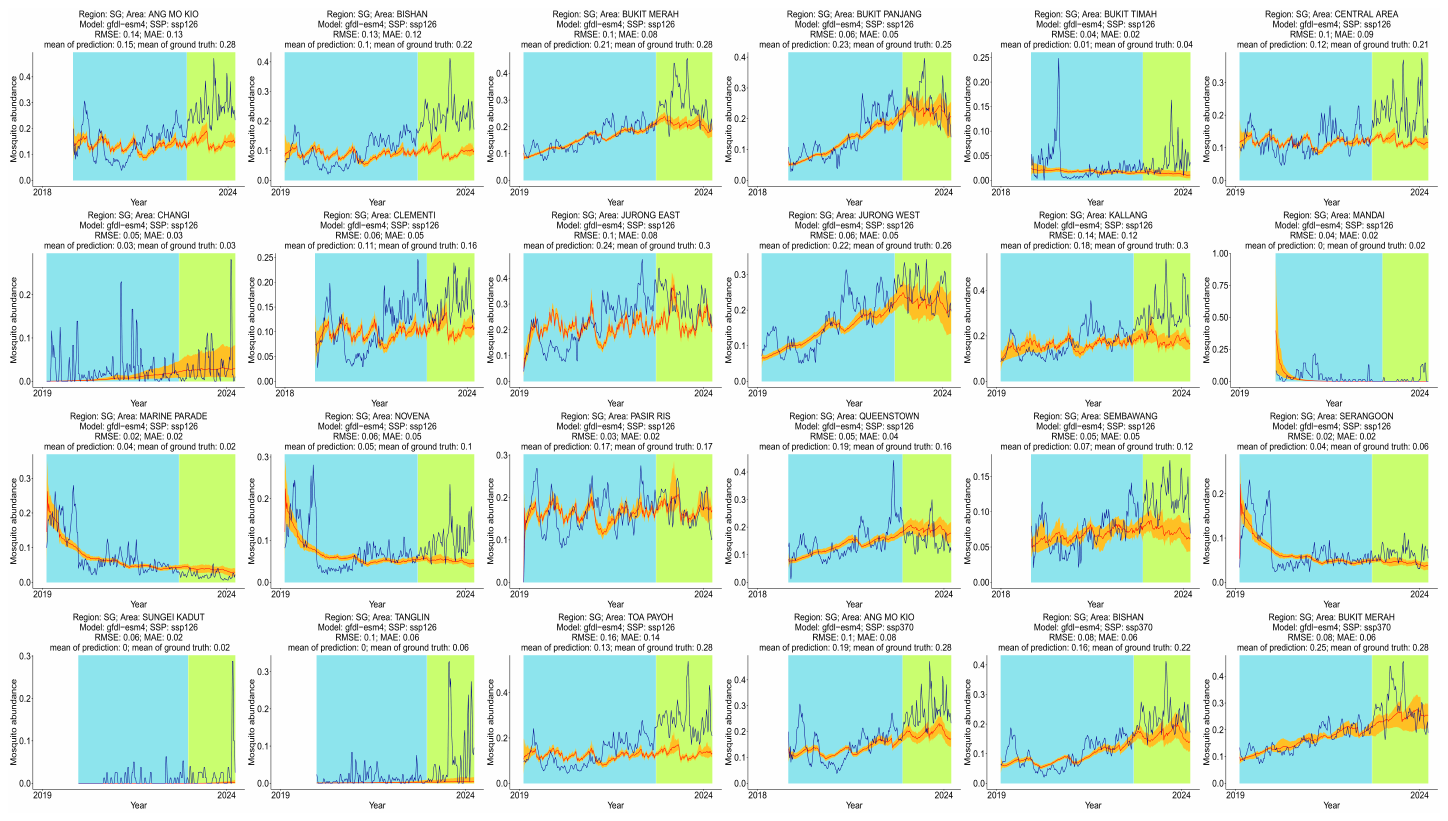

**Figure S18. Fitted and predicted values of mosquito abundance from lognormal likelihood model by area in Singapore (1 of 3).** The observed data is shown in black, while the model projection was visualized in the red line with the yellow band representing the 95% Crl. The model was calibrated on the first 70% of observed data and is represented in the blue shaded region. The model was then validated on the remaining 30% of test data represented in the green shaded area. Root mean square error (RMSE) and mean square error (MSE), as well as the mean of the predicted and observed values for the test set were reported in each panel.

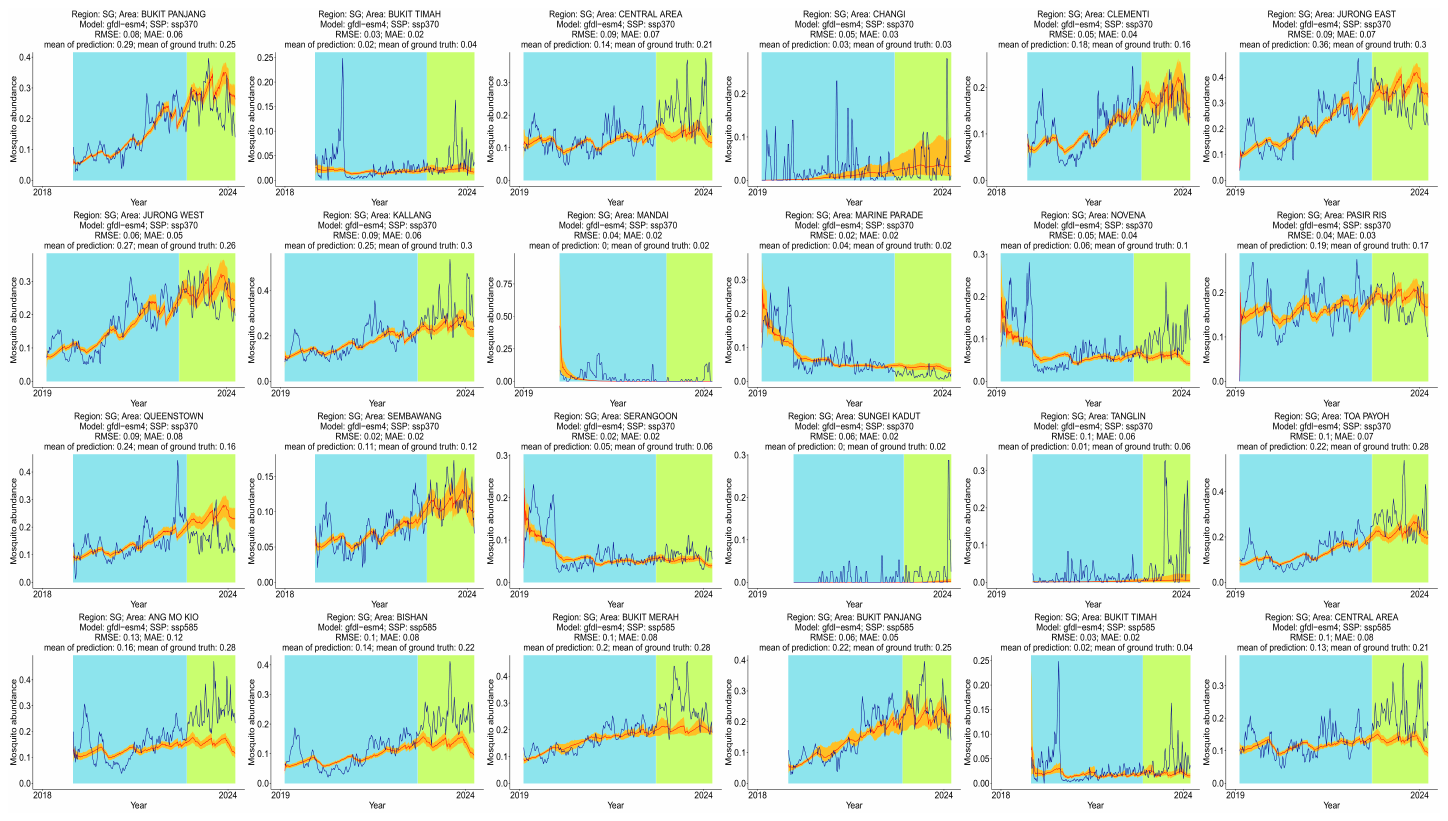

**Figure S19. Fitted and predicted values of mosquito abundance from lognormal likelihood model by area in Singapore (2 of 3).** The observed data is shown in black, while the model projection was visualized in the red line with the yellow band representing the 95% Crl. The model was calibrated on the first 70% of observed data and is represented in the blue shaded region. The model was then validated on the remaining 30% of test data represented in the green shaded area. Root mean square error (RMSE) and mean square error (MSE), as well as the mean of the predicted and observed values for the test set were reported in each panel.

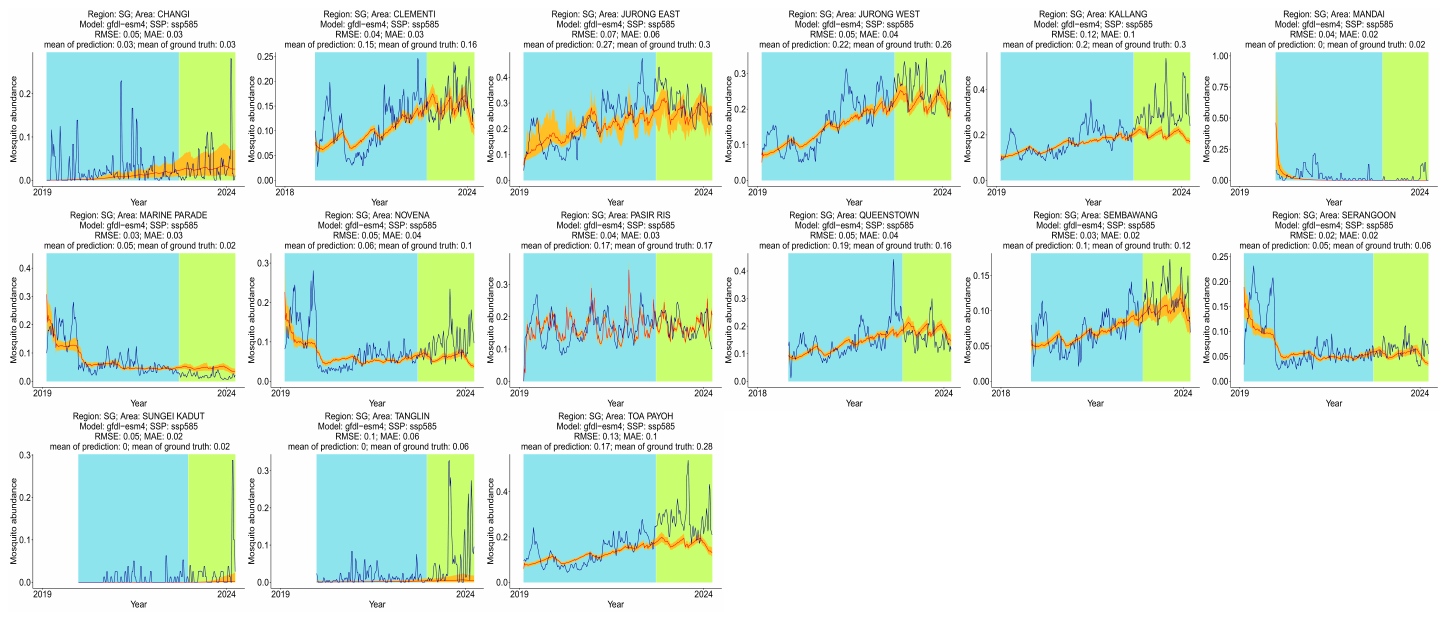

**Figure S20. Fitted and predicted values of mosquito abundance from lognormal likelihood model by area in Singapore (3 of 3).** The observed data is shown in black, while the model projection was visualized in the red line with the yellow band representing the 95% Crl. The model was calibrated on the first 70% of observed data and is represented in the blue shaded region. The model was then validated on the remaining 30% of test data represented in the green shaded area. Root mean square error (RMSE) and mean square error (MSE), as well as the mean of the predicted and observed values for the test set were reported in each panel.

**2.3 Cross validation on alternative carrying capacity function** To understand any potential influence of the specification of aquatic carrying capacity on model inputs, we conducted an additional sensitivity analysis which replaced the original function linking precipitation to the aquatic carrying capacity linearly with a quadratic precipitation function to describe the weekly environmental carrying capacity $f\left( K \right)$ as:

$$f\left( K \right)=\omega+\lambda{P\left( t \right)}^{2}$$

To ascertain model predictive accuracy, we split each location’s recorded mosquito abundance data into training and testing sets (70%, 30%). The model was calibrated to the training set and mosquito abundance was projected forward in time to assess concordance with the test-set observations. The mean of the predicted abundance and the ground truth during the testing period was calculated, as well as the root mean square error (RMSE) and mean absolute error (MAE) (Figure S21–S30). This assessment check was done using the GFDL-ESM4 climate model, which was the main climate model used to obtain the primary results of the study.

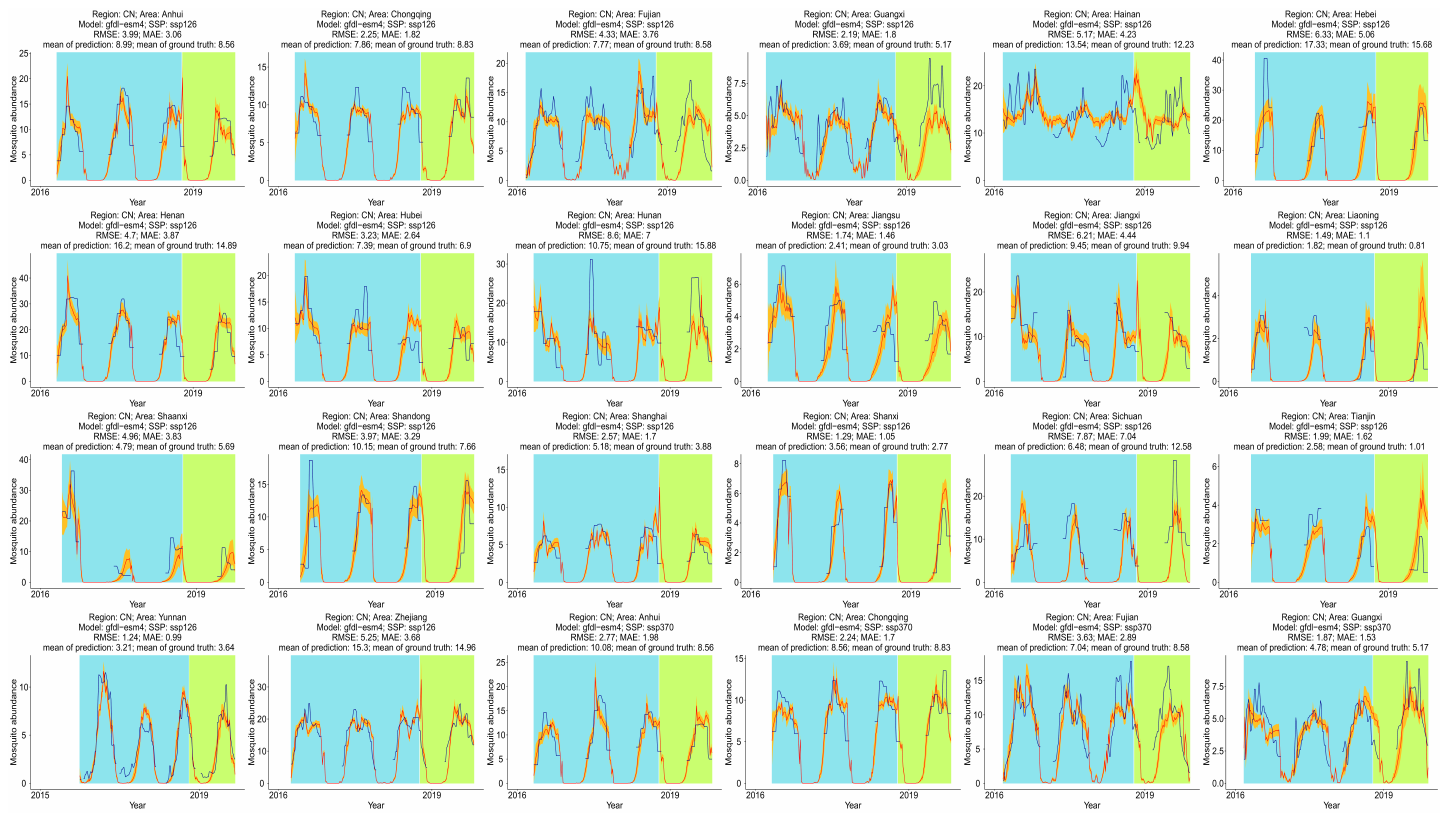

**Figure S21. Fitted and predicted values of mosquito abundance from quadratic precipitation-dependent carrying capacity model by area in mainland China (1 of 3).** The observed data is shown in black, while the model projection was visualized in the red line with the yellow band representing the 95% Crl. The model was calibrated on the first 70% of observed data and is represented in the blue shaded region. The model was then validated on the remaining 30% of test data represented in the green shaded area. Root mean square error (RMSE) and mean square error (MSE), as well as the mean of the predicted and observed values for the test set were reported in each panel.

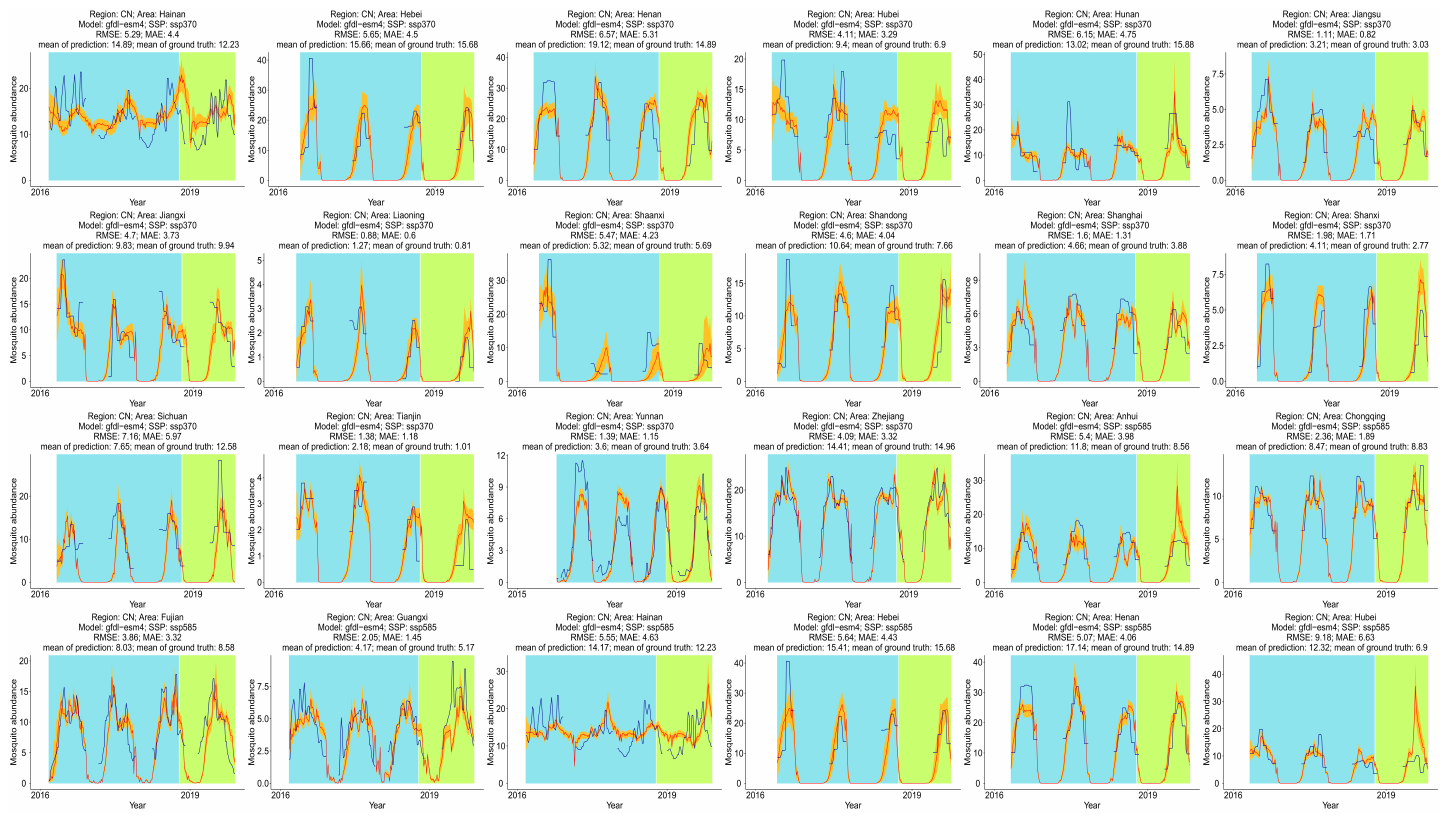

**Figure S22. Fitted and predicted values of mosquito abundance from quadratic precipitation-dependent carrying capacity model by area in mainland China (2 of 3).** The observed data is shown in black, while the model projection was visualized in the red line with the yellow band representing the 95% Crl. The model was calibrated on the first 70% of observed data and is represented in the blue shaded region. The model was then validated on the remaining 30% of test data represented in the green shaded area. Root mean square error (RMSE) and mean square error (MSE), as well as the mean of the predicted and observed values for the test set were reported in each panel.

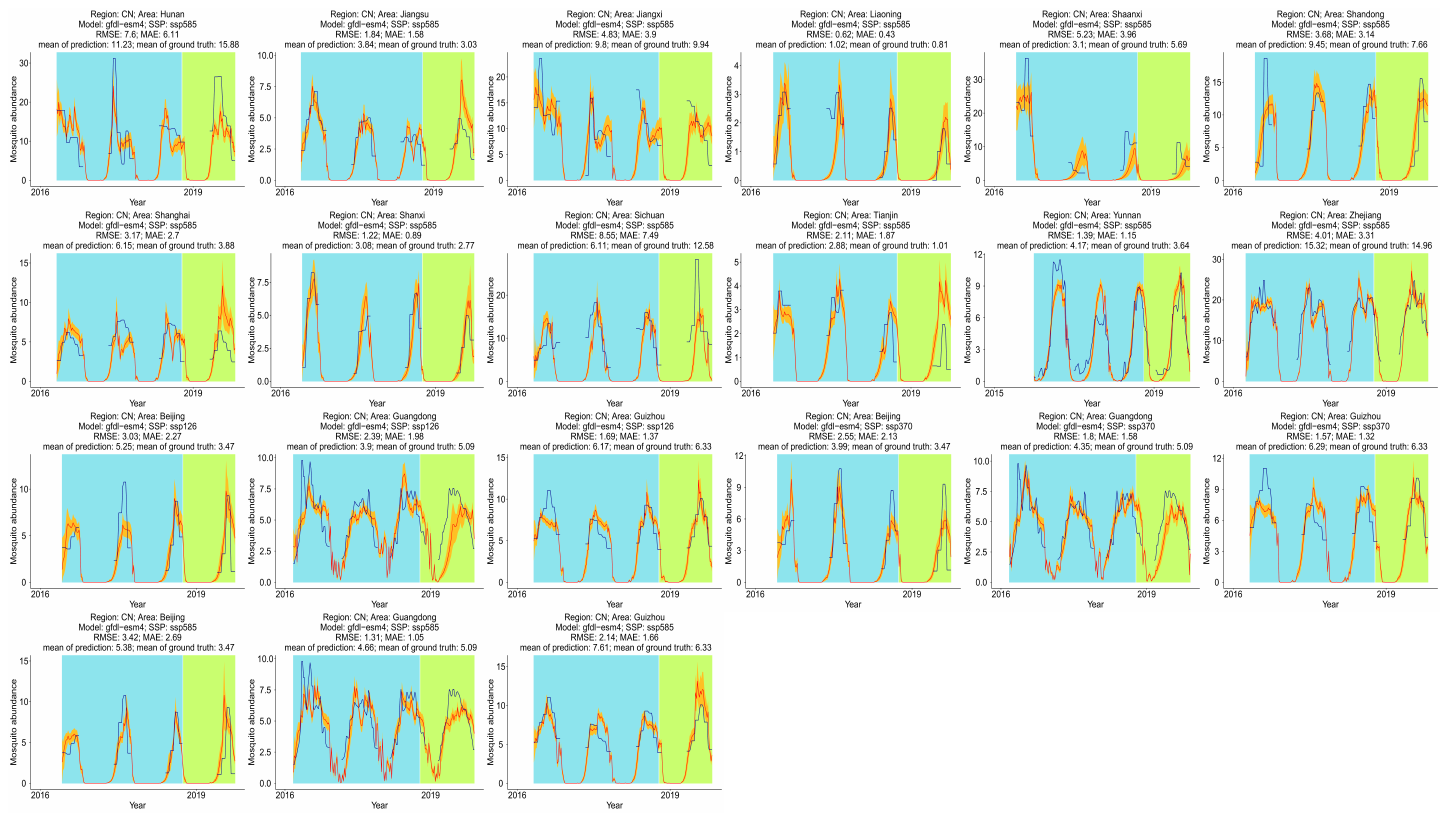

**Figure S23. Fitted and predicted values of mosquito abundance from quadratic precipitation-dependent carrying capacity model by area in mainland China (3 of 3).** The observed data is shown in black, while the model projection was visualized in the red line with the yellow band representing the 95% Crl. The model was calibrated on the first 70% of observed data and is represented in the blue shaded region. The model was then validated on the remaining 30% of test data represented in the green shaded area. Root mean square error (RMSE) and mean square error (MSE), as well as the mean of the predicted and observed values for the test set were reported in each panel.

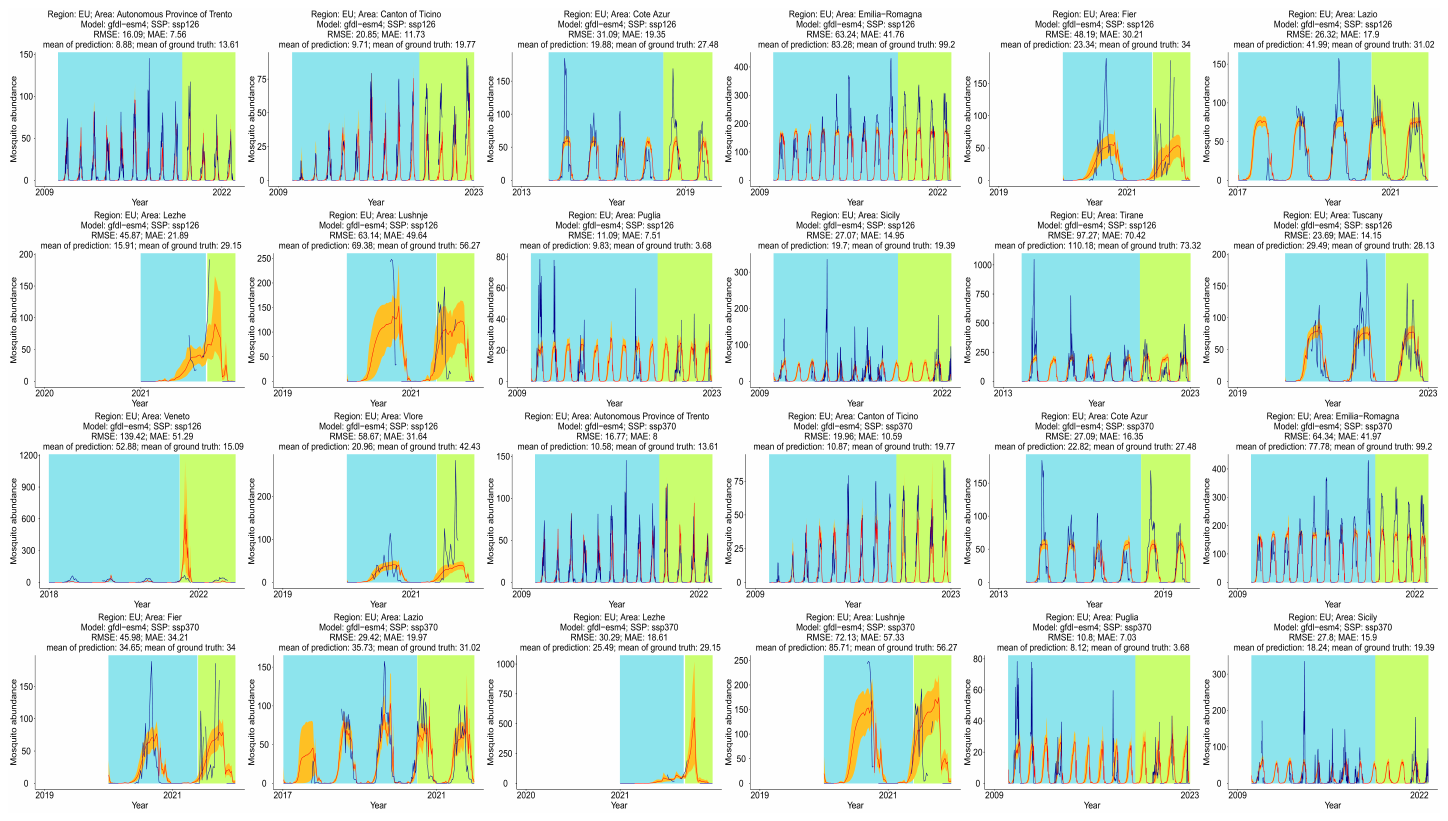

**Figure S24. Fitted and predicted values of mosquito abundance from quadratic precipitation-dependent carrying capacity model by area in Europe (1 of 2).** The observed data is shown in black, while the model projection was visualized in the red line with the yellow band representing the 95% Crl. The model was calibrated on the first 70% of observed data and is represented in the blue shaded region. The model was then validated on the remaining 30% of test data represented in the green shaded area. Root mean square error (RMSE) and mean square error (MSE), as well as the mean of the predicted and observed values for the test set were reported in each panel.

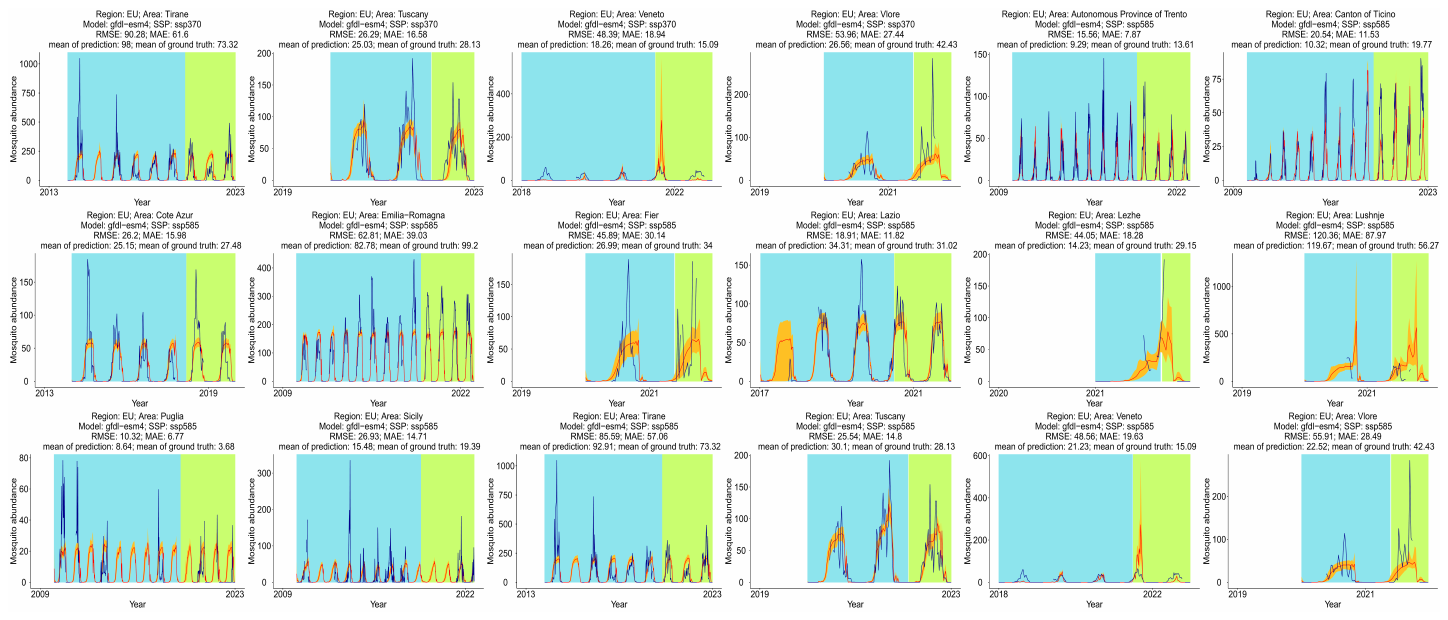

**Figure S25. Fitted and predicted values of mosquito abundance from quadratic precipitation-dependent carrying capacity model by area in Europe (2 of 2).** The observed data is shown in black, while the model projection was visualized in the red line with the yellow band representing the 95% Crl. The model was calibrated on the first 70% of observed data and is represented in the blue shaded region. The model was then validated on the remaining 30% of test data represented in the green shaded area. Root mean square error (RMSE) and mean square error (MSE), as well as the mean of the predicted and observed values for the test set were reported in each panel.

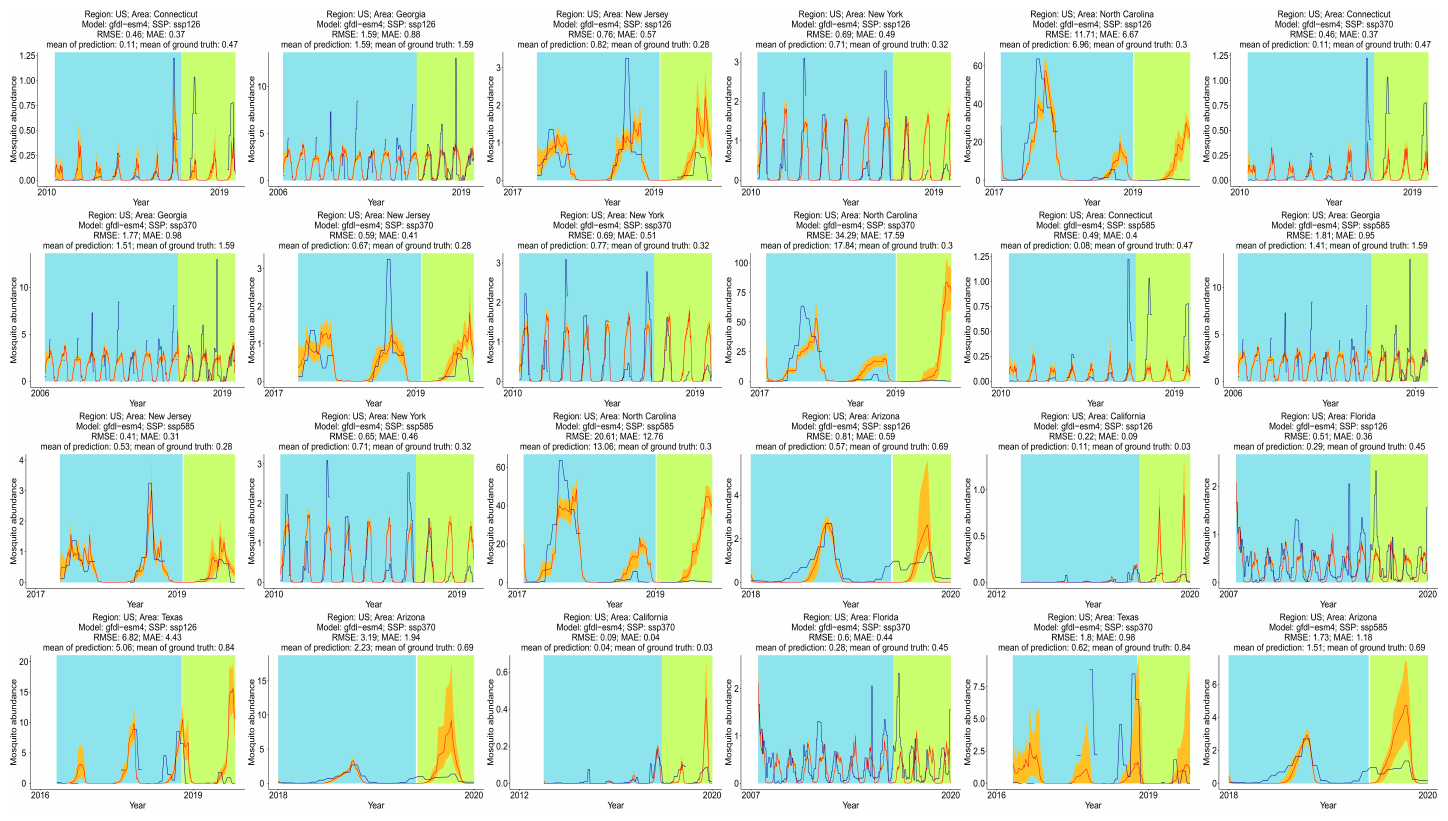

**Figure S26. Fitted and predicted values of mosquito abundance from quadratic precipitation-dependent carrying capacity model by area in the United States (1 of 2).** The observed data is shown in black, while the model projection was visualized in the red line with the yellow band representing the 95% Crl. The model was calibrated on the first 70% of observed data and is represented in the blue shaded region. The model was then validated on the remaining 30% of test data represented in the green shaded area. Root mean square error (RMSE) and mean square error (MSE), as well as the mean of the predicted and observed values for the test set were reported in each panel.

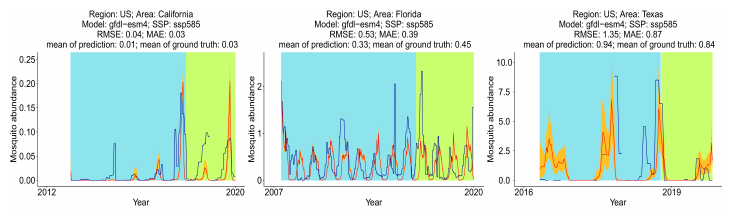

**Figure S27. Fitted and predicted values of mosquito abundance from quadratic precipitation-dependent carrying capacity model by area in the United States (2 of 2).** The observed data is shown in black, while the model projection was visualized in the red line with the yellow band representing the 95% Crl. The model was calibrated on the first 70% of observed data and is represented in the blue shaded region. The model was then validated on the remaining 30% of test data represented in the green shaded area. Root mean square error (RMSE) and mean square error (MSE), as well as the mean of the predicted and observed values for the test set were reported in each panel.

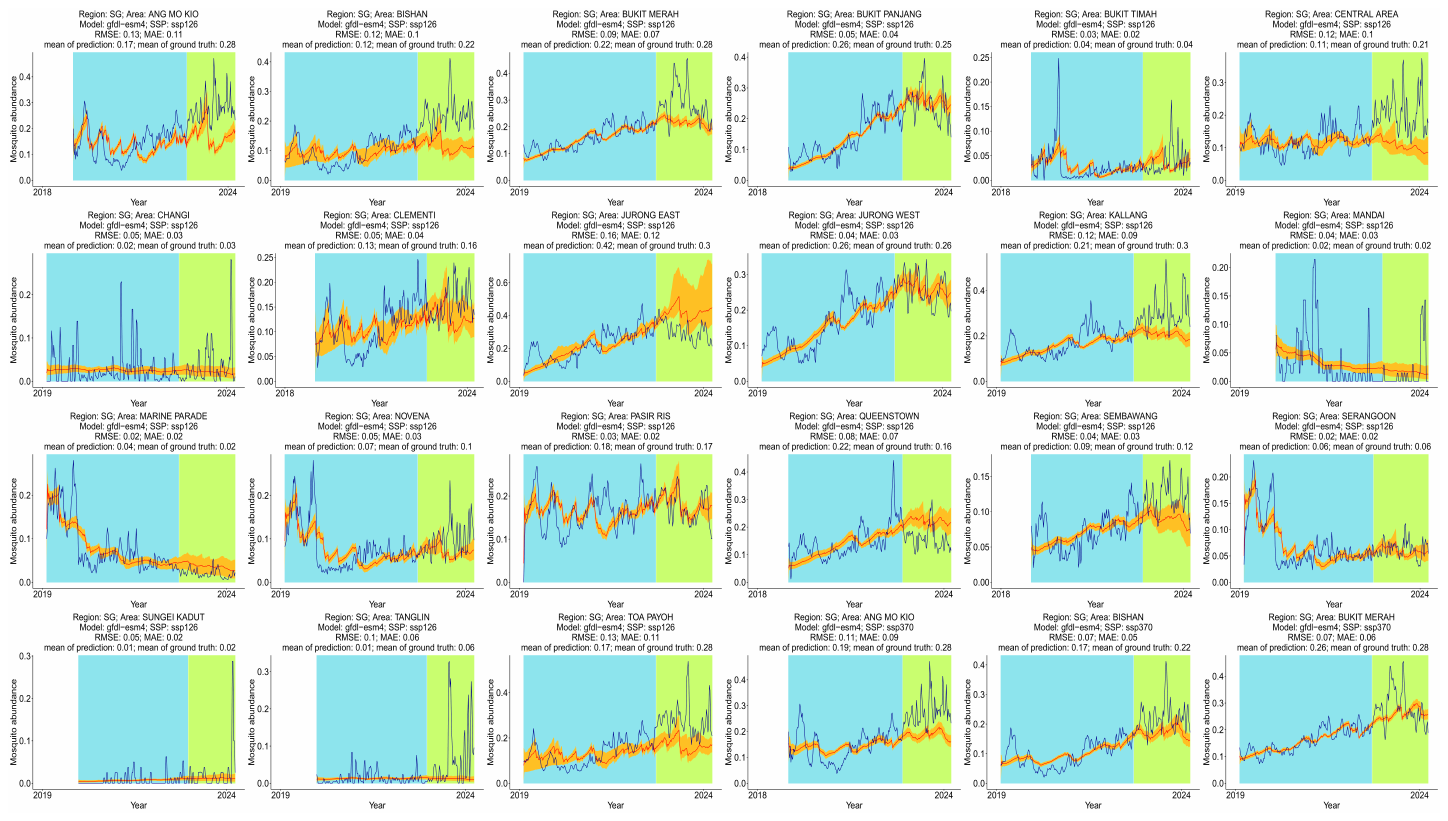

**Figure S28. Fitted and predicted values of mosquito abundance from quadratic precipitation-dependent carrying capacity model by area in Singapore (1 of 3).** The observed data is shown in black, while the model projection was visualized in the red line with the yellow band representing the 95% Crl. The model was calibrated on the first 70% of observed data and is represented in the blue shaded region. The model was then validated on the remaining 30% of test data represented in the green shaded area. Root mean square error (RMSE) and mean square error (MSE), as well as the mean of the predicted and observed values for the test set were reported in each panel.

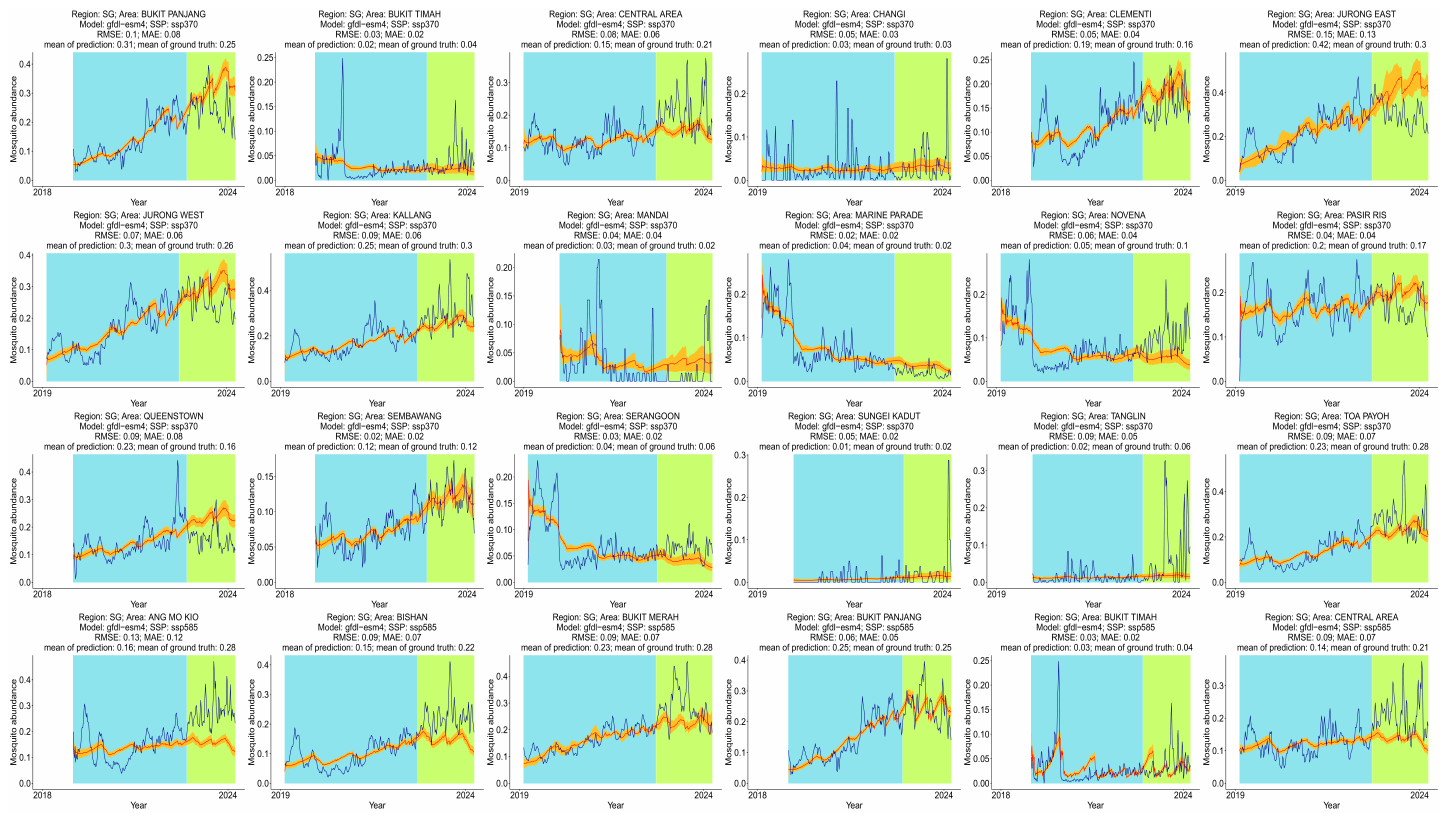

**Figure S29. Fitted and predicted values of mosquito abundance from quadratic precipitation-dependent carrying capacity model by area in Singapore (2 of 3).** The observed data is shown in black, while the model projection was visualized in the red line with the yellow band representing the 95% Crl. The model was calibrated on the first 70% of observed data and is represented in the blue shaded region. The model was then validated on the remaining 30% of test data represented in the green shaded area. Root mean square error (RMSE) and mean square error (MSE), as well as the mean of the predicted and observed values for the test set were reported in each panel.

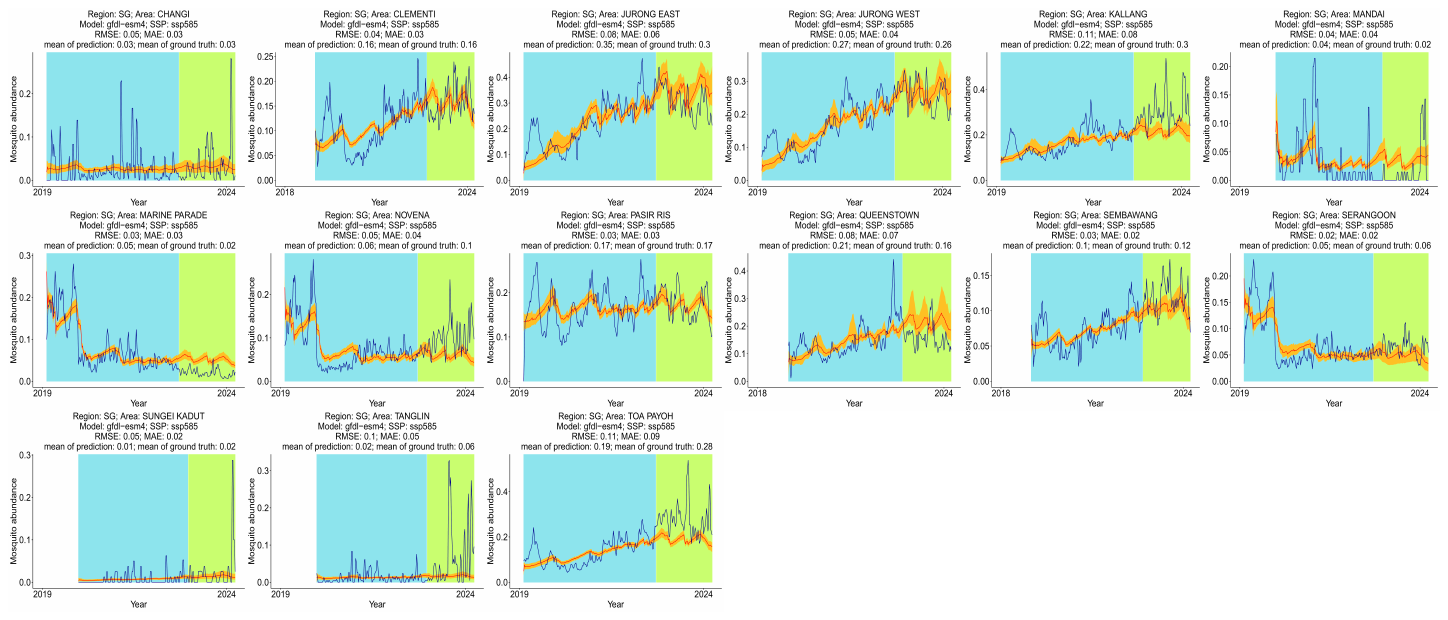

**Figure S30. Fitted and predicted values of mosquito abundance from quadratic precipitation-dependent carrying capacity model by area in Singapore (3 of 3).** The observed data is shown in black, while the model projection was visualized in the red line with the yellow band representing the 95% Crl. The model was calibrated on the first 70% of observed data and is represented in the blue shaded region. The model was then validated on the remaining 30% of test data represented in the green shaded area. Root mean square error (RMSE) and mean square error (MSE), as well as the mean of the predicted and observed values for the test set were reported in each panel.

**3. Sensitivity analysis**

Sensitivity analysis was conducted to examine the changes in mosquito abundance and intervention effectiveness when parameters were different from calibrated values. Here, we varied the climate model used to project future mosquito abundance in each region (subsection **3.1**) the inferred life-history traits (subsection **3.2**) andthe setup of the IIT release programme (subsection **3.3**) to examine the sensitivity of our model projections to the model input. Mosquito abundance in baseline and intervention scenarios were reprojected forward in time to calculate the change in abundance (CA) and intervention effectiveness (IE), and were summarised at the regional level. In each case, we observed minor differences in projected abundance, but intervention effectiveness remain robust across the studied regions.

**3.1 Varying the input climate model**

We replaced the general circulate models (GCMs) from the GFDL-ESM4 that was used in the main results with the other four (IPSL-CM6A-LR, MPI-ESM1-2-HR, MRI-ESM2-0, and UKESM1-0-LL) models. We retained each model to these new climate models and scenarios and projected the mosquito abundances in both baseline and intervention scenarios.

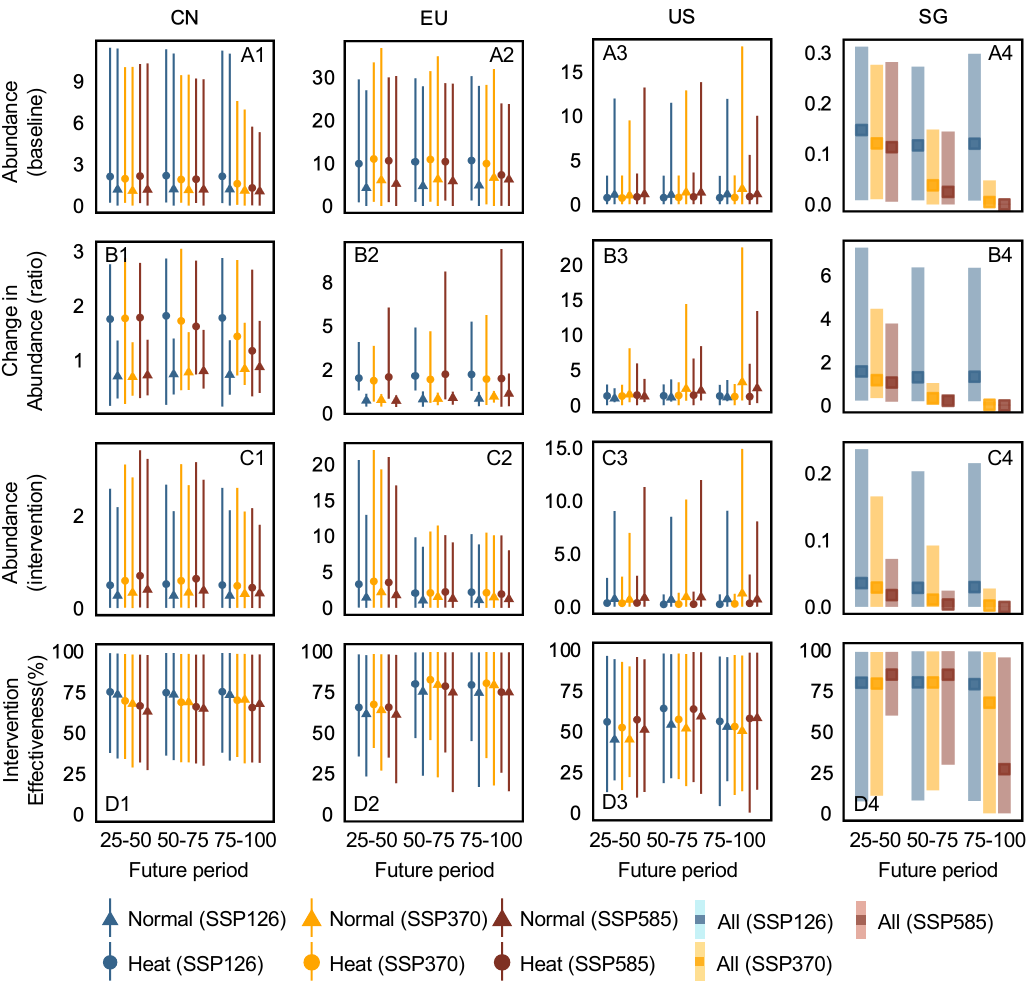

**Figure S31. Model projections with corresponding intervention effectiveness (IE) under the IPSL-CM6A-LR model.** The series of box plots summarized the projected abundance in the baseline scenario (A1-4), change in abundance (B1-4), abundance in intervention scenario (C1-4), and intervention effectiveness (D1-4) for each study region, namely, mainland China (CN), Europe (EU), the United States (US), and Singapore (SG) across three future time periods, 2025–2050, 2050–2075 and 2075–2100. Change in abundance was taken as the projected future mosquito abundance divided by historical mosquito abundance. Intervention effectiveness is defined as the **percentage reduction** in mosquito abundance achieved by implementing IIT interventions relative to the baseline scenario of no IIT interventions. Simulations assumed a 6:1 overflooding ratio. The figure visualized projections under the IPSL-CM6A-LR model. Each panel visualized seasonal changes in heat and normal conditions for EU, US and CN or the whole year for SG. Heat conditions were defined as the 12 consecutive warmest weeks of each year, while normal conditions included all other weeks. 95% credible intervals were showed as a line. The colour scheme included SSP126 in blue, SSP370 in yellow, and the SPP585 in red to compare projections across different emission scenarios.

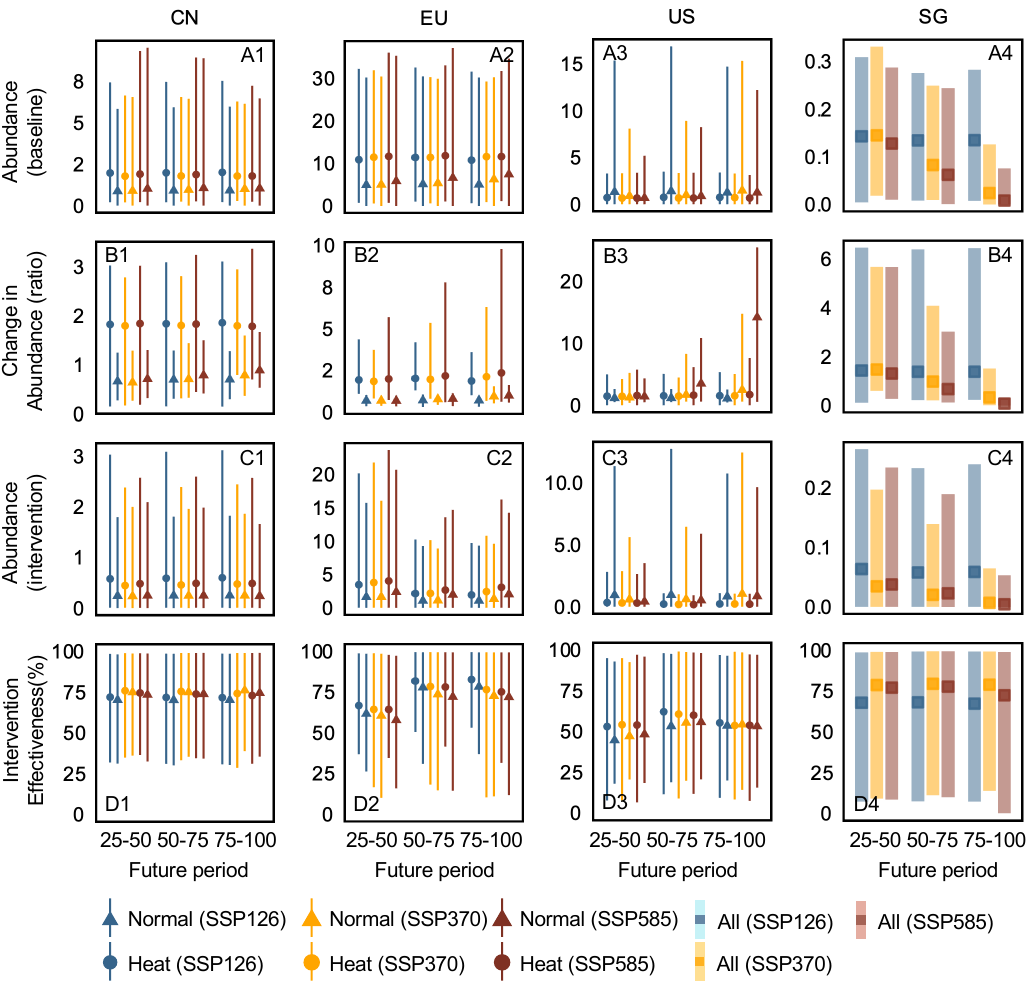

**Figure S32. Model projections with corresponding intervention effectiveness (IE) under the MPI-ESM1-2-HR model.** The series of box plots summarized the projected abundance in the baseline scenario (A1-4), change in abundance (B1-4), abundance in intervention scenario (C1-4), and intervention effectiveness (D1-4) for each study region, namely, mainland China (CN), Europe (EU), the United States (US), and Singapore (SG) across three future time periods, 2025–2050, 2050–2075 and 2075–2100. Change in abundance was taken as the projected future mosquito abundance divided by historical mosquito abundance. Intervention effectiveness is defined as the **percentage reduction** in mosquito abundance achieved by implementing IIT interventions relative to the baseline scenario of no IIT interventions. Simulations assumed a 6:1 overflooding ratio. The figure visualized projections under the MPI-ESM1-2-HR model. Each panel visualized seasonal changes in heat and normal conditions for EU, US and CN or the whole year for SG. Heat conditions were defined as the 12 consecutive warmest weeks of each year, while normal conditions included all other weeks. 95% credible intervals were showed as a line. The colour scheme included SSP126 in blue, SSP370 in yellow, and the SPP585 in red to compare projections across different emission scenarios.

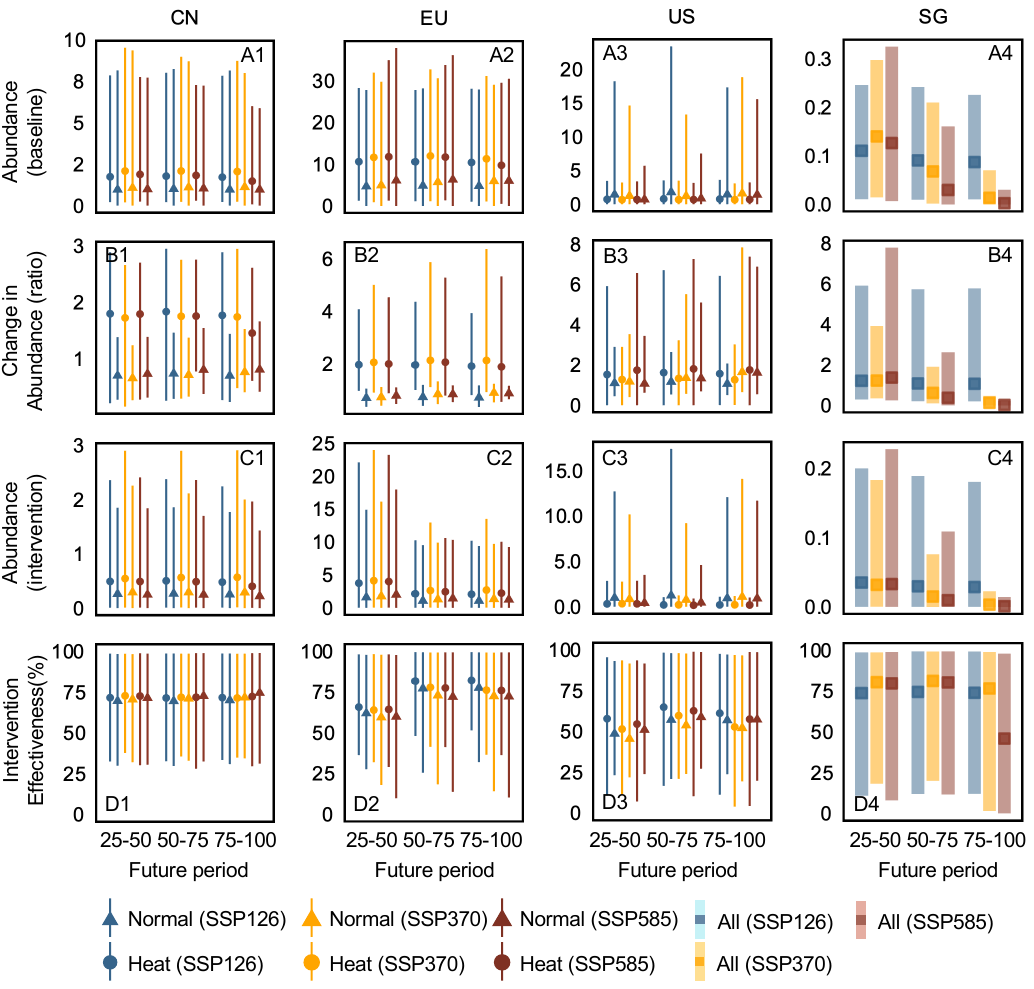

**Figure S33. Model projections with corresponding intervention effectiveness (IE) under the MRI-ESM2-0 model.** The series of box plots summarized the projected abundance in the baseline scenario (A1-4), change in abundance (B1-4), abundance in intervention scenario (C1-4), and intervention effectiveness (D1-4) for each study region, namely, mainland China (CN), Europe (EU), the United States (US), and Singapore (SG) across three future time periods, 2025–2050, 2050–2075 and 2075–2100. Change in abundance was taken as the projected future mosquito abundance divided by historical mosquito abundance. Intervention effectiveness is defined as the **percentage reduction** in mosquito abundance achieved by implementing IIT interventions relative to the baseline scenario of no IIT interventions. Simulations assumed a 6:1 overflooding ratio. The figure visualized projections under the MRI-ESM2-0 model. Each panel visualized seasonal changes in heat and normal conditions for EU, US and CN or the whole year for SG. Heat conditions were defined as the 12 consecutive warmest weeks of each year, while normal conditions included all other weeks. 95% credible intervals were showed as a line. The colour scheme included SSP126 in blue, SSP370 in yellow, and the SPP585 in red to compare projections across different emission scenarios.

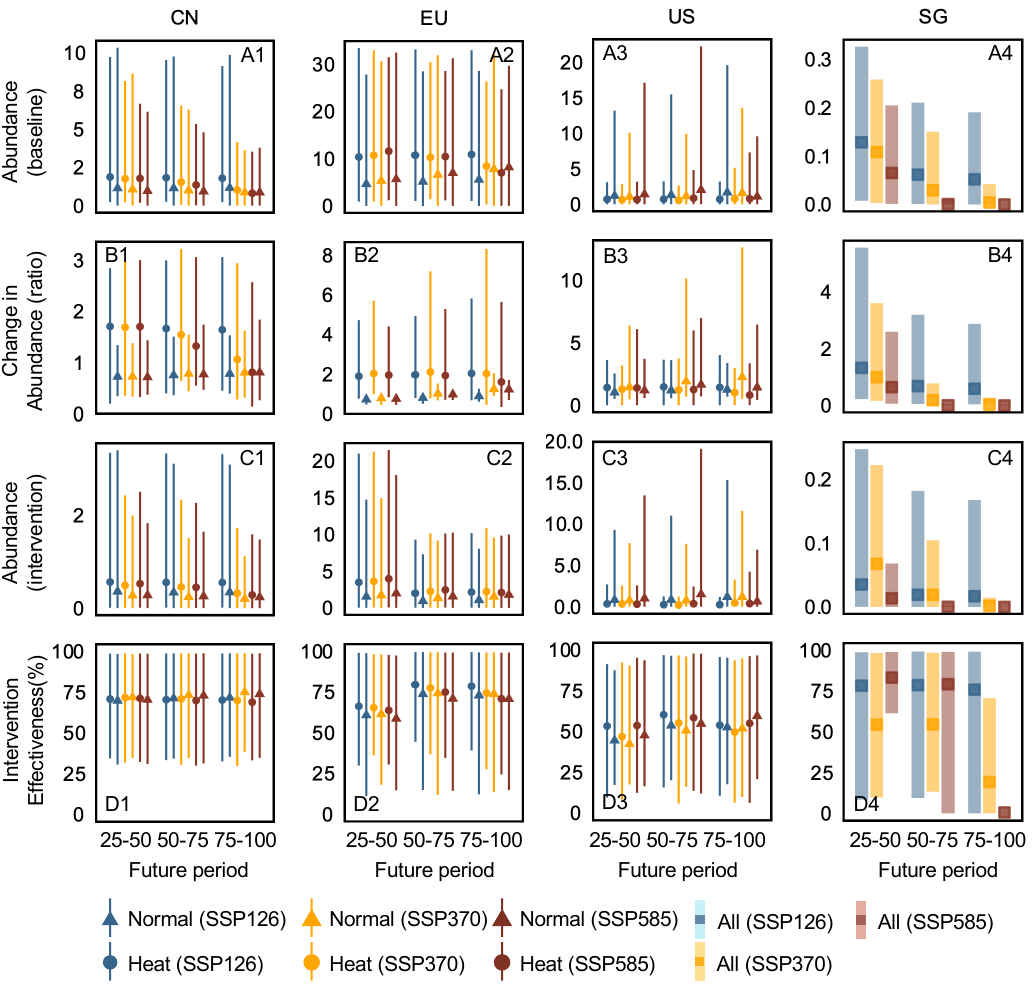

**Figure S34. Model projections with corresponding intervention effectiveness (IE) under the UKESM1-0-LL model.** The series of box plots summarized the projected abundance in the baseline scenario (A1-4), change in abundance (B1-4), abundance in intervention scenario (C1-4), and intervention effectiveness (D1-4) for each study region, namely, mainland China (CN), Europe (EU), the United States (US), and Singapore (SG) across three future time periods, 2025–2050, 2050–2075 and 2075–2100. Change in abundance was taken as the projected future mosquito abundance divided by historical mosquito abundance. Intervention effectiveness is defined as the **percentage reduction** in mosquito abundance achieved by implementing IIT interventions relative to the baseline scenario of no IIT interventions. Simulations assumed a 6:1 overflooding ratio. The figure visualized projections under the UKESM1-0-LL model. Each panel visualized seasonal changes in heat and normal conditions for EU, US and CN or the whole year for SG. Heat conditions were defined as the 12 consecutive warmest weeks of each year, while normal conditions included all other weeks. 95% credible intervals were showed as a line. The colour scheme included SSP126 in blue, SSP370 in yellow, and the SPP585 in red to compare projections across different emission scenarios.

**3.2 Sensitivity analysis on varying inferred life-history traits**

We separately varied the climate dependent entomological functions ($f_{\mu,a}$, $f_{\mu,f}$, $f_{\mu,m}, f_{\tau}$, $f_{\rho}$, $f_{K}$) and immigration rate ($\alpha$) by increasing or decreasing the estimated values by 5% while keeping all other parameters unchanged under GFDL-ESM4 climate model.

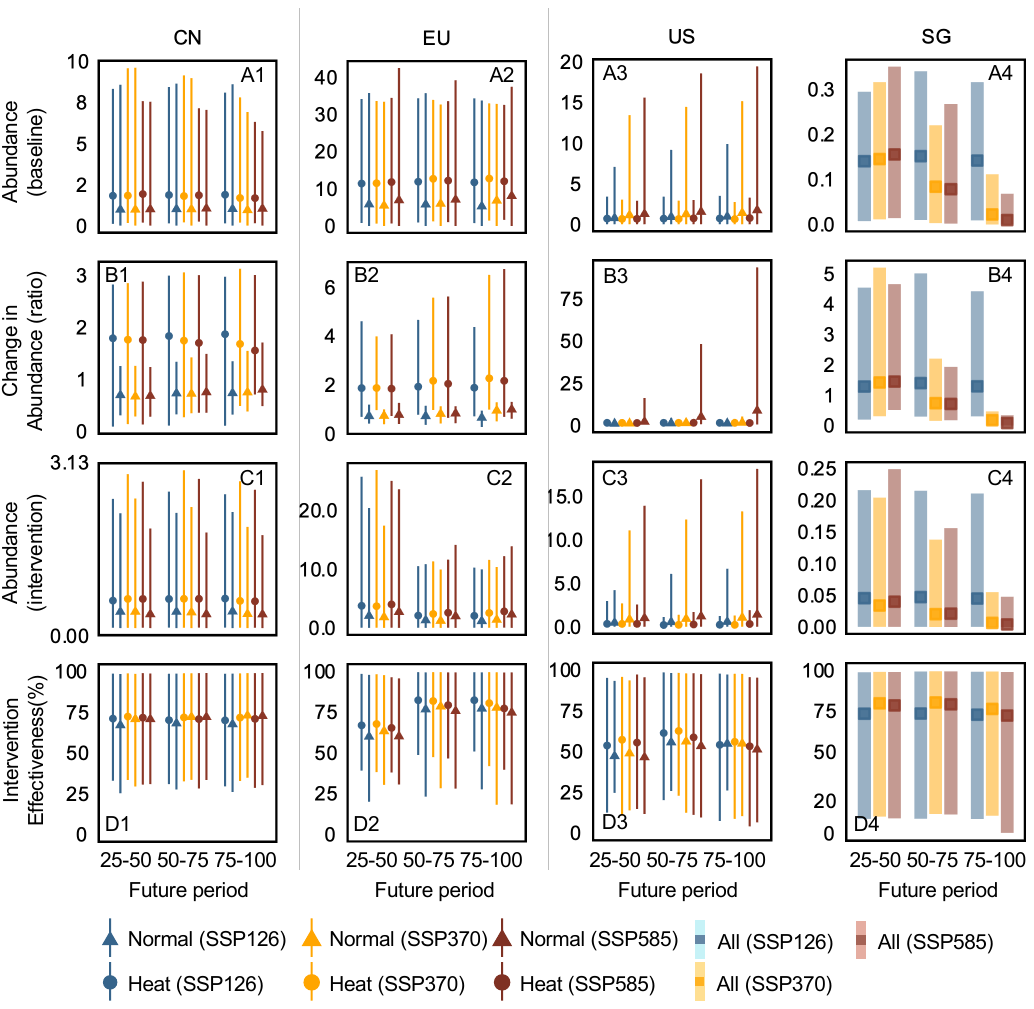

**Figure S35. Model projections with corresponding intervention effectiveness (IE) with a 5% increase in the aquatic mortality rate.** The series of box plots summarized the projected abundance in the baseline scenario (A1-4), change in abundance (B1-4), abundance in intervention scenario (C1-4), and intervention effectiveness (D1-4) for each study region, namely, mainland China (CN), Europe (EU), the United States (US), and Singapore (SG) across three future time periods, 2025–2050, 2050–2075 and 2075–2100. Change in abundance was taken as the projected future mosquito abundance divided by historical mosquito abundance. Intervention effectiveness is defined as the **percentage reduction** in mosquito abundance achieved by implementing IIT interventions relative to the baseline scenario of no IIT interventions. Simulations assumed a 6:1 overflooding ratio. The figure visualized projections under the GFDL-ESM4 model. Each panel visualized seasonal changes in heat and normal conditions for EU, US and CN or the whole year for SG. Heat conditions were defined as the 12 consecutive warmest weeks of each year, while normal conditions included all other weeks. 95% credible intervals were showed as a line. The colour scheme included SSP126 in blue, SSP370 in yellow, and the SPP585 in red to compare projections across different emission scenarios.

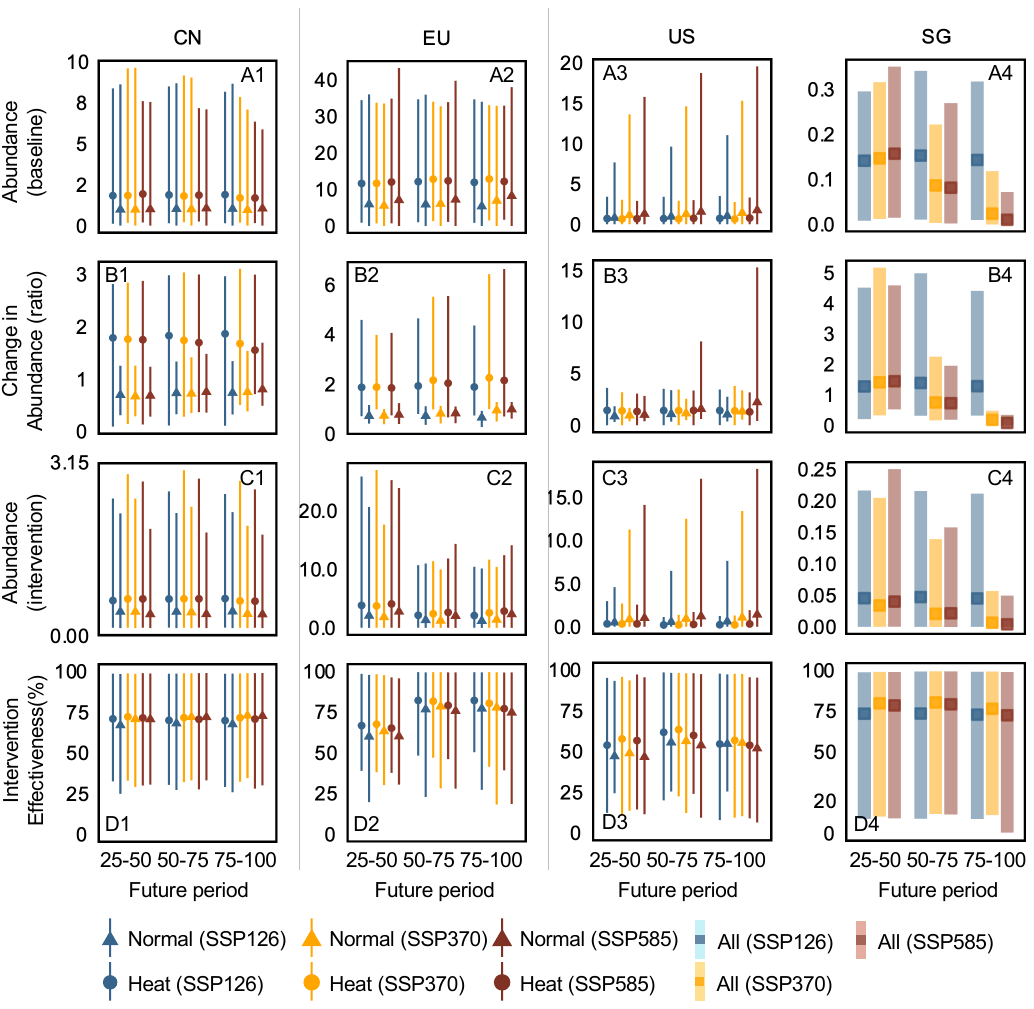

**Figure S36. Model projections with corresponding intervention effectiveness (IE) with a 5% decrease in the aquatic mortality rate.** The series of box plots summarized the projected abundance in the baseline scenario (A1-4), change in abundance (B1-4), abundance in intervention scenario (C1-4), and intervention effectiveness (D1-4) for each study region, namely, mainland China (CN), Europe (EU), the United States (US), and Singapore (SG) across three future time periods, 2025–2050, 2050–2075 and 2075–2100. Change in abundance was taken as the projected future mosquito abundance divided by historical mosquito abundance. Intervention effectiveness is defined as the **percentage reduction** in mosquito abundance achieved by implementing IIT interventions relative to the baseline scenario of no IIT interventions. Simulations assumed a 6:1 overflooding ratio. The figure visualized projections under the GFDL-ESM4 model. Each panel visualized seasonal changes in heat and normal conditions for EU, US and CN or the whole year for SG. Heat conditions were defined as the 12 consecutive warmest weeks of each year, while normal conditions included all other weeks. 95% credible intervals were showed as a line. The colour scheme included SSP126 in blue, SSP370 in yellow, and the SPP585 in red to compare projections across different emission scenarios.

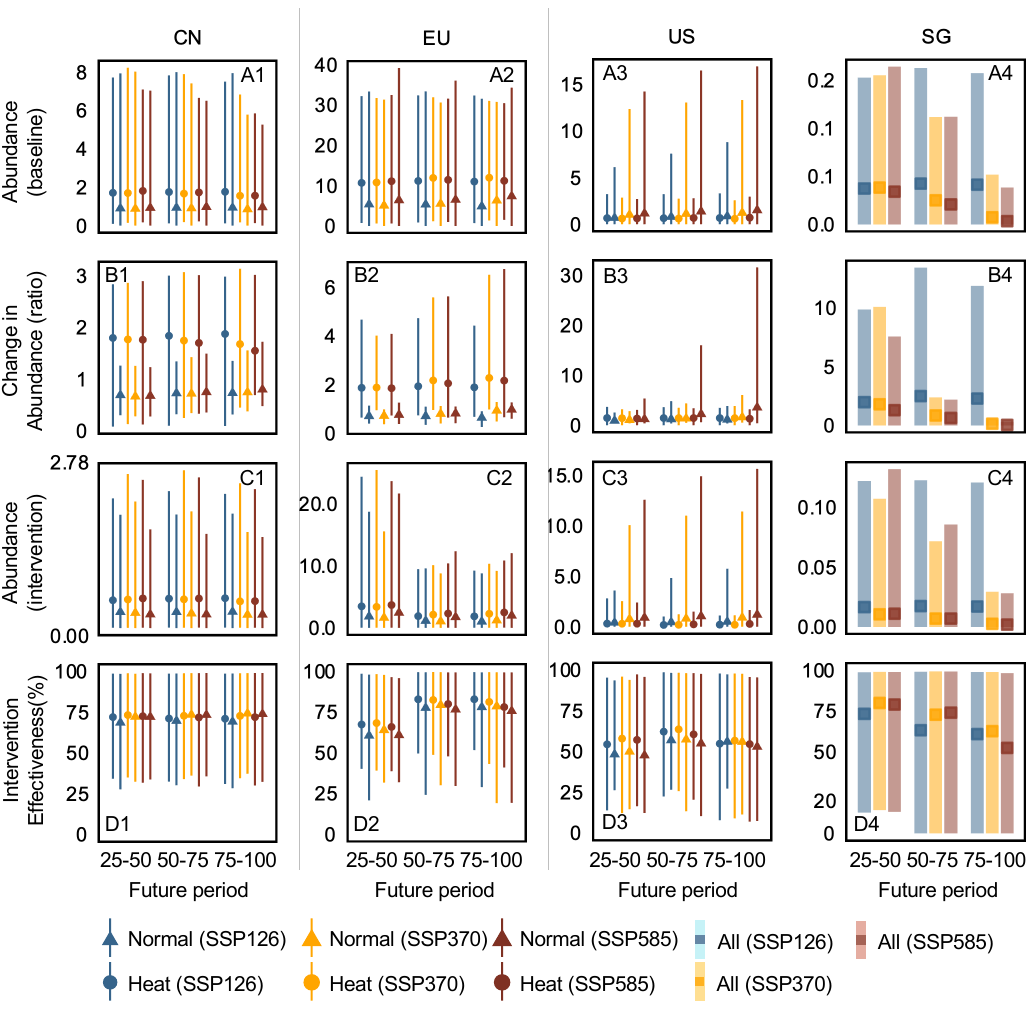

**Figure S37. Model projections with corresponding intervention effectiveness (IE) with a 5% increase in the adult mortality rate.** The series of box plots summarized the projected abundance in the baseline scenario (A1-4), change in abundance (B1-4), abundance in intervention scenario (C1-4), and intervention effectiveness (D1-4) for each study region, namely, mainland China (CN), Europe (EU), the United States (US), and Singapore (SG) across three future time periods, 2025–2050, 2050–2075 and 2075–2100. Change in abundance was taken as the projected future mosquito abundance divided by historical mosquito abundance. Intervention effectiveness is defined as the **percentage reduction** in mosquito abundance achieved by implementing IIT interventions relative to the baseline scenario of no IIT interventions. Simulations assumed a 6:1 overflooding ratio. The figure visualized projections under the GFDL-ESM4 model. Each panel visualized seasonal changes in heat and normal conditions for EU, US and CN or the whole year for SG. Heat conditions were defined as the 12 consecutive warmest weeks of each year, while normal conditions included all other weeks. 95% credible intervals were showed as a line. The colour scheme included SSP126 in blue, SSP370 in yellow, and the SPP585 in red to compare projections across different emission scenarios.

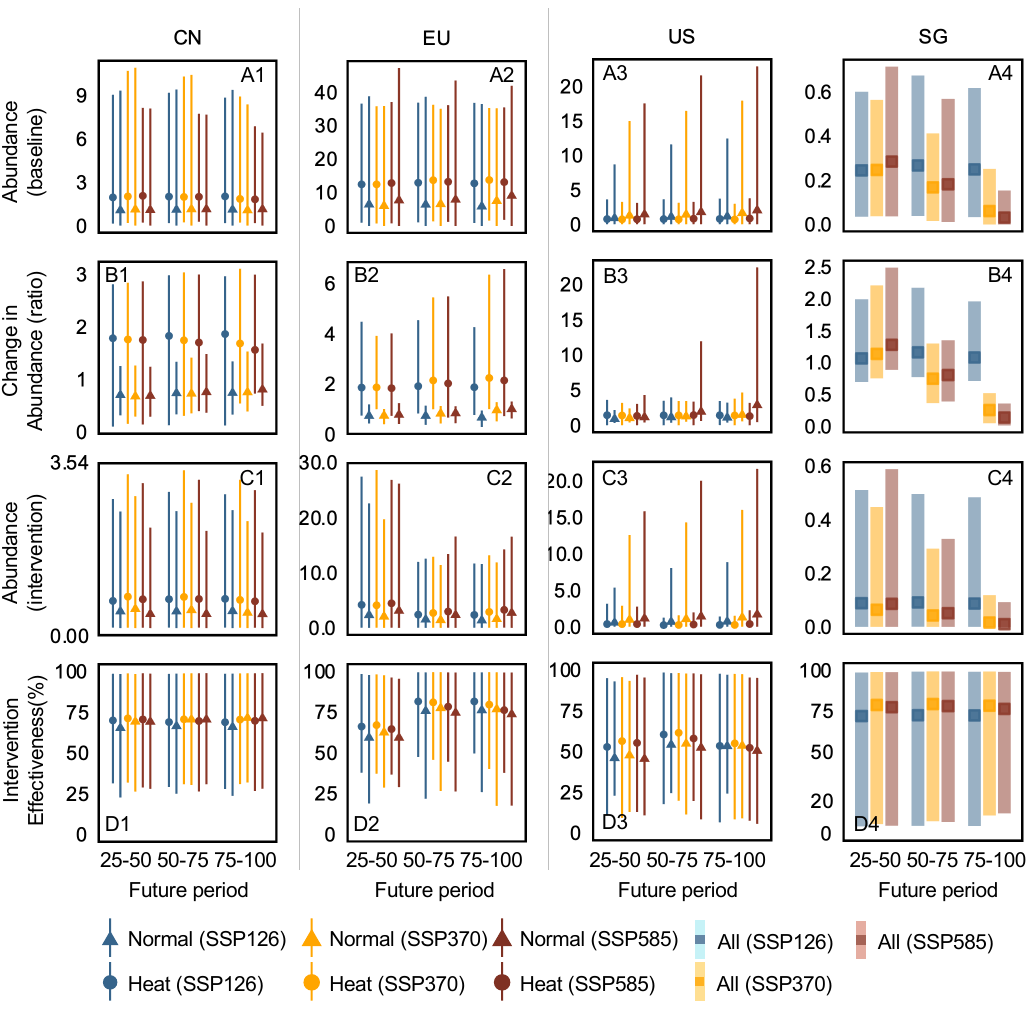

**Figure S38. Model projections with corresponding intervention effectiveness (IE) with a 5% decrease in the adult mortality rate.** The series of box plots summarized the projected abundance in the baseline scenario (A1-4), change in abundance (B1-4), abundance in intervention scenario (C1-4), and intervention effectiveness (D1-4) for each study region, namely, mainland China (CN), Europe (EU), the United States (US), and Singapore (SG) across three future time periods, 2025–2050, 2050–2075 and 2075–2100. Change in abundance was taken as the projected future mosquito abundance divided by historical mosquito abundance. Intervention effectiveness is defined as the **percentage reduction** in mosquito abundance achieved by implementing IIT interventions relative to the baseline scenario of no IIT interventions. Simulations assumed a 6:1 overflooding ratio. The figure visualized projections under the GFDL-ESM4 model. Each panel visualized seasonal changes in heat and normal conditions for EU, US and CN or the whole year for SG. Heat conditions were defined as the 12 consecutive warmest weeks of each year, while normal conditions included all other weeks. 95% credible intervals were showed as a line. The colour scheme included SSP126 in blue, SSP370 in yellow, and the SPP585 in red to compare projections across different emission scenarios.

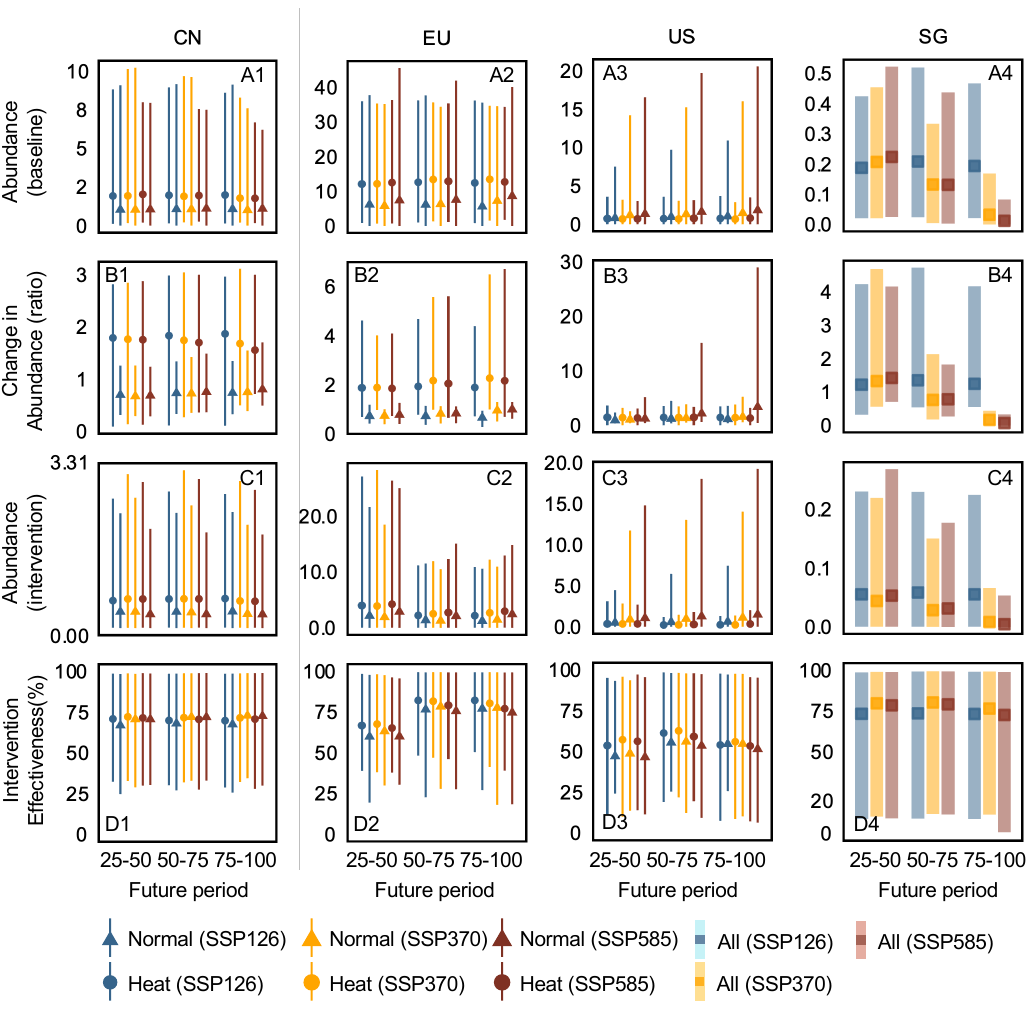

**Figure S39. Model projections with corresponding intervention effectiveness (IE) with a 5% increase in the development rate.** The series of box plots summarized the projected abundance in the baseline scenario (A1-4), change in abundance (B1-4), abundance in intervention scenario (C1-4), and intervention effectiveness (D1-4) for each study region, namely, mainland China (CN), Europe (EU), the United States (US), and Singapore (SG) across three future time periods, 2025–2050, 2050–2075 and 2075–2100. Change in abundance was taken as the projected future mosquito abundance divided by historical mosquito abundance. Intervention effectiveness is defined as the **percentage reduction** in mosquito abundance achieved by implementing IIT interventions relative to the baseline scenario of no IIT interventions. Simulations assumed a 6:1 overflooding ratio. The figure visualized projections under the GFDL-ESM4 model. Each panel visualized seasonal changes in heat and normal conditions for EU, US and CN or the whole year for SG. Heat conditions were defined as the 12 consecutive warmest weeks of each year, while normal conditions included all other weeks. 95% credible intervals were showed as a line. The colour scheme included SSP126 in blue, SSP370 in yellow, and the SPP585 in red to compare projections across different emission scenarios.

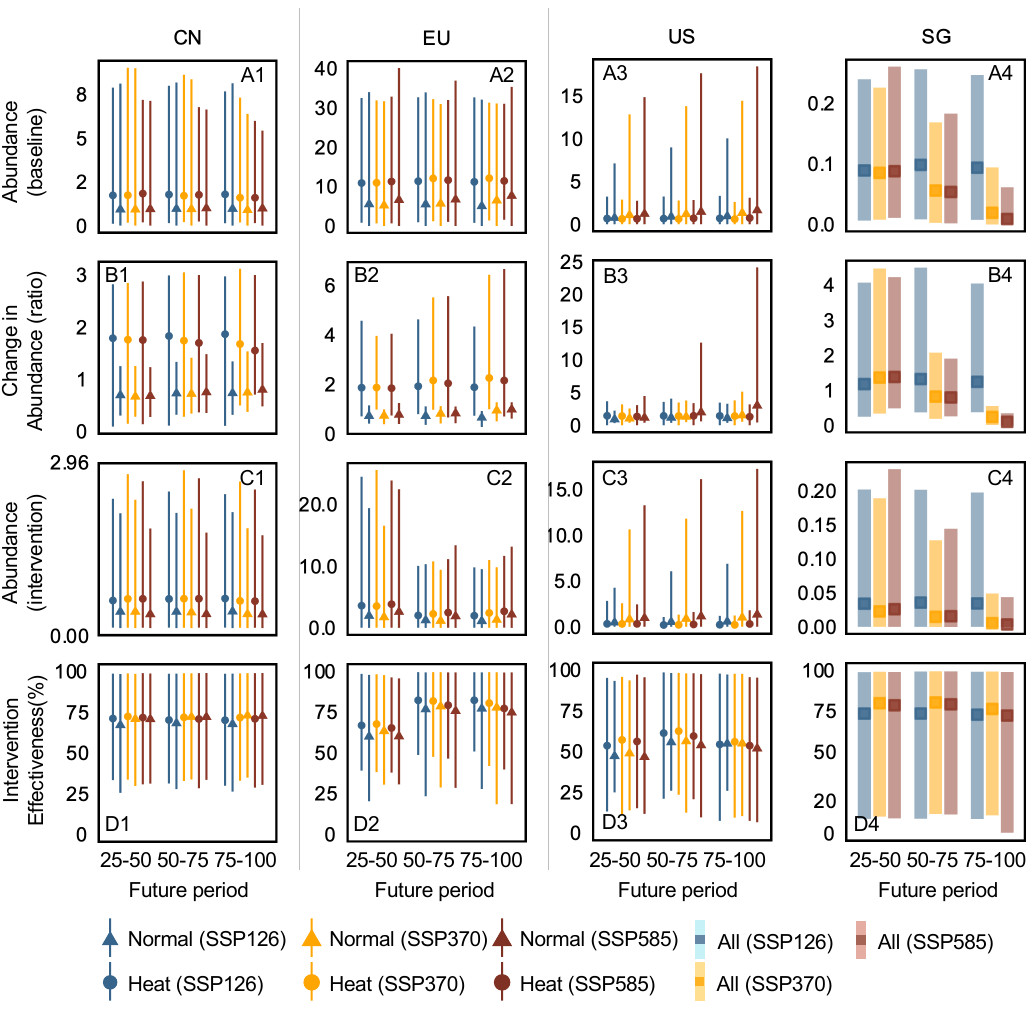

**Figure S40. Model projections with corresponding intervention effectiveness (IE) with a 5% decrease in the development rate.** The series of box plots summarized the projected abundance in the baseline scenario (A1-4), change in abundance (B1-4), abundance in intervention scenario (C1-4), and intervention effectiveness (D1-4) for each study region, namely, mainland China (CN), Europe (EU), the United States (US), and Singapore (SG) across three future time periods, 2025–2050, 2050–2075 and 2075–2100. Change in abundance was taken as the projected future mosquito abundance divided by historical mosquito abundance. Intervention effectiveness is defined as the **percentage reduction** in mosquito abundance achieved by implementing IIT interventions relative to the baseline scenario of no IIT interventions. Simulations assumed a 6:1 overflooding ratio. The figure visualized projections under the GFDL-ESM4 model. Each panel visualized seasonal changes in heat and normal conditions for EU, US and CN or the whole year for SG. Heat conditions were defined as the 12 consecutive warmest weeks of each year, while normal conditions included all other weeks. 95% credible intervals were showed as a line. The colour scheme included SSP126 in blue, SSP370 in yellow, and the SPP585 in red to compare projections across different emission scenarios.

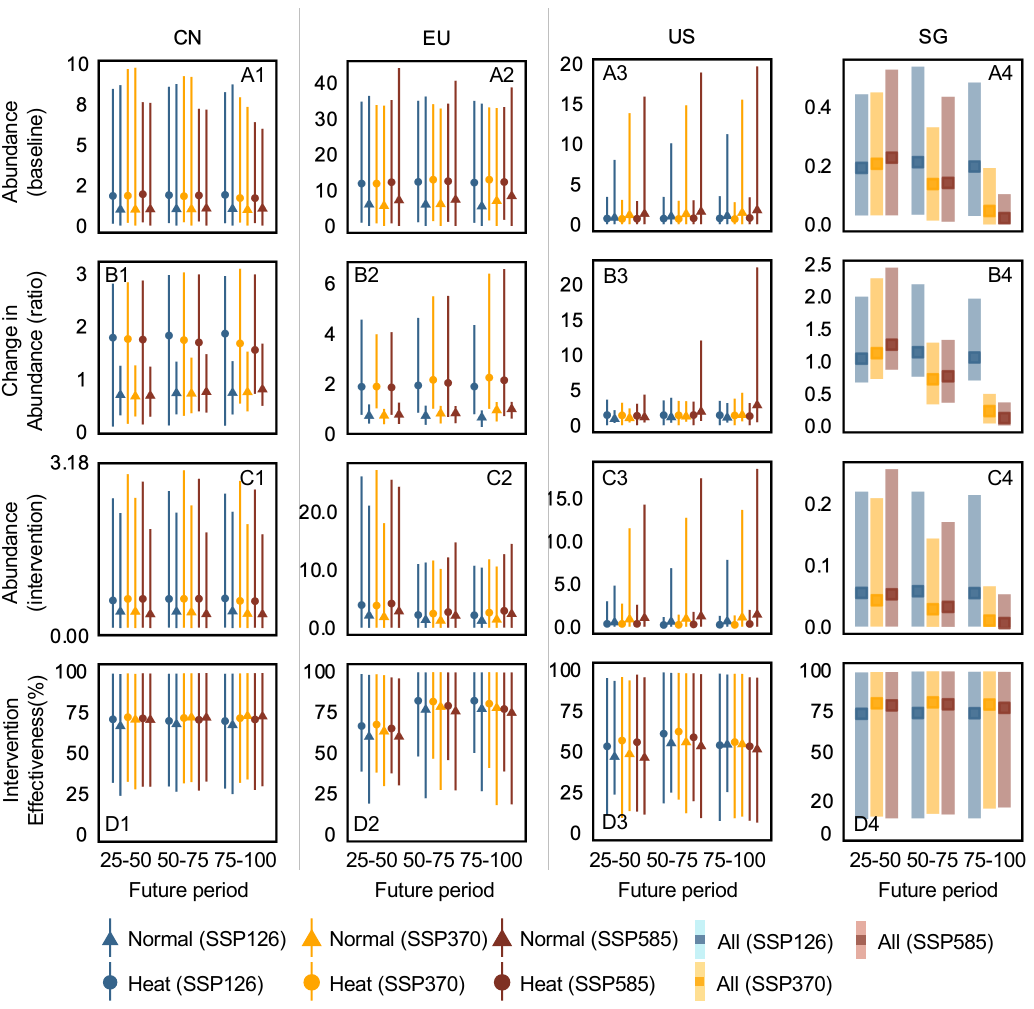

**Figure S41. Model projections with corresponding intervention effectiveness (IE) with a 5% increase in the reproduction rate.** The series of box plots summarized the projected abundance in the baseline scenario (A1-4), change in abundance (B1-4), abundance in intervention scenario (C1-4), and intervention effectiveness (D1-4) for each study region, namely, mainland China (CN), Europe (EU), the United States (US), and Singapore (SG) across three future time periods, 2025–2050, 2050–2075 and 2075–2100. Change in abundance was taken as the projected future mosquito abundance divided by historical mosquito abundance. Intervention effectiveness is defined as the **percentage reduction** in mosquito abundance achieved by implementing IIT interventions relative to the baseline scenario of no IIT interventions. Simulations assumed a 6:1 overflooding ratio. The figure visualized projections under the GFDL-ESM4 model. Each panel visualized seasonal changes in heat and normal conditions for EU, US and CN or the whole year for SG. Heat conditions were defined as the 12 consecutive warmest weeks of each year, while normal conditions included all other weeks. 95% credible intervals were showed as a line. The colour scheme included SSP126 in blue, SSP370 in yellow, and the SPP585 in red to compare projections across different emission scenarios.

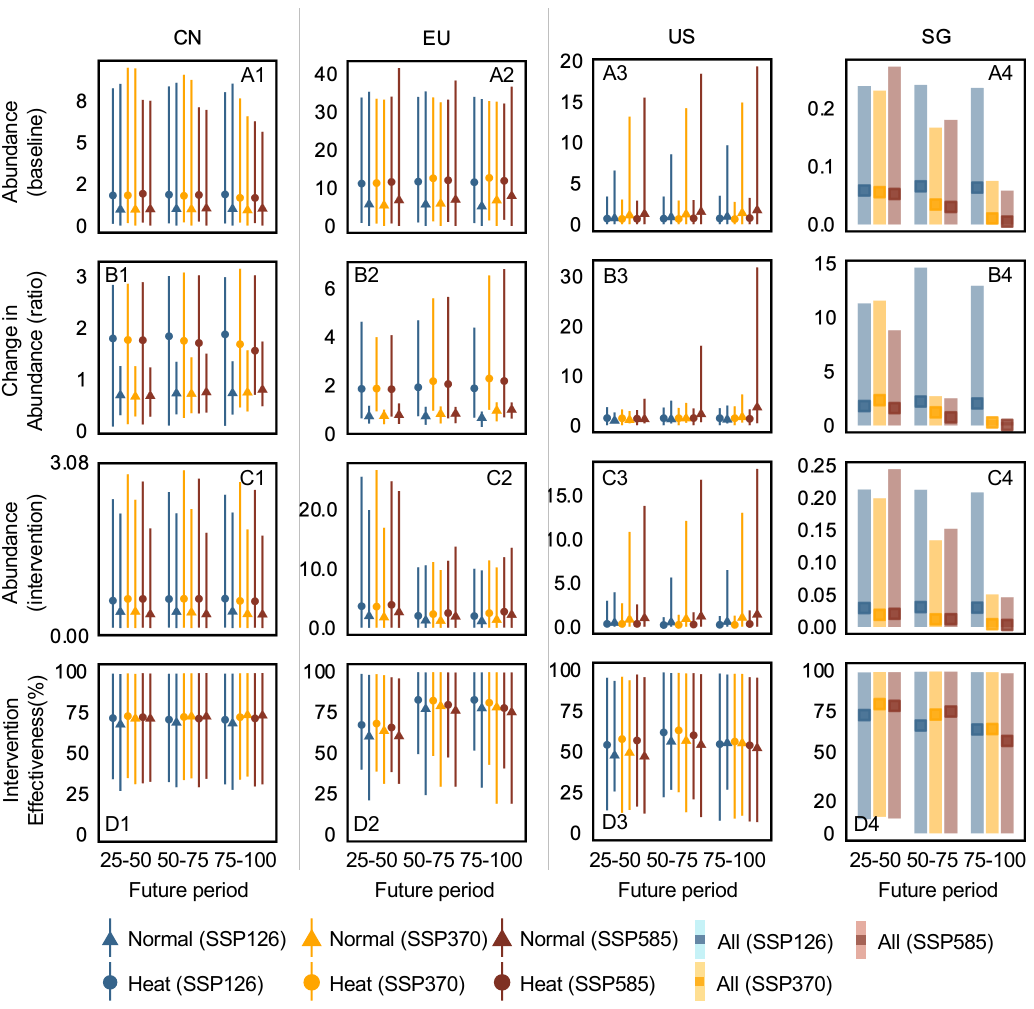

**Figure S42. Model projections with corresponding intervention effectiveness (IE) with a 5% decrease in the reproduction rate.** The series of box plots summarized the projected abundance in the baseline scenario (A1-4), change in abundance (B1-4), abundance in intervention scenario (C1-4), and intervention effectiveness (D1-4) for each study region, namely, mainland China (CN), Europe (EU), the United States (US), and Singapore (SG) across three future time periods, 2025–2050, 2050–2075 and 2075–2100. Change in abundance was taken as the projected future mosquito abundance divided by historical mosquito abundance. Intervention effectiveness is defined as the **percentage reduction** in mosquito abundance achieved by implementing IIT interventions relative to the baseline scenario of no IIT interventions. Simulations assumed a 6:1 overflooding ratio. The figure visualized projections under the GFDL-ESM4 model. Each panel visualized seasonal changes in heat and normal conditions for EU, US and CN or the whole year for SG. Heat conditions were defined as the 12 consecutive warmest weeks of each year, while normal conditions included all other weeks. 95% credible intervals were showed as a line. The colour scheme included SSP126 in blue, SSP370 in yellow, and the SPP585 in red to compare projections across different emission scenarios.

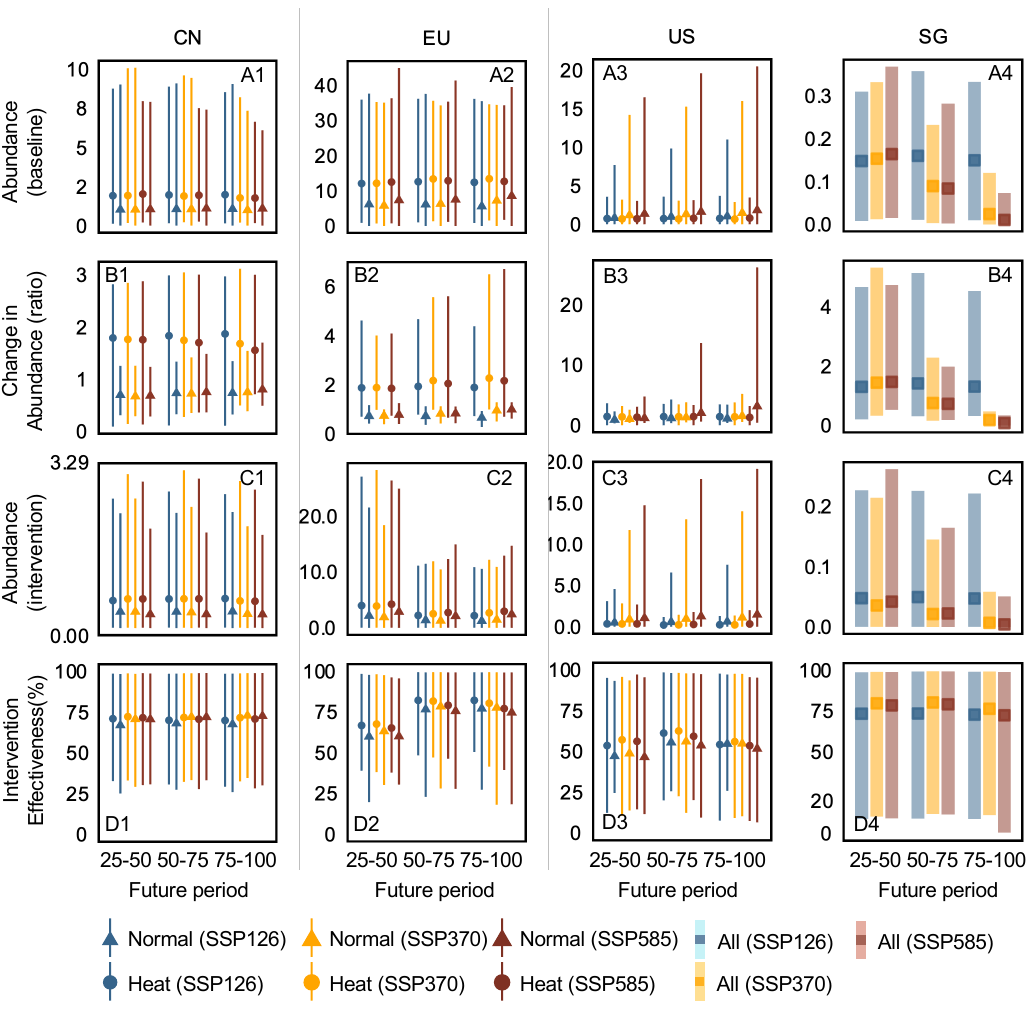

**Figure S43. Model projections with corresponding intervention effectiveness (IE) with a 5% increase in the environmental capacity.** The series of box plots summarized the projected abundance in the baseline scenario (A1-4), change in abundance (B1-4), abundance in intervention scenario (C1-4), and intervention effectiveness (D1-4) for each study region, namely, mainland China (CN), Europe (EU), the United States (US), and Singapore (SG) across three future time periods, 2025–2050, 2050–2075 and 2075–2100. Change in abundance was taken as the projected future mosquito abundance divided by historical mosquito abundance. Intervention effectiveness is defined as the **percentage reduction** in mosquito abundance achieved by implementing IIT interventions relative to the baseline scenario of no IIT interventions. Simulations assumed a 6:1 overflooding ratio. The figure visualized projections under the GFDL-ESM4 model. Each panel visualized seasonal changes in heat and normal conditions for EU, US and CN or the whole year for SG. Heat conditions were defined as the 12 consecutive warmest weeks of each year, while normal conditions included all other weeks. 95% credible intervals were showed as a line. The colour scheme included SSP126 in blue, SSP370 in yellow, and the SPP585 in red to compare projections across different emission scenarios.

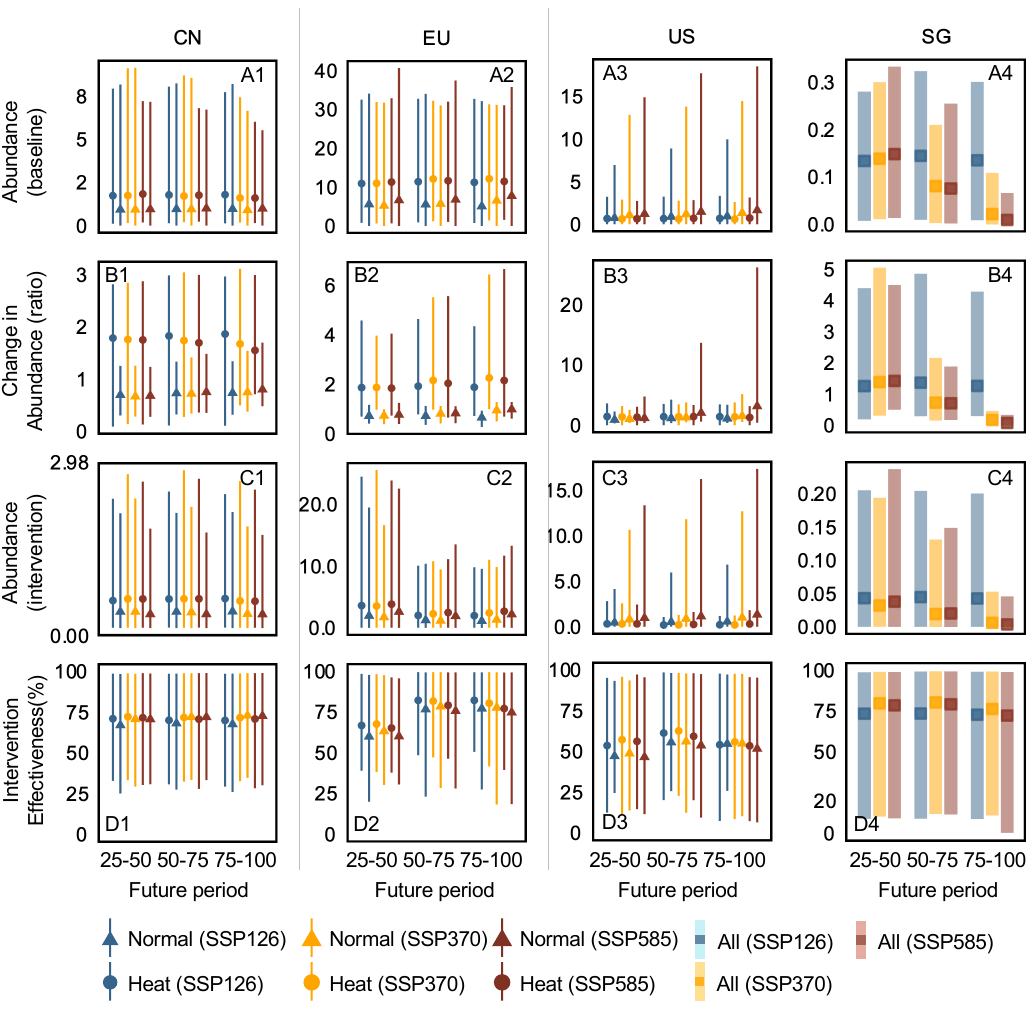

**Figure S44. Model projections with corresponding intervention effectiveness (IE) with a 5% decrease in the environmental capacity.** The series of box plots summarized the projected abundance in the baseline scenario (A1-4), change in abundance (B1-4), abundance in intervention scenario (C1-4), and intervention effectiveness (D1-4) for each study region, namely, mainland China (CN), Europe (EU), the United States (US), and Singapore (SG) across three future time periods, 2025–2050, 2050–2075 and 2075–2100. Change in abundance was taken as the projected future mosquito abundance divided by historical mosquito abundance. Intervention effectiveness is defined as the **percentage reduction** in mosquito abundance achieved by implementing IIT interventions relative to the baseline scenario of no IIT interventions. Simulations assumed a 6:1 overflooding ratio. The figure visualized projections under the GFDL-ESM4 model. Each panel visualized seasonal changes in heat and normal conditions for EU, US and CN or the whole year for SG. Heat conditions were defined as the 12 consecutive warmest weeks of each year, while normal conditions included all other weeks. 95% credible intervals were showed as a line. The colour scheme included SSP126 in blue, SSP370 in yellow, and the SPP585 in red to compare projections across different emission scenarios.

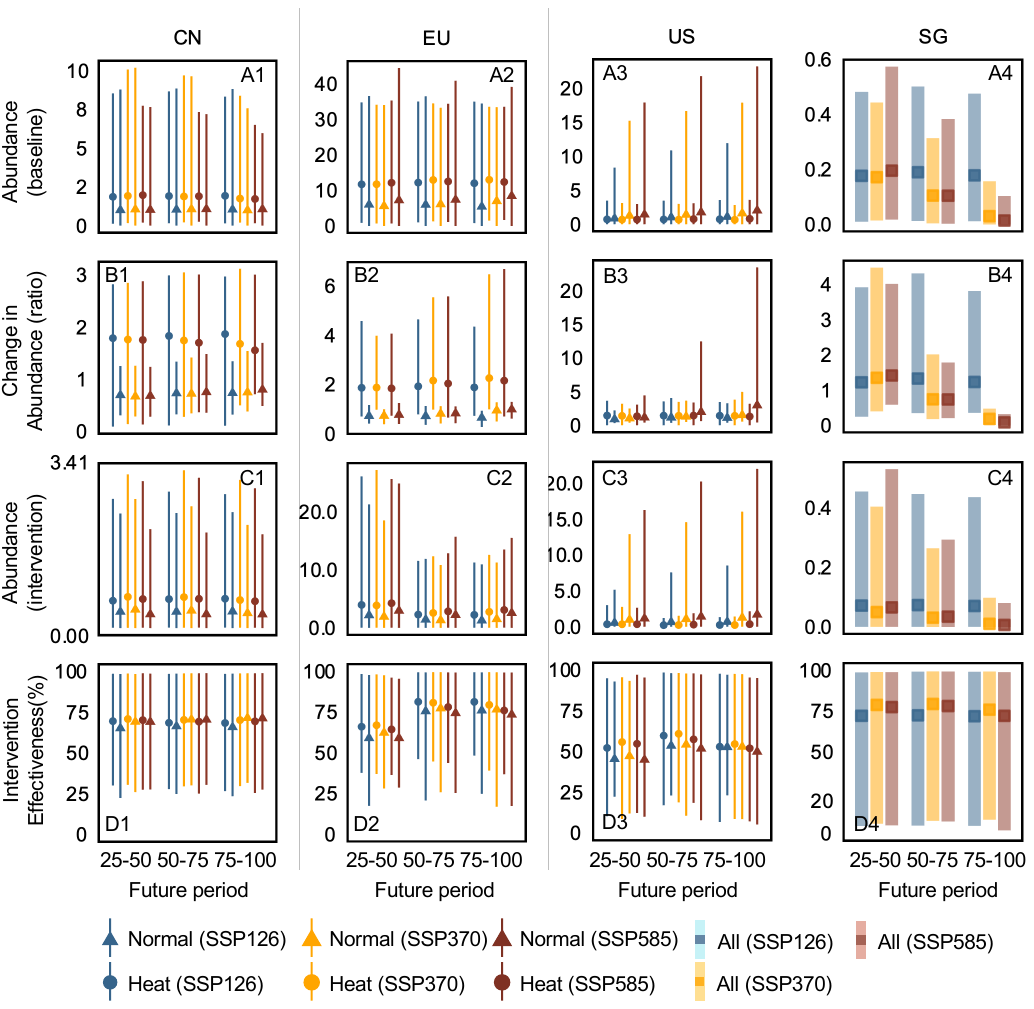

**Figure S45. Model projections with corresponding intervention effectiveness (IE) with a 5% increase in the migration rate.** The series of box plots summarized the projected abundance in the baseline scenario (A1-4), change in abundance (B1-4), abundance in intervention scenario (C1-4), and intervention effectiveness (D1-4) for each study region, namely, mainland China (CN), Europe (EU), the United States (US), and Singapore (SG) across three future time periods, 2025–2050, 2050–2075 and 2075–2100. Change in abundance was taken as the projected future mosquito abundance divided by historical mosquito abundance. Intervention effectiveness is defined as the **percentage reduction** in mosquito abundance achieved by implementing IIT interventions relative to the baseline scenario of no IIT interventions. Simulations assumed a 6:1 overflooding ratio. The figure visualized projections under the GFDL-ESM4 model. Each panel visualized seasonal changes in heat and normal conditions for EU, US and CN or the whole year for SG. Heat conditions were defined as the 12 consecutive warmest weeks of each year, while normal conditions included all other weeks. 95% credible intervals were showed as a line. The colour scheme included SSP126 in blue, SSP370 in yellow, and the SPP585 in red to compare projections across different emission scenarios.

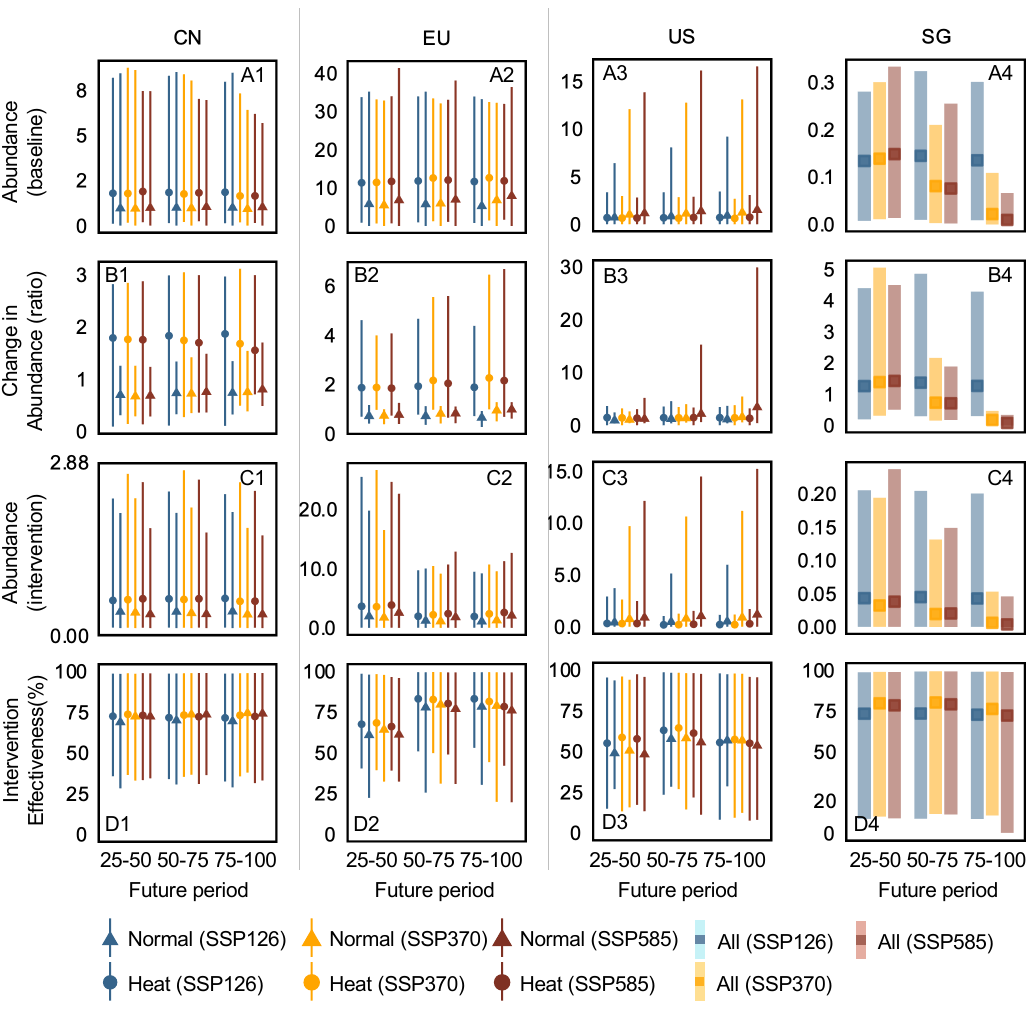

**Figure S46. Model projections with corresponding intervention effectiveness (IE) with a 5% decrease in the migration rate.** The series of box plots summarized the projected abundance in the baseline scenario (A1-4), change in abundance (B1-4), abundance in intervention scenario (C1-4), and intervention effectiveness (D1-4) for each study region, namely, mainland China (CN), Europe (EU), the United States (US), and Singapore (SG) across three future time periods, 2025–2050, 2050–2075 and 2075–2100. Change in abundance was taken as the projected future mosquito abundance divided by historical mosquito abundance. Intervention effectiveness is defined as the **percentage reduction** in mosquito abundance achieved by implementing IIT interventions relative to the baseline scenario of no IIT interventions. Simulations assumed a 6:1 overflooding ratio. The figure visualized projections under the GFDL-ESM4 model. Each panel visualized seasonal changes in heat and normal conditions for EU, US and CN or the whole year for SG. Heat conditions were defined as the 12 consecutive warmest weeks of each year, while normal conditions included all other weeks. 95% credible intervals were showed as a line. The colour scheme included SSP126 in blue, SSP370 in yellow, and the SPP585 in red to compare projections across different emission scenarios.

**3.3 Sensitivity analysis on varying the IIT release programme.**

We sequentially changed the overflooding ratio (OFR) from 6 in the main simulation to 1 and 3 (Figure S27–S28) or implemented an adaptive release scheme in Singapore (Figure S29) while keeping all life-history traits unchanged under GFDL-ESM4 climate model. OFR represents the release ratio of *Wolbachia* infected male mosquitoes given the historical mosquito abundance for the specific area.

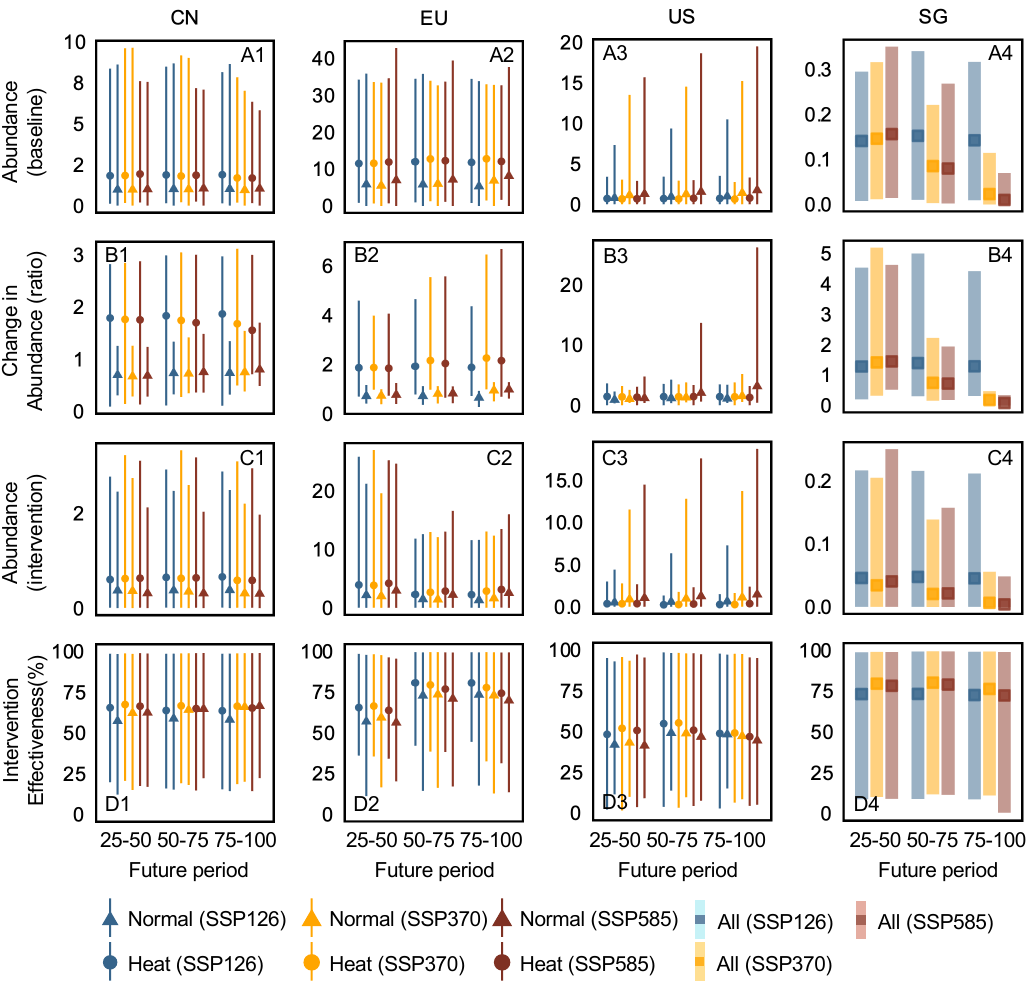

**Figure S47. Model projections with corresponding intervention effectiveness (IE) with OFR=3.** The series of box plots summarized the projected abundance in the baseline scenario (A1-4), change in abundance (B1-4), abundance in intervention scenario (C1-4), and intervention effectiveness (D1-4) for each study region, namely, mainland China (CN), Europe (EU), the United States (US), and Singapore (SG) across three future time periods, 2025–2050, 2050–2075 and 2075–2100. Change in abundance was taken as the projected future mosquito abundance divided by historical mosquito abundance. Intervention effectiveness is defined as the **percentage reduction** in mosquito abundance achieved by implementing IIT interventions relative to the baseline scenario of no IIT interventions. Simulations assumed a 2:1 overflooding ratio. The figure visualized projections under the GFDL-ESM4 model. Each panel visualized seasonal changes in heat and normal conditions for EU, US and CN or the whole year for SG. Heat conditions were defined as the 12 consecutive warmest weeks of each year, while normal conditions included all other weeks. 95% credible intervals were showed as a line. The colour scheme included SSP126 in blue, SSP370 in yellow, and the SPP585 in red to compare projections across different emission scenarios.

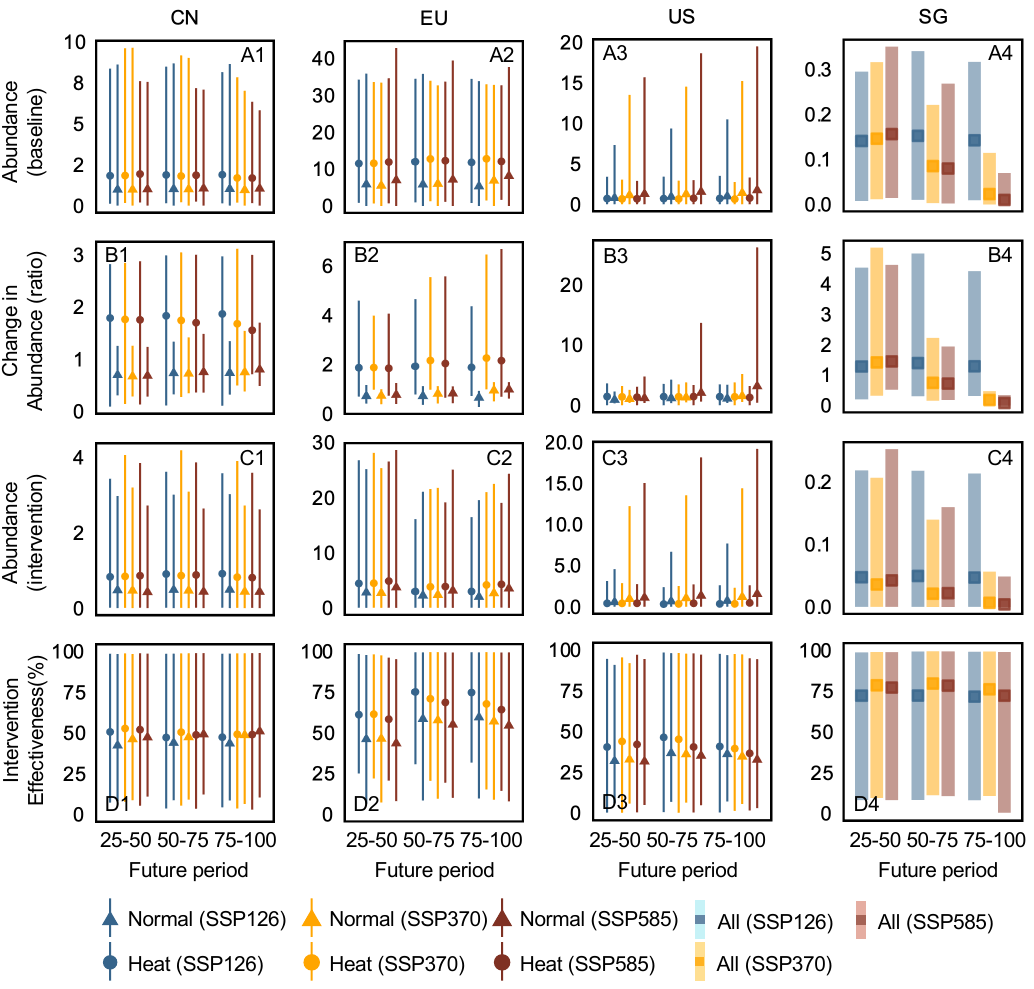

**Figure S48. Model projections with corresponding intervention effectiveness (IE) with OFR=1.** The series of box plots summarized the projected abundance in the baseline scenario (A1-4), change in abundance (B1-4), abundance in intervention scenario (C1-4), and intervention effectiveness (D1-4) for each study region, namely, mainland China (CN), Europe (EU), the United States (US), and Singapore (SG) across three future time periods, 2025–2050, 2050–2075 and 2075–2100. Change in abundance was taken as the projected future mosquito abundance divided by historical mosquito abundance. Intervention effectiveness is defined as the **percentage reduction** in mosquito abundance achieved by implementing IIT interventions relative to the baseline scenario of no IIT interventions.Simulations assumed a 4:1 overflooding ratio. The figure visualized projections under the GFDL-ESM4 model. Each panel visualized seasonal changes in heat and normal conditions for EU, US and CN or the whole year for SG. Heat conditions were defined as the 12 consecutive warmest weeks of each year, while normal conditions included all other weeks. 95% credible intervals were showed as a line. The colour scheme included SSP126 in blue, SSP370 in yellow, and the SPP585 in red to compare projections across different emission scenarios.

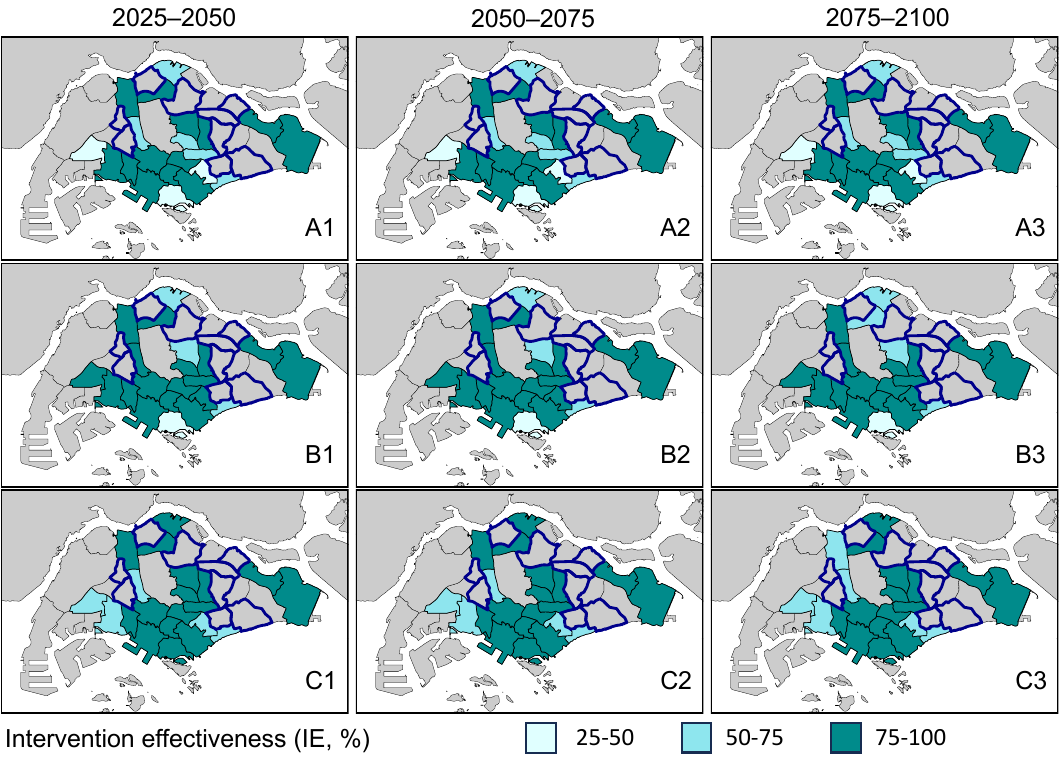

**Figure S49. Projected intervention effectiveness of adaptive IIT under future climate conditions by different timeframes in Singapore.** Maps show the projected intervention effectiveness (IE) for Singapore across three future time periods: 2025–2050 (column 1), 2050–2075 (column 2) and 2075–2100 (column 3). IE defined as the percentage reduction in female mosquito abundance in the intervention scenario compared to a baseline scenario without intervention, under future climate projections. Simulations imposed an adaptive release scheme, and are driven by the GFDL-ESM4 climate model. Rows show regional projections for SSP126 (A), SSP370 (B), and SSP585 (C). In Singapore, the areas with ongoing IIT-SIT deployment were excluded in the study, as data do not reflect natural *Aedes* population dynamics.
